# Describing health inequalities without distortion: Simple-Means MAIHDA vs Random-Effects MAIHDA

**DOI:** 10.64898/2026.08.17.26360592

**Authors:** Juan Merlo, Nasir Z Bashir, Merida Rodriguez-Lopez, Kani Khalaf, Johan Öberg, Raquel Perez-Vicente

**Affiliations:** Unit for Social Epidemiology, Faculty of Medicine, Lund University, Malmö, Sweden; Centre for Primary Health Care Research, Region Skåne, Malmö, Sweden; MRC Biostatistics Unit, University of Cambridge, Cambridge, United Kingdom; Universidad Icesi, Cali, Valle del Cauca, Colombia

## Abstract

Multilevel Analysis of Individual Heterogeneity and Discriminatory Accuracy (MAIHDA) describes health inequalities through three components: (i) specific contextual effects (SCE), (ii) general contextual effects (GCE), and (iii) discriminatory accuracy of the context. We present Simple-Means MAIHDA (S-MAIHDA), which estimates each stratum directly from its observed individuals, with no distributional assumption. The observed proportions are unbiased whatever the stratum size, and their confidence intervals report the uncertainty honestly. S-MAIHDA operationalises the three components on the probability scale. The SCE are the raw and composition-standardised prevalences, and the exact split of every observed stratum prevalence into a reference, the absolute risk difference (ARD) of its area, the ARD of its sociodemographic stratum common to all areas, and the modification of that ARD by the area. The GCE are the variance partition coefficient (VPC), the standardised contextual area under the ROC curve (coAUC*), which allows comparison across outcomes, and the contextual structuring of the between-stratum inequality, expressed as the contextual clustering of inequalities, the additive sociodemographic differences, and the contextual modification of inequalities (CMI). The contextual discriminatory accuracy is expressed by the contextual area under the ROC curve (coAUC), and the sensitivity and specificity at the population prevalence as the threshold for a possible intervention. Because its estimates are the observed data themselves, S-MAIHDA is the canonical description, and the *compare* diagnostic quantifies how Random-Effects MAIHDA (RE-MAIHDA), the usual implementation, departs from it: RE shrinkage pulls small strata towards the overall mean and can hide the very inequalities the analysis seeks. Every measure is computed from a table of counts per stratum, so the individual records need not be shared. The approach is implemented in the *smaihda* Stata command and reproduced in free Python and R code. We illustrate S-MAIHDA in a cross-sectional analysis of 2006 register data from Malmö, Sweden (43,291 individuals; 300 area-sociodemographic strata), showing how the three components separate two contrasting outcomes: psychotropic medication use, almost purely sociodemographic, stable across areas, with weak contextual structuring (VPC ≈ 4%, coAUC* ≈ 63%, CMI ≈ 0%); and choice of a private general practitioner, strongly geographical (VPC ≈ 11%, coAUC* ≈ 72%, CMI ≈ 17%), with the sociodemographic differences reshaped and amplified in wealthy areas. RE-MAIHDA attenuated inequalities. For describing inequalities, S-MAIHDA preserves what the data show.

## 1. Introduction

The Multilevel Analysis of Individual Heterogeneity and Discriminatory Accuracy (MAIHDA) is a general framework for social epidemiology that reorganises multilevel concepts to look beyond group averages and quantify how contexts structure individual heterogeneity around those averages. Its foundations were laid by uncovering the informative contrast between measures of variation and measures of association [1] and the use of the intraclass-correlation coefficient as (ICC) an expression of contextual phenomena [2]. It leads to a research framework that distinguished general from specific contextual effects and questioned the prevailing means-centric reductionism in epidemiology [3].

The same insight was almost simultaneously developed in the literature on the evaluation of individual-level markers, where it was shown that a strong measure of association can coexist with poor discrimination [4]. That work concerned individual-level indicators and the use of the area under the ROC curve (AUC) as a measure of discriminatory accuracy, while the counterpart we developed focused on social and geographical contexts and used the ICC together with a contextual AUC to measure contextual phenomena. In the rest of this paper the contextual AUC, coAUC, refers to the discriminatory accuracy of the context rather than of individual characteristics (they should not be conflated).

While both the AUC and the ICC convey similar information, they also complement each other, and discriminatory accuracy was introduced in social epidemiology and multilevel analyses [5, 6]. The three main elements in MAIHDA, association, variance and discriminatory accuracy, were brought together in 2014 [7]. The full acronym itself was coined in 2018 [8] when commenting on a multilevel analysis of intersectional interaction of effects [9]. Since then, random effects (RE) intersectional MAIHDA (RE I-MAIHDA) has popularised the framework [9–12] and with it the study of multidimensional contexts. However, intersectional MAIHDA without RE was published in 2017 [13, 14].

The MAIHDA framework is rooted in the geographical epidemiology developed by Duncan, Jones and Moon [15–19] in the UK, in the work of Davis and colleagues on health care in New Zealand [20–22] in the work of Boyle and Willms on public health in Canada [23], and in the multilevel analysis of variance developed by Goldstein [24]. We developed this work in social epidemiology [2, 3, 5–7, 13, 14, 25–33] and healthcare quality research [34–41]. The framework is being used and developed since then [8, 10, 33, 42–56].

Therefore, rather than being a specific analytical model, MAIHDA is an analytical framework for studying contextually structured individual inequalities. It is theoretically and statistically flexible, enabling context to be conceptualised in different ways and operationalised through a variety of statistical approaches. A plain-language account of MAIHDA is given elsewhere [52]

MAIHDA reflects a broader shift in how research reasons about populations and contexts: from a mean-centric approach to one informed by variance and discriminatory accuracy [5, 7, 32, 51, 57]. The mean-centric approach reduces a group to its average and reads that average as a property of its members. Used alone, averages can lead to ineffective interventions and to the unnecessary stigmatisation of areas and social groups. Average measures of association are abstractions that do not represent the heterogeneity of individual responses around the average. An approach informed by variance and discriminatory accuracy treats that heterogeneity as the substantive phenomenon. The variance is not a measure of statistical uncertainty, which is the role of the standard error, but a natural quantity that expresses the real interindividual heterogeneity of responses [7]. Read as content rather than as noise, it is what lets group context be studied as a force that structures individual lives. This recognition is not confined to epidemiology. The same argument has been made in fields as different as political science and evolutionary biology [7, 52]. A more extended genealogy is treated in a forthcoming JECH commentary [57]. MAIHDA is a consolidated analytical expression of this shift.

Within MAIHDA, the analysis is organised around three complementary components: (*i*) specific contextual effects (SCE), which map the average inequalities (where they lie and how large they are on average); (ii) general contextual effects (GCE), which quantify how strongly context structures individual heterogeneity; and (*iii*) contextual discriminatory accuracy (DA), which assesses how well context membership classifies individuals according to the outcome. The three components together provide a deeper picture of inequality than any single measure alone. MAIHDA is a way of thinking about inequality rather than a new statistical model.

The “M” in MAIHDA is conceptual. Individuals live in contexts, and so there is a multilevel structure. From this conceptual starting point, MAIHDA remains agnostic in three respects. First, it is agnostic about a specific substantive theory of how contexts come to matter. Second, it is agnostic about the dimensionality of context, which may be unidimensional, such as a single geographical level, or multidimensional, such as the intersection of several social positions. Third, it is agnostic about statistical technique. In practice, MAIHDA has most often been implemented through random effects, RE, multilevel models or RE-MAIHDA. RE models assume that contexts are exchangeable realisations of a common distribution with a grand mean and a between-context variance. To improve prediction’s reliability the context-specific estimates are therefore shrunken towards the grand mean, a process known as partial pooling or shrinkage, which is most severe for small strata when between-context variance is low. The same operation narrows the interval around each estimate, so an attenuated value can appear lower and more certain than the data warrant.

The problems caused by shrinkage in the study of inequalities are well known [58]. The epistemic critique of shrinkage has recently been renewed by Bashir [59] for intersectional analyses of small strata. Shrinkage smooths over precisely the inequalities that analyses in social epidemiology meant to capture, and the assumption of exchangeability conflicts with the principle that specific social positions give rise to distinct lived experiences. RE predictions describe a population defined by the model’s own assumptions of exchangeability, common variance and normally distributed intercepts, rather than the population the data show.

RE assumptions distort and smooth the very inequalities that social epidemiology aims to detect. However, that does not make RE wrong. It makes them suited to a different question: prediction under exchangeability, not the description of realised inequality. Nevertheless, MAIHDA remains unaffected as a framework. Its structural commitment, that individual heterogeneity is contextually organised in a nested data structure, is preserved under any statistical estimator faithful to that structure.

The MAIHDA approach can be operationalised not only through random-effects multilevel models, but also through fixed-effects regression and through the simple-means approach, as we have previously illustrated [53, 54, 60–66]. We denominate this approach simple-means MAIHDA (S-MAIHDA). Simple means impose no distributional assumption and estimate each stratum directly from its observed individuals.

Behind the choice of estimator in MAIHDA lies an epistemological question. Which reality should a study of inequality quantify: the reality observed in the population, or the reality inferred from a statistical model? The question is sharpest for the smallest strata, which are often the most marginalised. An observed stratum proportion is unbiased whatever the size of the stratum. What a small stratum lacks is precision, and the confidence interval reports that lack directly. Shrinkage does not supply the missing precision. It replaces it with an assumption about how the strata are distributed. This is not a balance to be attained between faithful description and statistical stability: description and prediction have different estimands, and for description the observed proportion is the estimand itself. When marginalised groups are small, measuring inequities may not be possible using quantitative methods, because of statistical noise. That is a problem which cannot be modelled away, and where the data cannot support a firm statement about a stratum, the scientific response is to report the imprecision rather than remove it by assumption.

Simple means are the observed stratum-specific averages computed directly from the individuals in each stratum. They are unbiased maximum likelihood estimates: the values that most faithfully represent what was observed in each stratum. This raises a practical question: do simple means suffice to operationalise the three MAIHDA components, SCE, GCE and DA, and thereby to describe contextual inequalities, or is the random-effects estimator needed? We investigate this question using data from Malmö, Sweden, previously published as part of a MAIHDA study [33]. Our study is empirical, but we also structure the paper in detail, so the reader understands all the concepts and obtains the Stata program, and its free Python and R versions, to repeat the analysis on the attached dataset or on their own data. We analyse two outcomes: use of psychotropic medication and choice of a private general practitioner. As we previously showed, the two outcomes contrast sharply in how strongly context structures them [33].

The remainder of the paper applies S-MAIHDA to both outcomes and compares the estimates against RE-MAIHDA, reflecting on the implications for inequality research. When doing so, we also develop the *smaihda* command, which performs a complete S-MAIHDA analysis. It also includes the *compare* diagnostic, which is the empirical instrument for deciding whether RE-MAIHDA is unbiased in any given dataset: it makes the shrinkage operation of RE-MAIHDA visible and measurable, allowing the reader to assess its appropriateness for their own analyses.

## 2. Study population and analytical approach

### 2.1 Study population and variables

This is a cross-sectional analysis of a register-based dataset covering one calendar year. The reporting follows the STROBE statement for cross-sectional studies (S1 Checklist).

In the present study we reanalyse a dataset originally published in a previous multilevel analysis of discriminatory accuracy, with contexts defined by neighbourhoods [33]. The dataset is fully anonymised and publicly available in the supplementary materials of that publication (PLoS ONE, DOI: 10.1371/journal.pone.0153778).

The study population comprises 43,291 individuals aged 35 to 64 years residing in Malmö, Sweden, in 2006, with at least one contact with primary healthcare during that year, comprising most of the population aged 35 to 64 in Malmö. The data are organised in a three-level nested structure: individuals are nested within area-sociodemographic (A-SD) strata, which are in turn nested within geographical areas (Fig 1).

**Fig 1.**
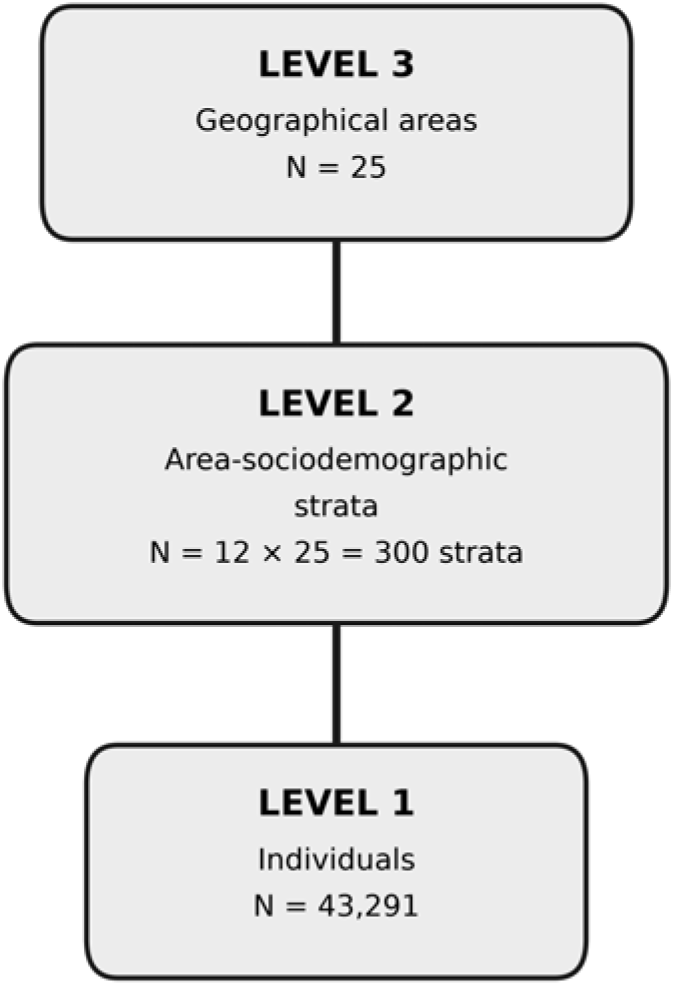
Three-level nested structure of the Malmö study. Individuals (Level 1, N=43,291) are nested within area-sociodemographic strata (Level 2, A-SD strata, N=300), which are in turn nested within geographical areas (Level 3, N=25). The A-SD strata are formed by the cross-classification of 12 sociodemographic categories (sex × age × income) with the 25 geographical areas. The figure illustrates that the present study has a multilevel structure in the conceptual sense, independent of the statistical estimator used to analyse it.

#### Individuals and outcomes (Level 1)

Two binary outcomes are examined. The first is use of *psychotropic medication* during 2006, defined according to ATC codes N05B, N05C, and N06A, with a population prevalence of 26.0%. The second is *choice of a private rather than a public general practitioner* during 2006, with a prevalence of 22.1%. These two outcomes were selected because in the previous study [33] we observed that they present very different patterns of contextual structuring, making them well suited for illustrating the full range of results that the analytical approach can produce.

#### Area Sociodemographic strata (Level 2)

Three sociodemographic characteristics define the sociodemographic component of the analytical strata: *sex* (women, men), *age* group (35 to 44, 45 to 54, 55 to 64 years), and *income* (low, defined as below the Malmö median equalised household disposable income; high, defined as above it). Their full cross-classification yields 12 *sociodemographic strata*. The combination of each sociodemographic (SD) stratum with each geographical area (A) produces 300 *area-sociodemographic* (A-SD) *strata*, each representing a specific social position in a specific place.

#### Geographical areas (Level 3)

For the purpose of this illustrative example, the 218 original Malmö neighbourhoods were grouped into 25 geographical areas by random assignment with a fixed seed. While the data were already fully anonymised, the random assignment further strengthens confidentiality. A poor area is defined as one whose proportion of low-income residents is above the median across the 25 areas; otherwise, the area is categorised as rich. This procedure yields 13 poor areas and 12 rich areas.

The analytic dataset is a complete register extract for the eligible population and contains no missing values on the variables used, so no imputation or exclusion was required.

### 2.2 Analytical measures

#### The stratum table

The first step of the analysis collapses the individual records into a table with one row per A-SD stratum and outcome. Each row holds the variables that define the stratum and its area, the number of individuals and the number with the outcome. For the two outcomes of this study, the 43,291 individual records become 600 rows. Every measure in this paper can be computed from this table, and the table contains nothing beyond what the analysis needs. Its disclosure risk depends on the granularity of the strata, not on the format of the file. Expanding it, one outcome at a time, gives back the individual records of that outcome. The table is provided as S5 File. All the programs that analyse these data start from it.

All analyses are based on simple observed proportions, computed directly from the individuals in each stratum on the probability scale. Both outcomes examined here are binary, so every quantity is a proportion, which is the mean of the individual binary outcomes (0 or 1). For a continuous outcome the same quantities are simple means, and the method is analogous (Section 4.1). No random-effects multilevel model is fitted. Each observed stratum proportion, *p_stratum_*, decomposes exactly into the population proportion, *p_pop_*, and two nested deviations:

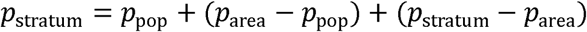

where *p_area_* is the crude proportion of the area, *p_area_*−*p_pop_* is the deviation of the area from the population proportion, and *p_stratum_*−*p_area_* is the deviation of the stratum from its area (printed by the command in its STEP 4, S1 Text). Both are expressed in percentage points. This is an exact additive identity, not a fitted model. The contextual measures use the same identity with the composition-standardised level of the area, *p*_area_*, in place of *p_area_*, and the grand mean *p\** in place of *p_pop_* (Section 2.2.1). There, all strata are given the same size and the within-area deviation is written *Δ_stratum_*. In Section 2.2.2 the individual level is added, and the deviations become the variance components of the general contextual effects.

The crude identity describes the population as it is, but its terms do not partition the inequality between strata. The crude prevalence of an area counts every individual once, so each stratum enters it in proportion to its size, and the plain mean of the deviations of the strata of an area is not zero but the composition gap of the area, the difference between its composition-standardised and its crude prevalence (Section 2.2.1). When the terms are squared and averaged, the cross-products then do not vanish, and their variances do not add up to the variance between strata: in the Malmö data they add up to 102.0% of it for psychotropic medication use and to 112.1% for private GP choice, before the noise correction. This is the classical problem of the two-way table with unequal numbers in the cells, which Yates resolved with the analysis of unweighted means, in which every cell is given the same weight whatever its size [67]. The plain mean of the stratum proportions of an area is the row mean of that analysis, and in the epidemiological tradition it is the equivalent average rate of Yule [68] (Section 2.2.1). The specific and the general contextual measures therefore take the composition-standardised prevalence of each area as its level. Standardising the level is not an adjustment added to the decomposition but the condition for an exact one. With this level every term averages zero within the level above it, and the variances of the terms add up to exactly 100% of the variance between strata. Within an area the crude and the standardised deviations differ only by the composition gap, so the ordering of the strata within an area is the same under both levels.

The probability-scale deviations are the working quantities of S-MAIHDA. Their variances in the study population feed every descriptive measure in this paper, both the Contextual Clustering of Inequalities and the variance partition coefficient (Section 2.2.2). S-MAIHDA never uses the logit scale for its measures. RE-MAIHDA is fitted on the logit scale, and both estimators are written as multilevel equations in Section 2.2.4, where the comparison between them is set out.

The two approaches differ in how the area and within-area-sociodemographic deviations are obtained. In RE-MAIHDA they are modelled as random variables drawn from a normal distribution, with variance of the areas *V_A_*, and variance of the strata *V_SD_* as estimated parameters of that distribution. In S-MAIHDA they are computed directly from observed stratum proportions, with *V_A_* and *V_SD_* as empirical descriptive statistics.

Under RE-MAIHDA, the normality assumption implies that the two random effects, one for the area and one for the sociodemographic stratum within the area, are treated as exchangeable realisations from a common distribution, which produces reliability-weighted shrinkage of the stratum estimates. Each stratum estimate is pulled towards the predicted level of its own area, and each area towards the grand mean, in which a context counts according to its reliability rather than to the number of people it represents. S-MAIHDA makes no such assumption. Each empirical deviation is computed from an observed mean, the population mean in the crude identity above or the composition-standardised mean in the decomposition used by the contextual measures, and is preserved as observed in the data, without smoothing. Fig 2 shows the two reference lines and what happens between them. For the descriptive analysis of health inequalities, this distinction matters: RE-MAIHDA attenuates contextual differences, whereas S-MAIHDA preserves them as observed, particularly in the smallest strata.

**Fig 2.**
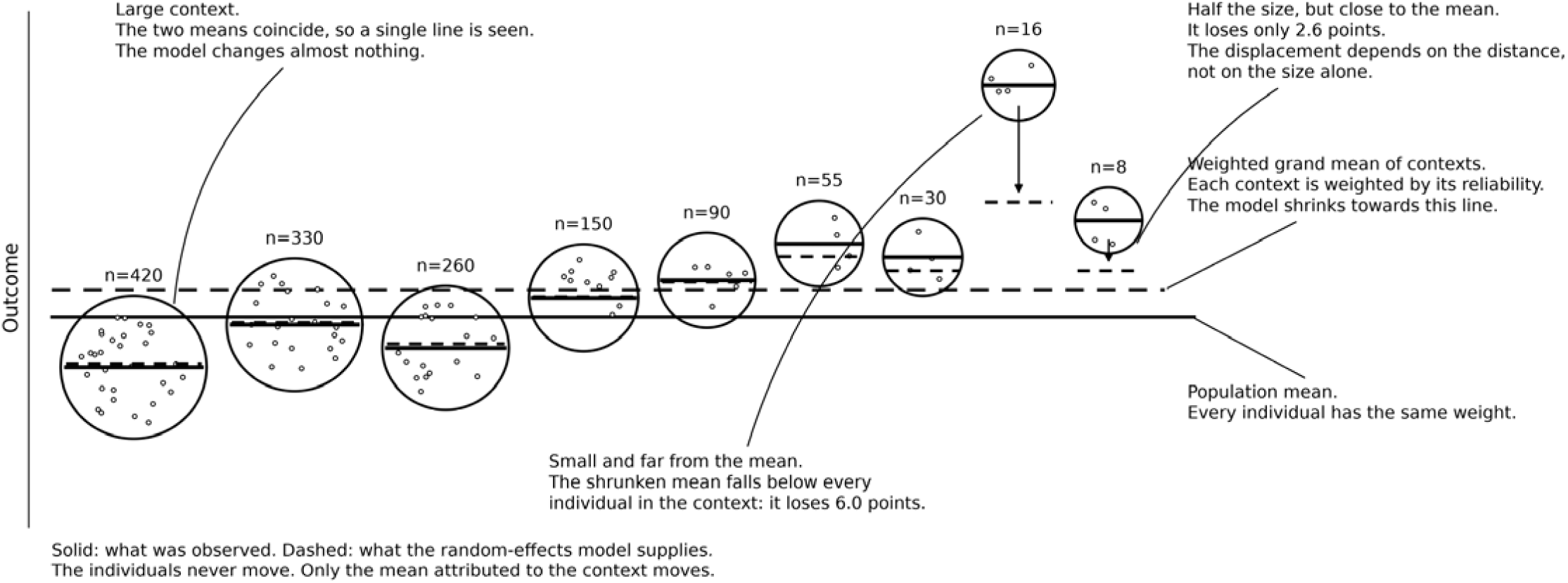
What the random-effects model changes, and what it leaves alone. The illustration uses a continuous outcome for pedagogical reasons. For a binary outcome the same reasoning applies unchanged. Each circle is a context. The area of the circle is proportional to the number of individuals it contains, and the points inside represent those individuals. The number of points drawn is only illustrative and is not equal to n. The vertical position of a point is the outcome of that individual, so the vertical spread inside a circle shows individual differences within the context. Two reference lines cross the figure. The solid line is the population mean, in which every individual has the same weight. The dashed line is the weighted grand mean of the contexts, in which each context is weighted by its reliability. This second line is what the random-effects model shrinks towards. It is neither the population mean nor a mean in which every context counts equally; it lies between them, and it approaches the second as the contexts grow large. The two reference lines differ whenever the size of a context is related to its outcome, which is the usual situation. Inside each circle the solid segment is the observed mean of that context. The dashed segment is the mean that the random-effects model attributes to it, usually called the shrunken mean. The individuals never move, and the observed mean never moves. Only the attributed (shrunken) mean moves, and it moves towards the grand mean. The displacement grows with the distance of the context from the grand mean and falls as the context grows. It is negligible in large contexts. In a small context far from the grand mean the attributed (shrunken) mean falls outside the circle, below every individual who lives there. A context of half that size but close to the grand mean is displaced less than half as much. The values are illustrative, but the displacements are not drawn by hand. Each one is computed from the standard reliability weight, which depends on the size of the context and on the variance between contexts (see S3 Text).

We operationalise the three MAIHDA components as follows.

#### 2.2.1 Specific contextual effects (SCE)

SCE map the inequalities: where they lie and how large they are on average. We operationalise the SCE in four steps. These descriptive measures parallel the quantities that RE-MAIHDA obtains from a fitted model. Here we read them directly from the observed proportions, with no model and no shrinkage. Section 2.2.4 sets the two approaches side by side.

Every SCE is a comparison of observed averages. The reference may be implicit, as when a stratum is compared with the population as a whole, or explicit, as when rich areas are compared with poor areas. Each comparison therefore carries an association, and Section 4.5 returns to what this implies for its precision.

Two conventions run through the whole section. We work on the probability scale, directly from the observed proportions. And, wherever possible, we standardise the contexts for their size, so that all strata are given the same size, whatever their number of individuals. A starred symbol denotes a standardised quantity; the same symbol without a star is a crude observed proportion. The information about how the individuals, and therefore the cases, are actually distributed across the contexts is not lost. It is reported separately by the contextual AUC (coAUC) in Section 2.2.3, which is computed from the individual counts, alongside its standardised counterpart coAUC*, in which all strata are given the same size. S8 File gives a small worked example, three areas and two strata, in a spreadsheet in which every measure is computed by formula, so that the reader can change the numbers and follow each step.

Two pairs of words are kept apart throughout. “Crude” means that a measure is computed from the individual counts, so the strata enter with their different sizes and the larger ones count more; “standardised” means that every stratum is given the same size, whatever its number of individuals. “Corrected” means that the binomial noise of the observed proportions has been removed from a variance component, and “uncorrected” that it has not (Section 2.2.2). The tables report the forms that answer each question, and the full output of the command gives all of them.

**First**, we report the observed outcome proportions for each of the 12 sociodemographic strata, the 25 areas, and the 300 area-sociodemographic (A-SD) strata. These observed proportions are the natural primary descriptive statistic for each stratum and require no model. We write the crude proportion of an A-SD stratum as *p_stratum_*, and the crude proportion of an area, its observed prevalence over all its individuals, as *p_area_*.

**Second**, to remove the influence of sociodemographic composition from the comparison between areas, we compute a standardised area proportion using the Equivalent Average Rate (EAR) method, giving equal weight to each of the 12 sociodemographic strata within each area [68]. The EAR-standardised area proportion is the plain average of the 12 sociodemographic stratum proportions within that area, each counted once,

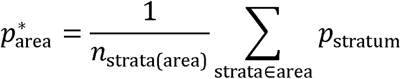

so it is the outcome the area would show if its sociodemographic groups were all of the same size. Comparing this standardised proportion with the crude area proportion *p_area_* from the first step shows how much of an area’s observed level is due to its sociodemographic composition: where the two diverge, composition matters; where they agree, the area’s level is not a compositional artefact. The EAR standardisation adjusts only for sociodemographic composition. It comes from the standardisation of rates, where it was introduced to improve the epidemiological interpretability of age-standardised comparisons [69]. The selection of an external standard population is arbitrary, and any skewness in that population influences the result. The EAR removes this problem by using a theoretical standard population with the same number of individuals in every group, which allows correctly balanced comparisons. What a single standardised figure cannot show is whether the sociodemographic differences themselves are larger in some areas than in others.

**Third**, we examine whether the sociodemographic differences vary between areas, in percentage points. From a descriptive perspective this asks whether these differences are the same everywhere; from a causal viewpoint a difference would constitute effect modification on the additive scale. For each A-SD stratum we compute its deviation from its area’s standardised proportion, on the probability scale,

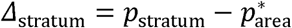

that is, how far a sociodemographic group sits above or below the level of its own area. Under a purely additive representation, the deviation of a given sociodemographic group should be similar in all areas. Systematic differences in these deviations across areas mean that the sociodemographic differences are larger in some areas than in others. This third step answers the question that RE-MAIHDA assigns to its interaction analysis. There, the answer is read from model-predicted stratum values, on the logit scale and after partial pooling. Here it is read from the observed deviations, in percentage points, with no transformation and no pooling. For describing realised inequality, this is the epistemologically correct form of that analysis: the values compared are the inequalities as observed, not as a model predicts them. These deviations *Δ_stratum_* are the empirical building blocks that Section 2.2.2 turns into the general contextual measures.

The deviations give every observed stratum proportion as the exact sum of four terms. Each is an absolute risk difference (ARD) in percentage points, taken relative to the average of the level above it and not to a reference category such as an unexposed group. The first term is the reference, p*, the plain mean of the 300 stratum proportions, all given the same size. The second is the area ARD*, the composition-standardised prevalence of the area minus the reference, p*area − p*; the star marks that the level of the area is standardised for composition. The third is the common ARD of the sociodemographic stratum, δSD, its deviation from the level of its area averaged over the 25 areas, which is the difference that is expected to repeat in every area. The level of the area plus the common ARD is the expected prevalence of the stratum if the area only shifted the level. The fourth term is the modification of the ARD by the area, γstratum, the observed minus the expected prevalence, which is what the area adds to or removes from the common difference. Where the common ARD is positive, a positive modification widens the difference and a negative one narrows it; where the common ARD is negative, the reverse. Thus pstratum = p* + (p*area − p*) + δSD + γstratum. The four terms form two blocks. The first two are area terms: the reference and the area ARD* add up to the level of the area, p*area, which says where the area stands in relation to the whole population. The last two are stratum terms: the common ARD and the modification add up to the deviation of the stratum from the level of its area, Δstratum, which says where the stratum stands within its area. The two stratum terms differ in kind. The common ARD is the part of the deviation that the stratum carries into every area. The modification belongs to the stratum in that area: it is the area-by-stratum interaction, what this particular area adds to or removes from the common difference. The expected prevalence is therefore the sum of the first three terms. Each term averages zero within the level above it, so the terms do not overlap, and their variances give the three shares of Section 2.2.2. These are the specific contextual effects of S-MAIHDA: one area ARD* per area, one common ARD per sociodemographic stratum and one modification per A-SD stratum. Fig 3 shows the four terms for one stratum in three areas, Section 3.1.3 follows them for a stratum that meets the floor of the scale, Fig 6 maps the deviations and the common ARDs, Fig 7 maps the modification of every stratum and Fig 8 draws it as observed and expected prevalences, and S8 File computes the chain by formula on a small example and draws it as in Fig 3.

**Fig 3.**
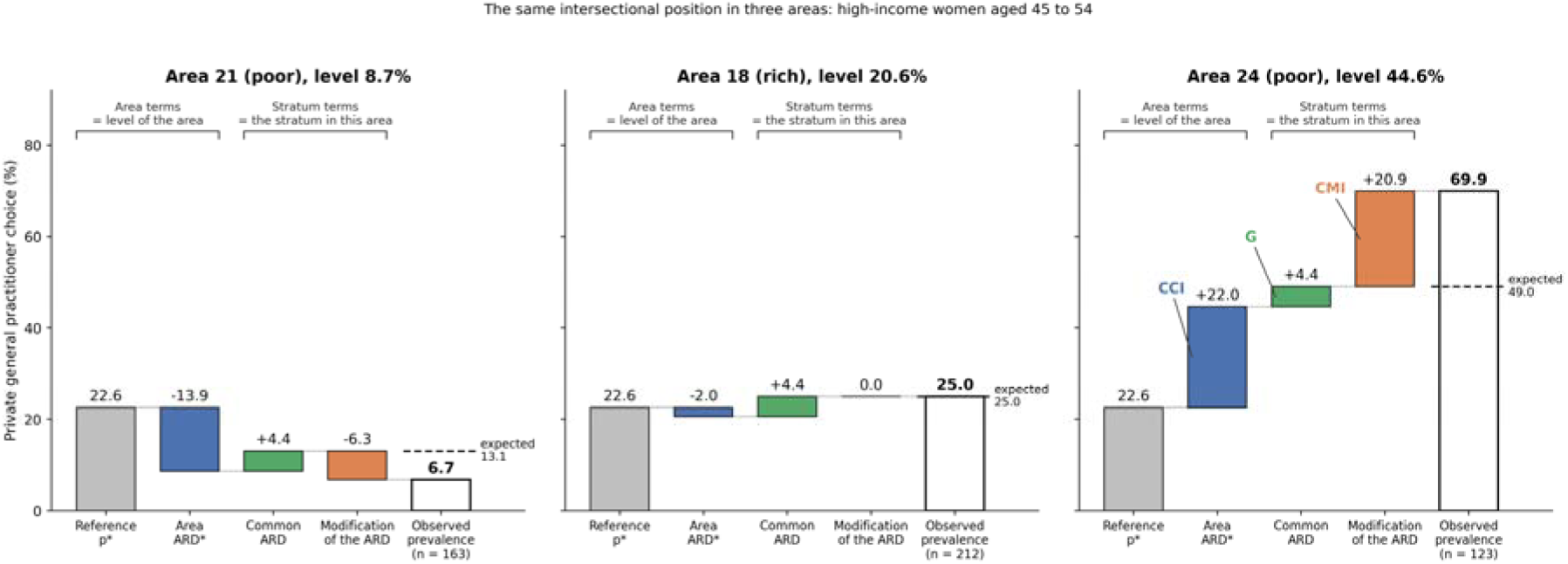
How an observed prevalence splits into four absolute differences, and what the general contextual measures summarise. The three panels show the same intersectional position, high-income women aged 45 to 54, in three areas ordered by their level, for private general practitioner (GP) choice. Malmö, Sweden, 2006. Each observed prevalence (white bar) is the exact sum of the four terms to its left: the reference p*, the plain mean of the 300 stratum proportions; the area ARD*, the composition-standardised prevalence of the area minus the reference; the common ARD of the stratum in all areas; and its modification by the area. No model is fitted. The first two terms are the area terms: together they give the level of the area (its EAR). The last two are the stratum terms: together they give the deviation of the stratum in that area from the level of its area. The common ARD is the part of that deviation the stratum carries into every area, and the modification belongs to the stratum in that area. The dashed line across the white bar marks the expected prevalence, the level of the area plus the common ARD; the distance from it to the top of the bar is the modification. All differences are absolute risk differences (ARD) in percentage points, each taken relative to the average of the level above it, not to a reference category. Across all strata, the variances of the area ARD* values, the common ARDs and the modifications give the three shares of the between-stratum inequality, CCI, G and CMI (Section 2.2.2), labelled in the third panel. In area 21 the modification reverses the common difference of the stratum, in area 18 it leaves it unchanged, and in area 24 it widens it from 4.4 to 25.3 points. S8 File draws the same figure for a small example, and the figure follows when its numbers are changed.

Each modification is read from its point estimate together with its precision, which depends on the size of its stratum. We measure the precision with the binomial standard error of the stratum proportion, computed with (cases + 1)/(n + 2) so that a stratum without cases does not get a zero error (S1 Text). The level of the area and the common ARD are averages of 12 and 25 proportions, so they add little uncertainty and are treated as fixed. To summarise the 300 modifications in one number, we count those that lie more than 1.96 standard errors from zero. If no area modified anything, about one modification in twenty, some 15 of the 300, would lie that far from zero by chance alone. The count is an exploratory description, not a hypothesis test, and the strata are read together rather than one by one.

A caution applies to the modification: the floor of the probability scale. A prevalence cannot fall below 0%, so near 0% a large negative modification cannot be fully expressed. Where the expected prevalence of a stratum is negative, its observed modification is partly the floor of the scale and not the influence of the area. The same holds at the ceiling of 100%. We report how many strata are affected and the share of the modification variance they carry (Section 3.2.2).

**Fourth**, we compare areas by their income level, as defined in Section 2.1: 13 poor and 12 rich areas. For each group we take the mean of its area proportions, and the absolute risk difference (ARD) is the difference between the two,

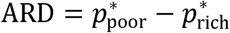

where *p*_poor_* is the mean of the 13 poor-area proportions and *p*_rich_* the mean of the 12 rich-area proportions. The relative risk (RR) is the ratio of the same two quantities. We report the ARD and RR primarily on the EAR-standardised area proportions, so that the comparison does not depend on the sociodemographic composition of the areas, and, for reference, on the crude proportions. The unit of comparison here is the area, 25 in total. The contrast describes these areas, so its interval reports how precisely each area proportion is measured, and not the variability of which areas were observed.

#### 2.2.2 General contextual effects (GCE)

GCE quantify how strongly context structures individual heterogeneity. We build them from a single variance decomposition of the outcome on the probability scale, and read from it two complementary things. First, the variance partition coefficient (VPC): the share of the total individual variation that lies between contexts. Second, a decomposition of that between-context inequality that informs on its sources: how much of the difference between the sociodemographic strata is due to the area; then, over and above the area differences, how much is a sociodemographic difference common to all areas, and how much are these differences reshaped by the area.

The purpose of the GCE is to describe how the outcome is shaped by the contextual boundaries, rather than how the burden of cases is distributed across contexts, which is the function of the discriminatory accuracy in Section 2.2.3. For this reason the contexts are standardised, so that every stratum weighs the same as the others. This isolates the influence of the contextual boundaries from context size. We report the standardised values both uncorrected and corrected for binomial noise.

##### The variance decomposition

The decomposition follows the law of total variance on the probability scale. It is the identity of Section 2.2 taken one step further: the deviations are now taken from the standardised means, so all strata are given the same size, and the comparison starts at the individual, which adds the within-stratum level. It rests on one telescoping comparison. The distance of each individual outcome *y* from the standardised grand proportion is the sum of three successive deviations,

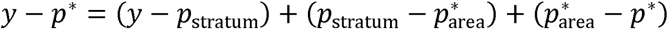

from the individual to their stratum, from the stratum to their area, and from the area to the whole. Squaring this distance and averaging gives the total individual variance, which by the law of total variance splits into three components, one per level, which can be added.

We compute each component standardised, so that all strata are given the same size, whatever their number of individuals. Let *n_strata_* be the number of A-SD strata (here 300), *n_area_* the number of areas (here 25), and *p_stratim_* the observed proportion in a stratum. From these we need two standardised averages. The grand mean *p\** is the plain average of all stratum proportions,

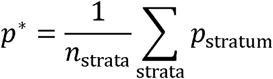

the prevalence that would be observed if every stratum held the same number of people. The area mean 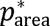 is the same average taken within one area, defined above in Section 2.2.1. Both give every stratum equal weight, so they describe the contexts themselves rather than how the population happens to fall across them.

**The between-area variance,** 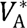. Each area carries the squared deviation of its standardised proportion from the grand mean, the same weight for every area,

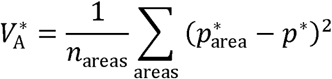

The sum runs over the 25 areas, each counted once. This is the variation between areas.

**The within-area, between-stratum variance,** *V*_SD_*. Each stratum carries the squared deviation of its proportion from the proportion of its own area,

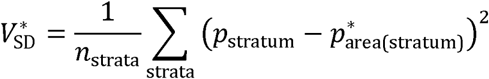

Where 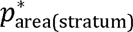 is the standardised proportion of the area to which the stratum belongs. The sum runs over the 300 strata, each counted once. This is the sociodemographic variation that remains once the area level is taken out, so we label it SD.

**The individual variance,** 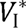. Each stratum carries the average squared deviation of its individuals from the stratum proportion. For a binary outcome this is the Bernoulli variance of the stratum,

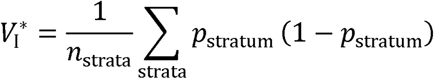

again with every stratum counted once.

The three components are the same kind of quantity, a mean of squared deviations, taken at three nested levels. They sum exactly to the total individual variance,

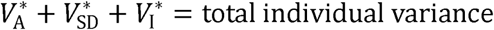

In the crude form a larger stratum contributes more to each component because it contains more individuals, so the crude form is the mean of the individual values. In the standardised form all strata are given the same size, exactly as the EAR gives every sociodemographic stratum the same size when standardising area proportions. Only the size given to the strata changes; the structure of the decomposition does not. Everything is computed on the probability scale, directly from the observed proportions.

Because the standardised form gives small strata the same weight as large ones, the sampling noise inside each observed proportion enters the between-stratum components more strongly than in the crude form. Each observed proportion contains a binomial sampling variance *p_stratum_* (1 - *p_stratum_*)/*N_stratum_*, where *N_stratum_* is the number of individuals in the stratum. We therefore report the standardised coefficients, written VPC*, in two versions: uncorrected, and corrected for this binomial noise. The correction does not remove the noise from the total, it reassigns it: the noise carried in the between-area and between-stratum components is subtracted from them and added to the individual component, so the identity is preserved exactly. What changes is where the variance is attributed, not how much there is. The exact noise terms are given in S1 Text, and S8 File shows the correction step by step on a small example. A corrected component is the observed variance minus its expected noise, so it can be negative when the true component is zero or close to it. We truncate the corrected components at zero and report a share computed from a truncated component at the boundary. An interval that reaches the boundary is truncated with it. A component at zero informs that the between-context variance is not distinguishable from the binomial noise. A share at 100% informs that the remaining components are not. At the boundary the substantive reading is what matters, and the exact width of the interval adds little to it.

*(a) The variance partition coefficient (VPC).*.

Using the variances we report three variance partition coefficients, one for each perspective on the design. Each has a crude and a standardised form; we show the standardised form here and give the crude form alongside it in the output.

The VPC at the area level is the share of the total individual variance that lies between areas,

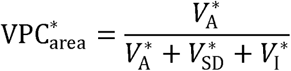

It also equals the intraclass correlation coefficient, the correlation in outcome between two individuals from the same area, computed here on the probability scale.

The combined VPC at the A-SD stratum level is the share of the total individual variance that lies between A-SD strata. It combines the area and the sociodemographic components, because every A-SD stratum is nested within an area, so the area variance is necessarily included,

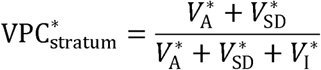

It also equals the correlation in outcome between two individuals from the same A-SD stratum.

The VPC at the sociodemographic level isolates the share attributable to the sociodemographic component on its own,

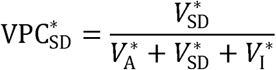

This one has no equivalent intraclass correlation, because two individuals from the same sociodemographic stratum can live in different areas and therefore do not form a single cluster in the data.

*(b) The contextual decomposition: clustering, additive differences, and modification.*.

The second general contextual measure describes how the between-stratum inequality is organised by geography. The between-stratum variance is 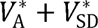, the two contextual components together. We report the decomposition standardised, giving every stratum equal weight, so that it captures the general contextual effect: the strength of the contextual boundaries themselves, independent of how many individuals fall on each side. How the burden of cases is actually distributed is a different question, answered by the discriminatory accuracy in Section 2.2.3.

The first share is the contextual clustering. The Contextual Clustering of Inequalities (CCI) is the proportion of the between-stratum variance that lies between areas,

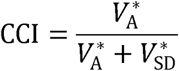

In the present paper the contextual dimension is geographical, the 25 areas of Malmö. The CCI is reported in two forms: crude, computed from the individual counts, and standardised, in which all strata are given the same size and the binomial noise is discounted. Only the standardised CCI has the split into G and CMI. In the rest of the paper, CCI without qualifier denotes the standardised form. Unlike the VPC, the CCI is built from the two contextual components alone; it omits the individual residual. A high CCI means that the between-stratum inequality is a matter of which area a stratum sits in, so that strata sharing an area resemble one another. The same logic generalises to any contextual stratification: hospitals (HCCI), schools (SCCI), workplaces (WCCI), time periods (TCCI), or any other meaningful dimension.

The remaining between-stratum variance, 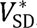, is the within-area sociodemographic variance, and we split it one step further. Recall the within-area deviation of a stratum, 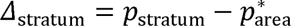 the total within-area sociodemographic difference. It splits into two parts. For each sociodemographic group, its average within-area deviation across the 25 areas, the common ARD of the group, is

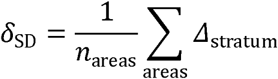

the deviation the group would show in every area if the area only shifted the level. The departure of a stratum’s actual deviation from this average is the modification of the ARD by the area,

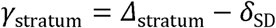

Because the modification averages to zero within each sociodemographic group, the within-area variance splits exactly into an additive part and a modification part. The additive variance is the average of the squared δ, and the modification variance the average of the squared γ,

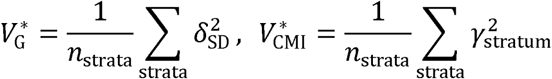

This gives three shares of the between-stratum inequality, which sum to one,

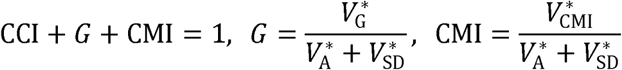

*G* is the additive part, the sociodemographic differences common to every area, and the Contextual Modification of Inequalities (CMI) is the area-by-sociodemographic interaction, how much the area reshapes these differences. A table of notation, with the default name of each term, its calculation, the footing on which it is read and the label printed by the command, is given in S1 Text (Table A).

Fig 3 shows the four terms with real data, for one intersectional position, high-income women aged 45 to 54, in three areas and for private GP choice. Each observed prevalence is the exact sum of the reference, the area ARD*, the common ARD of the stratum and its modification by the area. Across all strata, the variances of the last three terms are the three components of the decomposition: the area ARD* values give the clustering (CCI), the common ARDs the additive differences (G) and the modifications the contextual modification of inequalities (CMI). The figure groups the terms in two blocks, the area terms and the stratum terms (Section 2.2.1), and marks the expected prevalence on the observed bar.

##### Reading the three components

The additive differences *G* are the sociodemographic backbone of the inequality: the average differences between strata present in every area alike, independent of geography. The two contextual components describe what geography does to it. The CCI is a matter of level. It asks how much of the between-stratum inequality is simply which area a stratum sits in, so that strata sharing an area resemble one another. A high CCI means the areas differ in their overall prevalence and the strata cluster by area. The CMI is a matter of shape. It asks whether the same sociodemographic differences are reshaped from area to area, larger in some and smaller in others, rather than repeating identically; it is effect modification on the additive scale, the descriptive counterpart of an area-by-stratum interaction. A context can therefore act in two independent ways: it can raise or lower the level of a whole area (CCI), and it can reshape the differences within it (CMI). The additive differences say how much inequality there is to begin with; the CCI and the CMI say how much geography moves it as a block and how much it redraws its internal shape.

##### Noise correction

Each observed stratum proportion carries a binomial sampling variance. This matters most for the CMI, because the modification is a single-cell residual and carries the full sampling noise of its cell, so the uncorrected CMI is dominated by noise and overstates the modification. Unlike the VPC and the CCI, whose uncorrected form still represents the observed reality, the CMI is meaningful only once the noise is removed. We therefore report the CMI in its noise-corrected form, subtracting the expected contribution under no true modification, a binomial floor; the exact correction is given in S1 Text. The area and additive components each average over many cells, so their noise is small and their correction negligible.

##### The two forms, context conceptualisation, and when they differ

The two forms differ only in the size given to each stratum in the between-context variance, and in the reference mean. The crude form gives each stratum its number of individuals and takes deviations from the population mean *p_pop_*; the standardised form gives every stratum the same size and takes deviations from the standardised mean *p\**. They diverge when context sizes are unequal, and the divergence is largest when a small context has an extreme mean. In that case the boundary separating it from the rest is sharp, but few individuals belong to it, so its contribution to the crude variance is muted, while its contribution to the standardised variance is the same as that of any other context. This is the configuration in which a strong standardised GCE coexists with a weak crude GCE and a low discriminatory accuracy ([57], panel C; see also Section 2.2.3). The standardised form captures the GCE net of context size; the crude form, which tracks the discriminatory accuracy, does not. The two forms answer different and complementary questions and need not agree.

##### A note on the variance partition on the proportion scale

A common objection to the variance partition on the proportion scale is that it depends on the population prevalence and is therefore not a valid or comparable measure of clustering. We take the opposite view. The individual-level variance of a binary outcome is *p(1 - p)*, the variance of a Bernoulli variable. This variance is a real property of the data, and it genuinely depends on the prevalence: an outcome that affects 6% of the population has a smaller total individual variance than one that affects 50%. The proportion-scale variance partition reports this faithfully. When the same amount of between-group variance is set against a smaller total variance it represents a larger share of it, and against a larger total variance a smaller share. The dependence on prevalence is therefore not a defect of the measure. It is an honest consequence of describing the variance that the data actually contain. This individual variance, the average of the Bernoulli variances *p(1 - p)* across strata, is the same level-1 variance used in the simulation method of Goldstein, Browne and Rasbash for partitioning variance in binary multilevel models [24]. The proportion-scale partition we compute descriptively therefore matches the quantity their simulation approach targets.

The latent-scale variance partition avoids this dependence, but it does so by replacing the real individual variance with a fixed convention. It assumes that the binary outcome arises from an unobserved continuous variable thresholded at a cut-point, and fixes the individual residual variance of that latent variable at *π^2^*/3 by construction. Its prevalence-independence, and hence its comparability across outcomes and studies, is bought at the price of assuming a continuous latent variable that the data do not contain. For describing observed inequality in a binary outcome, where the quantity of interest is the variance that is actually present, the proportion scale is the faithful choice and the latent scale is an abstraction.

This has a practical consequence for comparison. Within a single study, and for the comparison between simple-means and random-effects estimates of the same outcome, the prevalence is held fixed, so the proportion-scale variance partition is directly comparable. Across outcomes or studies with very different prevalences it is not comparable, precisely because the underlying total variance differs. For that purpose, a rank-based measure such as the contextual AUC, which depends only on the ordering of strata and not on the scale of the variance, is the appropriate summary comparator. A rank-based measure does not depend on the base rate. The coAUC, however, still depends on how the population is distributed across the strata, since it counts pairs of individuals. The comparator across studies is therefore the standardised coAUC* introduced at the end of this section, which is the value that would be obtained if all strata were of equal size.

##### Reading the VPC on the probability scale

The categorisation of Table 1 was proposed for the VPC on the latent scale of a multilevel logistic model, a scale that has no direct meaning in terms of risk. On the probability scale the VPC has a meaning of its own, and it can be read in three equivalent ways (S1 Text).

**Table 1.** Epidemiological categorisation of VPC and AUC values (based on [44]).

|  |  |  |
| --- | --- | --- |
| **VPC, latent scale (%)** | **coAUC\* or coAUC (%)** | **Contextual structuring** |
| 0 to 1 | 50 to 55 | Absent |
| 1 to 5 | 55 to 61 | Very small |
| 5 to 10 | 61 to 66 | Small |
| 10 to 20 | 66 to 72 | Moderate |
| 20 to 30 | 72 to 77 | Large |
| 30 to 100 | 77 to 100 | Very large |

First, the VPC is the share of the uncertainty about the outcome of a person that is removed by knowing his or her stratum. For a binary outcome this uncertainty is the individual variance, p(1 − p). A VPC of 10% means that knowing the A-SD stratum of a person removes one tenth of it, and that nine tenths remain within the strata.

Second, the VPC is the correlation between the outcomes of two people drawn from the same stratum, the intraclass correlation of the binary outcome.

Third, the VPC is the difference between the average risk of the strata of the people with the outcome and the average risk of the strata of the people without it. Tjur [70] proposed this difference as a coefficient of determination for logistic regression models with individual predictors. In Simple-Means MAIHDA the only predictor is membership of a stratum, and the predicted risk of each person is the observed proportion of his or her stratum. With that prediction the coefficient of Tjur is the VPC on the probability scale, the between-stratum variance over the total individual variance of the binary outcome, so that it is also the intraclass correlation and the share of the individual variance explained by the strata. A VPC of 10% means that the people with the outcome live, on average, in strata whose prevalence is 10 points higher than that of the strata of the people without it. This reading links the general contextual effect with the discriminatory accuracy. The VPC measures how well the strata separate the people with and without the outcome by the difference between their average risks, and coAUC* measures the same separation by ranks. It also explains why the VPC depends on the prevalence, since when an outcome is rare both average risks are small, and so is the difference between them. coAUC*, based on ranks, barely depends on the prevalence, which is why it is the measure compared across outcomes.

The three readings are exact for the crude VPC, in which the strata enter with their different sizes. For VPC* they hold in a population in which all strata are given the same size, and the correction for binomial noise removes the part of the difference that small strata produce by chance (S1 Text). S8 File checks the third reading by formula on a small example.

##### The standardised contextual AUC (coAUC*) as a comparable reading of the GCE

The VPC is not the only possible summary of the general contextual effect. A rank-based summary is also available, and it is the one that survives a comparison across outcomes and studies. The contextual AUC (coAUC), defined in Section 2.2.3, is the probability that a randomly selected individual with the outcome belongs to a stratum with a higher observed proportion than a randomly selected individual without it. It counts pairs of individuals. A stratum of 3,000 individuals therefore contributes a million times more pairs than a stratum of 3, and the value obtained depends on how the population happens to be distributed across the strata, and not only on how sharply the strata separate individuals by risk. We also compute it with every individual weighted by the inverse of the size of their own stratum, so that every stratum contributes the same total weight. Following the convention introduced at the start of Section 2.2, we write it coAUC*, the standardised contextual AUC. It is the value that would be obtained if all strata were of equal size. It is the same expression as the coAUC (S1 Text, Step 8), with every stratum size set to 1, and ties counted at one half.

coAUC* is a standardised measure in the ordinary epidemiological sense and should be read as one. A rate standardised by age is real arithmetic on a hypothetical population whose age composition is fixed by the standard. coAUC* is real arithmetic on a hypothetical population whose strata are of equal size. Neither describes the population that was observed. Both are useful for the same reason, that is, they remove from the comparison a feature that is not the object of study. The star carries the same meaning here as it does for the composition-standardised area prevalences of Section 2.2.1 and for VPC* above.

coAUC* informs on the general contextual effect in a way that parallels VPC*. The two order the contexts alike, whatever the distribution of stratum sizes, while the coAUC departs from that ordering as the sizes become unequal (S7 Text). The VPC on the probability scale is not comparable across outcomes or studies of different prevalence, for the reason given above. coAUC* does not share that limitation, since a rank-based measure does not depend on the base rate. coAUC* is therefore the general contextual effect read on the discrimination scale, and it is the reading to use when the comparison crosses outcomes or studies.

coAUC* requires no continuity correction. It uses only the ordering of the observed stratum proportions, and a stratum with no cases has a well defined rank. coAUC* is computed on the strata with at least ten individuals, the minimum applied to every standardised measure, since giving every stratum the same size would otherwise give a stratum of 3 the weight of a stratum of 3,000. We report coAUC* at both contextual levels, area and A-SD stratum, with the same within-context bootstrap interval as every other measure. The coAUC is reported alongside it in Section 2.2.3, where it answers a different question, how the burden of cases is in fact distributed across the contexts.

At the area level, coAUC* gives every area the same weight but keeps its composition, since each area is scored by its crude prevalence. We therefore also report coAUC*EAR, in which every area is scored by its composition-standardised prevalence, the EAR of Section 2.2.1. It is the area-level coAUC with every stratum given the same size, standardised for size and composition, and it is the counterpart of the area VPC* on the discrimination scale. The difference between coAUC* and coAUC*EAR is the part of the general contextual effect of the area that comes from its composition. At the level of the A-SD strata the two coincide.

#### 2.2.3 Discriminatory accuracy of the context (DA)

DA assesses whether context membership is useful for classifying individuals according to their outcome. It is summarised by the contextual area under the receiver operating characteristic curve (coAUC), computed from the observed stratum proportions (S1 Text, Step 8). The coAUC is the probability that a randomly selected individual with the outcome belongs to a group with a higher predicted risk than a randomly selected individual without it. All individuals in the same stratum are assigned the same predicted risk. Under S-MAIHDA that predicted risk is simply the observed proportion of the stratum, so no model prediction is involved, and under RE-MAIHDA it is the shrunken predicted value that the model attributes to the stratum. DA is a property of the classification rather than of the data. Computed from contextual classifications, as in MAIHDA, it describes the discriminatory information carried by the context, not individual-level information, though the two are sometimes conflated. The coAUC is therefore a contextual AUC, the discriminatory accuracy of the context: the predictor is the individual’s context, and every individual in a stratum receives the same score. It should not be read as an individual-level AUC, which uses individual characteristics as predictors.

Because every individual is scored by the observed proportion of their stratum, the classification always operates on groups. The rows of the prediction matrix are strata, not individuals, and each stratum contributes a single predicted risk shared by all who belong to it. What the coAUC measures is therefore the discriminatory accuracy of the groups: how well context membership orders individuals by risk. An individual-level AUC would need predictors that vary within a stratum, which the proportion-based approach does not use. Using proportions, the coAUC is always a group measure, whatever label is attached to it.

Three AUCs therefore need to be distinguished. (i) An individual-level AUC, not used here, is computed from the risks that a model with individual-level variables, such as a logistic regression, predicts for each person, and measures the discriminatory accuracy of those characteristics. (ii) The contextual AUC, coAUC, is computed from the observed prevalences of the contexts, so every individual in a stratum receives the same score, and it measures the discriminatory accuracy of the context. (iii) The standardised contextual AUC, coAUC*, is the coAUC with all strata given the same size, and it measures the general contextual effect, like VPC* (Section 2.2.2).

To calculate the coAUC a sliding risk threshold is moved across the range of observed stratum prevalences. At each threshold, individuals belonging to strata with a prevalence above the threshold are classified as predicted positives, and the rest as predicted negatives. This yields the classification table (true positives, false positives, true negatives, false negatives), from which sensitivity (true positive rate) and 1−specificity (false positive rate) are derived. The coAUC summarises the trade-off between sensitivity and 1−specificity across all thresholds. S8 File computes the coAUC and coAUC* by counting pairs of individuals on a small example.

The coAUC is a crude measure, computed from the individual counts, and therefore informs on how the burden of cases is distributed across the contexts. Its standardised counterpart coAUC*, introduced in Section 2.2.2 and reported with the other general contextual measures in Table 5, gives all strata the same size and informs on the general contextual effect. Both are reported.

##### Classification at a threshold: true and false positives

The coAUC summarises discriminatory accuracy across all possible thresholds. A concrete decision, however, requires a single threshold, at which each stratum is classified as high-risk or not. The two readings are complementary: the coAUC describes the ranking of the strata, and the classification at a threshold describes what acting on that ranking would achieve.

We use the population prevalence as the default threshold. This is not an arbitrary choice but a parameter of the data, the mean outcome across all individuals. A stratum is classified as high-risk when its predicted proportion exceeds the population prevalence, and every individual is classified through the stratum they belong to.

Against the observed outcome this yields the same four cells used to trace the ROC curve above, now held at a single fixed threshold and counted in individuals. A true positive is an individual with the outcome in a high-risk stratum. A false positive is an individual without the outcome in a high-risk stratum. A false negative is an individual with the outcome in a stratum not flagged. A true negative is an individual without the outcome in a stratum not flagged. We report the counts, together with the number of strata flagged. From the cells follow the sensitivity, the share of all cases that fall in the high-risk strata, the specificity, and the positive predictive value, the share of flagged individuals who have the outcome.

The false negatives, the individuals with the outcome who live in strata not flagged, operationalise Rose’s prevention paradox, since they are the cases that any strategy directed at the high-risk strata leaves out. The false positives, the individuals without the outcome who live in high-risk strata, are those that the same strategy would reach without need. The larger these two shares, the stronger the case for a universal component in the intervention, proportionate to the position of the strata.

The threshold can be set otherwise. In many applications it follows an a priori criterion of the decision at hand, for example a recommended proportion of patients to be treated, a clinical cut-point, or a quality benchmark. The population prevalence is the natural default because it separates the strata that sit above the average from those below it, but the machinery is unchanged for any threshold the user specifies.

This classification makes the discriminatory accuracy actionable, and it is where the choice of estimator becomes visible in practice. Under S-MAIHDA the predicted proportion is the observed one; under RE-MAIHDA it is the shrunken value. When shrinkage moves a small high-risk stratum below the threshold, that stratum is no longer flagged, and its cases move from true positives to false negatives. Comparing the two classifications at the same threshold therefore shows, in counts of people, what the random-effects assumption would change (Section 2.2.4).

##### Levels of DA

We distinguish two levels of DA: an area-level DA computed using area prevalences, and an A-SD-level DA computed using A-SD stratum prevalences. The former quantifies the discriminatory information carried by geography alone; the latter, by geography and sociodemographic position jointly.

Together with the VPCs and the CCI, the area-level and A-SD-level DA complete the three perspectives on contextual structuring proposed in this paper: how much of the total individual variance lies between contexts (VPC), how that between-stratum inequality is geographically structured, both clustered by area level and reshaped within it (CCI and CMI), and how useful context membership is for classifying individual outcomes (DA).

##### Epidemiological interpretation of coAUC* and coAUC

To facilitate the epidemiological interpretation of the general contextual effect and of the discriminatory accuracy, we use a categorisation similar to that proposed by Merlo, Wagner and Leckie [44], shown in Table 1, which translates these statistical measures into substantive judgements about the degree of contextual structuring. The categorisation was proposed for the VPC on the latent scale of a multilevel logistic model and for its equivalent AUC. We apply it to coAUC* to rate the general contextual effect (Section 3.2.2) and to the coAUC to rate the discriminatory accuracy (Section 3.3). The VPC on the probability scale is reported without a rating, because it has a direct reading of its own (Section 2.2.2).

#### 2.2.4 Comparison between Simple-Means MAIHDA and Random-Effects MAIHDA

S-MAIHDA and RE-MAIHDA differ in the distributional assumption applied to the contextual components. RE-MAIHDA assumes that stratum-specific intercepts are exchangeable realisations of a common normal distribution and, accordingly, pulls each prediction toward the grand mean by an amount that scales inversely with stratum size and with the between-stratum variance. The smallest strata, which often represent the most marginalised subpopulations, are shrunk the most. The empirical consequences of this shrinkage are not assumed in advance; they are evaluated for any given dataset through the compare diagnostic of the smaihda command, which produces a quantitative report on the agreement and divergence between the two estimators across the three MAIHDA components.

Section 4.4 illustrates how shrinkage operates and why it matters for descriptive inequality.

The compare diagnostic fits a standard Random-Effects MAIHDA via melogit on the same A-SD strata. It returns nine quantitative measures distributed across the three MAIHDA components: five for SCE, one for GCE, and three for DA. All nine are formally defined, with their values for both outcomes, in S1 Text and S3 Text. In the main text we restrict attention to the six measures listed in Box 1. The set is extensible: one could add, for example, a measure of how much narrower the random-effects confidence intervals are for the smallest strata, an apparent precision that accompanies the shrinkage of the point estimate.

For the comparison to be meaningful, both estimators must be placed on the same footing, in scale and in weighting. In scale, the Simple-Means VPC is already on the probability scale. The Random-Effects VPC is placed on the same scale by simulation: stratum residuals are drawn from the fitted normal distribution, each draw is converted to a probability, and the VPC is formed as the variance of these probabilities over their mean Bernoulli variance. This is the simulation method of Goldstein, Browne and Rasbash [24].

In weighting, the general contextual effect describes how strongly the contextual boundaries structure risk, independently of how many individuals each context contains. It is therefore compared using the Simple-Means VPC*, in which all strata are given the same size. Giving all strata the same size matches the way the random-effects τ² treats strata as exchangeable units rather than weighting them by the number of individuals they contain, and the command reports it uncorrected and corrected for binomial noise. Once both estimators are on the probability scale and expressed as VPC*, their difference reflects the distributional assumption, not a difference of scale or weighting. The relation between log-odds and probabilities is not linear, so the same latent-scale VPC corresponds to different degrees of heterogeneity in observed risk depending on the prevalence of the outcome. Because inequalities are experienced as differences in risk, not in log-odds, we read them on the probability scale.

The six selected measures reflect that shrinkage acts first on the predictions for individual strata. There are 300 such predictions, one for each A-SD stratum, so S-MAIHDA and RE-MAIHDA cannot be compared through a single difference. The comparison is made between the two sets of 300 predictions, those of S-MAIHDA and those of RE-MAIHDA, and it is summarised in two ways: how much the spread of the values is compressed, and how far the most displaced stratum has moved. The VPC and the coAUC are single numbers, so S-MAIHDA and RE-MAIHDA are compared directly on them, and they serve as summary-level agreement checks. The sensitivity and the specificity at the population-prevalence threshold are also single numbers. They expose a different consequence of shrinkage: a stratum whose prediction crosses the threshold changes the classification, even when the coAUC is unchanged. The values of these six measures for both outcomes are presented in Table 8 in Section 3.4. The operational implementation through the smaihda Stata package is provided in S1 File. A standalone conceptual guide to the nine measures, without Stata code, is provided in S3 Text.

A short example shows why both summaries are needed. Suppose that in one stratum the observed proportion, which is what S-MAIHDA reports, is 30%, and the RE-MAIHDA prediction for that stratum is 27%. Suppose that in a second stratum the observed proportion is 10% and the RE-MAIHDA prediction is 13%. Each stratum has moved by three percentage points, but the distance between the two strata has fallen from twenty percentage points to fourteen. The average across the two strata is unchanged, because the movements cancel. The range compression factor records that the spread has narrowed. The maximum absolute attenuation records the largest single movement, together with the size of the stratum in which it occurred.

**Box 1.**
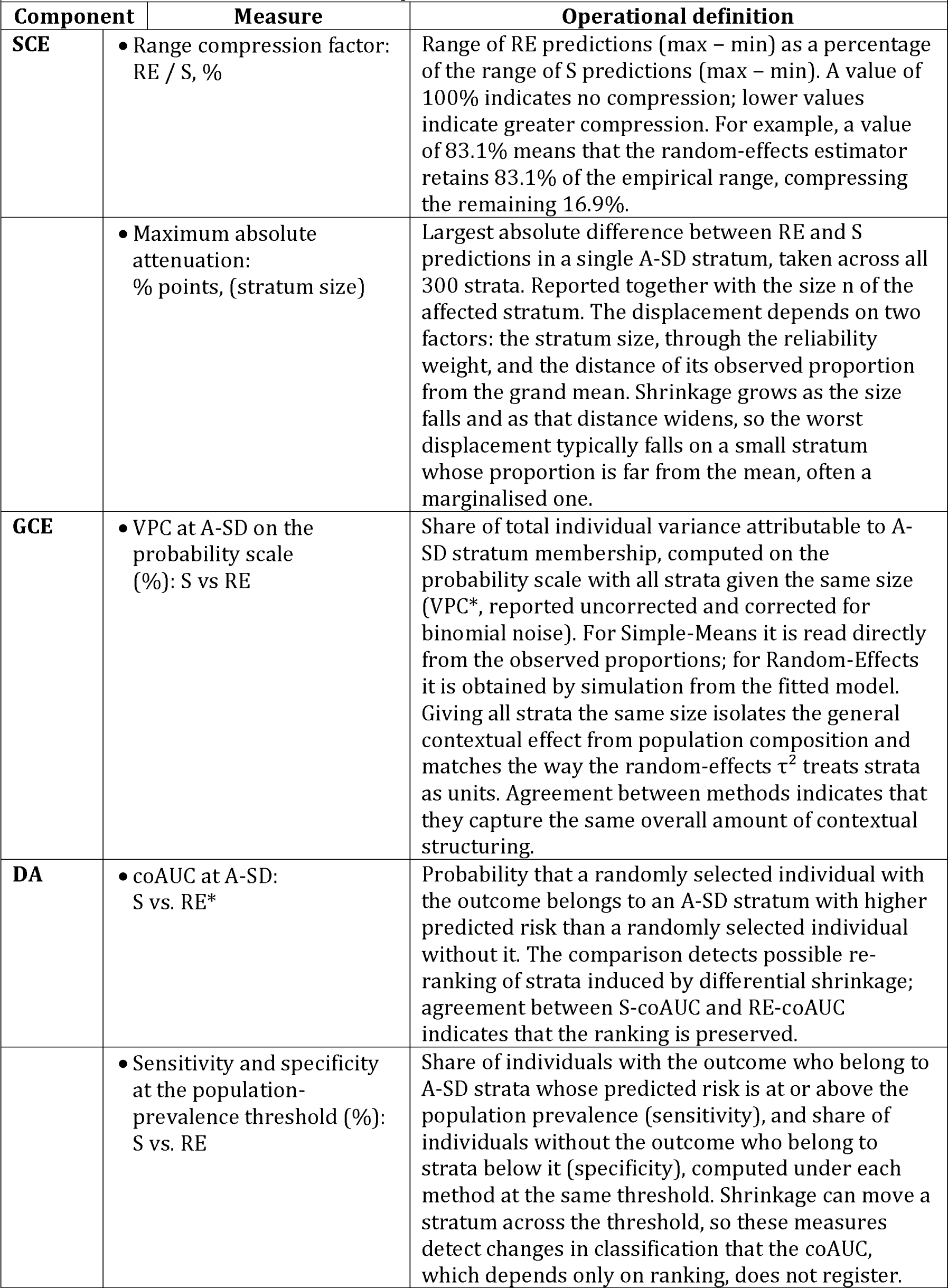
The six measures from the compare output reported in the main text. RE denotes Random-Effects MAIHDA; S denotes Simple-Means MAIHDA.

### 2.3 Software and reproducibility

All simple-means analyses were performed using Python and corroborated in R. The smaihda command for Stata version 17 (StataCorp, College Station, TX) was corroborated against them in its version 1.8.0. Some parts of the new version 1.9.0, provided in S1 File, had not yet been run in Stata when this version was posted. The random-effects comparison was fitted in Stata and R.

The S-MAIHDA pipeline is implemented as a single user-callable Stata command, smaihda, developed alongside this paper. Given a binary outcome and at least one contextual variable (an area, a sociodemographic stratum, or both), and optionally an area-level covariate, the command executes the full analytical pipeline in one call. The simple-means measures are also provided as free Python and R scripts (S2 File and S3 File).

It computes the observed proportions at each level and, from them, the EAR-standardised area proportions, the ARD and RR for the area covariate, the A-SD stratum deviations, the CCI, the VPC and ICC (each crude and standardised, the latter uncorrected and corrected for binomial noise), the contextual decomposition of the between-stratum inequality into CCI, G and CMI (with the CMI noise-corrected), and the discriminatory accuracy, including a threshold classification with its sensitivity, specificity, positive predictive value and cell counts.

Uncertainty is reported as confidence intervals, and one principle governs all of them. The contexts are the observed set and are held fixed, so what is estimated is a property of these 25 areas and these 300 strata, and the only uncertainty is the uncertainty of measuring each proportion. No interval reported here includes the variability of which areas were observed, since the analysis does not generalise to other areas. This is the same choice that separates simple means from random effects, applied to inference rather than to estimation. Within that principle the method follows the sampling structure of each measure. Observed proportions use Wilson intervals [71], the score interval for a binomial proportion, which unlike the usual large-sample interval keeps a sensible width when nobody, or everybody, in the stratum had the outcome. The standardised area proportions and the contrasts derived from them use analytical formulae. The summary contextual measures (CCI, VPC, the contextual decomposition and the coAUC) use a non-parametric within-context bootstrap with 2,000 replicates and a fixed seed (default 2026). The areas and the strata are the observed set, so they are held fixed and the individuals are resampled inside each cell. For a binary outcome, resampling the individuals of a cell is the same as drawing its number of cases from a binomial distribution with the size and the observed proportion of the cell. The bootstrap therefore also works from the stratum table. Sensitivity and specificity at the population-prevalence threshold are reported as point estimates. The method used for each measure, with the reasons for the choice, is given in S1 Text.

The dataset is the eligible population rather than a probability sample, so no survey weights apply. Uncertainty for the summary measures is estimated by a within-context bootstrap that holds the areas and the strata fixed and resamples the individuals inside each area-sociodemographic cell.

Both area() and sdstratum() are optional. Specifying only one runs a two-level analysis of individuals within contexts, whether the contexts are geographical or institutional units or intersectional strata. In that case the command reports the single VPC, crude and standardised, together with the coAUC, and omits the CCI, which requires a second contextual dimension.

An optional *compare* keyword fits a standard RE-MAIHDA on the same A-SD strata and reports the comparative measures described in Section 2.2.4. Full documentation is accessible through the standard Stata help system (*help smaihda*).

The two analyses reported in this paper are reproduced in Stata from the stratum table (S5 File) with:

~~~
. smaihda cases if outcome == “psycmed”, area(area) sdstratum(strata_sociodem_id) areacov(area_income) n(n) compare
. smaihda cases if outcome == “private”, area(area) sdstratum(strata_sociodem_id) areacov(area_income) n(n) compare
~~~

The step-by-step Stata implementation underlying the *smaihda* command, with each measure presented as a numbered step with commented code, is provided as S1 Text.

The *smaihda* package (the .ado source file, the .sthlp help file, the .pkg installation manifest and a test do-file) is provided as S1 File. The command reads the stratum table (S5 File) with the option n(), and individual records without it.

A complete reproduction of the simple-means measures in Python, using only the numpy library and requiring no proprietary software, is provided as S5 Text. It reads the stratum table, or individual records, and returns the same figures from either, including VPC* and its bootstrap intervals, so the analysis can be checked end to end without a Stata licence.

An independent reproduction in base R, with no packages, is provided as S6 Text. Like the Python script, it reads the stratum table or individual records. Its point estimates agree with the Python implementation at every printed figure, and its bootstrap intervals agree within Monte Carlo error. The values reported here are from version 1.9.0 of the Python and R scripts. The programs that draw Figs 3 to 10 from the stratum table (S5 File) are provided as S6 File, together with a program that computes, and checks against the text, the numbers of Sections 2.2, 3.1.3 and 3.2.2 that the command does not print. The latent-scale quantities of Section 3.4 are printed by the program of Fig 10.

Once the command is installed (placed in Stata’s PERSONAL ado-path), the entire analysis presented in this paper can be reproduced from the stratum table with the two command lines shown above.

## 3. Results

### 3.1 Specific contextual effects: mapping and understanding average risk

#### 3.1.1 Sociodemographic strata

Table 2 shows consistent and substantial income differences for both outcomes across all sex and age groups.

**Table 2.**
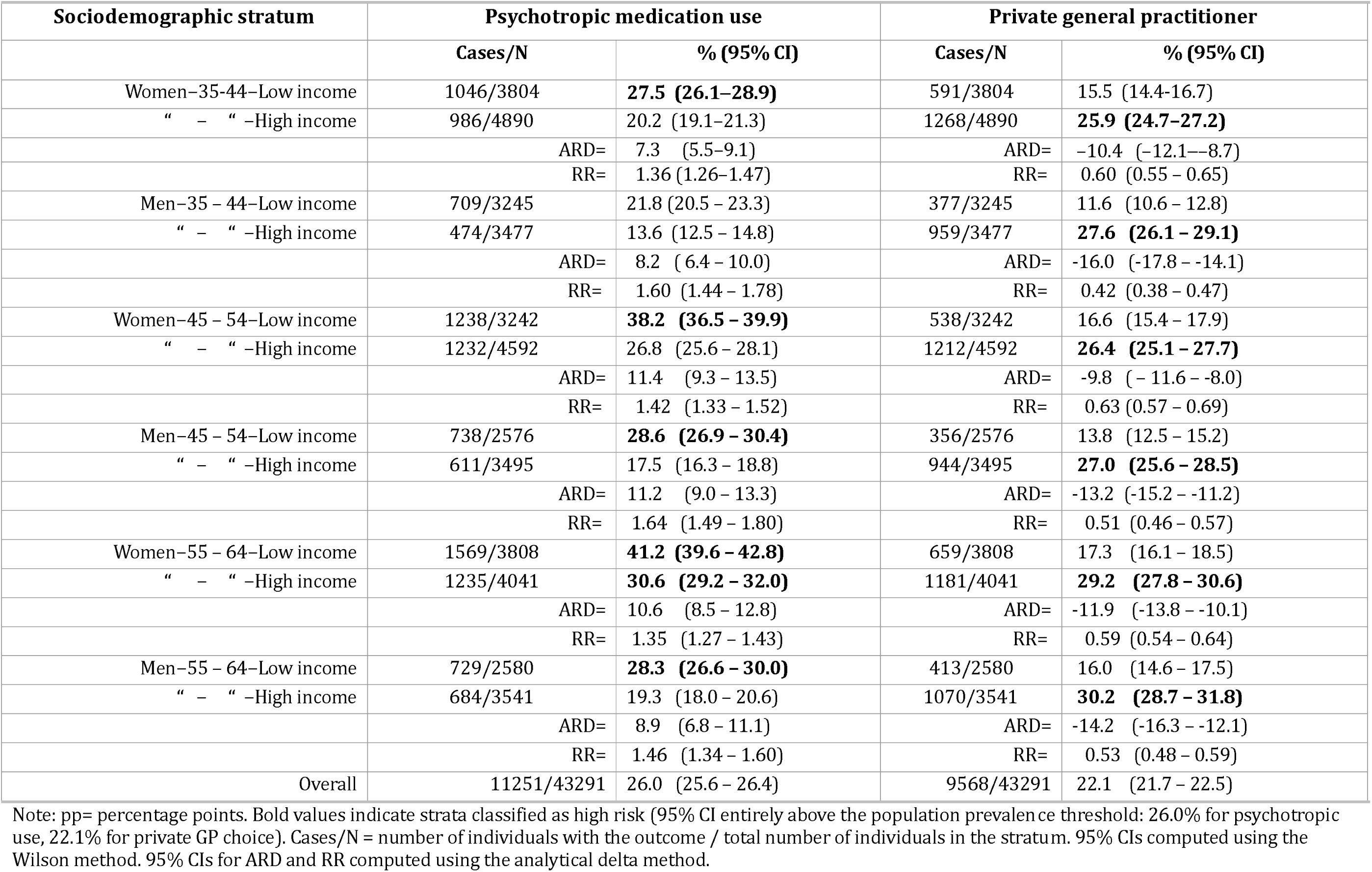
Crude prevalences (%) of psychotropic medication use and choice of a private general practitioner as well as Absolute Risk Differences (ARD) in percentage points and Relative Risks (RR) with 95% confidence intervals (CI) comparing low versus high income individuals of same sex and age group, Malmö, Sweden, 2006.

For psychotropic medication use, individuals in low-income strata have systematically higher prevalence than those in high income strata, with absolute risk differences ranging from 7.3 to 11.4 percentage points in women and from 8.2 to 11.2 percentage points in men. The pattern is reversed for private GP choice: individuals in high income strata use private GP more than those in low-income strata in every stratum, with absolute risk differences ranging from 9.8 to 11.9 percentage points in women and from 13.2 to 16.0 percentage points in men. Strata classified as high risk (95% CI entirely above the population prevalence threshold) are highlighted in bold and are revisited in the discriminatory accuracy analysis (Section 3.3).

#### 3.1.2 Geographical areas

Table 3 shows the crude area prevalences by area income group. For psychotropic medication use, area prevalences range from 22.0% to 30.4%.

**Table 3.** Crude prevalences of psychotropic medication use (Psy) and private general practitioner choice (Priv), with 95% confidence intervals (CI), by geographical area income. Malmö, Sweden, 2006.

| Area | Income | Psy n/N | Psy % (95% CI) | Priv n/N | Priv % (95% CI) |
| --- | --- | --- | --- | --- | --- |
| 3 | Poor | 702/2633 | 26.7 (25.0–28.4) | 428/2633 | 16.3 (14.9–17.7) |
| 4 | “ | 559/1994 | 28.0 (26.1–30.0) | 705/1994 | 35.4 (33.3–37.5) |
| 8 | “ | 412/1452 | 28.4 (26.1–30.7) | 336/1452 | 23.1 (21.0–25.4) |
| 9 | “ | 520/1957 | 26.6 (24.7–28.6) | 216/1957 | 11.0 (9.7–12.5) |
| 10 | “ | 594/2166 | 27.4 (25.6–29.3) | 280/2166 | 12.9 (11.6–14.4) |
| 12 | “ | 557/2161 | 25.8 (24.0–27.7) | 259/2161 | 12.0 (10.7–13.4) |
| 14 | “ | 605/2252 | 26.9 (25.1–28.7) | 416/2252 | 18.5 (16.9–20.1) |
| 16 | “ | 530/1989 | 26.6 (24.7–28.6) | 223/1989 | 11.2 (9.9–12.7) |
| 17 | “ | 628/2348 | 26.7 (25.0–28.6) | 538/2348 | 22.9 (21.3–24.7) |
| 20 | “ | 436/1796 | 24.3 (22.3–26.3) | 265/1796 | 14.8 (13.2–16.5) |
| 21 | “ | 560/1842 | 30.4 (28.3–32.5) | 152/1842 | 8.3 (7.1–9.6) |
| 23 | “ | 467/1691 | 27.6 (25.5–29.8) | 225/1691 | 13.3 (11.8–15.0) |
| 24 | “ | 365/1266 | 28.8 (26.4–31.4) | 589/1266 | 46.5 (43.8–49.3) |
| 1 | Rich | 296/1186 | 25.0 (22.6–27.5) | 211/1186 | 17.8 (15.7–20.1) |
| 2 | “ | 453/1772 | 25.6 (23.6–27.6) | 165/1772 | 9.3 (8.0–10.8) |
| 5 | “ | 425/1665 | 25.5 (23.5–27.7) | 486/1665 | 29.2 (27.1–31.4) |
| 6 | “ | 283/1266 | 22.4 (20.1–24.7) | 598/1266 | 47.2 (44.5–50.0) |
| 7 | “ | 367/1340 | 27.4 (25.1–29.8) | 291/1340 | 21.7 (19.6–24.0) |
| 11 | “ | 344/1521 | 22.6 (20.6–24.8) | 652/1521 | 42.9 (40.4–45.4) |
| 13 | “ | 444/1682 | 26.4 (24.3–28.6) | 480/1682 | 28.5 (26.4–30.7) |
| 15 | “ | 380/1652 | 23.0 (21.0–25.1) | 371/1652 | 22.5 (20.5–24.5) |
| 18 | “ | 436/1726 | 25.3 (23.3–27.4) | 396/1726 | 22.9 (21.0–25.0) |
| 19 | “ | 279/1217 | 22.9 (20.7–25.4) | 503/1217 | 41.3 (38.6–44.1) |
| 22 | “ | 296/1293 | 22.9 (20.7–25.3) | 228/1293 | 17.6 (15.7–19.8) |
| 25 | “ | 313/1424 | 22.0 (19.9–24.2) | 555/1424 | 39.0 (36.5–41.5) |
| Average rate Poor (n=13) |  |  | 27.25 (26.69–27.80) |  | 18.93 (18.45–19.41) |
| Average rate Rich (n=12) |  |  | 24.24 (23.60–24.87) |  | 28.33 (27.68–28.98) |
| ARD (Poor – Rich) % pp |  |  | +3.01 (2.16 to 3.85) |  | –9.40 (–10.21 to –8.59) |
| RR (Poor / Rich) |  |  | 1.124 (1.087–1.162) |  | 0.668 (0.646–0.691) |
| Overall |  | 11251/43291 | 26.0 (25.6–26.4) | 9568/43291 | 22.1 (21.7–22.5) |

Footnote: Areas are classified as poor (n=13) or rich (n=12) according to whether the proportion of low-income residents in the area falls above or at-or-below the median of that proportion across the 25 areas. The average rate is the unweighted mean of the crude area proportions within each income group. Per-area 95% CIs computed using the Wilson method. 95% CIs for the average rate use the standard error of a weighted sum of independent binomial proportions [72], computed with J. Ranstam's Confint spreadsheet, version 51.4. 95% CIs for the ARD and RR computed using the analytical delta method.

The crude area prevalence is 27.25% in poor areas and 24.24% in rich areas, yielding an ARD of 3.01 percentage points (95% CI 2.16 to 3.85) and a RR of 1.12. The crude values show that poor areas have only slightly higher psychotropic use than rich areas.

For private GP choice the picture is strikingly different. Area prevalences range from 8.3% to 47.2%, a difference of 38.9 percentage points between the extreme areas. That figure should be read together with the GCE and the DA, which are reported below. The crude area income difference is substantial: 18.93% in poor areas versus 28.33% in rich areas, yielding an ARD of −9.40 percentage points (95% CI −10.21 to −8.59) and a RR of 0.67. Rich areas have substantially higher private GP use than poor areas. This is consistent with the sociodemographic income differences in Table 2, where high-income strata use private GP more, and it reflects the geographical segregation of private healthcare provision in Malmö, where private practices are not uniformly distributed across the city.

Whether the strong area-level association reflects geography per se or simply the sociodemographic composition of areas requires knowing, within each area, the joint distribution of sociodemographic position and outcome across the strata. This is precisely what the three-level design provides, and it lives at the level of the cells. The analysis collapses the individual records to the 300 A-SD strata, each with its size and its observed proportion, and no measure reported here loses anything in the collapse. The EAR standardisation in Table 4 uses exactly this cell-level information: it reweights the stratum proportions to give equal representation to the 12 sociodemographic groups within each area. This is not an ecological analysis. An ecological analysis keeps only the area margins and cannot separate composition from geography. Here the full joint of area and sociodemographic position is retained in the cells, and that is what makes the separation possible. It is individual-level data reweighted for comparability. After EAR standardisation, the adjusted value, ARD-EAR, for psychotropic use is 2.14 percentage points (95% CI 1.27 to 3.02), confirming that the small crude difference was not driven by composition. For private GP choice, the adjusted ARD-EAR is −7.98 percentage points (95% CI −8.79 to −7.17), only modestly attenuated compared with the crude. The strong association between area income and private GP use is therefore not explained by sociodemographic composition; it reflects a genuine geographical contextual effect, plausibly anchored in the location of private healthcare infrastructure itself.

**Table 4.** EAR-standardised (Std) prevalences of psychotropic medication use (Psy) and private general practitioner (GP) choice (Priv), with 95% confidence intervals (CI), by geographical area income. Malmö, Sweden, 2006.

| Area | Income | Std % Psy (95% CI) | Std % Priv (95% CI) |
| --- | --- | --- | --- |
| 3 | Poor | 26.1 (24.5–27.8) | 16.3 (14.8–17.7) |
| 4 | “ | 27.9 (26.0–29.9) | 35.6 (33.5–37.7) |
| 8 | “ | 28.2 (25.9–30.5) | 23.5 (21.3–25.8) |
| 9 | “ | 26.2 (24.2–28.1) | 11.3 (9.9–12.8) |
| 10 | “ | 27.2 (25.3–29.0) | 12.6 (11.3–14.0) |
| 12 | “ | 25.6 (23.7–27.4) | 11.8 (10.5–13.2) |
| 14 | “ | 27.4 (25.6–29.3) | 18.1 (16.5–19.7) |
| 16 | “ | 26.0 (24.1–28.0) | 10.9 (9.5–12.2) |
| 17 | “ | 26.4 (24.6–28.2) | 22.8 (21.1–24.5) |
| 20 | “ | 24.4 (22.4–26.4) | 14.3 (12.7–15.9) |
| 21 | “ | 29.5 (27.4–31.5) | 8.7 (7.3–10.0) |
| 23 | “ | 26.4 (24.3–28.4) | 13.4 (11.7–15.0) |
| 24 | “ | 29.8 (27.3–32.3) | 44.6 (42.2–47.1) |
| 1 | Rich | 25.8 (23.2–28.5) | 20.4 (17.8–23.0) |
| 2 | “ | 25.4 (23.3–27.4) | 9.1 (7.7–10.4) |
| 5 | “ | 26.2 (23.9–28.4) | 24.9 (23.0–26.9) |
| 6 | “ | 23.4 (20.8–26.1) | 43.5 (40.6–46.5) |
| 7 | “ | 29.0 (26.5–31.5) | 21.9 (19.6–24.1) |
| 11 | “ | 23.4 (21.1–25.7) | 40.1 (37.5–42.7) |
| 13 | “ | 26.6 (24.4–28.8) | 28.2 (25.9–30.5) |
| 15 | “ | 23.3 (21.2–25.4) | 21.0 (19.1–22.9) |
| 18 | “ | 25.4 (23.3–27.5) | 20.6 (18.7–22.4) |
| 19 | “ | 22.9 (20.5–25.3) | 38.3 (35.6–41.0) |
| 22 | “ | 24.0 (21.5–26.4) | 15.9 (13.9–17.8) |
| 25 | “ | 23.1 (20.7–25.4) | 37.1 (34.5–39.7) |
| Overall EAR Poor areas (n=13) |  | 27.01 (26.46–27.56) | 18.77 (18.30–19.24) |
| Overall EAR Rich areas (n=12) |  | 24.87 (24.19–25.54) | 26.75 (26.09–27.41) |
| ARD-EAR (Poor – Rich) %pp |  | +2.14 (1.27 to 3.02) | –7.98 (–8.79 to –7.17) |
| RR-EAR (Poor / Rich) |  | 1.086 (1.050–1.124) | 0.702 (0.677–0.727) |
| Overall |  | 26.0 (25.6–26.4) | 22.1 (21.7–22.5) |

*Footnote: Sociodemographic standardised proportions are computed using the Equivalent Average Rate (EAR) method, giving equal weight to each of the 12 sociodemographic strata within each area. Each standardised proportion (Std %) represents the geographical risk of that area net of its sociodemographic composition. The overall area-income EAR rate is the unweighted mean of the standardised proportions within each area income group. The ARD-EAR (Poor – Rich) %pp is the Absolute Risk Difference between the EAR standardised risks expressed in percentage points (pp.)*

#### 3.1.3 Area-sociodemographic strata

**Fig 4** (panels a and b) shows the observed prevalences for all 300 A-SD strata grouped by sociodemographic stratum and coloured by area income. Each dot is a directly observed proportion from the individuals in that stratum in that area, with its 95% confidence interval. In each band the black tick marks the prevalence of the sociodemographic stratum in all areas pooled. If an area only shifted the level, its dots would sit at about the same distance from the tick in every band. A distance that changes from band to band suggests that the area also changes the sociodemographic differences, and Figs 5 to 7 turn this impression into measures.

**Fig 4a.**
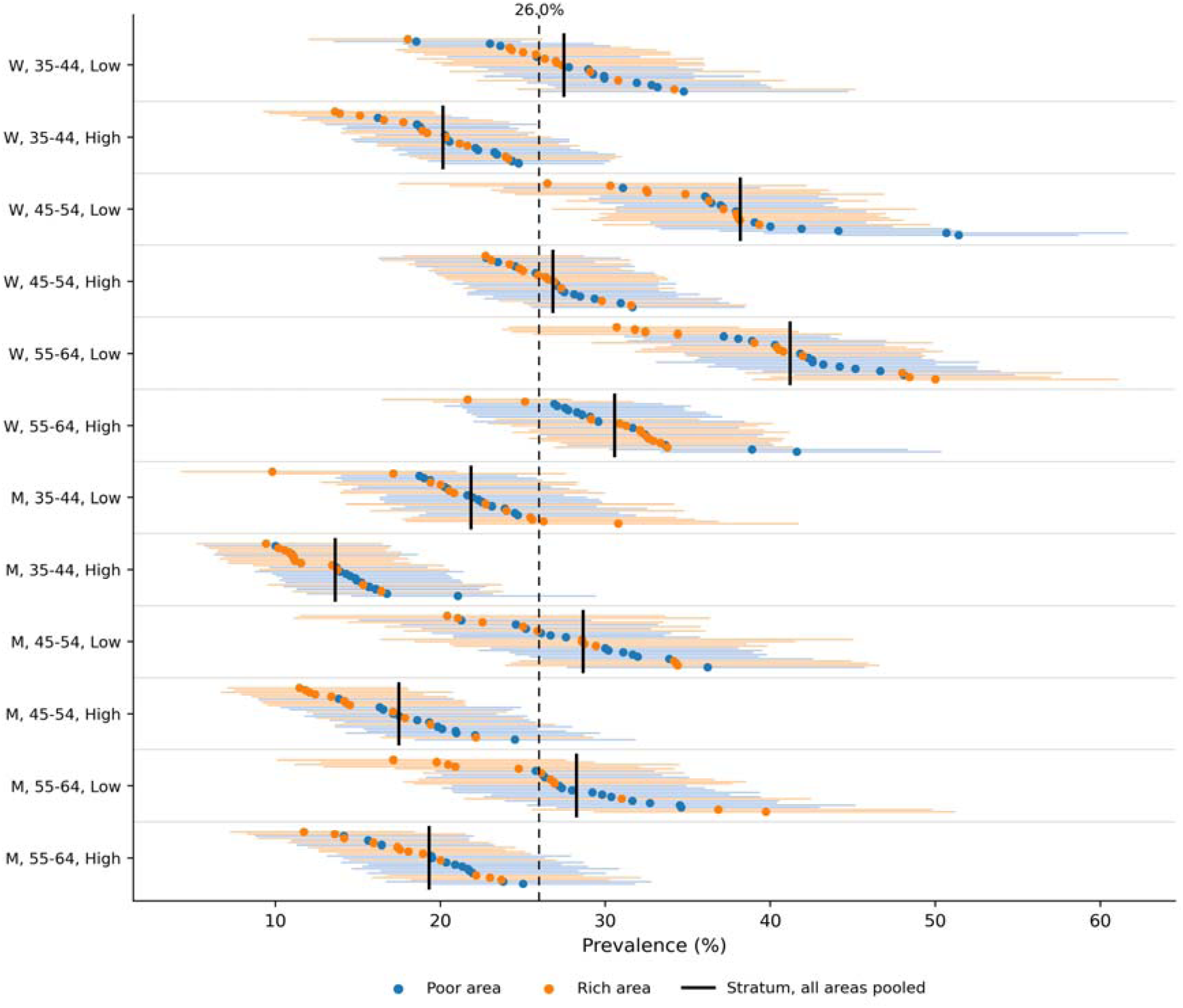
Prevalence of psychotropic medication use across the 300 area-sociodemographic strata. Malmö, Sweden, 2006. Each dot represents one of the 300 area-sociodemographic strata, showing the observed prevalence with its 95% confidence interval. Strata are grouped by sociodemographic stratum and sorted by prevalence within each group. Blue dots indicate poor areas, and orange dots indicate rich areas. The black tick marks the prevalence of the sociodemographic stratum in all areas pooled. The vertical dashed line marks the population prevalence threshold (26.0%). W = women; M = men; Low = low income; High = high income.

**Fig 4b.**
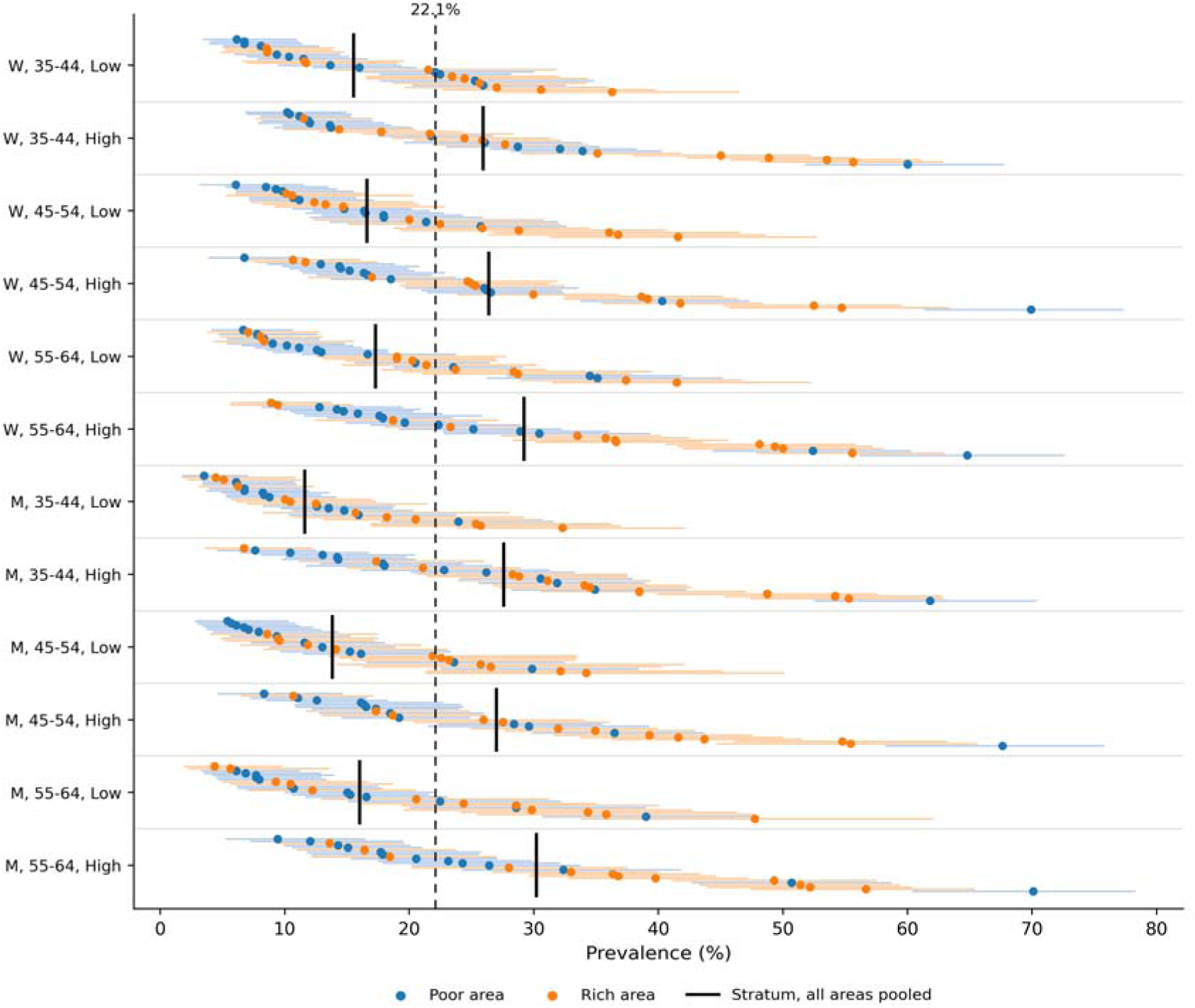
Prevalence of private general practitioner (GP) choice across the 300 area-sociodemographic strata. Malmö, Sweden, 2006. Each dot represents one of the 300 area-sociodemographic strata, showing the observed prevalence with its 95% confidence interval. Strata are grouped by sociodemographic stratum and sorted by prevalence within each group. Blue dots indicate poor areas, and orange dots indicate rich areas. The black tick marks the prevalence of the sociodemographic stratum in all areas pooled. The vertical dashed line marks the population prevalence threshold (22.1%). W = women; M = men; Low = low income; High = high income.

**Fig 5a.**
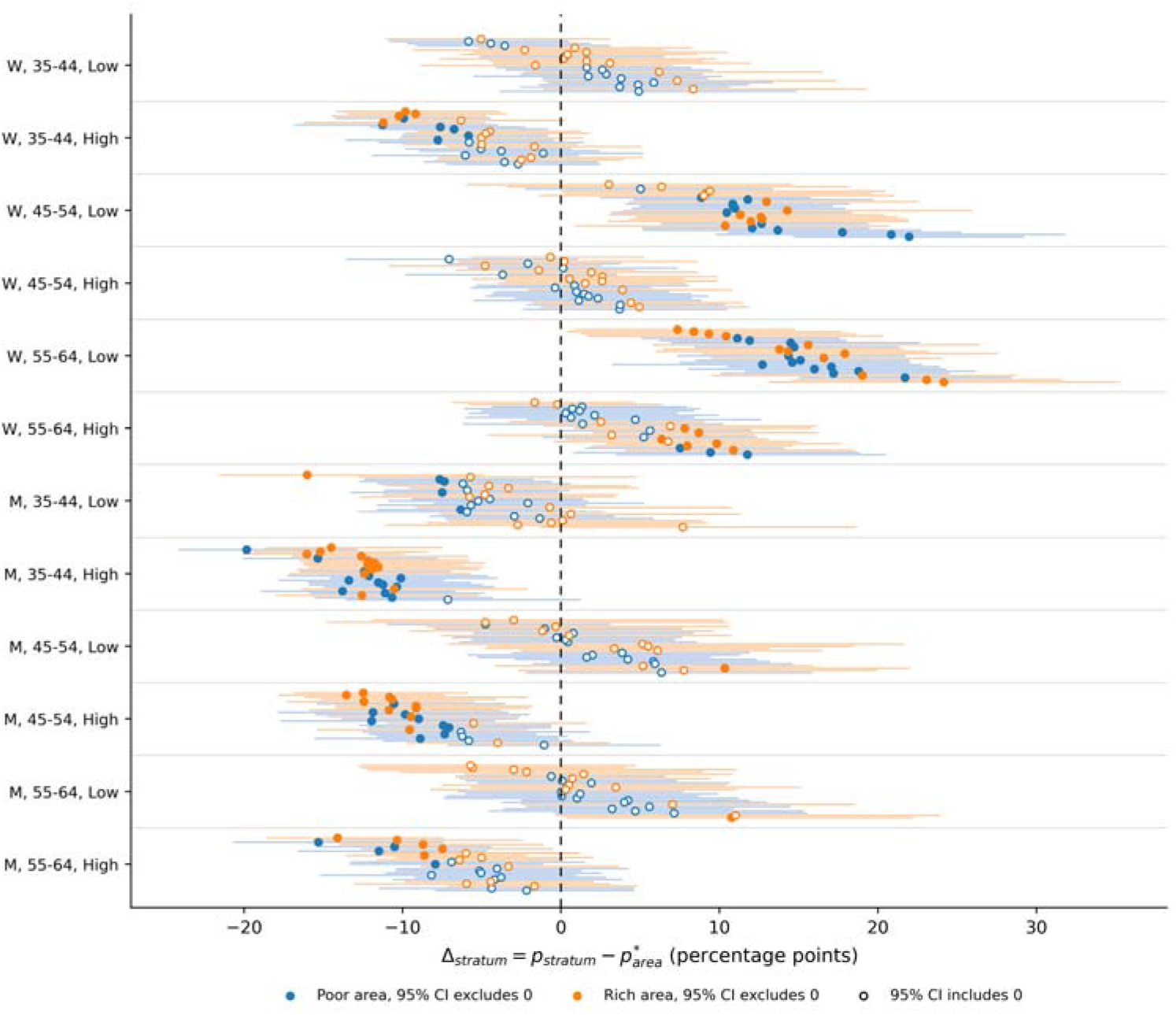
Deviation of area-sociodemographic stratum prevalence from the composition-standardised area prevalence (*Δ_stratum_*) for psychotropic medication use, by sociodemographic stratum. Malmö, Sweden, 2006. Each dot represents one of the 25 geographical areas for a given sociodemographic stratum. The x-axis shows the deviation of that A-SD stratum prevalence from its area’s composition-standardised prevalence (EAR, the plain mean of the 12 stratum proportions of the area) in percentage points. A positive value indicates higher prevalence than the area level; a negative value indicates lower. Blue dots indicate poor areas, and orange dots indicate rich areas. Horizontal lines show 95% confidence intervals. Solid dots indicate statistically meaningful deviations whose interval excludes zero; open dots indicate deviations whose interval includes zero. The vertical dashed line marks zero deviation. W = women; M = men; Low = low income; High = high income. Tables A to D of S2 Text give the deviations from the crude area prevalence.

**Fig 5b.**
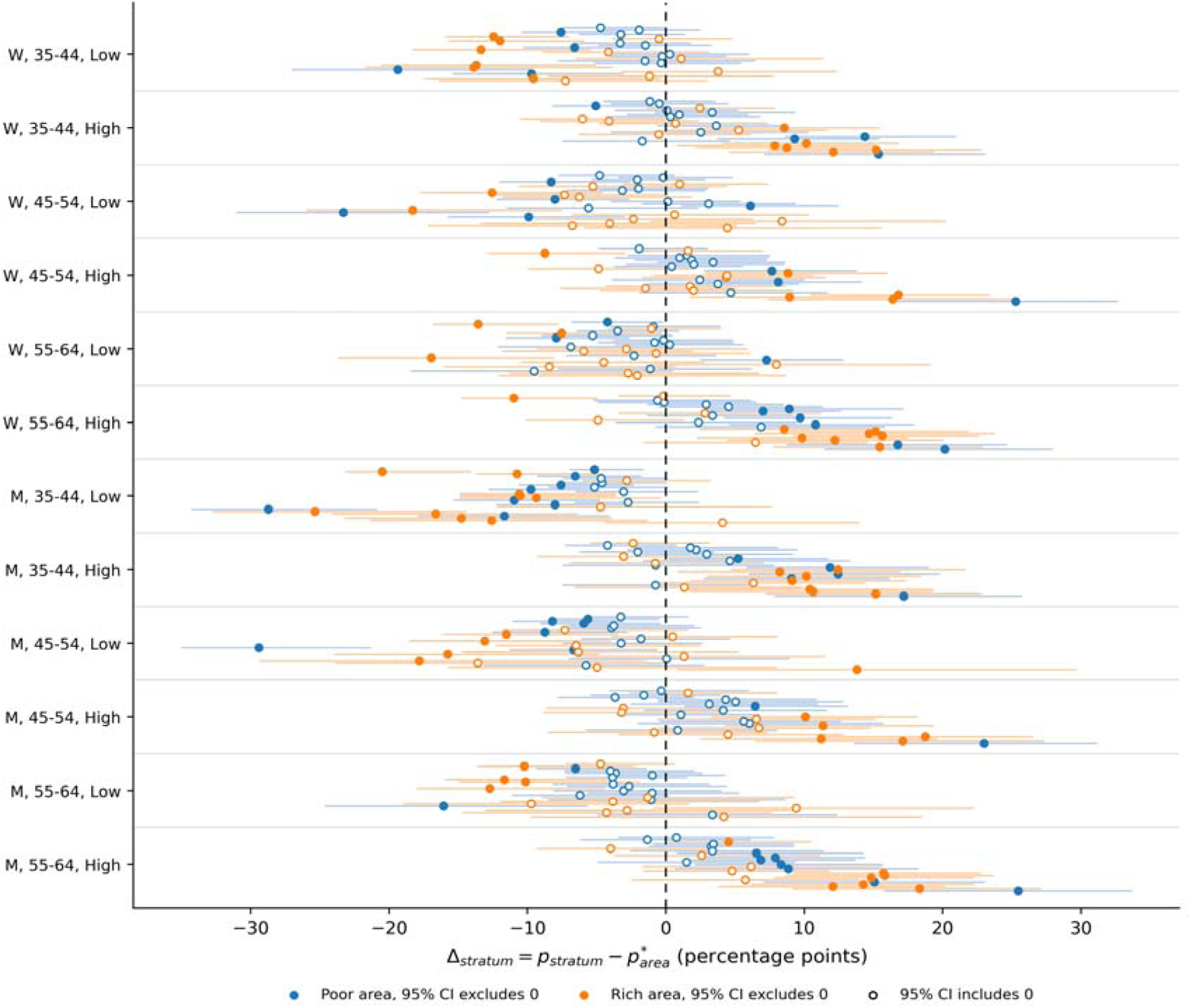
Deviation of area-sociodemographic stratum prevalence from the composition-standardised area prevalence (*Δ_stratum_*) for private general practitioner (GP) choice, by sociodemographic stratum. Malmö, Sweden, 2006. Each dot represents one of the 25 geographical areas for a given sociodemographic stratum. The x-axis shows the deviation of that A-SD stratum prevalence from its area’s composition-standardised prevalence (EAR, the plain mean of the 12 stratum proportions of the area) in percentage points. A positive value indicates higher prevalence than the area level; a negative value indicates lower. Blue dots indicate poor areas, and orange dots indicate rich areas. Horizontal lines show 95% confidence intervals. Solid dots indicate statistically meaningful deviations whose interval excludes zero; open dots indicate deviations whose interval includes zero. The vertical dashed line marks zero deviation. W = women; M = men; Low = low income; High = high income. Tables A to D of S2 Text give the deviations from the crude area prevalence.

**Fig 6a.**
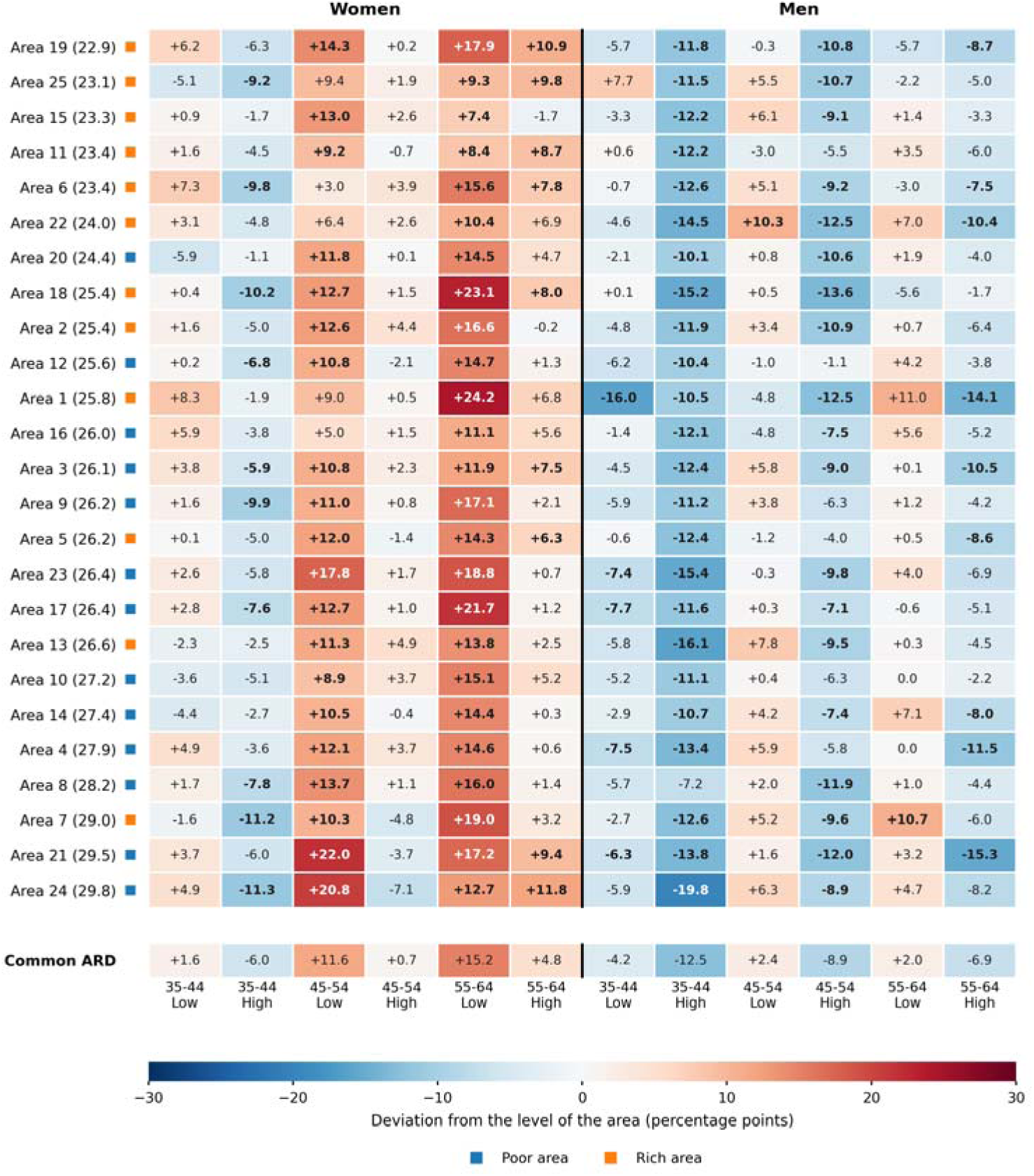
Deviation of each area-sociodemographic stratum from the level of its area, psychotropic medication use, in percentage points. Malmö, Sweden, 2006. The level of the area is its composition-standardised prevalence (in brackets), the plain mean of its 12 stratum proportions, so the deviations of every area average zero. Rows: the 25 areas, ordered by their level. Columns: the 12 sociodemographic strata, women to the left of the vertical line and men to the right. Bottom row: the common ARD of each sociodemographic stratum, the mean of its column over the 25 areas. Red, the stratum sits above the level of its area; blue, below it. Bold: the 95% confidence interval of the deviation excludes zero, as in Fig 5a. Blue square, poor area; orange square, rich area. Low = low income; High = high income.

**Fig 6b.**
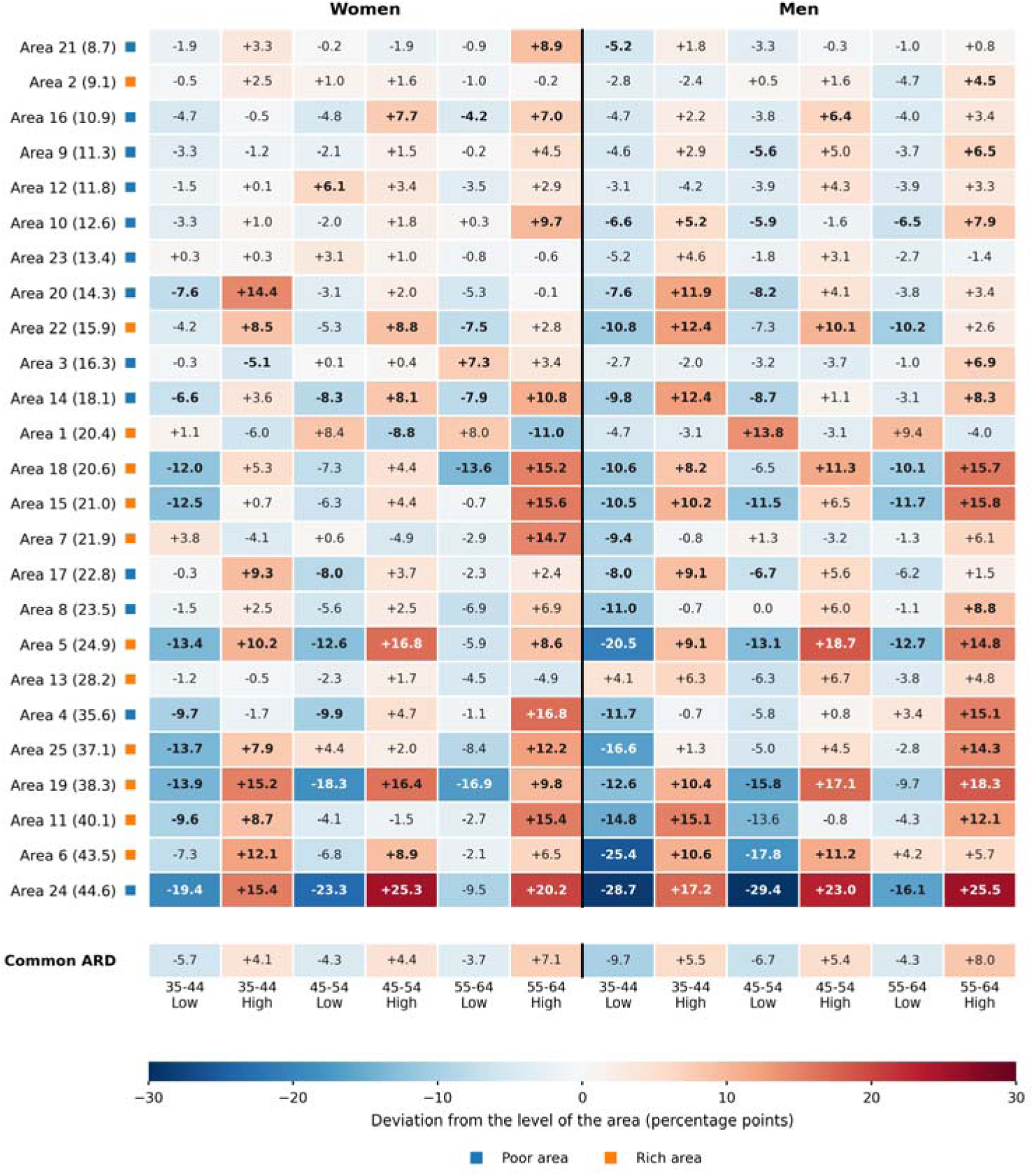
Deviation of each area-sociodemographic stratum from the level of its area, private general practitioner (GP) choice, in percentage points. Malmö, Sweden, 2006. The level of the area is its composition-standardised prevalence (in brackets), the plain mean of its 12 stratum proportions, so the deviations of every area average zero. Rows: the 25 areas, ordered by their level. Columns: the 12 sociodemographic strata, women to the left of the vertical line and men to the right. Bottom row: the common ARD of each sociodemographic stratum, the mean of its column over the 25 areas. Red, the stratum sits above the level of its area; blue, below it. Bold: the 95% confidence interval of the deviation excludes zero, as in Fig 5b. Blue square, poor area; orange square, rich area. Low = low income; High = high income.

**Fig 7a.**
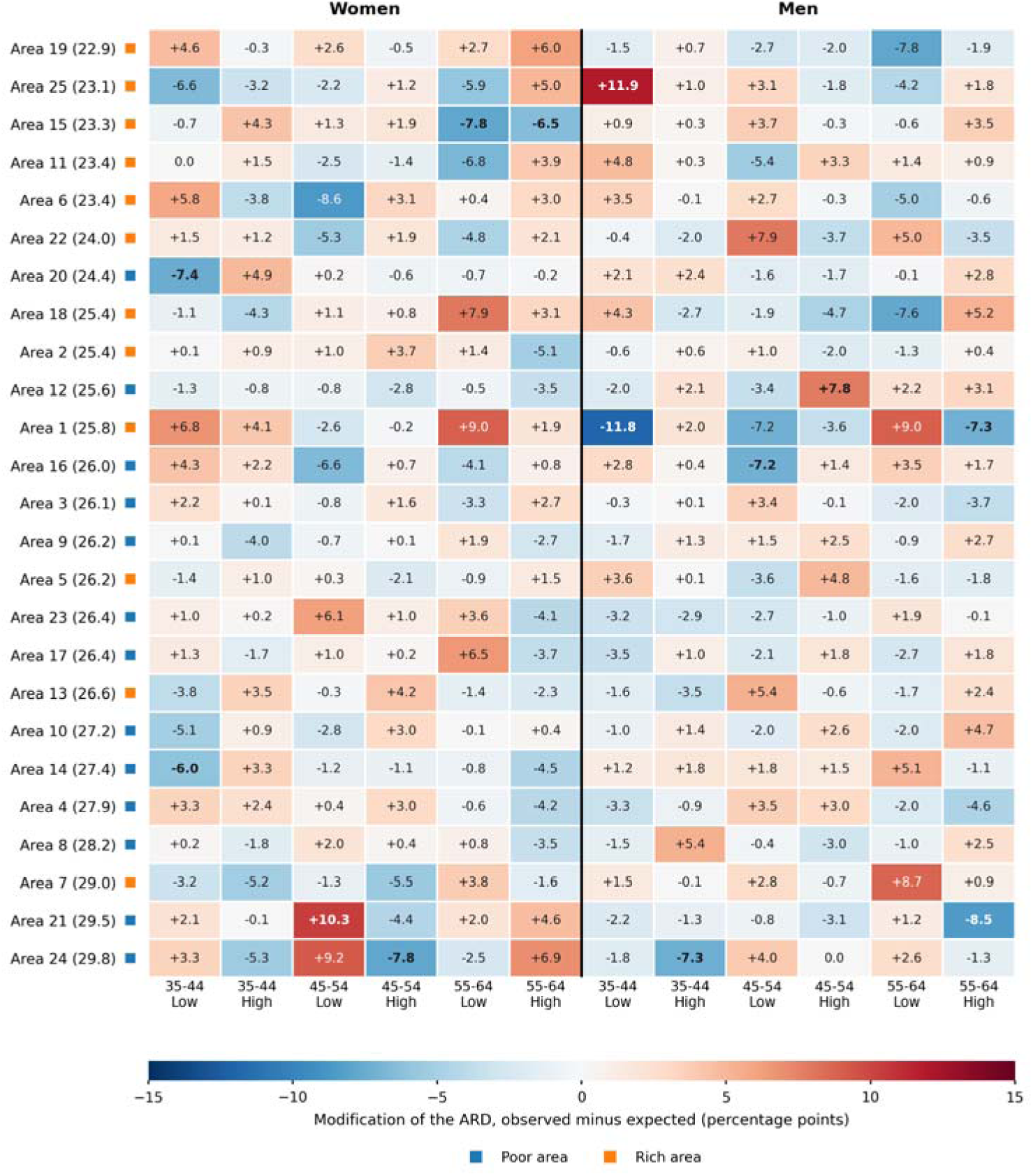
Modification of the sociodemographic differences by the area, psychotropic medication use, map of the 25 areas by the 12 sociodemographic strata. Malmö, Sweden, 2006. Each cell gives the modification of the ARD, γ, of one area-sociodemographic stratum in percentage points: the observed prevalence minus the prevalence expected if the area only shifted the level, which is the level of the area (in brackets) plus the common ARD of the stratum. It is the deviation of Fig 6a minus the common ARD of its column. Red, the stratum is above its expected prevalence; blue, below it. Where the common ARD is positive, red widens the common difference and blue narrows it; where it is negative, the reverse. The modifications sum to zero over the 12 strata of an area and over the 25 areas of a stratum. Bold: |γ| exceeds 1.96 binomial standard errors of the stratum (13 of 300 cells). Rows ordered by the level of the area. Blue square, poor area; orange square, rich area. Low = low income; High = high income.

**Fig 7b.**
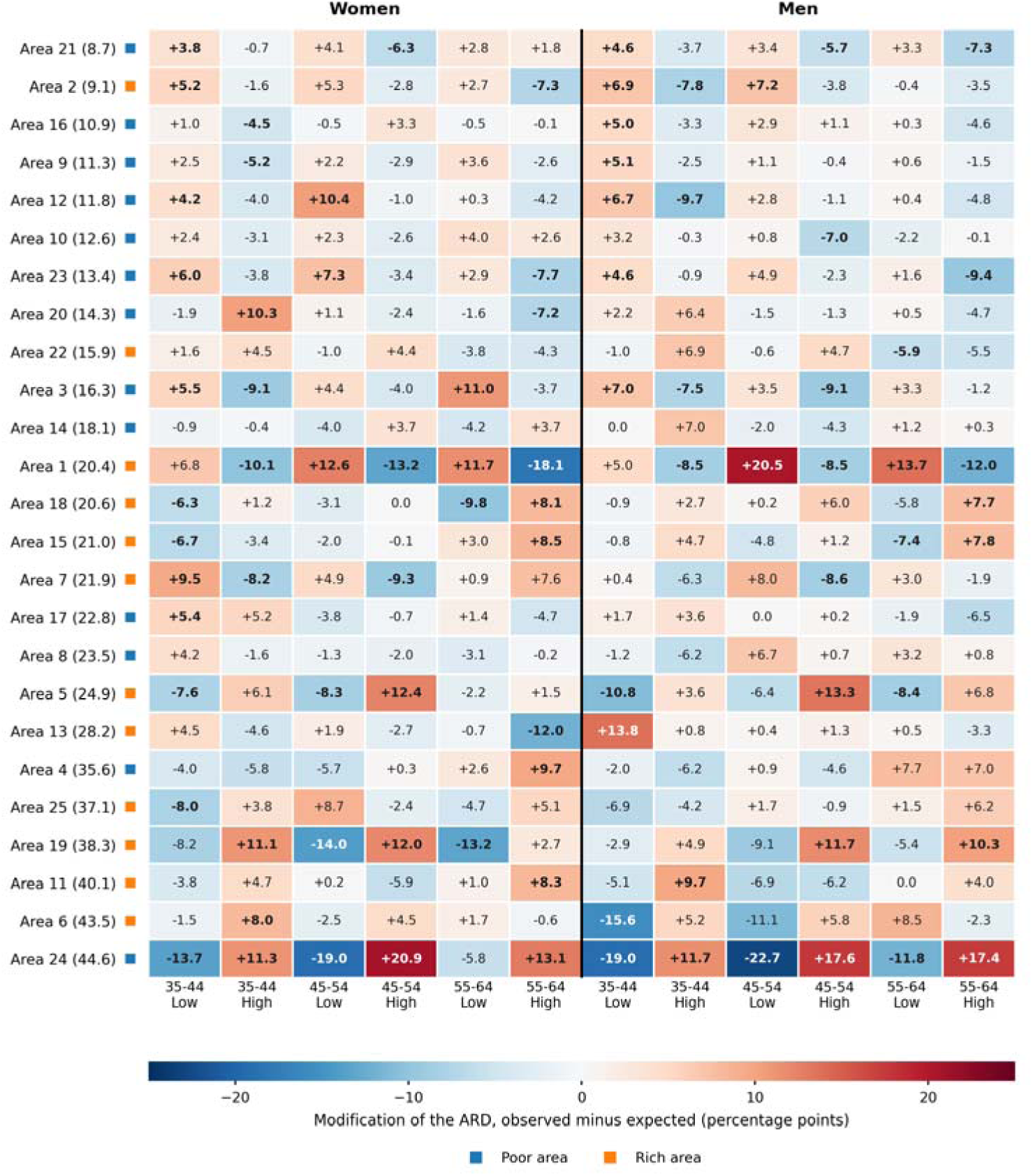
Modification of the sociodemographic differences by the area, private general practitioner (GP) choice, map of the 25 areas by the 12 sociodemographic strata. Malmö, Sweden, 2006. Each cell gives the modification of the ARD, γ, of one area-sociodemographic stratum in percentage points: the observed prevalence minus the prevalence expected if the area only shifted the level, which is the level of the area (in brackets) plus the common ARD of the stratum. It is the deviation of Fig 6b minus the common ARD of its column. Red, the stratum is above its expected prevalence; blue, below it. Where the common ARD is positive, red widens the common difference and blue narrows it; where it is negative, the reverse. The modifications sum to zero over the 12 strata of an area and over the 25 areas of a stratum. Bold: |γ| exceeds 1.96 binomial standard errors of the stratum (87 of 300 cells). Rows ordered by the level of the area. Blue square, poor area; orange square, rich area. Low = low income; High = high income.

**For psychotropic medication use,** four patterns are visible simultaneously. First, area income has only a modest influence on stratum-specific prevalence. Within each sociodemographic stratum group, orange dots (rich areas) and blue dots (poor areas) are largely interleaved, with blue dots tending to sit only slightly above orange dots. Whether a low-income woman aged 45 to 54 lives in a rich or a poor area makes only a small difference to her probability of using psychotropic medication.

Second, the sociodemographic income differences are consistent and geographically uniform. Low-income strata have systematically higher prevalence than high-income strata of the same sex and age, and these differences are similar in rich and poor areas alike.

Third, there is a geographical spread of approximately 16 percentage points within each stratum group, from the lowest to the highest area prevalence, reflecting between-area differences that are only weakly organised by area income.

Fourth, because of the consistent income differences, low-income strata more frequently exceed the population threshold of 26.0% and are classified as high risk.

**For private GP choice** the picture is strikingly different. The sociodemographic income differences are reversed: high-income strata have consistently higher prevalence than low-income strata of the same sex and age. Area income is also visible at the A-SD level. Within each sociodemographic stratum group the orange dots (rich areas) sit on average at higher prevalence than the blue dots (poor areas), and the gap is wider within the high-income strata, consistently across age groups and both sexes. This is in keeping with the concentration of private provision in the wealthier parts of the city. One area breaks the pattern, and the figure shows it plainly: the highest-prevalence dots in every high-income stratum belong to a single poor area whose high-income residents use private GP more than in any rich area, while its low-income residents fall well below the area average, giving it the largest internal income difference of all. A random-effects model would have pulled this area towards the overall mean and attenuated the contrast; the directly observed proportions keep it in full view.

**Fig 5** plots the deviation of each sociodemographic stratum from the level of its own area, its composition-standardised prevalence (Section 2.2.1) (*Δ_stratum_*), with one dot per area. Whereas Fig 4 shows the stratum prevalences against the population mean, Fig 5 removes the area level and shows only the within-area deviation of each stratum. For the same stratum, this deviation is larger in some areas and smaller in others, and it can even change sign, so the spread of the 25 dots within a stratum shows how strongly the area reshapes that stratum’s position. In the terms of Fig 3, the centre of a stratum’s 25 dots is its common ARD, δ, and their spread across areas is the modification of the ARD, γ. Because the level of each area is the plain mean of its 12 stratum proportions, the deviations of every area average zero, so the centre and the spread do not overlap (Section 2.2).

Two features are read directly from Fig 5. First, the deviations are centred on zero, the level of each area, and which side of zero a stratum falls on is shaped by its sociodemographic position, though the dominant dimension differs by outcome. For private GP choice (panel b) income governs the sign almost alone: high-income strata sit above their area average and low-income strata below it, while men and women differ negligibly. For psychotropic use (panel a) no single dimension dominates: sex, income and age shift the deviation by comparable amounts, with women, low-income and older strata upward, so the largest deviations combine all three, as in low-income women aged 55 to 64. Second, and more telling, the area dots are far more spread out within each stratum for private GP than for psychotropic use. For psychotropic use the 25 area deviations within a stratum span about 15 percentage points on average, widest in the low-income strata; for private GP choice the same dots span roughly twice as wide, about 29 points, so the same sociodemographic stratum can fall well below or well above its area average depending on the area. This spread is the visible signature of geographical modification of the sociodemographic differences, quantified by the contextual modification of inequalities (CMI, Section 2.2.2), the orange term of Fig 3. Area income accounts for only a small part of it: for psychotropic use the deviations are essentially the same in rich and poor areas, and for private GP choice the low-income strata sit on average 1.3 percentage points further below the level of their area in rich areas, and the high-income strata 1.3 points further above it, still small against the total spread. The modification is therefore not reducible to a simple contrast between rich and poor areas. It can be extreme: in one poor area, low-income strata aged 45 to 54 fall about 23 to 29 percentage points below the level of their area, while high-income strata of the same age rise about 23 to 25 points above it. Tables A to D of S2 Text give the deviations from the crude prevalence of each area, as printed by the smaihda command. Within an area they differ from those of Fig 5 by the composition gap of the area, on average 0.6 percentage points for psychotropic medication use and 1.2 for private GP choice, and at most 4.2. The ordering of the strata within an area is the same, but the crude level carries the composition of the area into the deviations: for private GP choice it doubles the apparent shortfall of the low-income strata in rich areas, from 1.3 to 2.8 points, and hides the excess of the high-income strata.

The four terms of Section 2.2.1 can be followed for one stratum, low-income men aged 35 to 44, and private GP choice. The reference is 22.6%, and the common ARD of the stratum is −9.7 percentage points: in the average area these men sit almost ten points below the level of their area. In area 18, a rich area with a level of 20.6%, the expected prevalence is 10.9%, and the observed 10.0% (11 of 110) departs from it by −0.9 points, so the area barely modifies the common difference. In area 24, the poor area with the highest level, 44.6%, the expected prevalence is 34.9% and the observed one 15.9% (18 of 113), a modification of −19.0 points that triples the common difference: 22.6 + 22.0 − 9.7 − 19.0 = 15.9%. In area 21, with a level of 8.7%, the expected prevalence is −1.0%, below what the scale allows. The observed 3.5% (7 of 199) gives a modification of +4.6 points that is partly the floor of the scale and not a narrowing of the difference by the area (Section 2.2.1).

Fig 6 draws the same deviations as a map of the 25 areas, ordered by their level, by the 12 sociodemographic strata, with the common ARD of each stratum in the bottom row. Read across a row, the deviations of an area average zero; read down a column, their mean is the common ARD. For psychotropic medication use the columns keep their colour from top to bottom: low-income women aged 45 to 64 sit above the level of every area, and high-income men aged 35 to 44 below it. For private GP choice the colours change down the columns, most of all in the areas with the highest levels, where the low-income strata fall far below the level of their area and the high-income strata rise far above it.

The detailed values for all 300 A-SD strata are provided as Tables A and B (Supplementary Information S2 Text).

### 3.2 General contextual effects

We report the two general contextual measures in the order introduced in the Methods: first the variance partition coefficient (VPC), then the Contextual Clustering of Inequalities (CCI).

#### 3.2.1 Variance partition coefficients

For psychotropic medication use the VPC_A_ = ICC_A_ is 0.22% (95% CI 0.13 to 0.32%), statistically above zero, but the areas organise very little of the individual heterogeneity. The purely sociodemographic component VPC_SD_ is 3.70% (95% CI 3.33 to 4.08%), so sociodemographic position accounts for only a small share of individual variation. The combined VPC**_SD+_**_A_ = ICC_SD+A_ is 3.92% (95% CI 3.56 to 4.30%). Almost all of this small contextual share is sociodemographic, and geography contributes next to nothing.

For private GP choice the pattern is strikingly different. VPC_A_ = ICC_A_ is 7.37% (95% CI 6.89 to 7.89%), a clearly geographical component with no parallel for psychotropic use. The VPC_SD_ is 3.80% (95% CI 3.43 to 4.18%), similar in magnitude to that observed for psychotropic use, indicating that sociodemographic position contributes comparably to individual variation in both outcomes. The combined VPC**_SD+_**_A_ = ICC_SD+A_ is 11.17% (95% CI 10.56 to 11.77%). The geographical component dominates the contextual structuring of private GP choice in a way that has no parallel for psychotropic use.

Table 5 also reports the standardised variance partition, VPC*, in which all strata are given the same size (Section 2.2.2). In the balanced Malmö design the two forms almost coincide. For psychotropic medication use the combined VPC_SD+A is 3.92% crude, 4.06% standardised without correction, and 3.30% after the binomial-noise correction. For private GP choice it is 11.17%, 11.44%, and 10.72%. The agreement is expected here, because each of the 25 areas contains the same 12 sociodemographic strata, so giving all strata the same size and counting individuals are almost the same operation. What the standardised version isolates is the strength of the contextual boundaries themselves, independent of how many individuals fall on each side. The two forms diverge when stratum sizes are unequal, the situation examined in the intersectional reanalysis in S4 Text.

**Table 5.** General contextual effects (GCE) estimated by crude and standardised variance partition coefficients (VPC and VPC*) and by the standardised contextual AUC (coAUC*) from observed proportions, for psychotropic medication use and private GP choice, Malmö 2006.All values are shown with 95% within-context bootstrap confidence intervals (B = 2,000, seed 2026), recentred on the estimate.

| Measure | Crude (95% CI) | Standardised, |  |
| --- | --- | --- | --- |
|  |  | uncorrected | Corrected |
| Panel A. Psychotropic medication use (p = 26.0%) |  |  |  |
| $VPC_A = ICC_A$ | 0.22% (0.13 to 0.32) | 0.20% (0.10 to 0.30) | 0.14% (0.04 to 0.24) |
| $VPC_{SD}$ | 3.70% (3.33 to 4.08) | 3.86% (3.46 to 4.25) | 3.16% (2.76 to 3.56) |
| $VPC_{SD} + A = ICC_{SD} + A$ | 3.92% (3.56 to 4.30) | 4.06% (3.65 to 4.47) | 3.30% (2.88 to 3.71) |
| coAUC*, A-SD strata (%) | See Table 7 | 62.97 (62.37 to 63.55) |  |
| coAUC*, area only,<br>standardised for size (%) | See Table 7 | 53.22 (52.60 to 53.85) |  |
| coAUC*EAR, area only,<br>standardised for size and<br>composition (%) | See Table 7 | 52.86 (52.21 to 53.51) |  |
| Panel B. Private GP choice (p = 22.1%) |  |  |  |
| $VPC_A = ICC_A$ | 7.37% (6.89 to 7.89) | 6.91% (6.39 to 7.48) | 6.86% (6.33 to 7.42) |
| $VPC_{SD}$ | 3.80% (3.43 to 4.18) | 4.53% (4.05 to 5.02) | 3.87% (3.39 to 4.37) |
| $VPC_{SD} + A = ICC_{SD} + A$ | 11.17% (10.56 to 11.77) | 11.44% (10.80 to 12.09) | 10.72% (10.08 to 11.38) |
| coAUC*, A-SD strata (%) | See Table 7 | 71.90 (71.34 to 72.46) |  |
| coAUC*, area only,<br>standardised for size (%) | See Table 7 | 68.38 (67.80 to 68.97) |  |
| coAUC*EAR, area only,<br>standardised for size and<br>composition (%) | See Table 7 | 67.46 (66.84 to 68.09) |  |

The binomial-noise correction has a larger effect than the standardisation. For private GP choice it accounts for most of the movement in the combined coefficient, from 11.44% to 10.72%. With much smaller strata the correction would dominate. It does not remove variance from the total. It reassigns the sampling component from the between-stratum levels to the individual level, preserving the exact identity (Section 2.2.2).

The VPC on the probability scale also reads as a difference of average risks (Section 2.2.2). The people who chose a private GP lived in A-SD strata whose prevalence was on average 30.8%, against 19.6% for the people who did not, a difference of 11.2 points, which is the crude VPC. For psychotropic medication use the two averages were 28.9% and 25.0%, a difference of 3.9 points. With all strata given the same size the differences are 11.4 and 4.1 points, the uncorrected VPC* of Table 5, and the correction for binomial noise lowers them to 10.7 and 3.3 points.

#### 3.2.2 How context structures inequality: clustering, additive differences, and modification

We decompose the between-stratum inequality into three shares that sum to 100%: the between-area clustering (CCI), the additive sociodemographic differences (G), and the geographical modification of those differences (CMI). All three are standardised; the CMI is reported noise-corrected, with 95% confidence intervals (Table 6). These three shares summarise over all strata the coloured terms of Fig 3, and can be read off Figs 4 and 5: the area level (Fig 4) is the clustering (CCI), while the position and the spread of the stratum deviations in Fig 5 are the additive differences (G) and their modification (CMI). Figs 6 and 7 map the same terms area by area.

**Table 6.** Contextual decomposition of the between-stratum inequality into clustering (CCI), additive differences (G) and modification (CMI), standardised, with the CMI noise-corrected, Malmö 2006. Shares of the total between-stratum variance, with 95% within-context bootstrap CIs. Corrected variance components are truncated at zero. A share at the boundary carries an interval truncated with it.

|  |  |  |
| --- | --- | --- |
| **Share** | **Psychotropic medication use** | **Private GP choice** |
| CCI, contextual clustering of inequalities (%) | 4.1 (1.9 to 6.6) | 63.9 (60.5 to 67.3) |
| G, additive (shared) sociodemographic differences (%) | 95.9 (90.7 to 100.0) | 19.0 (16.7 to 21.4) |
| CMI, contextual modification of inequalities (%) | 0.0 (0.0 to 5.0) | 17.1 (14.3 to 19.9) |
| Notes. $CCI = V_A^* / (V_A^* + V_{SD}^*)$, the share of the between-stratum variance that lies between areas. $G = V_G^* / (V_A^* + V_{SD}^*)$, with $V_G^*$ the mean of the squared common sociodemographic differences $\delta_{SD}$. $CMI = V_{CMI}^* / (V_A^* + V_{SD}^*)$, with $V_{CMI}^*$ the mean of the squared modifications $\gamma$, noise-corrected. $CCI + G + CMI = 1$ (Section 2.2.2). The three shares are computed with all strata given the same size, whatever their number of individuals, so composition does not enter them; an asterisk marks a quantity of that kind. The crude VPC, the coAUC and the classification measures are computed from the individual counts, so composition does enter them, and the coAUC asks how well the context separates people in the population as it is. The full notation is in S1 Text (Table A).<br>Fig 7 maps the modification of every stratum, the deviation of Fig 6 minus the common ARD of its column, which sums to zero across every row and down every column. For psychotropic medication use the map is pale, and 13 of the 300 cells exceed 1.96 standard errors. For private GP choice 87 do. In area 24, the poor area with the highest level, every low-income stratum lies below its expected prevalence, by 5.8 to 22.7 points, and every high-income stratum above it, by 11.3 to 20.9 points, so the area widens the income gap in private GP use. In several of the areas with the lowest levels the sign is reversed, mainly in the low-income strata aged 35 to 44, which lie above their expected prevalence. |  |  |

For psychotropic medication use the inequality is almost entirely additive sociodemographic differences: G is 95.9% (95% CI 90.7 to 100.0), while the CCI is only 4.1% (1.9 to 6.6) and the CMI is indistinguishable from zero (0.0%, 0.0 to 5.0). Geography neither clusters the inequality nor reshapes it; the same differences operate in every area.

For private GP choice geography acts twice over. The CCI is 63.9% (60.5 to 67.3): two thirds of the between-stratum inequality is a matter of which area a stratum sits in. And of the sociodemographic differences, nearly half is reshaped by the area: the CMI is 17.1% (14.3 to 19.9), comparable to the differences themselves (G 19.0%, 16.7 to 21.4). This is the quantitative counterpart of Fig 5: the income gap in private GP use is amplified in rich areas and dampened in poor ones.

The CCI is the clustering share, here in its standardised form; the decomposition retains it and adds the modification (CMI) that clustering alone cannot capture. The contrast between the two outcomes is sharp: psychotropic use is sociodemographic and geographically stable, whereas private GP choice is geographical both in level and in the shape of its sociodemographic differences.

The VPC and the CCI give a consistent picture: for psychotropic use virtually all contextual structuring is sociodemographic, while for private GP choice geography dominates. These results are obtained entirely from observed proportions without any multilevel model, and they are coherent, analytically transparent, and directly interpretable.

Figs 8a and 8b display the modification cell by cell. For psychotropic medication use the observed prevalences follow the additive expectation almost everywhere: 13 of the 300 cells, 4.3%, depart from it by more than 1.96 binomial standard errors, close to what sampling variation alone would produce. For private general practitioner choice 87 cells, 29.0%, do so, and the departures are concentrated in one sector of the ordering rather than scattered.

**Fig 8a.**
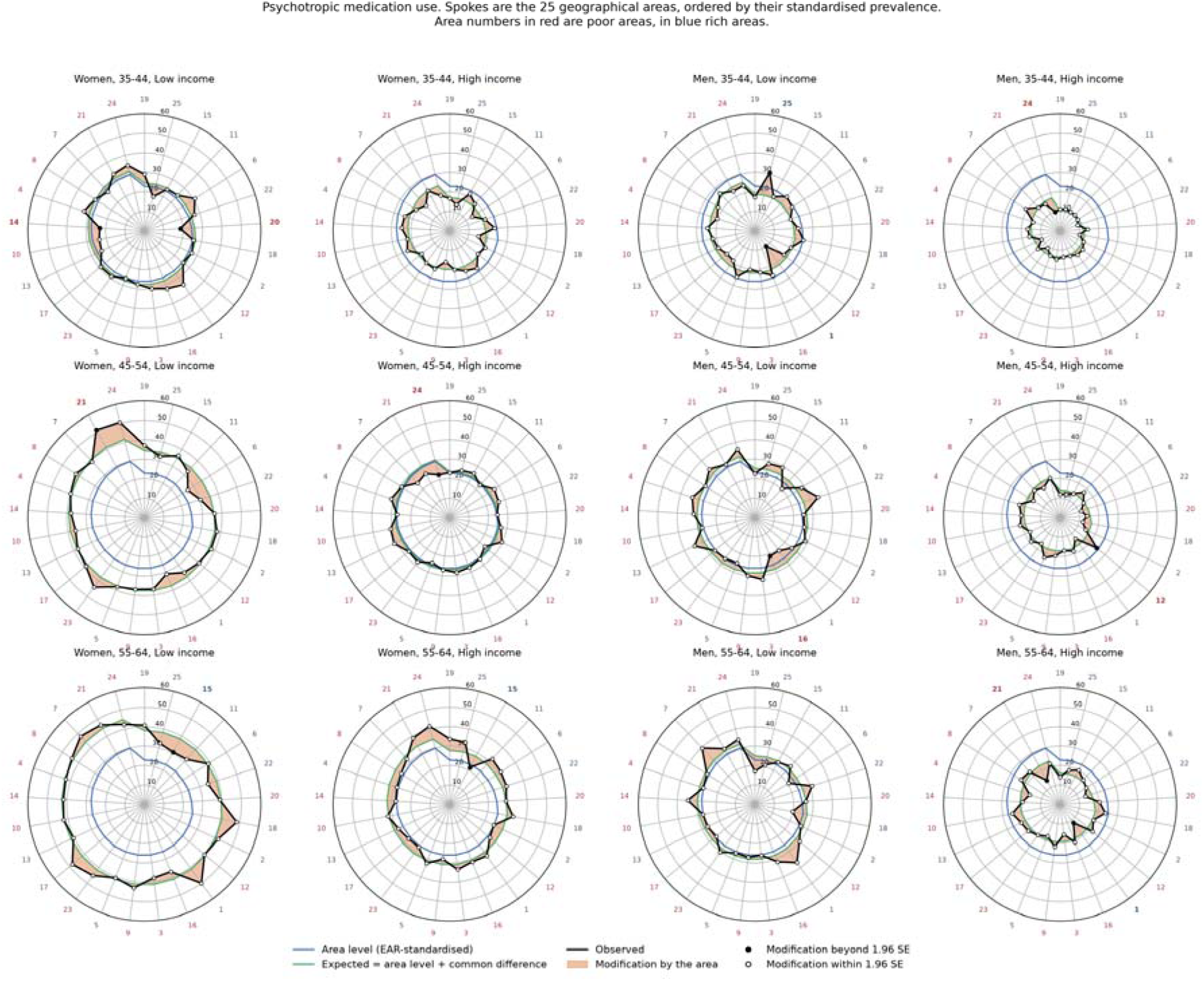
Modification of the sociodemographic differences by the area, psychotropic medication use. One panel per sociodemographic stratum. The 25 geographical areas are the spokes, ordered clockwise from the lowest to the highest standardised area prevalence; the order is the same in every panel. Blue: the standardised prevalence of the area, giving each of the 12 sociodemographic strata equal weight. Green: the prevalence expected under a purely additive representation, the area level plus the difference of that stratum common to all areas. Black: the observed prevalence. The orange area between the green and the black lines is the modification of the difference by the area. Filled markers and bold area numbers indicate cells whose modification exceeds 1.96 binomial standard errors; open markers and plain numbers, cells within that bound. Area numbers in red are poor areas, in blue rich areas. The areas are random groupings of the 218 original neighbourhoods, so the position of a spoke carries no geographical meaning. Of the 300 cells, 13 (4.3%) exceed the bound.

**Fig 8b.**
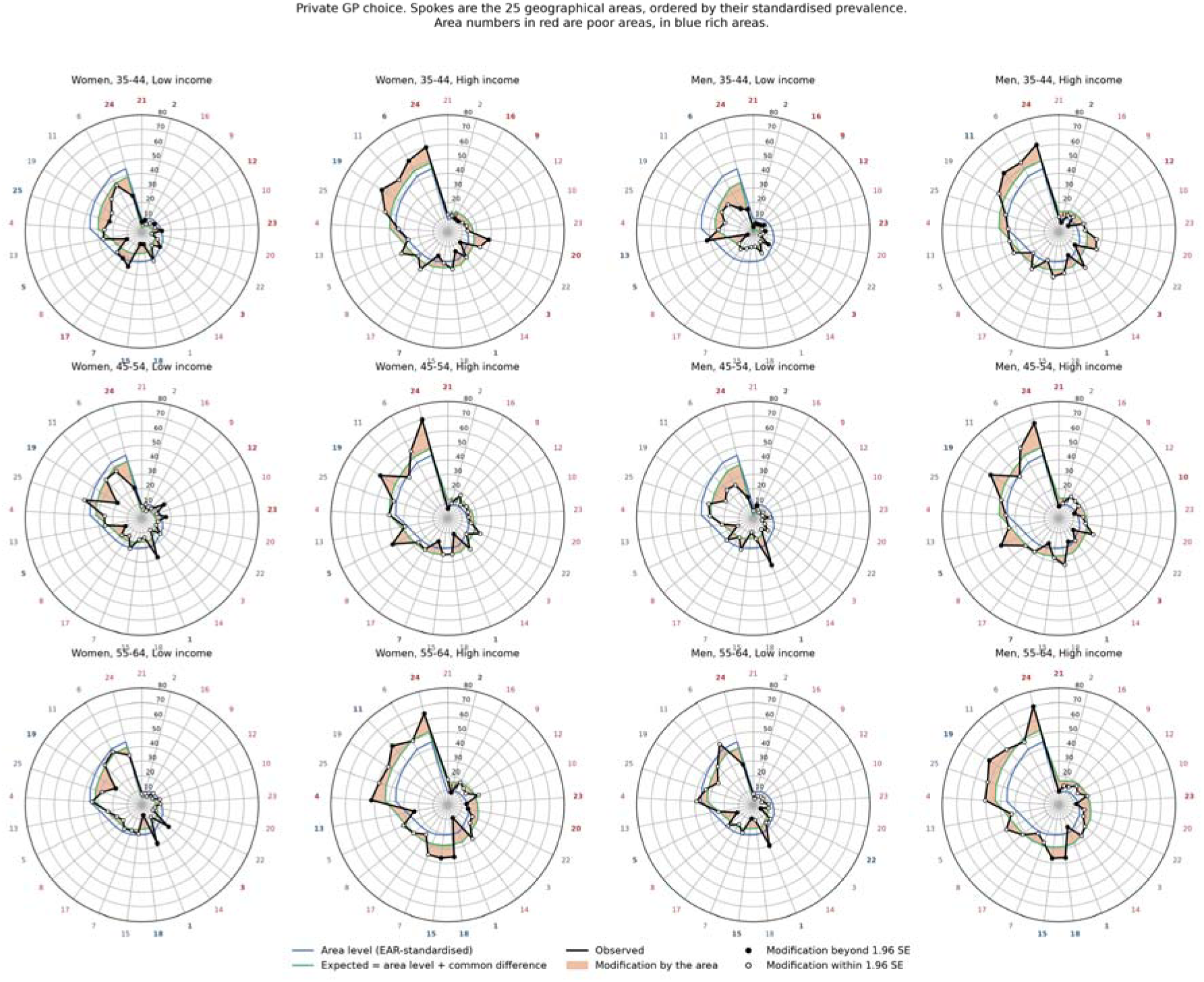
Modification of the sociodemographic differences by the area, private general practitioner choice. One panel per sociodemographic stratum. The 25 geographical areas are the spokes, ordered clockwise from the lowest to the highest standardised area prevalence; the order is the same in every panel. Blue: the standardised prevalence of the area, giving each of the 12 sociodemographic strata equal weight. Green: the prevalence expected under a purely additive representation, the area level plus the difference of that stratum common to all areas. Black: the observed prevalence. The orange area between the green and the black lines is the modification of the difference by the area. Filled markers and bold area numbers indicate cells whose modification exceeds 1.96 binomial standard errors; open markers and plain numbers, cells within that bound. Area numbers in red are poor areas, in blue rich areas. The areas are random groupings of the 218 original neighbourhoods, so the position of a spoke carries no geographical meaning. Of the 300 cells, 87 (29.0%) exceed the bound.

The floor of the probability scale barely touches these results. The expected prevalence under additivity was below zero in 2 of the 300 cells for private GP choice, which carry 0.5% of the modification variance, and in none for psychotropic medication use, and no expected prevalence exceeded 100%.

#### 3.2.3 The general contextual effect on the discrimination scale

The standardised contextual AUC, coAUC*, reads the same general contextual effect on the discrimination scale (Section 2.2.2). For psychotropic medication use it is 62.97% (95% CI 62.37 to 63.55) for the A-SD strata and 53.22% (52.60 to 53.85) for the areas alone. For private GP choice it is 71.90% (71.34 to 72.46) and 68.38% (67.80 to 68.97). The ordering of the two outcomes is the one given by VPC*, and Table 1 places the A-SD values in the small and in the moderate band respectively, and the area values in the absent and in the moderate band (Table 5). With the composition of the areas also standardised, coAUC*EAR was 52.86% (52.21 to 53.51) for psychotropic medication use and 67.46% (66.84 to 68.09) for private GP choice, in the same bands, so that the composition of the areas carried little of their general contextual effect.

Standardisation moves the contextual AUC by at most 0.5 percentage points in these data (Table 7). That agreement is itself informative. The 300 A-SD strata are close to uniform in size, with a mean and a median of 144 individuals and a range from 35 to 299, so counting individuals and giving all strata the same size give the same answer. The two readings diverge when the population is concentrated in a few contexts (S7 Text).

**Table 7.** Discriminatory accuracy measures for psychotropic medication use and private GP choice, Malmö 2006.

|  |  |  |
| --- | --- | --- |
| *Measure* | *Psychotropic medication* | *Private GP choice* |
| coAUC (A-SD strata) | 62.7% (95% CI: 62.2 to 63.3) | 71.8% (95% CI: 71.2 to 72.3) |
| coAUC (area level only) | 53.0% (95% CI: 52.4 to 53.5) | 67.9% (95% CI: 67.3 to 68.5) |
| Categorisation | Small | Moderate |
| Risk threshold | 26.0% | 22.1% |
| - True positives (TP) | 6,717 | 6,366 |
| - False positives (FP) | 13,352 | 11,583 |
| - False negatives (FN) | 4,534 | 3,202 |
| - True negatives (TN) | 18,688 | 22,140 |
| - Sensitivity | 59.7% | 66.5% |
| - Specificity | 58.3% | 65.7% |

The coAUC reported in Section 3.3 answers a different question. It counts individuals, so it describes how the burden of cases is in fact distributed across the contexts, which is what a classification at a threshold acts upon. coAUC* gives all strata the same size, so it describes how sharply the contextual boundaries separate individuals by risk, whatever the number of individuals on each side. The two are reported side by side in Table 5 and read together.

### 3.3 Discriminatory accuracy

Table 7 presents the discriminatory accuracy results for both outcomes including the contextual AUC and the threshold-based classification.

The coAUC of the A-SD strata is the probability that a randomly selected individual with the outcome belongs to an A-SD stratum with a higher observed proportion than a randomly selected individual without the outcome. For psychotropic use this probability is 62.7%, corresponding to small discriminatory accuracy. The area-level coAUC alone is 53.0%, barely above the 50% that would indicate complete absence of geographical discriminatory accuracy. Adding sociodemographic information to geography raises the coAUC by 9.7 percentage points, a meaningful contribution.

For private GP choice the coAUC at the A-SD level is 71.8%, corresponding to moderate discriminatory accuracy. The area-level coAUC alone is already 67.9%, which Table 1 places in the moderate band. Adding sociodemographic information raises it by only 3.9 percentage points, indicating that geography dominates the discriminatory accuracy for private GP choice.

The threshold-based classification makes Rose’s prevention paradox concrete and quantifiable. For psychotropic use, a targeted intervention directed only at high-risk A-SD strata would reach 6,717 cases (sensitivity 59.7%) but miss 4,534, representing 40.3% of all individuals with the outcome. At the same time 13,352 individuals without the outcome would be unnecessarily included. For private GP choice sensitivity is higher at 66.5%, but 3,202 cases (33.5%) would still be missed and 11,583 individuals without the outcome unnecessarily included. For both outcomes a large share of cases arises outside the designated high-risk strata, confirming that targeted strategies alone are insufficient and that universal approaches addressing the underlying sociodemographic and geographical determinants are needed alongside any targeted action.

It bears emphasising that the CCI, VPC and coAUC values reported here are conservative estimates of the true geographical contextual structuring. The 25 areas in this dataset were constructed by random grouping of the 218 original Malmö neighbourhoods, which substantially dissolves the true geographical structure of the city. In the original data with 218 meaningful SAMS the coAUC for private GP choice was approximately 90%[33]. Even with this degraded area structure, the simple proportions approach correctly identifies private GP choice as strongly geographically structured and psychotropic use as not.

Fig 9 shows the corresponding receiver operating characteristic curves. Within each panel the distance between the two curves is what the sociodemographic information adds to the area alone.

**Fig 9.**
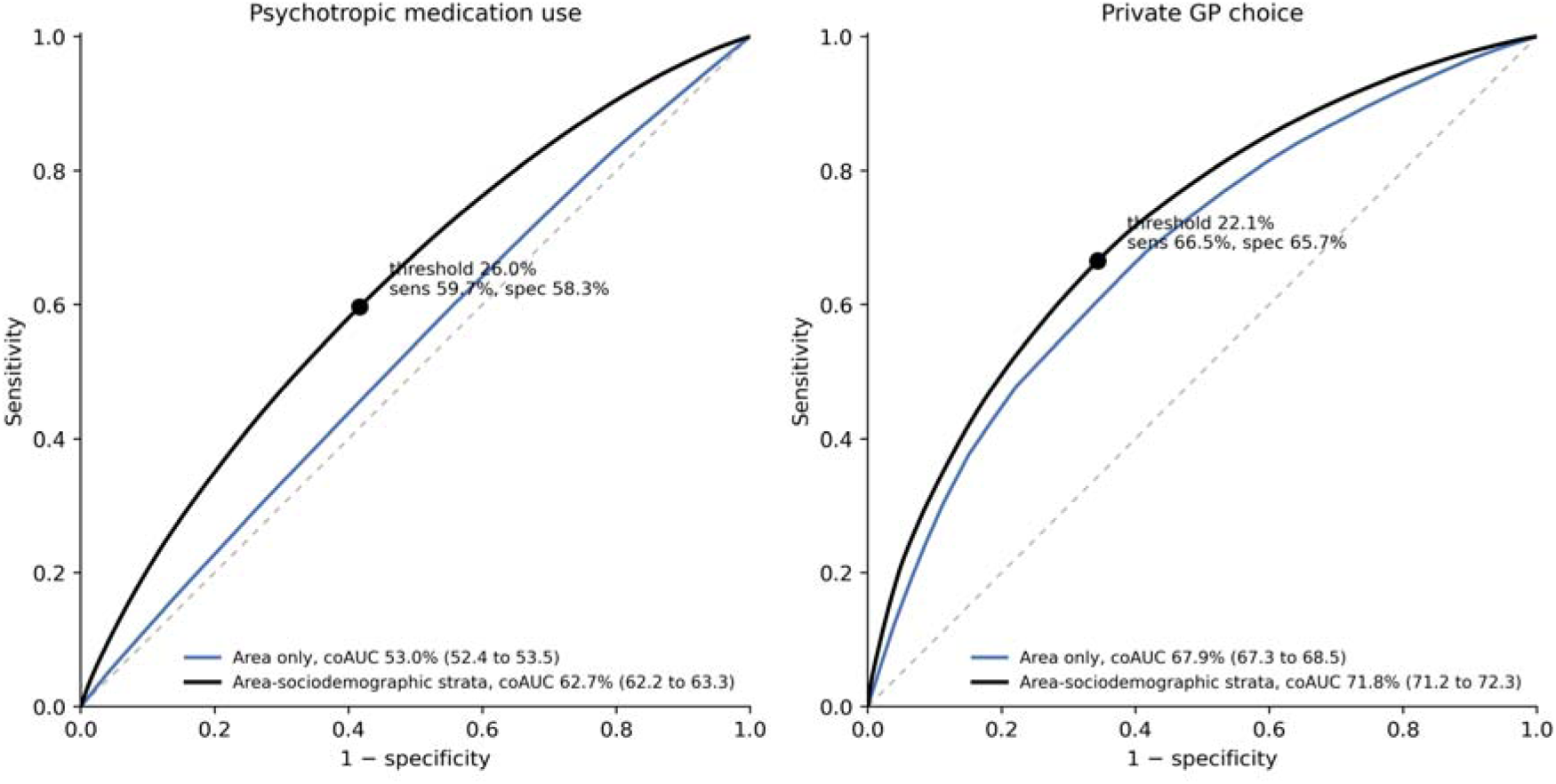
Discriminatory accuracy of the context. Receiver operating characteristic curves for the two outcomes. Black: individuals ranked by the observed prevalence of their area-sociodemographic stratum. Blue: ranked by the observed prevalence of their area alone. The area under each curve is given with its 95% confidence interval. The dashed diagonal marks the absence of discrimination. The black dot is the classification at the population prevalence taken as the threshold for a possible intervention, with its sensitivity and specificity. The curves join the vertices, so the area under the curve as plotted equals the reported coAUC.

### 3.4 Comparison between Simple-Means MAIHDA and Random-Effects MAIHDA

Table 8 presents the comparison between simple-means and RE-MAIHDA on the same A-SD strata using six of the nine measures proposed in the S1 Text. The pattern is consistent across both outcomes: in our study, the two methods agree closely on the summary-level measures (VPC and coAUC) but diverge at the per-stratum level.

**Table 8.** Comparison of simple-means (S) and random-effects (RE) MAIHDA on the same area-sociodemographic (A-SD) strata, for psychotropic medication use and private GP choice, Malmö 2006.

| *Measure* | *Psychotropic medication* | *Private GP choice* |
| --- | --- | --- |
| Range compression factor (RE / S, %) | 83.1 | 94.3 |
| Maximum absolute attenuation:<br>% points (stratum size) | 6.44 (n = 75) | 4.14 (n = 44) |
| VPC at A-SD on the probability scale, standardised, uncorrected (%): S / RE | 4.06 / 3.28 | 11.44 / 9.76 |
| VPC at A-SD on the probability scale, standardised, corrected for binomial noise (%): S / RE | 3.30 / 3.28 | 10.72 / 9.76 |
| coAUC at A-SD (%): S / RE | 62.7 / 62.7 | 71.8 / 71.8 |
| Sensitivity at the population-prevalence threshold (%): S / RE | 59.7 / 57.9 | 66.5 / 65.5 |
| Specificity at the population-prevalence threshold (%): S / RE | 58.3 / 60.1 | 65.7 / 66.6 |

Notes. S = Simple-Means MAIHDA; RE = Random-Effects MAIHDA. The RE estimates were obtained via *melogit* on the A-SD strata. The RE VPC at A-SD is placed on the probability scale by simulation from the fitted *melogit* (Method B); see S3 Text. Sensitivity and specificity for RE use the shrunken empirical-Bayes stratum predictions. The S estimates retain the 95% CIs reported in the tables of Sections 3.1, 3.2 and 3.3; the RE estimates are single point estimates without bootstrap CIs. Full technical details, together with the remaining three measures from the *compare* output in the *smaihda* Stata program, are provided in S1 Text. An explanation of *compare* is in S3 Text.

At the summary level the two methods agree closely. The comparison uses VPC* at the A-SD stratum level on the probability scale, in which all strata are given the same size, because that is how the random-effects τ² treats the strata (Section 2.2.4). Before the binomial-noise correction, simple means give 4.06% against 3.28% for random effects for psychotropic medication use, and 11.44% against 9.76% for private GP choice. The observed proportions retain the sampling noise that the model discounts, so the like-for-like comparison is the corrected form: 3.30% against 3.28%, and 10.72% against 9.76%. For psychotropic medication use the two values coincide. For private GP choice a difference of one percentage point remains, which reflects the distributional assumption of the random-effects model (S3 Text). The crude VPCs of Table 5 (3.92% and 11.17%) count every individual once and are not the comparator.

The random-effects model also gives the latent-scale quantities of the conventional multilevel logistic analysis, which read the same between-stratum variance in two ways. The median odds ratio (MOR) reads it as heterogeneity between the strata: the median of the odds ratios between two strata drawn at random, the higher-risk against the lower-risk one. The latent intraclass correlation reads it as clustering within the strata [2]. For the A-SD strata the between-stratum variance was 0.18 for psychotropic medication use and 0.64 for private GP choice, a MOR of 1.50 and 2.15 and a latent intraclass correlation of 5.2% and 16.4%. For the areas alone the MOR was 1.10 and 1.89 and the latent intraclass correlation 0.3% and 12.0% (lme4::glmer, adaptive quadrature with 7 points). The two outcomes keep the ordering given by VPC* and coAUC*. These quantities describe the normal distribution of strata that the model assumes, and they are reported for comparison with the literature; the simple-means analysis does not use them.

In our study S-MAIHDA and RE-MAIHDA capture the same overall contextual structuring, and the ranking of strata by predicted risk is preserved. Part of this agreement is structural. Because the coAUC is a rank-based summary that integrates over all thresholds, it is relatively insensitive to differential shrinkage: uniform shrinkage preserves the ordering and leaves the coAUC unchanged, while differential shrinkage alters it only by reordering strata. When group sizes differ widely, this reordering can in principle lower the random-effects coAUC. In our study, where the strata were more uniform in size, it did not (i.e., 300 A-SD strata, mean and median n = 144, range 35 to 299, with only four strata below 50 individuals). Threshold-specific measures such as sensitivity and specificity are more exposed: a single stratum crossing the classification threshold (the population prevalence in our study) shifts both, so classification can diverge between methods even when the coAUC agrees. In our data this divergence is small, because shrinkage moves only a few strata across the threshold (Table 8).

Fig 10 displays the comparison stratum by stratum. The upper panels plot the observed proportion against the random-effects prediction: the cloud departs from the identity line at both extremes and lies on it in the centre, which is the range compression reported above. The lower panels plot the displacement produced by the model against the distance of the stratum from the population prevalence. The displacement grows with that distance, with a correlation of 0.89 for psychotropic medication use and 0.83 for private general practitioner choice, whereas its correlation with the number of individuals in the stratum is −0.41 and −0.40. The largest displacement falls on a stratum of 75 individuals in the first outcome and of 44 in the second, and neither is among the smallest. The displacement is the distance from the mean multiplied by one minus the reliability weight, so it does not depend on stratum size alone.

**Fig 10.**
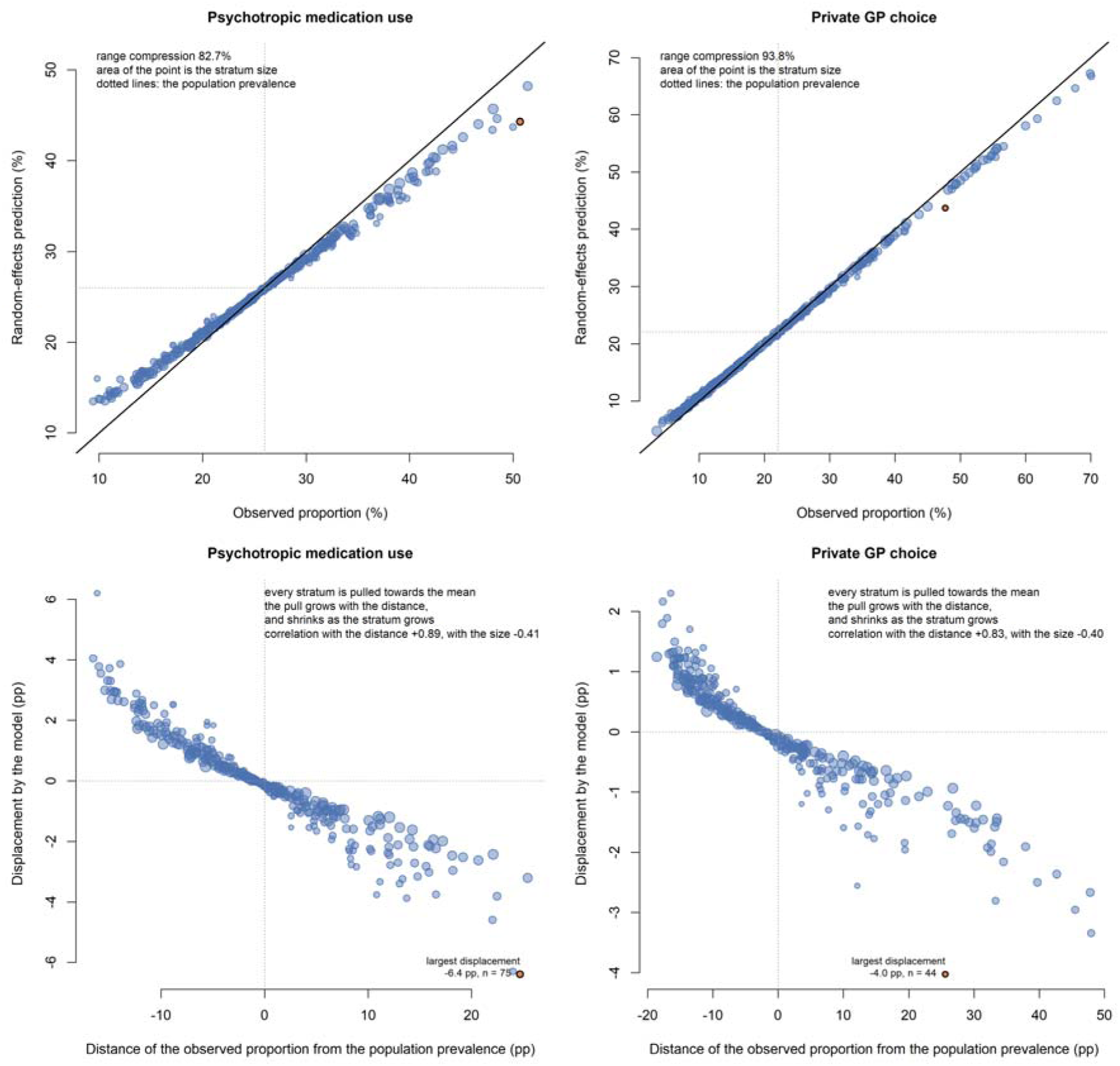
The compare diagnostic, stratum by stratum. Upper panels: the observed proportion of each of the 300 area-sociodemographic strata against the prediction of the random-effects model, with the identity line; the area of each point is proportional to the number of individuals in the stratum, and the dotted lines mark the population prevalence. Lower panels: the displacement produced by the model against the distance of the observed proportion from the population prevalence. The stratum with the largest displacement is marked in orange. The random-effects model was fitted with the glmer function of the lme4 package (S6 File); it agrees closely with the range compression factor and the maximum absolute attenuation of Table 8, including the size of the stratum at which that maximum falls.

## 4. Discussion

### 4.1 We found contrasting inequalities from unmodelled data

Using population data from Malmö and the three-level design, we showed that simple observed proportions deliver coherent, precise, and directly interpretable results for all three MAIHDA components.

The SCE reveal, for psychotropic medication use, consistent income differences across all sex and age groups that are stable between rich and poor areas. For private GP choice, the difference between rich and poor areas persists after EAR standardisation, so it is not explained by the sociodemographic composition of the areas as measured here. The pattern also differs across areas: the same income differences are larger in some areas and smaller in others. We describe these patterns rather than identify them causally, and the directly observed proportions display them without shrinkage.

The GCE put the contrast on two complementary footings. The crude VPCs of Table 5 express the share of total individual variation that lies between contexts: for psychotropic use only about 4% is contextual and almost all of it sociodemographic (3.7%, 0.2%), whereas for private GP choice about 11% is contextual and dominated by geography (7.4%, 3.8%). The CCI asks the complementary question, not how much of individual variation is contextual but how much of the contextual inequality is itself geographically organised: 4.1% for psychotropic use against 63.9% for private GP choice (Table 6). The VPC contextualises individuals, the CCI (geographically) contextualises the (sociodemographical) contexts, and both point the same way *VPC_SD_VPC_A_VPC_A_VPC_SD_*.

The DA show a sharp contrast between the two outcomes. Geography alone gives an area-level coAUC of 67.9% for private GP choice, which Table 1 places in the moderate band, against 53.0% for psychotropic use, barely above the 50% that indicates no discrimination at all. Being a rank-based summary, the coAUC agreed closely between simple means and random effects, while the threshold-based classification was more exposed. That classification also makes Rose’s prevention paradox concrete: for both outcomes a large share of cases falls outside the high-risk strata, so targeted strategies alone are insufficient. The three components are complementary and must be read together [52, 56, 57].

Table 5 also reports the standardised contextual AUC, coAUC*. In these data it barely differs from the coAUC, because the 300 A-SD strata are close to uniform in size. Its value is for comparison rather than for description. Two studies of the same outcome whose populations are differently distributed across the strata report a different coAUC for the same contextual structure, and the same coAUC*, so it is the coAUC* that can be carried from one study to another (S7 Text). Like any standardised measure it is read on a hypothetical population, here one in which the strata are of equal size. It also buys that comparability at a price in precision, since the smallest and noisiest strata carry full weight. Where the strata are of similar size, as in the present data, the price is nil and the two readings agree.

Throughout, no multilevel model was needed, no exchangeability assumption was imposed, and no shrinkage was applied. In these data RE-MAIHDA would have agreed closely, but the compare diagnostic and the worked example in S4 Text show that the agreement breaks down when strata are small, precisely the conditions of intersectional research. What was observed is what was reported.

When inequalities in health, or elsewhere, are studied from a social-epidemiological perspective, the contextually structured individual heterogeneity is the object of interest rather than noise, so shrinkage should be avoided. In simple-means MAIHDA, stratum size governs precision and nothing else. A small stratum yields a wide confidence interval that openly signals insufficient data. This matters epistemically, because absence of evidence is not evidence of absence. Reporting a sparse stratum as observed, with its full uncertainty, leaves the question honestly open; what shrinkage does instead is set out in Section 4.4.

Because they report the data as observed, simple means are the natural gold standard for descriptive inequality research; random-effects MAIHDA is acceptable when, and only when, it reproduces them, as the compare diagnostic confirms.

As we set out in the Methods (Section 2.2.2), the dependence of the proportion-scale variance partition on prevalence is not a defect. The Bernoulli variance p(1 − p) is a valid representation of reality, not an artifact and not a convention: it is the variance the data actually contain. The prevalence-independence of the latent scale is the convention, bought by fixing the individual variance at π²/3, a value that belongs to an assumed latent variable, not to the outcome. None of this is new in spirit. Variance-based measures of how an outcome is distributed across groups have a long history in the study of segregation and inequality, from the correlation ratio applied by Farley [73] to the Neighborhood Sorting Index of Jargowsky [74]; our contribution is to organise and apply these classical tools within the MAIHDA framework, not to invent a new measure.

Although illustrated with binary outcomes, the simple-means design is analogous for continuous outcomes: specific contextual effects become differences between observed stratum means, and the variance partition and the CCI are obtained directly on the natural scale, from a table with the size, the mean and the standard deviation of each stratum. For the discriminatory accuracy the natural extension is a rank-based contextual concordance, in the family of Harrell’s c-index and Somers’ D, of which the AUC is the binary special case; we develop this extension in forthcoming work.

### 4.2 Three perspectives in favour of simple means

Before setting out three perspectives in favour of simple means, a word on scale. Every quantity reported here is on the probability scale, and every comparison is a difference in percentage points. The decomposition is additive throughout: a stratum proportion is the population proportion plus two deviations. This is a deliberate choice rather than a technical convenience. Health inequality is experienced, counted and acted upon as a difference in risk: how many more people are affected in one stratum than in another. Quantities on the log-odds scale answer a different question, and they do not translate into that count without further assumptions. We therefore avoid transformations and pooling operations that move the reported quantities off the additive scale on which the inequality is defined. The same reasoning governs the comparison of deviations between areas: whether the sociodemographic differences are the same everywhere is a question about risk differences, and we answer it in those units.

From an epistemological perspective, accurate measurement of health inequalities is a precondition for equitable public health action. Take, for example, a low-income older woman living in a deprived area: RE-MAIHDA pulls her stratum’s estimate towards the population average within a hypothetical common distribution, while S-MAIHDA describes her stratum’s risk as the data show it. The present paper demonstrates empirically that simple means can do everything that RE-MAIHDA does for descriptive inequality research without that distortion.

From a pragmatic perspective, simple means are accessible, transparent, and reproducible without specialist multilevel software. The complete analytical pipeline is delivered as a documented Stata command and as free Python and R scripts, together with the stratum table, allowing any researcher to replicate every result and extend the approach.

From an empirical perspective, the Malmö illustration uses an artificially constructed area structure that substantially dissolves the true geographical signal. This is a conservative test of the method (Section 3.3). Even so, simple means correctly identify private GP choice as strongly geographically structured and psychotropic use as not. The example was designed for teaching and illustration, not for causal inference. Its purpose was to ask if the method works. It does.

A practical consequence follows from working with observed proportions. Every measure reported here is computed from two numbers per stratum: the number of individuals and the number with the outcome. For each outcome, the 43,291 individual records collapse to 300 rows, one per A-SD stratum, and every quantity in this paper can be recovered from that table (Section 2.2). Nothing that the analysis uses is lost in the collapse, and the individual records of each outcome can be recovered from the table.

This makes the approach portable. S-MAIHDA can be applied to results that have already been published, provided the source reports stratum sizes together with stratum-specific proportions or counts. S4 Text does exactly that, reanalysing published data on prescription opioid misuse without access to the original records. The portability is not exclusive to simple means, since a random-effects model can also be fitted from stratum counts in binomial form, but S-MAIHDA needs nothing beyond the counts themselves and fits no model to them.

The collapsed form offers no additional confidentiality. Within a stratum the individual records are indistinguishable except for the outcome, so the two forms present the same disclosure risk. What governs that risk is the granularity of the strata, not the format of the file. What the table changes is what has to leave the data holder. Only the counts that the analysis needs are shared, not the individual records. As ethics committees tighten the conditions for access to individual records, this is a practical solution.

The same property makes cross-national comparison tractable. Each country can compute the stratum table from its own records, under its own legal basis, and only the table needs to be shared. What must be agreed in advance is the definition of the strata, the definition of the outcome and the period, and the granularity of the strata is bounded by the disclosure rules of the participating registers.

### 4.3 Relation to previous work and what we add

Concern about random-effects shrinkage in RE-MAIHDA has a long history. Bingenheimer and Raudenbush [58] set out the mechanism two decades ago, in the context of health care provider performance. The empirical Bayes estimator is pulled away from the unbiased cluster mean by a fraction of its distance from the grand mean, and that fraction depends on the cluster size. The displacement is therefore governed by the size and the distance together, and it varies across clusters whenever the sizes differ. With equal sizes the rank order is preserved. With unequal sizes the ranks are reordered, and small providers that perform well are penalised while small providers that perform poorly are rewarded. The same mechanism operates on sociodemographic strata, where the most attenuated stratum is not the smallest one but one of limited size lying far from the average (Section 4.4). Lizotte and colleagues [75] returned to the issue in the context of intersectional MAIHDA, framed as an interpretive problem about how stratum residuals should be read against the fixed-effect baseline of a hypothetical equally sized population. Their proposed alternative was single-level regression with penalisation for high-dimensional intersectional designs. However, Lizotte’s concern was interpretive, not about whether shrinkage attenuates the description of inequalities. In a response, Evans, Leckie and Merlo [76] clarified that MAIHDA produces precision-weighted grand means and defended this property as an eclectic compromise between population means and unweighted grand means. However, the underestimation of inequalities was then not discussed.

Leckie and co-authors [77] subsequently revisited the issue with detailed simulations of intersectional RE-MAIHDA. They showed that the practical impact of shrinkage depends strongly on stratum size. When strata are reasonably large, random-effects predictions remain close to observed proportions, and the statistical advantages of RE-MAIHDA (precision, parsimony, protection against false positives) are preserved. The implication of their analysis is that the shrinkage critique is most consequential when stratum sizes are small, which is precisely the situation in which intersectional MAIHDA is most often applied. Bashir [59] articulated this concern epistemically in the context of intersectional RE-MAIHDA. His core argument is that the foundational assumption of RE-MAIHDA, that strata are exchangeable realisations of a common distribution, is incompatible with the conceptual premise of inequality research, because the groups whose differences are studied are structurally distinct entities, not interchangeable instances of a common process. In this line, several previous empirical studies under the MAIHDA framing dropped random effects in favour of fixed-effects regression and stratified simple-proportion analyses [53, 54, 60–65, 78], implicitly adopting the epistemic position later articulated by Bashir [59].

This paper adds four contributions. First, it provides a complete empirical demonstration that simple observed proportions are sufficient to operationalise all three MAIHDA components: SCE, GCE, and DA. Second, it introduces the contextual decomposition of the between-stratum inequality into clustering (CCI), additive differences (G) and modification (CMI), with the CMI corrected for binomial noise. Third, the compare option of the smaihda command makes the shrinkage trade-off identifiable in any dataset, turning a long-standing theoretical critique into a routine empirical check that should always be present in RE-MAIHDA studies. Nevertheless, visual inspection of the S-MAIHDA and RE-MAIHDA predictions is valuable too, and Fig 10 provides it: it shows both faces of shrinkage at once, the point pulled towards the mean and the pull growing with the distance from it. Fourth, the whole analysis works from a stratum table that holds only the number of individuals and the number of cases in each stratum, so only that table needs to leave the data holder. The pipeline is delivered in Stata, Python and R together with the table.

Leckie and colleagues [77] set out the statistical case for Random-Effects MAIHDA. With analytic expressions and a worked example, they show that the model’s predicted means recover the true population mean of each intersection more accurately than simple means, judged by mean squared error. This holds most clearly for the version that keeps the additive effects of the identities (their Model 2). The model accepts a small bias towards the additive pattern in return for a large drop in sampling noise, and the benefit is greatest for the small strata, where simple means are least stable. We accept this result. It is correct for its question, the prediction of each stratum’s underlying mean. However, descriptive inequality research asks a different question. It does not try to predict a hidden mean. It describes the inequality that is really there in the population we observe. On near-complete population data the observed stratum proportions are exact. They are the realised inequality, so there is no hidden target to recover and no noise to trade away. This is the point Bashir [59] makes: the method should fit the question. For realised inequality the fitting choice is the observed proportion, which is unbiased.

The same analysis [77] also shows where the advantage of RE-MAIHDA for prediction stops. It is largest when the between-stratum VPC is low, when some strata are small, particularly those whose observed proportions lie far from the overall proportion, and when the inequalities are mostly additive. It almost disappears when all strata are large. At the extremes it can even reverse. Leckie and colleagues [77] give a case where the model that shrinks towards the overall mean (their Model 1) is less accurate than the simple mean for a stratum far from the average. Those far strata are often the multiply marginalised groups that inequality research most wants to see. The version that keeps the additive effects (Model 2) does not shrink those, but it does shrink the part that departs from additivity, and that part is what many conceptualise as the intersectional inequality itself. For predicting a hidden mean this helps. For describing the real departures it dampens the very thing we want to show. The authors also note that the frequentist version understates its own uncertainty, so the smallest strata are reported both shrunk and over-confident.

Leckie and colleagues themselves [77] say that researchers who value unbiasedness over prediction may prefer simple means. The description of health inequalities is exactly that case. So, the disagreement is not about whether Random-Effects MAIHDA is sound. It is sound. It is about which question the description of health inequalities needs. For that question, Simple-Means MAIHDA is the faithful choice.

### 4.4 From conceptual critique to empirical visibility of shrinkage: the compare diagnostic

Random-Effects MAIHDA does not report a stratum’s own mean. It reports a compromise between that mean and the grand mean, weighted by how reliably the stratum is estimated. The reliability weight, *w = σ^2^u/(σ^2^u + σ^2^e/n)*, rises towards one as the between-stratum variance σ²_u grows and as the stratum size n increases. S3 Text sets out the algebra; here the consequence is what matters.

The GCE, reported as the VPC, rests on this same between-stratum variance. So when the GCE is large, w is close to one, the compromise barely moves, and Random-Effects MAIHDA reproduces the simple means. In that case shrinkage is irrelevant, because a strong GCE protects the stratum-specific estimates. The concern is the opposite case, a small or moderate GCE together with small strata, where w falls well below one and the estimate is pulled hard towards the grand mean. A small GCE does not rule out a real SCE in a particular small stratum. The between-stratum variance is an average of squared departures, in which one stratum barely registers, so the model, keyed to that small variance, shrinks a genuine effect away. That is where the description can be lost.

Shrinkage then does two things at once. It moves the point towards the grand mean, and for the smallest strata it draws the interval in around it, because the model borrows precision from the other strata. The result is a tight interval around a displaced point, which reads as confidence in an attenuated estimate.

A single example fixes the idea. Consider a small neighbourhood of five residents whose mean systolic blood pressure is 145 mmHg, against a population mean of 130. Simple-Means MAIHDA reports 145, with a wide interval of about 128 to 163 that honestly signals how little information five residents carry. Read the same neighbourhood through Random-Effects MAIHDA under two assumptions about the between-neighbourhood variance. When that variance is large, so neighbourhoods genuinely differ, w is about 0.83, the estimate barely moves to 142.5, and its interval stays close to the simple-means one. The SCE is preserved. When the between-neighbourhood variance is near zero, w falls to about 0.05, the estimate collapses to 130.7, almost the population mean, and its interval narrows to about 127 to 135, excluding the observed 145 entirely. The data on the neighbourhood are identical in both readings. What changes is one model-estimated quantity, the between-neighbourhood variance, and with it the GCE. A real elevation in a small stratum is preserved when the GCE is strong and erased, with false confidence, when it is weak.

Conventional benchmarks, proposed for the VPC on the latent scale, place a “large” VPC at 20% or above (Table 1), yet most MAIHDA studies report values between 3% and 15%. Whether shrinkage matters, however, is governed not by the VPC but by the reliability weight, which also depends on stratum size. At typical VPC values shrinkage is negligible only for large strata; for the small strata that intersectional inequality research most concerns, it remains material. Whether shrinkage matters in a given study is therefore not a question to settle in the abstract. It is an empirical one, set by the reliability weight and so by the GCE and the stratum sizes.

What has been lacking in the field is not the conceptual critique of shrinkage but a routine empirical procedure to make its consequences visible in any dataset. The compare option of the smaihda command closes this gap. It fits a standard RE-MAIHDA on the same area-sociodemographic strata used by the simple-means analysis and reports the two sets of results side by side, allowing the applied researcher to see how much the random-effects estimator has displaced the observed proportions.

The Malmö data offer a relatively favourable scenario for RE-MAIHDA: stratum sizes are reasonably uniform (mean n = 144 per A-SD stratum) and the population is large. Even so, the compare output (Section 3.4, Table 8; full details in S1 Text) makes the behaviour of shrinkage visible. The per-stratum attenuation depends on two things at once: how strongly each stratum is shrunk and how far it sits from the population average. Empirical-Bayes shrinkage removes a fraction of each stratum’s departure from the mean, and that fraction is larger for the smaller strata, which carry less information. The displacement in percentage points, however, is this fraction multiplied by the departure itself. The most attenuated stratum is therefore not the smallest one, but the one that combines a limited size with a large distance from the average. In our data the single largest attenuation, 6.44 percentage points, fell on a stratum of moderate size whose observed prevalence lay far above the population average, while the smallest strata, when they lay close to the average, were barely displaced. This pattern confirms rather than weakens our argument. The strata that random effects attenuate most are the strata that depart most from the average, that is, the strata where the inequality is strongest. Shrinkage does not merely smooth random noise; it pulls in the very strata that carry the inequality signal. Attenuation was also greater for the outcome with smaller contextual variance, psychotropic medication use, because a smaller τ² yields a smaller shrinkage factor. The summary measures (VPC and coAUC) agreed closely between methods, because Malmö’s reasonably uniform stratum sizes preserve the ranking of strata. RE-MAIHDA distorts per-stratum predictions most where the contextual signal is weakest, even under these favourable conditions.

The picture changes when stratum sizes differ widely, as is typical in intersectional and high-dimensional MAIHDA: the shrinkage factor is differential, leaving the smallest strata shrunk substantially while the largest are barely touched. A published MAIHDA-style study illustrates this issue.

It is an intersectional MAIHDA study of opioid misuse [45], reanalysed in S4 Text. The design has 72 strata defined by sex, race/ethnicity, income, and age, with stratum sizes ranging sixtyfold (50 to 3,030 individuals). Range compression reaches 70.2%, and the maximum absolute attenuation of 5.76 percentage points falls on young African-American women with high income (n = 50), one of the most marginalised intersectional positions. Mean attenuation in the smallest quartile of strata is 1.62 percentage points, against 0.16 percentage points in the largest quartile, a tenfold difference. The summary measures still agree closely between methods (coAUC 0.641 versus 0.640): a reader looking only at summary-level diagnostics would conclude that the two methods are interchangeable. The threshold-based classification at the population prevalence (6.08%) tells a different story. Simple means flag 28 strata as high-risk and capture 74.2% of cases; random effects flag only 25 and capture 65.4%. The nine-percentage-point sensitivity gap corresponds to roughly 230 cases that the random-effects high-risk strategy would miss, concentrated in the smallest and most marginalised intersectional strata. This is precisely the distortion that RE-MAIHDA is structurally prone to produce in intersectional research: the groups the analysis is most intended to bring into view are the ones it most systematically attenuates.

Read together, the two empirical illustrations make the same point at different intensities and in different MAIHDA designs. Random-Effects shrinkage introduces a systematic bias against the smallest groups, which are typically the most marginalised, and the magnitude of this bias scales with the very conditions in which inequality research most needs a transparent reading of the data [59]. The theoretical critique developed in Section 4.3 is therefore no longer something to be taken on trust: it is visible, quantifiable, and dataset-specific. We recommend that compare (or an equivalent diagnostic) be reported as standard practice in any MAIHDA study, so that readers can judge what the random-effects estimator is doing to the findings.

One limit of the diagnostic should be stated. It contrasts the simple means against the variance components model. It does not contrast them against the model that adds the additive main effects of the identity variables. That comparison would require treating the departure from additivity as the quantity of interest, and that is the decomposition this paper declines.

### 4.5 The unit of information in contextual analysis

Every SCE is a comparison of observed averages. Sometimes the reference is implicit. The deviation of an area from the population proportion compares that area with the population as a whole, or with other areas. Sometimes the reference is explicit. The comparison of rich with poor areas, standardised by EAR, contrasts one group of areas with another. In both cases the quantity reported is a difference between averages, and in both cases the comparison carries an association.

This matters for how the precision of an SCE should be judged. Describing one named context is one operation. The observed proportion of a given stratum is exact for that stratum, and its confidence interval reflects the number of individuals it contains. Generalising across contexts is a different operation. When we ask whether area income is associated with the outcome, the units that carry the information are the areas. In the present study there are 25 of them, not 43,291.

Conventional individual-level regression ignores this. It treats the individuals as independent and produces confidence intervals that are too narrow. Multilevel regression corrects the error through the design effect, one plus the average cluster size minus one, multiplied by the intraclass correlation, which here is the VPC. Dividing the number of individuals by the design effect gives the effective sample size. As the GCE grows, the effective sample size falls from the number of individuals towards the number of contexts.

This correction produces a paradox that we have described elsewhere [56]. When the GCE is small, the effective sample size stays close to the number of individuals, the confidence interval around the SCE is narrow, and the association is easily declared statistically significant. When the GCE is large, the effective sample size is small, and the same SCE may be reported as inconclusive. In other words, the less the context matters, the easier it is to publish a significant contextual association. An investigator who reports SCE without reporting the GCE has no way of knowing which situation applies.

S-MAIHDA does not need this correction, because it never made the error that the correction repairs. Its unit of analysis is the context from the outset. The observed area proportions are computed one area at a time, the EAR standardisation adjusts each area for its sociodemographic composition, and the comparison between rich and poor areas is a comparison of 25 units. The bootstrap used for the CCI, the VPC and the coAUC follows the same logic. The 25 areas are the observed set, not a sample of exchangeable areas, so they are held fixed and the uncertainty estimated is the uncertainty of the observed proportions themselves. Resampling whole areas would treat them as exchangeable draws from a wider set of areas, which is the assumption this framework declines in the estimation, and it answers a different question, the variability expected over other sets of areas like these. The number of individuals enters only where it belongs, in the precision of each stratum proportion.

The general point returns to the theme of this paper. It is worth being explicit about what the random-effects model estimates. Its parameters are the mean of the assumed distribution of strata and its variance. The stratum values are not parameters. The likelihood integrates over them, and they are recovered afterwards as posterior predictions. In social epidemiology these predictions have often been unpacked and read as descriptions of the strata themselves. That reading gives them a status the model does not confer. S-MAIHDA declines the whole operation. Its intervals are generally wider, and they are wider for a reason the reader can see.

### 4.6 Intersectionality analysed as a whole, geography as a structural axis

MAIHDA, specifically intersectional MAIHDA (I-MAIDA, is often used for quantitative operationalisation of intersectionality. It is therefore worth stating precisely how the present design treats the intersection, and what role geography plays in it.

S-MAIHDA treats each intersectional position as a whole. A stratum such as low-income women aged 55 to 64 enters the analysis as one social position with one observed proportion. Nothing in the estimation decomposes that position into main effects of sex, age and income plus an interaction term. The between-stratum variance is the variance of whole strata, not a residual left after additive effects of separate variables have been removed. This is deliberate, and it is more than a technical preference. Intersectionality theory holds that social positions are lived jointly, not as a sum of separable exposures [79]. Decomposing the intersection into main effects and an interaction term is therefore an epistemological error: it presumes the very separability that intersectionality denies. The stratum, not the variable, is the unit of description. The decomposition of Section 2.2.2 does not contradict this: G and the CMI split the within-area deviation of each stratum into a part common to all areas and a part reshaped by the area. They partition the geography of the intersectional inequality, never the intersection itself.

Geography plays a different role. Adding the area to the design does not decompose the intersection either. It contextualises it spatially: the same intersectional position is lived in different places, and the design observes it in each place. This is the quantitative counterpart of structural intersectionality. The area is not a confounder to be adjusted away, and not a covariate that explains the intersection. It lies outside the stratum and does not define it. It is a structural axis along which the same stratum is observed, and it can raise the risk of that stratum, lower it, or reshape the differences between strata. Interaction belongs here, between the axis and the whole stratum, and not within the stratum.

The three shares of Section 2.2.2 quantify this role. The CCI measures how much of the intersectional inequality is organised by place, so that strata resemble their area. G measures the differences that repeat in every area, the portion of the inequality that place does not touch. The CMI measures how much the area reshapes those differences. In the Malmö illustration the two outcomes sit at the two poles: for psychotropic medication use place organises almost nothing (CCI 4.1%, CMI 0.0%), whereas for private GP choice place both clusters the inequality (CCI 63.9%) and reshapes it (CMI 17.1%). The same three numbers, read in an intersectional register, say how strongly the structures that produce inequality are themselves located.

One balance should be kept. Treating place as a structural axis does not make every geographical difference structural, and the CCI and the CMI describe how the inequality is organised, not why. The mechanisms behind a high CCI or a high CMI, from segregation to the location of services, remain to be identified by other designs. The description does not name the mechanisms. It shows where they operate, and therefore where to look for them.

In RE-MAIHDA the proportional change in variance (PCV) from Model 1 to Model 2 reports the additive share of the between-stratum variance, and its complement the interaction share. Our three shares play the same role along a different axis. The interaction behind the PCV is between the dimensions that define the stratum, so obtaining it requires splitting the stratum. Ours is between the stratum and a structural axis external to it, so the stratum is never split. S-MAIHDA therefore does describe interaction. It describes the kind that does not require dismantling the position.

### 4.7 Limitations

Several limitations of the present study should be acknowledged. The study is cross-sectional, based on a single city in 2006, and the two outcomes examined are specific to the Swedish primary healthcare context. The findings are intended as an empirical illustration of the analytical approach rather than as conclusions about Malmö itself, and the paper makes no causal claims about why the observed inequalities exist.

Both outcomes examined here are common, with prevalences above 20%, and this bounds what the illustration can show. The information a stratum carries about its own risk depends not on the number of individuals it contains but on its expected number of events, the product of stratum size and prevalence. A stratum of 100 individuals expects 20 events when the prevalence is 20%, one event when it is 1%, and none when it is 0.1%. The shrinkage follows the events. Rare outcomes such as maternal mortality are precisely where inequality research often needs a stratum-specific description, and a low prevalence is a real limit: it either demands large strata or it leaves no stratum-specific description to make. It is there that S-MAIHDA and RE-MAIHDA would diverge most, so the divergence documented in this paper is a lower bound for such settings rather than an upper one.

A further point concerns what can be described at all. With a rare outcome spread across many strata, the expected number of events in each stratum may be so small that no method can describe stratum-specific inequality reliably. This is a property of the data. It is a constraint on descriptive observational epidemiology, not a defect of S-MAIHDA.

S-MAIHDA and RE-MAIHDA differ in how visible they make that constraint. S-MAIHDA displays it directly, as wide confidence intervals and as strata containing no events. RE-MAIHDA produces output that looks complete: the model converges, every stratum receives a point estimate, and every point estimate carries an interval. In sparse strata that interval is narrow because the assumed distribution supplies what the data do not. A reader who takes convergence and completeness as evidence that the question has been answered will be misled.

Where the expected number of events per stratum is too small, the honest response in descriptive work is to reduce the granularity of the strata, or to report that the data cannot support a stratum-specific description. Where the aim is prediction rather than description, RE-MAIHDA remains the appropriate tool, and its treatment of sparsity is a feature of that task rather than a distortion.

Sparsity also sets a limit on the standardised measures, which give all strata the same size. When many strata hold only a few individuals, their sampling noise enters those measures at full strength and no correction recovers the between-stratum components; the crude measures are less exposed because they are computed from the individual counts, so the large strata dominate. The smaihda command therefore computes the standardised partition and the CCI, G and CMI shares on strata with at least ten individuals, and reports how many strata and individuals are left out. In the present data no stratum is left out.

One feature of the design, by contrast, is better described as an explicit didactic move than as a limitation. The 25 areas are not the original Malmö neighbourhoods but random groupings of the 218 neighbourhoods, adopted to protect confidentiality. In doing so we deliberately reproduce a limitation that is common in applied epidemiology: the contextual units available to the analyst are predefined administrative boundaries that need not coincide with the collective body that actually conditions individual outcomes [31]. When the unit of analysis misses the relevant collective body, geographical effects are diluted and can appear weak or absent even where they are strong, which is why the geographical measures reported here are conservative (Section 3.3). That the simple-means approach still separates the strongly geographical outcome from the non-geographical one under this degraded structure is part of the demonstration, not a caveat to it.

The additive variance decomposition of simple-means MAIHDA assumes that each individual belongs to a single stratum, a clean partition. In its current form it does not extend to multiple-membership or other non-nested structures. In epidemiological practice this restricts less than it may seem: the contextual structuring of inequalities can usually be described with a clean partition of the population, as here. Multiple-membership designs keep their value for questions where individuals genuinely belong to several contexts at once; those are different questions, not the descriptive one this paper addresses.

Continuous individual-level covariates raise a related point. They cannot enter the partition directly, so adjusting for them requires categorisation, which adds dimensions and can create sparse cells. Modelling the covariate instead does not remove the sparsity; it fills it invisibly, extrapolating a common functional form into regions where a stratum has no data. An empty cell at least declares itself. Neither condition arises here, where the strata form a complete partition and the aim is to describe realised inequality rather than to adjust it away.

S-MAIHDA suits strata defined by a few dimensions, since every added dimension multiplies the number of strata and reduces their size. When many characteristics must be taken into account, as the patient mix in a comparison of hospitals, they can be summarised in a score obtained from a regression model, and the categories of the score can enter the definition of the strata. The need for enough individuals in every stratum is also a strength: rather than extrapolating from a model, the analysis shows where the information is scarce, with a wide interval or an undefined value.

The decomposition is additive on the probability scale. Near 0% a prevalence has little room to fall, so where the level of an area is low the modification of the strata with the lowest risk partly reflects the floor of the scale (Sections 2.2.1 and 3.2.2), and on a relative scale the modification would be different. The absolute scale is, however, the scale of the cases an intervention would have to prevent. Finally, S-MAIHDA describes the observed contexts and does not predict for contexts that were not observed. That is the question of the random-effects model, which assumes the exchangeability that the description of these contexts does not need (Section 4.8).

### 4.8 Implications for inequality research and public health

The case for simple means in MAIHDA reaches beyond the question of which estimator is technically preferable. It bears on the kind of public health science that MAIHDA can contribute to.

Random-Effects models have legitimate uses when outcome clustering is a nuisance to be adjusted for (twin studies, repeated measurements within individuals) or when prediction under a mean-squared-error criterion is the goal. Shrinkage is defensible when contextual variation is treated as noise and problematic when contextual variation is the object of interest. Descriptive inequality research is the second case.

The distinction can be put as two scenarios. In the first, the data are a random sample from a population, and the aim is to predict the unknown population means of the strata. In that scenario the empirical Bayes predictions are preferable to the observed sample means. In the second, the dataset in front of us is the finite population, as in the register data analysed here, and the aim is to report the means of that population. In that scenario the observed proportions are preferable, because they are those means. The finite population is the sharpest version of the argument, but not its boundary. Even in a random sample, when the question is descriptive, the estimand is each stratum’s own mean, the observed proportion is its unbiased estimator, and the confidence interval reports the imprecision honestly. Shrinkage does not supply the missing precision; it replaces it with an assumption. What separates the two scenarios is the question being asked; the finite population merely makes the answer exact.

It is worth being precise about what the random-effects machinery predicts. Its targets are the stratum means themselves, predicted as compromises between each stratum’s own data and the grand mean. Within this machinery the between-stratum variance and the reliability weights are not substantive quantities; they are instruments that calibrate how far each stratum is pulled towards the grand mean. Under exchangeability, the variance serves the mean.

Whether that service is justified depends on a second distinction: the substantive warrant for exchangeability. Consider repeated measurements within individuals, a legitimate home of random effects. There the context is the individual, and the common distribution has a mechanistic foundation: a shared physiology, a homeostatic set point around which measurements fluctuate. The latent mean has a real referent, the fluctuation around it is largely noise, and pooling borrows strength across individuals who share a biological machinery. Exchangeability is not a convenience there; it is a defensible substantive claim. The reliability weight completes the picture. In physiological quantities the between-individual variance is typically large relative to measurement noise, so the intraclass correlation is high, and a patient with a pathological mean keeps most of it even when measured only a few times: with an ICC of 0.7 and three measurements, about ninety per cent of the deviation is retained. The value is smoothed, not hidden. Descriptive social epidemiology sits at the opposite corner: the variance partition coefficients typically reported in MAIHDA studies lie between 3% and 15%, so the reliability weight collapses exactly where the strata are small, and a deviating stratum is not smoothed but largely erased. The same machinery is gentle where its assumption is defensible and severe where it is not.

Social strata offer no such foundation. The differences between strata are not random dispersion around a common social set point; they are produced by structured and directional forces, among them discrimination, racism, segregation, and the unequal location of services. There is no latent stratum essence for the observed proportion to be noise around: the realised proportion is the phenomenon. Treating strata as exchangeable draws from a common distribution is therefore not a mild approximation but a substantive claim that the data-generating process contradicts, and shrinking their differences towards a grand mean treats the very forces under study as noise.

The two polar cases now line up. In repeated measures within individuals, the question is predictive and exchangeability is mechanistically warranted: both considerations favour random effects. In descriptive research on social inequality, the question is descriptive and exchangeability lacks warrant: both considerations favour the observed proportions. The question commands in every case; the warrant for exchangeability sets the price of choosing prediction. None of this leaves description without inference: the uncertainty of the observed figures is estimated outside the random-effects machinery, by Wilson intervals and the within-context bootstrap, and their sampling noise is discounted by explicit accounting, as in the binomial-noise correction, rather than by a distributional assumption. A worked contrast between a patient and a stratum is given in Supplementary Information S3 Text.

S-MAIHDA and RE-MAIHDA answer different questions, and the choice follows from the question rather than from the statistical method.

A descriptive objective is appropriate when the aim is to quantify the magnitude of the health inequalities that exist in a population. S-MAIHDA serves this aim. The observed stratum proportion is the estimand itself, not an approximation to something else. This is the situation in monitoring inequalities across social groups and geographical areas, in comparing indicators between regions or over time, in evaluating progress towards goals such as the Sustainable Development Goals, universal health coverage, and national equity targets, and in reporting the epidemiological situation to policymakers through surveillance systems.

An application of S-MAIHDA to five rounds of the European Social Survey in 24 member states of the European Union reports the same measures with the country as the geographical context, together with what survey data add: post-stratification and analysis weights, participation that may depend on health and on social position, and the modification of the sociodemographic inequalities by the country (Merlo et al., in preparation medRxiv).

A predictive objective is appropriate when the aim is to estimate the expected risk of a stratum under an assumed distribution of strata, rather than the risk that was observed. RE-MAIHDA serves this aim, provided the strata can be treated as exchangeable. Shrinkage reduces the influence of random variation and yields more stable predictions. This is a genuine gain when prediction is the goal.

There is no such thing as a balance between statistical stability and faithful description in the study of social inequalities. The two belong to different estimands, and an estimand is not negotiable: shrinkage does not stabilise the description, it changes what is being described. Reducing random error is desirable when the target is an expected value. It is not desirable when the target is the inequality the population actually experienced, because the operation that reduces the error also displaces the estimate. Where a stratum is too small to support a firm statement, the confidence interval says so, and that is the correct scientific report.

This has an ethical dimension that is worth stating plainly. The strata that shrinkage displaces most are the small ones, and the small ones are usually those of historically marginalised populations. An analysis that smooths these strata towards the population average reports a more comfortable picture than the data contain, and it does so precisely where the inequality matters most.

Discriminatory accuracy at the population-prevalence threshold operationalises Rose’s prevention paradox within the MAIHDA framework. The connection between MAIHDA, Rose’s high-risk versus population strategy, and Marmot’s proportionate universalism has been developed in prior work [45, 56, 66] and the threshold-based classification reported here makes the trade-off empirically visible in the present dataset.

The implications for applied MAIHDA practice are direct. For descriptive inequality research, observed proportions should be the primary estimator. RE-MAIHDA produces reasonable approximations to the observed inequalities when stratum sizes are sufficiently large and uniform, and the compare diagnostic is the empirical tool that allows researchers to verify this condition in any specific dataset. Where this condition cannot be verified, or where small strata are present, the compare output must be reported transparently so that readers can judge what shrinkage has done to the findings. Even when RE-MAIHDA agrees closely with simple means, simple means remain epistemologically more suitable because they describe the data as observed rather than as smoothed by an exchangeability assumption.

The apparent flexibility of random-effects models can hide two operations. Strata with little information receive a prediction borrowed from the overall mean, which is a form of extrapolation, and every stratum is shrunk towards that mean, most strongly the small and the deviating ones, which are those where inequality lives. Both operations rest on the assumption that the strata are exchangeable draws from a common population. Where the two approaches agree, the model adds no information. Where they disagree, the difference is the cost of its assumption.

S-MAIHDA also extends naturally to the study of the interaction of effects on the additive scale [80], a central concern for research using multidimensional strata; we will develop this extension, grounded in the factorial decomposition of analysis of variance, in forthcoming work.

The axis in that extension is external to the stratum. Geography is one such axis, as shown here. The healthcare provider and the calendar period are others: the question in each case is whether the same sociodemographic differences repeat across the axis or are reshaped by it. S-MAIHDA studies how an intersectional position interacts with something outside it. It never studies how a position interacts with its own parts.

Finally, intersectional designs raise the problem of very small strata, and it helps to separate honest imprecision from bias. In S-MAIHDA a small stratum gives an unbiased point estimate with a wide confidence interval, but the point sits where the data place it, and the width only reports how little information is available. The wide interval is therefore a disclosure to report, not a defect. The shrinkage bias is the opposite case: a narrow interval placed in the wrong location (Section 4.4). Where more precision is genuinely needed it must come from more information, through designs that oversample marginalised groups or aggregation of strata decided on theoretical grounds before the outcome is seen, not from a model that supplies it by assumption. Simple means are not a methodological retreat. They are a commitment to letting the data speak for the people the analysis is meant to represent [51].

Documenting how health is distributed is a civic responsibility and a core public health function [51]. That monitoring is incomplete if it stops at averages. Distribution implies heterogeneity, and heterogeneity is rarely random. A substantial share of the differences between individuals is structured by their contexts, and S-MAIHDA makes that structure visible from the observed proportions, in terms that decision makers can read. In this sense it lets epidemiology act both as a mirror, describing the inequalities as they are, and as a compass, indicating where universal and proportionate action is needed.

**Box.**
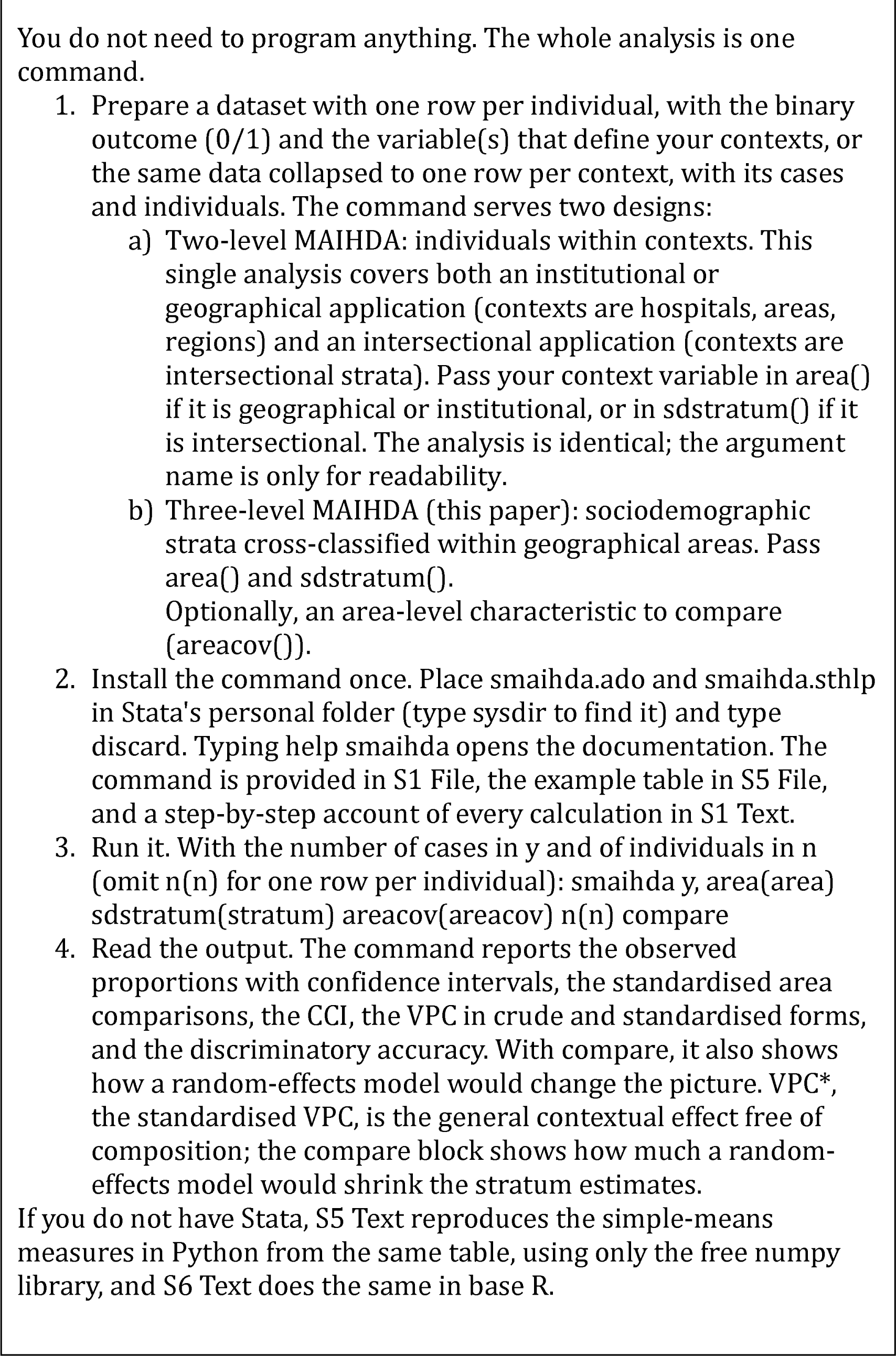
Running this analysis on your own data.

### 4.9 Anticipated objections and where they are addressed

Table 9 gathers the objections most often raised against describing realised inequality with observed proportions, together with a short response and the section where the argument is developed.

**Table 9.** Anticipated objections to Simple-Means MAIHDA and where each is addressed in this paper.

| Objection | Response | Where it is addressed |
| --- | --- | --- |
| **A. On the estimator** |  |  |
| Shrunken predictions are on average closer to the true stratum means, so simple means are the less accurate choice. | Correct for the predictive question, and we accept it. Descriptive inequality research does not predict a hidden mean. It describes the inequality realised in the population observed, and on near-complete population data the observed proportions are exact. | 4.3 |
| The mean squared error advantage does not require exchangeability, so the epistemic objection does not defeat it.[81, 82] | Also correct. The guarantee is for the total squared error across the ensemble, and the total is reduced by allowing individual components to be worse. The disagreement is about the loss function, not the theorem. Only the total is guaranteed to improve, and a small group whose true value lies far from the rest can end up worse estimated than by its own crude mean. | 4.3 |
| A stratum's true mean would have to be extremely deviant before shrinkage harmed it. | The threshold is not far out. Shrinkage begins to harm a stratum once its true value lies beyond a distance from the grand mean that is never below about 1.4 standard deviations. With 48 exchangeable strata the most extreme is expected to lie about 2.2 standard deviations out, which is its average position rather than an unusual one. | 4.3 |
| Shrinking towards the additive means rather than the grand mean removes the problem. | This is the conditional reading of exchangeability, applied to intersectional strata: name the reasons the groups differ; model them as covariates, and assume exchangeable only the residuals [83, 84]. Two things follow. That rescue names attributes a group has, whereas the reasons named here are the dimensions that constitute it, so modelling them does not adjust the group but dissolves it into its parts, and the residual whose exchangeability is then assumed is the part of a position left unexplained by its own definition. And the assumption only moves one level in: the residuals must now be judged exchangeable, that is, no particular intersection may be expected in advance to depart from additivity more than another; which is the negation of the claim the field exists to examine. The model also keeps the additive part while shrinking the departure from additivity, so for describing real departures it dampens what the analysis sets out to show. | 4.3, 4.6 |
| The dispersion of the observed proportions is inflated by sampling noise, so simple means exaggerate the inequality. | The arithmetic is granted: the variance between the observed proportions is the true dispersion plus the sampling noise. What does not follow is that the remedy is to shrink every stratum. Shrinkage alters three hundred values to correct one summary, and it overshoots, because the variance of the shrunken values is the reliability weight times the true dispersion; the picture moves from inflated to deflated rather than becoming right. We subtract the noise where it arises, in the variance itself, using the binomial variance $p(1-p)/n$, which is a fact of the counting process and not an assumption about society. The general contextual effect is reported uncorrected and noise-corrected side by side, and every stratum is left where the data put it. | 2.2.2, 3.2.1 |
| Bayesian estimation with weakly informative priors avoids the problem. | The prior is the exchangeability assumption in another form. A weaker prior reduces how much the strata are pooled, but not the direction of the pooling, and it does not supply the warrant. The question is not how strongly the strata are pooled but whether they should be pooled at all. | 4.2 |
| Simple means are unstable in the small strata that intersectional research most needs. | A small stratum gives an unbiased point estimate with a wide confidence interval. The point sits where the data place it, and the width reports how little information is available. The wide interval is a disclosure, not a defect. The alternative is a narrow interval placed in the wrong location. | 4.7 |
| Without a model there is no inference. | Uncertainty is estimated outside the random-effects machinery, by confidence intervals for each stratum and the within-context bootstrap for the summary measures, and known binomial noise is discounted by explicit accounting rather than by deforming each stratum's value. | 2.3, 4.5 |
| **B. On the assumptions** |  |  |
| Hospitals, schools and areas are not exchangeable either, yet random effects are standard there. | We agree on the premise. The disagreement is about the response. Where the estimand is an ensemble average the pooling is defensible. Where each named context is the object of the report, it is not. | 4.2 |
| Intersectional strata are strongly ordered, since more privileged positions have better outcomes, so the ordering should be built into the model. | Building it in means decomposing the position into main effects plus an interaction term, which presumes the separability that intersectionality denies. The stratum, not the variable, is the unit of description. | 4.6 |
| Variance components are estimated consistently even when the random effects are not normally distributed. | Accepted, and it answers a different question. Ours is what the between-stratum variance refers to when the strata are not draws from anything, which is prior to how well it is estimated. | 4.2 |
| A substantial intraclass correlation shows that the grouping matters and therefore justifies the random effects. | Structuration and exchangeability are different things. Persons nested in five-year age groups give a high intraclass correlation, and nobody would call the age groups exchangeable. The measure reports how much the grouping structures the outcome. It says nothing about whether the groups are draws from a common distribution. | 4.2 |
| **C. On the framework** |  |  |
| Is this not stratified analysis or fixed-effects regression under a new name? | It is not a regression. It is an exact decomposition of observed proportions, and it operationalises all three MAIHDA components, including the general contextual effects and the discriminatory accuracy of the context, which stratified description does not deliver. | 1, 2.2 |
| Dropping random effects abandons MAIHDA. | The commitment of MAIHDA is conceptual: individual heterogeneity is contextually organised in a nested structure. That commitment is preserved under any estimator faithful to the structure. MAIHDA is agnostic about statistical technique. | 1 |
| You decompose an interaction too, so you also split the intersection. | Not the same interaction. The three shares split the geography of the inequality into a part common to all areas and a part the area reshapes, and the stratum stays whole throughout. Intersectionality claims that the dimensions of a social position are lived jointly. It makes no such claim about a position and a place. | 4.6 |
| Why the probability scale rather than the logit scale? | Health inequality is experienced, counted and acted upon as a difference in risk. Quantities on the log-odds scale answer a different question and do not translate into that count | 4.2 |
|  | without further assumptions. |  |
| **D. On practice** |  |  |
| With 300 strata some will look extreme by chance, and simple means offer no protection. | The reported quantities are ensemble measures rather than a search for the extreme stratum, and the components most exposed to sampling noise are reported noise-corrected. Each stratum carries its own interval, so an extreme point resting on few individuals is visible as such. | 2.2.2, 3.2.2 |
| When strata are large the two approaches agree, so the choice does not matter. | Where they agree the simpler procedure is preferable, since it answers the question without an assumption the data do not need. Agreement cannot be assumed in advance, and establishing it for a given dataset is what the compare diagnostic does. | 4.4, 4.8 |
| The approach does not extend to multiple-membership or other non-nested structures. | Correct, and stated as a limitation. Those designs answer different questions from the descriptive one addressed here. | 4.7 |

## Ethics statement

This study used the LOMAS (Longitudinal Multilevel Analysis in Scania) database, constructed by the National Board of Health and Welfare and Statistics Sweden by record linkage of routine registers using the unique Swedish personal identification number. The database was delivered to the research group without personal identification numbers to preserve the anonymity of the individuals. The study was approved by the Regional Ethical Review Board in Lund, Sweden (Dnr 2014/856, 18 December 2014), and the data safety committees of the National Board of Health and Welfare and of Statistics Sweden approved the construction of the database. For the present analysis we used a fully anonymised sample, with all individual and area identifiers removed and replaced, which permits replication while preventing the identification of any individual or area.

## Data availability statement

The data supporting the findings of this study are available as S5 File: the stratum table, with the number of individuals and the number of cases in each area-sociodemographic stratum, which is all the analysis needs. It was built from a fully anonymised dataset published with a previous study (PLoS ONE, DOI: 10.1371/journal.pone.0153778), as described in S1 Text. A worked example in a spreadsheet, with every measure computed by formula, is S8 File. The Stata command is available as S1 File, the Python and R implementations as S2 File and S3 File, the programs that draw the result figures as S6 File, and the simulation of S7 Text as S7 File. The table contains no personal identifiers and cannot be linked back to individuals or areas. The underlying individual-level register data are held by Swedish authorities and are governed by Swedish legislation and data protection regulations; access requires application to the relevant register-holding authorities and ethical approval. The stratum counts of the opioid misuse reanalysis in S4 Text are available as S4 File.

## Declaration of generative AI and AI-assisted technologies in the writing process

During the preparation of this work the authors used a generative AI assistant (Claude, Anthropic) to support language and readability and to help draft and refine the wording of the manuscript and its supplementary material. The ideas and the interpretation are the authors’; the execution and the verification were assisted. After using this tool, the authors reviewed and edited the content as needed and take full responsibility for the content of the publication.

## Supporting information

**S1 Text.** Stata programs. The step-by-step implementation underlying the smaihda command, with each measure presented as a numbered step and the full output for both outcomes.

**S1 File.** The smaihda Stata package: the .ado source file, the .sthlp help file, the .pkg installation manifest and the test do-file.

**S2 Text.** Supplementary tables. Tables A and B, giving the prevalences of the area-sociodemographic strata in the poorest and the richest areas.

**S3 Text.** The compare measures. A standalone conceptual guide to the nine measures of the compare diagnostic, with the algebra of the attenuation.

**S4 Text.** Reanalysis of published data. A worked example on the intersectional analysis of opioid misuse, showing what shrinkage does when strata are small.

**S5 Text.** Python programs. A complete reproduction of the simple-means measures in Python, using only the numpy library and requiring no proprietary software.

**S6 Text.** R programs. An independent reproduction of the same measures in base R.

**S7 Text.** Simulation supporting the standardised contextual AUC (coAUC*). Four questions on coAUC*: its agreement with VPC*, its invariance to the distribution of stratum sizes, its behaviour under the prevalence of the outcome, and what size standardisation costs in precision.

**S2 File.** The Python implementation as a runnable script.

**S3 File.** The R implementation as a runnable script.

**S4 File.** The 72 intersectional strata of the opioid misuse reanalysis, with the number of individuals and the number of cases in each, as a comma-separated file.

**S5 File.** The data of this paper: the stratum table of the Malmö dataset, one row per area-sociodemographic stratum and outcome with the variables that define the stratum and its area, the number of individuals and the number of cases, as a comma-separated file.

**S6 File.** The programs that draw the result figures, Figs 3 to 10, from the stratum table of S5 File, in Python and in R, and the program that computes the numbers of Sections 2.2, 3.1.3 and 3.2.2 not printed by the command.

**S7 File.** The simulation as a runnable Python script, requiring only the numpy library.

**S8 File. A worked toy example of Simple-Means MAIHDA in a spreadsheet: three areas and two sociodemographic strata, with every measure of the paper computed by formula and a check cell for each identity.**

**S1 Checklist.** STROBE checklist for cross-sectional studies, with the section of the manuscript where each item is addressed.

## Supporting information

S1 Text. Stata programs

S1 File. The smaihda Stata package

S2 File. Python implementation

S3 Text. The compare measures

S3 File. R implementation

S4 Text. Reanalysis of published data (opioid misuse)

S4 File. Strata of the opioid misuse reanalysis (CSV)

S5 Text. Python programs

S5 File. The stratum table (data of this paper)

S6 Text. R programs

S6 File. Programs that draw the result figures

S7 Text. Simulation supporting the standardised contextual AUC

S7 File. The simulation as a runnable Python script

S8 File. A worked toy example in a spreadsheet

S1 Checklist. STROBE checklist

