## Supplementary material for "Describing health inequalities without distortion: Simple-Means MAIHDA vs Random-Effects MAIHDA": S1 Text. Stata programs

### Simple-Means MAIHDA: complete Stata programs with explanations

*Companion to the smaihda command, version 1.9.0. The command reports the standardised variance partition (VPC* and CCI), uncorrected and corrected for binomial noise, alongside the crude measures, and the contextual decomposition of the between-stratum inequality into clustering (CCI), additive differences (G) and modification (CMI). The Python and R programs (S5 Text and S6 Text) give the same point estimates. Their bootstrap intervals agree with those of the command within Monte Carlo error, because each program draws its own random numbers.*

**Dataset. Malmö, Sweden 2006, given as the stratum table of S5 File.** N = 43,291 individuals aged 35-64. 25 geographical areas (12 rich, 13 poor). 12 sociodemographic strata. 300 A-SD strata. Outcomes: psychotropic medication use (26.0%) and private GP choice (22.1%).

**Structure of programs**

- Notation and scales
- Confidence intervals: which method for which measure
- Step 0 - Build the stratum table from the original PLoS ONE data
- Step 1 - Records from the stratum table, and data verification
- Step 2 - Measure 1: Crude proportions with Wilson 95% CI
- Step 3 - Measure 2: Standardised area proportions (EAR method)
- Step 4 - Measure 3: Area income effect (ARD and RR with Ranstam CI)
- Step 5 - Measure 4: Deviation Δ_A,SD with Wilson-shift CI
- Step 6 - CCI: Contextual Clustering of Inequalities
- Step 7 - VPC: Variance partition coefficients (probability scale, variance decomposition)
- Step 7b - Contextual decomposition: CCI, G and CMI (noise-corrected)
- Step 8 - Discriminatory accuracy (coAUC and threshold classification)
- Step 9 - Figures (Figure 4 of the paper and the deviations from the crude area prevalence)
- Step 10 - Within-context bootstrap for CCI, VPC and coAUC
- Step 11 - Comparison with random-effects MAIHDA (compare option)

### Installing the smaihda command

The steps below reproduce every measure with plain Stata code, so that each calculation can be read and checked. The same analysis can be run in two lines with the smaihda command, provided in S1 File. The package contains four files: smaihda.ado, smaihda.sthlp, smaihda.pkg and smaihda_v1.9.0_test.do, a test that checks that the command gives the same results from the table and from the records.

The file names matter. In Stata the name of the ado-file is the name of the command, so the two files must be called exactly smaihda.ado and smaihda.sthlp. If they have been downloaded under any other name, rename them before use. Place both in Stata’s PERSONAL ado-path, which the command sysdir will display, and then restart Stata or type discard. Typing which smaihda should then report version 1.9.0.

The data are the stratum table, dataset-smaihda_stratum_table.csv (S5 File). Step 1 recovers the individual records from it and saves them as records-from-table.dta, which the later steps read by that name. The table and the records carry the same information, so every step gives the same result from either.

### Correspondence between these steps and the command output

The steps in this document are numbered 0 to 11 and follow the logic of the analysis. The smaihda command prints six numbered steps, which group several of them together. The correspondence is as follows.

- Step 0, building the table, and Step 1, the records and their verification, have no printed counterpart. With n(), the command recovers the records from the table itself. The command performs the checks of Step 1 internally and stops with an error message if they fail, but prints nothing when they pass.
- Step 2 corresponds to the command’s STEP 1, crude proportions with Wilson 95% CI.
- Step 3 corresponds to the command’s STEP 2, EAR-standardised area proportions.
- Step 4 corresponds to the command’s STEP 3, the area covariate effect.
- Step 5 corresponds to the command’s STEP 4, the deviation Delta_A,SD.
- Steps 6, 7, 7b and 8, which compute the CCI, the VPC, the contextual decomposition and the discriminatory accuracy, are reported together in the command’s STEP 6, Summary statistics.
- Step 9, the figures, is produced by the graph option rather than by a numbered step.
- Step 10 corresponds to the command’s STEP 5, the within-context bootstrap.
- Step 11 corresponds to the compare option, which is reported after the summary statistics.

### Notation and scales

Two kinds of quantity run through this document and through the main paper, and they are read on different footings. The measures that describe the contextual boundaries give all strata the same size, whatever their number of individuals: the standardised grand mean, the EAR-standardised area proportions, the standardised variance components, and everything built from them, that is, VPC*, the CCI, G and CMI, coAUC* and coAUC*EAR. Composition does not enter them. An asterisk marks a quantity of that kind. The measures that describe the population as it is are computed from the individual counts: the crude prevalences, the crude VPC and CCI, the coAUC and the threshold classification. Composition does enter them: the AUC asks how well the context separates people in the population as it is. Both readings are correct and they answer different questions.

Table A gives, for every symbol of the main paper, its default name in words, how it is computed, which of the two footings it follows, and the label under which the smaihda command prints it. The symbols are fixed. The name in words is a default: a study may choose another, provided it gives the symbol.

**Table A.** Notation of Simple-Means MAIHDA: symbol, default name, calculation, the footing on which each quantity is read, and the label printed by the smaihda command.

| Symbol | Default name | How it is computed | Given the same weight | Label printed by smaihda (Step in this document) |
| --- | --- | --- | --- | --- |
| p_stratum_ | Observed proportion (crude prevalence) of a stratum: an area, a sociodemographic stratum or an A-SD stratum | Cases divided by individuals in the stratum, with a Wilson 95% CI | Individual | STEP 1, crude proportions (Step 2) |
| p_pop_ | Population prevalence | Cases divided by all individuals. It is the default classification threshold | Individual | threshold (pop. prevalence) (Step 8) |
| p*_area_ | EAR-standardised area proportion (equivalent average rate) | Plain mean of the proportions of the 12 sociodemographic strata of the area | Stratum | STEP 2, EAR-standardised area proportions (Step 3) |
| p* | Standardised grand mean | Plain mean of the proportions of the 300 A-SD strata: the prevalence that would be observed if every stratum held the same number of people | Stratum | Used inside STEP 6 (Step 7) |
| ARD, RR | Absolute risk difference and relative risk for the area covariate | Difference and ratio between the two groups of areas, crude and EAR-standardised | Individual (crude), stratum (standardised) | STEP 3, area covariate effect (Step 4) |
| Δ_A,SD_ | Deviation of an A-SD stratum from its area | p_A,SD_ − p_A_, the stratum proportion minus the crude proportion of its area, with a Wilson-shift 95% CI (STEP 4 of the output and Tables A to D of S2 Text) | Individual (reference: the crude area prevalence) | STEP 4, deviations (Step 5) |
| Δ_stratum_, δ_SD_, γ_stratum_ | Within-area deviation, common sociodemographic difference and modification: the three terms of the contextual decomposition | Δ_stratum_ = p_stratum_ − p*_area_; δ_SD_ = mean of Δ_stratum_ over the 25 areas for the same sociodemographic stratum; γstratum = Δstratum − δSD; Δstratum is drawn in Figs 5a and 5b of the main paper | Stratum | Used inside the contextual decomposition (Step 7b) |
| V*_A_, V*_SD_, V*_I_ | Standardised variance components: between areas, between strata within areas, individual | Means of squared deviations with all strata given the same size; V*_A_ + V*_SD_ + V*_I_ = p*(1 − p*). Reported uncorrected and corrected for binomial noise, which is reassigned from the two contextual components to V*_I_ | Stratum | Through the standardised VPCs (Step 7) |
| VPC_A_, VPC_SD_, VPC_A+SD_ | Crude variance partition coefficients | Share of the total individual variance between areas, between strata within areas, and between A-SD strata, with every individual counted once | Individual | VPC_A, crude; VPC_SD, crude; VPC/ICC_A+SD, crude (Step 7) |
| VPC*_A_, VPC*_SD_, VPC*_A+SD_ | Standardised variance partition coefficients | The same shares built from V*_A_, V*_SD_ and V*_I_, uncorrected and corrected for binomial noise | Stratum | VPC_A std, uncorrected / corrected; VPC_SD std, uncorrected / corrected; VPC/ICC_A+SD std, uncorr / corrected (Step 7) |
| CCI | Contextual clustering of inequalities | V*_A_ / (V*_A_ + V*_SD_): the share of the between-stratum variance that lies between areas. A crude form, with every individual counted once, is also printed | Stratum (crude form: individual) | CCI, std, uncorrected / corrected; CCI, crude (Steps 6 and 7b) |
| G | Additive (shared) sociodemographic differences | V*_G_ / (V*_A_ + V*_SD_), with V*_G_ the mean of δ_SD_^2^: the share carried by the sociodemographic differences common to all areas | Stratum | G additive (Step 7b) |
| CMI | Contextual modification of inequalities | V*_CMI_ / (V*_A_ + V*_SD_), with V*_CMI_ the mean of γ_stratum_^2^, noise-corrected: the share carried by the area-by-sociodemographic modification. CCI + G + CMI = 1 | Stratum | CMI modification (Step 7b) |
| coAUC | Discriminatory accuracy of the context | Probability that a randomly chosen case belongs to a stratum with a higher proportion than a randomly chosen non-case, at the A-SD level and at the area level | Individual | coAUC (A-SD strata); coAUC (area level) (Step 8) |
| coAUC* | Standardised contextual AUC | The same probability with every stratum given the same total weight: the AUC that would be observed if all strata were of equal size | Stratum | coAUC* (A-SD strata); coAUC* (area, size) (Step 8) |
| coAUC*EAR | Contextual AUC standardised for size and composition | The area-level coAUC* with every area scored by its composition-standardised prevalence (EAR) instead of its crude prevalence | Area | coAUC*EAR (area) (Step 8) |
| Sensitivity, specificity, PPV | Classification at the population-prevalence threshold | A stratum is high-risk when its proportion exceeds the population prevalence; the cells TP, FP, FN and TN are counted in individuals | Individual | sensitivity / specificity; positive predictive value; TP / FP / FN / TN (Step 8) |

An asterisk marks a quantity computed with all strata given the same size. The random-effects counterparts reported by the compare option are described in Step 11 and in S3 Text.

### Confidence intervals: which method for which measure

One principle governs every interval reported here. The contexts are the observed set and are held fixed, so what is estimated is a property of these 25 areas and these 300 strata, and the only uncertainty is the uncertainty of measuring each proportion. Each area has one true proportion, and the individuals of that area are what allow it to be measured, so they are not redundant and the number of areas does not enter. No interval includes the variability of which areas were observed, since the analysis does not generalise to other areas. That would require the areas to be exchangeable draws from a wider set, which is the assumption this framework declines in the estimation and declines again here. Within that principle, the method used for each measure depends on its sampling structure (Table B). The reasoning is set out here once, and each step below applies it to the measure it computes.

Stratum-specific proportions use Wilson intervals, which remain within [0, 1] and maintain good coverage even for small strata or extreme proportions (Step 2).

The EAR-standardised area proportions follow the analytical formula for the variance of a weighted sum of independent binomial proportions (Ranstam 1998) (Step 3). The absolute risk difference (ARD) and the relative risk (RR) are derived from those standardised proportions and use large-sample approximations of their standard errors, with the RR computed on the log scale so that the bounds stay positive (Step 4).

The deviation of an A-SD stratum from its area, Δ_A,SD, uses the Wilson interval of the stratum proportion shifted by the area proportion, which is treated as known because it rests on the whole area (Step 5). A delta-method approximation, even with a correction for the dependence between the stratum and the area proportion, has zero width for a stratum in which everyone or no one has the outcome, so that such a stratum is always declared significant. The Wilson shift never degenerates.

The summary contextual measures (CCI, VPC, the contextual decomposition and the coAUC) use a non-parametric within-context bootstrap with 2,000 replicates (Davison and Hinkley 1997) (Step 10). The areas and the strata are the observed set, not a sample of exchangeable contexts, so each replicate holds them fixed and resamples the individuals with replacement inside each A-SD cell. Resampling whole areas would treat them as exchangeable draws from a wider set of areas, which is the assumption this framework declines in the estimation, and it answers a different question, the variability expected over other sets of areas like these. A replicate is drawn from proportions that already carry one dose of binomial noise, so it carries two, and every measure that depends on the dispersion of the proportions is centred above the estimate by one dose while its spread is the sampling spread. The reported interval is therefore the percentile interval recentred on the estimate and truncated to the admissible range of the measure.

Sensitivity and specificity at the population-prevalence threshold are reported as point estimates without confidence intervals. They summarise the consequences of a deterministic classification rule applied to the observed data, and are presented as descriptive characteristics of that rule rather than as inferential estimates.

**Table B.** Confidence interval methods used for each measure.

| Measure | CI method | Step |
| --- | --- | --- |
| Group-specific proportions for area, SD and A-SD | Wilson 95% CI | 2 |
| EAR-standardised area proportions | Ranstam (1998) analytical standard error | 3 |
| ARD and RR for the area covariate (crude and EAR-standardised) | Large-sample analytical method (Ranstam 1998) | 4 |
| Deviation of the A-SD stratum from its area, Δ_A,SD | Wilson interval of the stratum shifted by the area proportion (Wilson shift) | 5 |
| CCI, VPC, contextual decomposition (CCI, G, CMI) and coAUC | Within-context bootstrap (B = 2,000, contexts fixed, individuals resampled) | 10 |
| Sensitivity and specificity at the population-prevalence threshold | Point estimate (no CI) | 8 |

### Step 0: Build the Stratum Table from the Original PLoS ONE Data

**PURPOSE:** Reproduce the three-level Malmö dataset from the original Merlo et al. (2016) PLoS ONE data. The 218 SAMS neighbourhoods of Malmö are grouped into 25 geographical areas by random assignment with seed = 42, ensuring full reproducibility. Sociodemographic strata are defined by sex x age x income (12 strata). Area income is classified as rich or poor based on whether the proportion of low-income individuals in each area falls below the Malmö median across the 25 areas. The individual records are then collapsed into the stratum table, one row per A-SD stratum and outcome, which is the data of the paper.

**Input:** pone_0153778_s004.dta (original PLoS ONE supplementary dataset)

**Output: dataset-smaihda_stratum_table.csv (S5 File)**

```
  * ============================================================
  * Step 0: Build the stratum table from the original PLoS ONE data
  * Seed = 42 for reproducibility
  * ============================================================
  use "pone_0153778_s004.dta", clear

  * Rename variables for clarity
  rename Neigh     sams
  rename Id        id
  rename Psycmed   psycmed
  rename Private   private
  rename agegroup  agegroup6
  rename male      sex
  rename rich      income
  rename richarea  sams_income

  * Create 3 age groups from 6 original five-year groups
  * Groups 1-2 = age 35-44, 3-4 = 45-54, 5-6 = 55-64
  gen age3 = ceil(agegroup6 / 2)
  label define age3_lbl 1 "35-44" 2 "45-54" 3 "55-64"
  label values age3 age3_lbl

  * Create 25 areas by random SAMS assignment with seed=42
  * 218 SAMS: 18 areas of 9 SAMS and 7 areas of 8 SAMS
  preserve
    keep sams
    duplicates drop
    set seed 42
    gen rand = runiform()
    sort rand
    gen area = .
    local idx = 1
    forval a = 1/25 {
      local size = cond(`a' <= 18, 9, 8)
      local end_idx = `idx' + `size' - 1
      replace area = `a' in `idx'/`end_idx'
      local idx = `end_idx' + 1
    }
    keep sams area
    save "sams_area_map.dta", replace
  restore
  merge m:1 sams using "sams_area_map.dta", nogen

  * Area-level income: classify rich/poor by Malmö median
  * Rich area = proportion low-income below median across 25 areas
  bysort area: egen area_lowinc_prop = mean(1 - income)
  bysort area: gen area_tag = (_n == 1)
  quietly sum area_lowinc_prop if area_tag, det
  local malmo_median = r(p50)
  gen area_income = (area_lowinc_prop < `malmo_median')
  drop area_lowinc_prop area_tag

  * Verify: should give 12 rich and 13 poor areas
  bysort area: gen vtag = (_n == 1)
  tab area_income if vtag
  drop vtag

  * Create 12 SD strata: sex(2) x age3(3) x income(2)
  egen strata_sociodem_id = group(sex age3 income)

  * Create 300 A-SD strata: area(25) x SD stratum(12)
  egen strata_global_id = group(area strata_sociodem_id)

  * Label variables
  label variable area               "Geographical area (1-25)"
  label variable age3               "Age group"
  label variable sex                "Sex (0=women, 1=men)"
  label variable income             "Individual income (0=low, 1=high)"
  label variable area_income        "Area income (0=poor, 1=rich)"
  label variable strata_sociodem_id "SD stratum (1-12)"
  label variable strata_global_id   "A-SD stratum (1-300)"

  * The stratum table (S5 File): one row per A-SD stratum and outcome,
  * with the variables that define the stratum and its area, the number
  * of individuals n and the number of cases. It is the data of the paper.
  tempfile tab
  local first = 1
  foreach y in psycmed private {
    preserve
    collapse (count) n = `y' (sum) cases = `y', ///
      by(area strata_sociodem_id strata_global_id sex age3 income area_income)
    gen outcome = "`y'"
    gen byte order_y = (`first' == 0)
    if `first' == 0 append using `tab'
    save `tab', replace
    local first = 0
    restore
  }
  use `tab', clear
  sort area strata_sociodem_id order_y
  drop order_y
  order area strata_sociodem_id strata_global_id sex age3 income ///
    area_income outcome n cases
  export delimited using "dataset-smaihda_stratum_table.csv", nolabel replace
```

### Step 1: Records from the Stratum Table, and Data Verification

**PURPOSE: Recover the individual records from the stratum table and verify them before beginning the analysis. The records of each outcome are those of the table, expanded.** Confirm N = 43,291 with 25 areas, 12 SD strata, and 300 A-SD strata. Check that outcome prevalences are 26.0% for psychotropic medication and 22.1% for private GP. Check stratum size distributions to ensure adequate sizes throughout.

* The data: the stratum table (S5 File), one row per A-SD stratum and
 * outcome, with the number of individuals n and the number of cases
 import delimited using "dataset-smaihda_stratum_table.csv", clear
 tabstat n cases, by(outcome) statistics(count sum)

 * The individual records. Each stratum is expanded to its n individuals,
 * and the first ones, as many as the cases, have the outcome. Both
 * outcomes have the same n in every stratum, so they share the records.
 * Which individual has which pair of outcomes cannot be recovered from
 * the table, and no measure depends on it.
 reshape wide cases, i(strata_global_id) j(outcome) string
 expand n
 bysort strata_global_id: gen byte psycmed = (_n <= casespsycmed)
 by strata_global_id: gen byte private = (_n <= casesprivate)
 drop n casespsycmed casesprivate
 save "records-from-table.dta", replace

 * Overall structure
 count
 sum psycmed private
 tab area_income

 * Strata counts
 codebook strata_global_id strata_sociodem_id area, compact

 * A-SD stratum sizes (should range 35-299, mean ~144)
 bysort strata_global_id: gen asd_n = _N
 sum asd_n, detail

### Step 2: Measure 1 - Crude Proportions with Wilson 95% CI

**PURPOSE:** Compute observed proportions with Wilson confidence intervals for (a) the 12 SD strata, (b) the 25 areas, and (c) the 300 A-SD strata. The Wilson method is appropriate across the full range of proportions and stratum sizes (Agresti and Coull 1998). This is the gold standard estimator for descriptive inequality research: unbiased, model-free, and transparent. Strata whose CI lies entirely above the population prevalence threshold (26.0% for psycmed, 22.1% for private GP) are classified as high risk.

Saved files: sd_strata_psycmed.dta, sd_strata_private.dta, areas_crude_psycmed.dta, areas_crude_private.dta, asd_strata_psycmed.dta, asd_strata_private.dta

```
  use "records-from-table.dta", clear

  * Thresholds are the population prevalences, taken from the data
  quietly summarize psycmed
  local thresh_psycmed = r(mean)
  quietly summarize private
  local thresh_private = r(mean)

  * Wilson CI program
  capture program drop wilson_ci
  program define wilson_ci, rclass
    args n events
    local p   = `events' / `n'
    local z   = 1.96
    local den = 1 + `z'^2 / `n'
    local ctr = (`p' + `z'^2/(2*`n')) / `den'
    local mrg = (`z'/`den') * sqrt(`p'*(1-`p')/`n' + `z'^2/(4*`n'^2))
    return scalar p  = `p'
    return scalar lo = `ctr' - `mrg'
    return scalar hi = `ctr' + `mrg'
  end

  * ----- 2a: SD strata (12 strata) -----
  foreach outcome in psycmed private {
    preserve
    collapse (count) n=`outcome' (sum) events=`outcome', ///
      by(strata_sociodem_id sex age3 income)
    gen p=.
    gen lo=.
    gen hi=.
    gen high_risk=.
    forval i = 1/`=_N' {
      wilson_ci `=n[`i']' `=events[`i']'
      replace p  = r(p)  in `i'
      replace lo = r(lo) in `i'
      replace hi = r(hi) in `i'
      replace high_risk = (lo > `thresh_`outcome'') in `i'
    }
    format p lo hi %6.3f
    list strata_sociodem_id sex age3 income n events p lo hi high_risk, noobs
    save "sd_strata_`outcome'.dta", replace
    restore
  }

  * ----- 2b: Areas (25 areas) -----
  foreach outcome in psycmed private {
    preserve
    collapse (count) n=`outcome' (sum) events=`outcome', ///
      by(area area_income)
    gen p=.
    gen lo=.
    gen hi=.
    gen high_risk=.
    forval i = 1/`=_N' {
      wilson_ci `=n[`i']' `=events[`i']'
      replace p  = r(p)  in `i'
      replace lo = r(lo) in `i'
      replace hi = r(hi) in `i'
      replace high_risk = (lo > `thresh_`outcome'') in `i'
    }
    format p lo hi %6.3f
    save "areas_crude_`outcome'.dta", replace
    restore
  }

  * ----- 2c: A-SD strata (300 strata) -----
  foreach outcome in psycmed private {
    preserve
    collapse (count) n=`outcome' (sum) events=`outcome', ///
      by(strata_global_id area strata_sociodem_id area_income)
    gen p=.
    gen lo=.
    gen hi=.
    gen high_risk=.
    forval i = 1/`=_N' {
      wilson_ci `=n[`i']' `=events[`i']'
      replace p  = r(p)  in `i'
      replace lo = r(lo) in `i'
      replace hi = r(hi) in `i'
      replace high_risk = (lo > `thresh_`outcome'') in `i'
    }
    sort area strata_sociodem_id
    save "asd_strata_`outcome'.dta", replace
    restore
  }
```

### Step 3: Measure 2 - Standardised Area Proportions (EAR Method)

**PURPOSE:** Compute sociodemographically standardised area proportions using the Equivalent Average Rate (EAR) method (Merlo et al. 1994). The EAR gives each of the 12 SD strata equal weight within each area. This is equivalent to direct standardisation against a hypothetical population with equal stratum sizes and requires no external reference population. The standardised proportion p_std_A represents the area’s average risk net of sociodemographic composition.

Uncertainty is quantified analytically using variance propagation. The SE of the EAR equals sqrt(1/144 sum(v_k)), where v_k is the variance implied by the Wilson interval of stratum k, ((hi − lo)/3.92)², and 144 = 12 squared is the squared number of strata. The variance implied by the Wilson interval is used rather than p(1-p)/n, which is zero for a stratum in which everyone or no one has the outcome. The 95% CI uses the normal approximation.

Saved files: area_std_psycmed.dta, area_std_private.dta

```
  use "records-from-table.dta", clear

  foreach outcome in psycmed private {
    * A-SD stratum proportions
    bysort area strata_sociodem_id: gen n_asd = _N
    bysort area strata_sociodem_id: gen p_asd = sum(`outcome') / _N

    * EAR = unweighted mean of 12 SD strata per area
    bysort area strata_sociodem_id: gen asd_tag = (_n == 1)
    gen aux1 = p_asd if asd_tag == 1
    bysort area: egen p_std = mean(aux1)
    drop aux1

    * Variance propagation: SE = sqrt(sum(v_k) / 144), v_k from the
    * Wilson interval width of each stratum
    gen wden = 1 + 1.96^2 / n_asd
    gen wmrg = (1.96 / wden) * sqrt(p_asd*(1-p_asd)/n_asd + 1.96^2/(4*n_asd^2))
    gen var_stratum = (wmrg / 1.96)^2
    drop wden wmrg
    gen aux2 = var_stratum if asd_tag == 1
    bysort area: egen sum_var = sum(aux2)
    drop aux2
    gen se_std    = sqrt(sum_var / 144) if asd_tag == 1
    gen p_std_lo  = p_std - 1.96 * se_std
    gen p_std_hi  = p_std + 1.96 * se_std

    * Collapse to area level and save
    preserve
    keep if asd_tag == 1
    collapse (first) area_income p_std se_std p_std_lo p_std_hi, by(area)
    format p_std p_std_lo p_std_hi %6.3f
    sort area_income area
    save "area_std_`outcome'.dta", replace
    restore

    drop p_asd asd_tag p_std sum_var var_stratum se_std ///
         p_std_lo p_std_hi n_asd
  }
```

### Step 4: Measure 3 - Area Income Effect (ARD and RR)

**PURPOSE:** Describe the observed difference between rich and poor areas for each outcome. Rich and poor areas are compared using equal-weighted means, giving each area the same weight regardless of size. The crude comparison uses raw area proportions. The adjusted comparison uses EAR-standardised proportions from Step 3, removing the influence of sociodemographic composition.

Confidence intervals use the Ranstam (1998) large-sample method. For each group (rich/poor), the SE of the equal-weighted mean equals sqrt(sum(p(1-p)/n) / k^2) where k is the number of areas in the group. ARD CI uses the normal approximation. RR CI is computed on the log scale to ensure bounds are positive.*

```
  foreach outcome in psycmed private {

    * --- Crude ARD and RR ---
    use "areas_crude_`outcome'.dta", clear

    * Equal-weighted means by income group
    bysort area_income: egen mean_p = mean(p)
    gen var_p = p * (1 - p) / n
    bysort area_income: egen sum_var = sum(var_p)
    bysort area_income: gen k = _N
    gen se_mean = sqrt(sum_var / k^2)
    bysort area_income: keep if _n == 1

    * ARD with 95% CI
    local poor_c  = mean_p[1]
    local rich_c  = mean_p[2]
    local se_p    = se_mean[1]
    local se_r    = se_mean[2]
    local ARD_c   = (`poor_c' - `rich_c') * 100
    local se_ARD  = sqrt(`se_p'^2 + `se_r'^2)
    local ARD_lo  = `ARD_c' - 1.96*`se_ARD'*100
    local ARD_hi  = `ARD_c' + 1.96*`se_ARD'*100

    * RR with 95% CI (log scale)
    local RR_c    = `poor_c' / `rich_c'
    local se_lRR  = sqrt(`se_p'^2/`poor_c'^2 + `se_r'^2/`rich_c'^2)
    local RR_lo   = `RR_c' * exp(-1.96*`se_lRR')
    local RR_hi   = `RR_c' * exp( 1.96*`se_lRR')

    di "CRUDE `outcome':"
    di "  ARD = " %5.2f `ARD_c' " pp (" %5.2f `ARD_lo' " to " %5.2f `ARD_hi' ")"
    di "  RR  = " %5.3f `RR_c'  " (" %5.3f `RR_lo'  " to " %5.3f `RR_hi'  ")"

    * --- Adjusted ARD and RR ---
    use "area_std_`outcome'.dta", clear
    bysort area_income: egen mean_std = mean(p_std)
    gen var_std = se_std^2
    bysort area_income: egen sum_var_s = sum(var_std)
    bysort area_income: gen k_s = _N
    gen se_mean_s = sqrt(sum_var_s / k_s^2)
    bysort area_income: keep if _n == 1
    local poor_s  = mean_std[1]
    local rich_s  = mean_std[2]
    local se_ps   = se_mean_s[1]
    local se_rs   = se_mean_s[2]
    local ARD_s   = (`poor_s' - `rich_s') * 100
    local se_ARDs = sqrt(`se_ps'^2 + `se_rs'^2)
    local ARDs_lo = `ARD_s' - 1.96*`se_ARDs'*100
    local ARDs_hi = `ARD_s' + 1.96*`se_ARDs'*100
    local RR_s    = `poor_s' / `rich_s'
    local se_lRRs = sqrt(`se_ps'^2/`poor_s'^2 + `se_rs'^2/`rich_s'^2)
    local RRs_lo  = `RR_s' * exp(-1.96*`se_lRRs')
    local RRs_hi  = `RR_s' * exp( 1.96*`se_lRRs')

    di "ADJUSTED `outcome':"
    di "  ARD = " %5.2f `ARD_s' " pp (" %5.2f `ARDs_lo' " to " %5.2f `ARDs_hi' ")"
    di "  RR  = " %5.3f `RR_s'  " (" %5.3f `RRs_lo'  " to " %5.3f `RRs_hi'  ")"
  }
```

### Step 5: Measure 4 - Deviation Δ_A,SD with Wilson-Shift CI

**PURPOSE:** For each of the 300 A-SD strata, compute the deviation of the stratum proportion from its area’s crude proportion: Δ_A,SD = p_A,SD − p_A. A positive value means the stratum has higher risk than the area average. A negative value means lower risk. This measures how the sociodemographic gradient differs across geographical areas, which is the signature of geographical effect modification.

Confidence intervals use the Wilson interval of the stratum proportion, shifted by the area proportion: dev_lo = lo_ASD − p_A and dev_hi = hi_ASD − p_A, where lo_ASD and hi_ASD are the Wilson 95% limits of Step 2. The area proportion is treated as known, because it rests on the whole area (1,000 to 3,000 individuals here). A delta-method interval, dev ± 1.96 SE with Var(Δ) = p_ASD(1-p_ASD)/n_ASD + p_A(1-p_A)/n_A − 2p_ASD(1-p_ASD)/n_A, has zero width for a stratum in which everyone or no one has the outcome, so that such a stratum is always declared significant. The Wilson shift never degenerates. In Malmö, 125 strata deviate significantly for psychotropic medication use and 122 for private GP choice (Tables A and B in S2 Text).

Saved files: dev_asd_psycmed.dta, dev_asd_private.dta

```
  use "records-from-table.dta", clear

  foreach outcome in psycmed private {
    preserve
    collapse (count) n_asd=`outcome' (sum) ev_asd=`outcome', ///
      by(strata_global_id area strata_sociodem_id area_income)
    gen p_asd = ev_asd / n_asd

    * Merge area-level proportions
    merge m:1 area using "areas_crude_`outcome'.dta", ///
      keepusing(n p) nogen
    rename n n_area
    rename p p_area

    * Deviation from area mean
    gen dev = p_asd - p_area

    * Wilson interval of the stratum, shifted by the area proportion,
    * which is treated as known
    gen wden = 1 + 1.96^2 / n_asd
    gen wctr = (p_asd + 1.96^2/(2*n_asd)) / wden
    gen wmrg = (1.96 / wden) * sqrt(p_asd*(1-p_asd)/n_asd + 1.96^2/(4*n_asd^2))
    gen dev_lo  = (wctr - wmrg) - p_area
    gen dev_hi  = (wctr + wmrg) - p_area
    gen se_dev  = (dev_hi - dev_lo) / (2*1.96)
    drop wden wctr wmrg

    * Significant if CI excludes zero
    gen sig = (dev_lo > 0 | dev_hi < 0)
    format dev dev_lo dev_hi %6.3f
    sort area strata_sociodem_id
    save "dev_asd_`outcome'.dta", replace
    restore
  }
```

### Step 6: CCI - Contextual Clustering of Inequalities

**PURPOSE:** Compute the Contextual Clustering of Inequalities (CCI). This rests on a standard variance decomposition of a binary outcome. We take the between-stratum variance and ask how much of it lies between geographical areas. V_A is the weighted sum of squared deviations of area proportions from the population prevalence, divided by N. V_SD is the weighted sum of squared deviations of A-SD stratum proportions from their area proportion, divided by N. CCI = V_A / (V_A + V_SD). A value near 0 means sociodemographic inequalities are similar across areas. A value near 1 means they are strongly clustered geographically. Uncertainty is estimated by the within-context bootstrap (Step 10).

```
  use "records-from-table.dta", clear

  foreach outcome in psycmed private {

    * Grand mean and N
    quietly sum `outcome'
    local grand = r(mean)
    local N     = r(N)

    * V_A: variance between area means
    preserve
    collapse (count) n_area=`outcome' (sum) ev=`outcome', by(area)
    gen p_area = ev / n_area
    gen va_contrib = n_area * (p_area - `grand')^2
    quietly sum va_contrib
    local V_A = r(sum) / `N'
    restore

    * V_SD: variance within areas (weighted squared deviations)
    preserve
    use "dev_asd_`outcome'.dta", clear
    gen vsd_contrib = n_asd * dev^2
    quietly sum vsd_contrib
    local V_SD = r(sum) / `N'
    restore

    * CCI
    local CCI = `V_A' / (`V_A' + `V_SD')

    di "`outcome':"
    di "  V_A   = " %8.6f `V_A'
    di "  V_SD  = " %8.6f `V_SD'
    di "  CCI  = " %5.1f (`CCI'*100) "%"
    di "  1-CCI= " %5.1f ((1-`CCI')*100) "%"
  }

  * 95% CI for CCI is computed in Step 10 using the within-context bootstrap.
```

The command also reports the standardised CCI, in which all occupied A-SD strata are given the same size, whatever their number of individuals. This isolates the geographical structuring of inequality from population composition, as the EAR (Step 3) standardises area proportions across sociodemographic strata. It is reported in two forms, uncorrected and corrected for binomial noise, and returned by the command as r(GCCI_s) and r(GCCI_c) with their bootstrap CIs. The block below is the underlying computation.

```
  * --- Standardised CCI (weight 1 per A-SD stratum) ---
  foreach outcome in psycmed private {
    preserve
      use "dev_asd_`outcome'.dta", clear
      quietly count
      local K = r(N)
      bysort area: egen p_a_star = mean(p_asd)   // unweighted area mean p*_a
      quietly sum p_asd
      local pbar = r(mean)                        // grand standardised mean
      gen va_s  = (p_a_star - `pbar')^2
      quietly sum va_s
      local V_A_s = r(sum) / `K'
      gen vsd_s = (p_asd - p_a_star)^2
      quietly sum vsd_s
      local V_SD_s = r(sum) / `K'
      local GCCI_s = `V_A_s' / (`V_A_s' + `V_SD_s')
      * binomial-noise correction (reassigned, not removed)
      gen nz = p_asd*(1 - p_asd) / n_asd
      quietly sum nz
      bysort area: gen _Sa = _N                   // strata in the area (12 here)
      gen _nzS = (1 - 1/_Sa) * nz                 // within-area noise factor, per stratum
      quietly sum _nzS
      local nz_SD = r(sum) / `K'
      bysort area: egen nz_am = mean(nz)
      bysort area: gen _first = _n == 1
      gen _nzAw = (1 - _Sa/`K') * nz_am             // v1.9.0: the area is compared with pbar, the mean of the same K strata
      quietly sum _nzAw if _first
      local nz_A = r(sum) / `K'
      local V_A_c  = `V_A_s'  - `nz_A'
      local V_SD_c = `V_SD_s' - `nz_SD'
      local GCCI_c = `V_A_c' / (`V_A_c' + `V_SD_c')
      di "`outcome' (standardised CCI): uncorrected = " ///
         %5.1f (`GCCI_s'*100) "%   corrected = " %5.1f (`GCCI_c'*100) "%"
    restore
  }
```

### Step 7: VPC - Variance Partition Coefficients (probability scale)

**PURPOSE:** Estimate variance partition coefficients from the observed proportions, on the probability scale, by direct variance decomposition. No multilevel model is required. A binary outcome has total variance p(1-p). We decompose it into three parts: the variance between areas (V_A), the variance between sociodemographic strata within areas (V_SD), and the residual individual variance within strata (V_I). Each part is a weighted mean of squared deviations across individuals.

The command computes the standardised components (V_A*, V_SD*, V_I*, their corrected forms and the standardised CCI) on the A-SD strata with at least minn() individuals, default 10, and prints how many strata and individuals are excluded. The crude components, the coAUC and the classification always use all strata. With many very small strata the standardised measures are dominated by binomial sampling noise and no noise term recovers the between-stratum components; a minimum stratum size does. In Malmö no stratum is excluded. The command also prints a cell-size diagnostic in STEP 1: median and minimum stratum size, strata and individuals with n < 5, < 10 and < 30, and strata with no or with all events.

V_A and V_SD are the same components computed for the CCI in Step 6. V_I is the within-stratum Bernoulli variance, the weighted mean of p_A,SD (1 - p_A,SD). The three parts add up to the total: V_A + V_SD + V_I = p_pop (1 - p_pop). The coefficients are VPC_A = V_A / (V_A + V_SD + V_I), VPC_SD = V_SD / (V_A + V_SD + V_I), and VPC_A+SD = (V_A + V_SD) / (V_A + V_SD + V_I). There is no logit transformation and no fixed pi^2/3 residual. 95% CI computed in Step 10.

```
  use "records-from-table.dta", clear

  foreach outcome in psycmed private {

    * Population prevalence (grand mean) and total N
    quietly sum `outcome'
    local grand = r(mean)
    local N     = r(N)

    * --- V_A: between-area variance (same V_A as in Step 6) ---
    preserve
      collapse (count) n_area=`outcome' (sum) ev_area=`outcome', by(area)
      gen p_area     = ev_area / n_area
      gen va_contrib = n_area * (p_area - `grand')^2
      quietly sum va_contrib
      local V_A = r(sum) / `N'
    restore

    * --- V_SD and V_I: from the 300 A-SD strata (Step 5 file) ---
    preserve
      use "dev_asd_`outcome'.dta", clear
      * V_SD: within-area between-stratum variance (same V_SD as in Step 6)
      gen vsd_contrib = n_asd * dev^2
      quietly sum vsd_contrib
      local V_SD = r(sum) / `N'
      * V_I: within-stratum Bernoulli residual, p_asd*(1-p_asd)
      gen vi_contrib = n_asd * p_asd * (1 - p_asd)
      quietly sum vi_contrib
      local V_I = r(sum) / `N'
    restore

    * --- Variance partition coefficients on the probability scale ---
    local V_total = `V_A' + `V_SD' + `V_I'
    local VPC_A   = `V_A'  / `V_total'
    local VPC_SD  = `V_SD' / `V_total'
    local VPC_ASD = (`V_A' + `V_SD') / `V_total'

    * --- Identity check: components sum to p_pop(1-p_pop) ---
    local check = `grand' * (1 - `grand')

    di "`outcome':"
    di "  V_A               = " %8.6f `V_A'
    di "  V_SD              = " %8.6f `V_SD'
    di "  V_I               = " %8.6f `V_I'
    di "  V_A+V_SD+V_I       = " %8.6f `V_total'
    di "  p_pop(1-p_pop)     = " %8.6f `check'  "   <- identity check"
    di "  VPC_A    = " %5.2f (`VPC_A'*100)   "%   (= ICC_A)"
    di "  VPC_SD   = " %5.2f (`VPC_SD'*100)  "%"
    di "  VPC_A+SD = " %5.2f (`VPC_ASD'*100) "%   (= ICC_A+SD)"
  }
```

The command also reports the standardised VPC, VPC*, with weight one per A-SD stratum, in two forms: uncorrected, and corrected for binomial noise. The standardised components satisfy the same identity, V_A* + V_SD* + V_I* = pbar*(1 - pbar*), preserved exactly because the noise is reassigned from the between-stratum levels to the individual level, not removed. The command returns r(ICC_ASD_s) and r(ICC_ASD_c) with their bootstrap CIs, alongside the crude r(ICC_ASD). The block below is the underlying computation.

```
  * --- Standardised VPC (weight 1 per A-SD stratum) ---
  foreach outcome in psycmed private {
    preserve
      use "dev_asd_`outcome'.dta", clear
      quietly count
      local K = r(N)
      bysort area: egen p_a_star = mean(p_asd)
      quietly sum p_asd
      local pbar = r(mean)
      gen va_s  = (p_a_star - `pbar')^2
      gen vsd_s = (p_asd - p_a_star)^2
      gen vi_s  = p_asd * (1 - p_asd)
      quietly sum va_s
      local V_A_s = r(sum) / `K'
      quietly sum vsd_s
      local V_SD_s = r(sum) / `K'
      quietly sum vi_s
      local V_I_s = r(sum) / `K'
      local den_s = `pbar' * (1 - `pbar')
      * binomial-noise correction
      gen nz = p_asd*(1 - p_asd) / n_asd
      quietly sum nz
      bysort area: gen _Sa = _N                   // strata in the area (12 here)
      gen _nzS = (1 - 1/_Sa) * nz                 // within-area noise factor, per stratum
      quietly sum _nzS
      local nz_SD = r(sum) / `K'
      bysort area: egen nz_am = mean(nz)
      bysort area: gen _first = _n == 1
      gen _nzAw = (1 - _Sa/`K') * nz_am             // v1.9.0: the area is compared with pbar, the mean of the same K strata
      quietly sum _nzAw if _first
      local nz_A = r(sum) / `K'
      local V_A_c  = `V_A_s'  - `nz_A'
      local V_SD_c = `V_SD_s' - `nz_SD'
      local V_I_c  = `V_I_s'  + `nz_A' + `nz_SD'
      di "`outcome' (standardised VPC):"
      di "  identity uncorr = " %8.6f (`V_A_s'+`V_SD_s'+`V_I_s') ///
         "   pbar*(1-pbar*) = " %8.6f `den_s'
      di "  VPC_A+SD uncorr = " %5.2f ((`V_A_s'+`V_SD_s')/`den_s'*100) "%"
      di "  VPC_A+SD corr   = " %5.2f ((`V_A_c'+`V_SD_c')/`den_s'*100) "%"
    restore
  }
```

**Weighting standard.** The code gives weight one to each occupied A-SD stratum. In the balanced Malmö design (25 areas, each with the same 12 sociodemographic strata) this coincides with equal weight per area. In unbalanced designs the two differ, and the choice must be declared.

**Three readings of the VPC on the probability scale. Let p_j be the proportion of stratum j and w_j its weight, N_j/N for the crude VPC and 1/K for VPC*, with pbar = Σ w_j p_j. The average risk of the strata of the individuals with the outcome is Σ w_j p_j² / pbar, and that of the individuals without it is Σ w_j p_j (1 − p_j) / (1 − pbar). Their difference is (Σ w_j p_j² − pbar²) / [pbar (1 − pbar)], the between-stratum variance over the total individual variance, which is the VPC. It is the coefficient of discrimination of Tjur (2009). The same ratio is the correlation between the outcomes of two individuals drawn from the same stratum, the intraclass correlation of the binary outcome, and the share of the individual variance that is removed by knowing the stratum. For private GP choice, with the crude weights the two average risks are 30.8% and 19.6%, and their difference, 11.2 points, is the crude VPC of the A-SD strata. With all strata given the same size they are 31.5% and 20.0%, a difference of 11.4 points, the uncorrected VPC*, which the correction for binomial noise brings to 10.7%. For psychotropic medication use the differences are 3.9 points (crude) and 4.1 points (VPC*, uncorrected), and the corrected VPC* is 3.3%. S8 File checks the crude identity by formula on a small example.**

### Step 7b: Contextual Decomposition - CCI, G and CMI (noise-corrected)

**PURPOSE:** Split the between-stratum inequality into the three shares reported in Section 3.2.2 of the paper, which sum to 100%: the contextual clustering (CCI, the between-area share; in this geographical application the CCI), the additive sociodemographic differences (G, the part common to every area), and the Contextual Modification of Inequalities (CMI, the area-by-sociodemographic interaction). All three are standardised and built on the noise-corrected components of Step 7. The command reports them in STEP 6 of its output and returns r(CCI), r(Gshare) and r(CMI) with bootstrap CIs (Step 10).

The three shares are computed on the strata retained by minn() (see Step 7). In Malmö all 300 strata are retained. A flag (!) is printed when a share lies outside its own percentile interval.

The decomposition works on the composition-standardised within-area deviations, Delta_stratum = p_asd - p_a_star (Table A), not on the crude deviations of Step 5. For each sociodemographic group, its average deviation across the 25 areas is the additive part, delta_SD. The departure of a stratum’s actual deviation from this average is the modification, gamma = Delta_stratum - delta_SD. The variance of the delta terms is V_G*, the variance of the gamma terms is V_CMI*, and V_G* + V_CMI* = V_SD* exactly, because the gamma average to zero within each sociodemographic group.

**The exact noise terms.** Each observed stratum proportion carries a binomial sampling variance nz = p_asd(1 - p_asd)/n_asd. The corrections used in Step 7 and here are:

- nz_SD = mean over the A-SD strata of (1 - 1/S_a) x nz, subtracted from V_SD*, where S_a is the number of strata in the area of the stratum (12 in every area here). The within-area deviation of a stratum is taken from the mean of the S_a strata of its own area, so its expected sampling noise is (1 - 1/S_a) x nz rather than (1 - 1/K) x nz, which applies to a deviation from the grand mean of the K strata.
- nz_A = (1/K) x sum over areas of (1 - S_a/K) x the within-area mean of nz, subtracted from V_A*. Each area is compared with the standardised grand mean pbar*, which is the mean of the same K strata, so part of the noise of the area is also in pbar* and cancels. With one stratum per area (S_a = 1) the term is (1 - 1/K) x mean(nz), the term of the two-level designs.
- Both are added back to the individual component, so the identity V_A* + V_SD* + V_I* = pbar*(1 - pbar*) is preserved exactly: the noise is reassigned, not removed.

**The CMI noise floor.** The modification gamma is a single-cell residual, so its variance is inflated by the full sampling noise of its cell, and the uncorrected V_CMI overstates the modification. Under no true modification, the expected value of V_CMI is the binomial floor

floor_CMI = (1 - 1/A) x (1 - 1/S) x mean(nz)

with A = 25 areas and S = 12 sociodemographic strata. The corrected modification variance is V_CMI_c = max(V_CMI - floor_CMI, 0), capped at V_SD_c. The G share is the remainder, so the three shares always sum to 100%:

CCI = V_A_c / (V_A_c + V_SD_c), CMI = V_CMI_c / (V_A_c + V_SD_c), G = (V_SD_c - V_CMI_c) / (V_A_c + V_SD_c)

```
  * --- Contextual decomposition: CCI / G / CMI (standardised) ---
  foreach outcome in psycmed private {
    preserve
      use "dev_asd_`outcome'.dta", clear
      quietly count
      local K = r(N)
      quietly levelsof area, local(al)
      local A : word count `al'
      quietly levelsof strata_sociodem_id, local(sl)
      local S : word count `sl'
      bysort area: egen p_a_star = mean(p_asd)
      quietly sum p_asd
      local pbar = r(mean)
      * corrected components (as in Step 7)
      gen va_s  = (p_a_star - `pbar')^2
      gen vsd_s = (p_asd - p_a_star)^2
      gen nz    = p_asd*(1 - p_asd) / n_asd
      quietly sum va_s
      local V_A_s = r(sum) / `K'
      quietly sum vsd_s
      local V_SD_s = r(sum) / `K'
      quietly sum nz
      local meannz = r(mean)
      bysort area: gen _Sa = _N                   // strata in the area (12 here)
      gen _nzS = (1 - 1/_Sa) * nz                 // within-area noise factor, per stratum
      quietly sum _nzS
      local nz_SD = r(sum) / `K'
      bysort area: egen nz_am = mean(nz)
      bysort area: gen _first = _n == 1
      gen _nzAw = (1 - _Sa/`K') * nz_am             // v1.9.0: the area is compared with pbar, the mean of the same K strata
      quietly sum _nzAw if _first
      local nz_A = r(sum) / `K'
      local V_A_c  = `V_A_s'  - `nz_A'
      local V_SD_c = `V_SD_s' - `nz_SD'
      * additive part (delta) and modification (gamma)
      gen dstar = p_asd - p_a_star
      bysort strata_sociodem_id: egen delta = mean(dstar)
      gen gamma = dstar - delta
      gen vc = gamma^2
      quietly sum vc
      local V_CMI = r(sum) / `K'
      * CMI noise floor and the three shares
      local floor_cmi = (1 - 1/`A') * (1 - 1/`S') * `meannz'
      local V_CMI_c = max(`V_CMI' - `floor_cmi', 0)
      if `V_CMI_c' > `V_SD_c' local V_CMI_c = `V_SD_c'
      local tot = `V_A_c' + `V_SD_c'
      di "`outcome' contextual decomposition (standardised, CMI corrected):"
      di "  CCI clustering   = " %5.1f (`V_A_c'/`tot'*100) "%"
      di "  G   additive     = " %5.1f ((`V_SD_c'-`V_CMI_c')/`tot'*100) "%"
      di "  CMI modification = " %5.1f (`V_CMI_c'/`tot'*100) "%"
    restore
  }
```

**Expected values** (Malmö dataset, B = 2000, seed 2026; CIs from the within-context bootstrap of Step 10). Psychotropic medication use: CCI 4.1% (1.9 to 6.6), G 95.9% (90.7 to 100.0), CMI 0.0% (0.0 to 5.0). Private GP choice: CCI 63.9% (60.5 to 67.3), G 19.0% (16.7 to 21.4), CMI 17.1% (14.3 to 19.9). The same values are returned by the Python reproduction (S5 Text). Stata and numpy generate different bootstrap resamples, so the CIs agree within Monte Carlo error rather than digit for digit.

### Step 8: Discriminatory Accuracy (coAUC and Threshold Classification)

**PURPOSE: Compute the contextual AUC (coAUC) from** observed A-SD stratum proportions and from area proportions alone. Each individual is assigned the proportion of their A-SD stratum as predicted risk. The coAUC equals the probability that a randomly selected case belongs to a higher-risk stratum than a randomly selected non-case. This is the contextual public health AUC, not a clinical diagnostic AUC. coAUC* is the same probability with all strata given the same size, and coAUC*EAR scores every area by its EAR.

Categories for coAUC* and coAUC, as in Table 1 of the paper (Merlo, Wagner and Leckie 2019): 0.50-0.55 Absent | 0.55-0.61 Very small | 0.61-0.66 Small | 0.66-0.72 Moderate | 0.72-0.77 Large | 0.77-1.00 Very large.

The threshold classification uses the population prevalence as the risk threshold (26.0% for psycmed, 22.1% for private GP). Strata above the threshold are high-risk. For each individual: TP (has outcome AND in high-risk stratum), FP (no outcome, high-risk), FN (has outcome, low-risk stratum), TN (no outcome, low-risk). Sensitivity = TP/(TP+FN), Specificity = TN/(TN+FP). This makes Rose’s prevention paradox concrete and quantifiable.

```
  use "records-from-table.dta", clear
  * Thresholds are the population prevalences, taken from the data
  quietly summarize psycmed
  local thresh_psycmed = r(mean)
  quietly summarize private
  local thresh_private = r(mean)

  foreach outcome in psycmed private {
    * Merge A-SD stratum proportions onto individual data
    merge m:1 strata_global_id using "asd_strata_`outcome'.dta", ///
      keepusing(p) nogen
    rename p pred_asd

    * Merge area proportions
    merge m:1 area using "areas_crude_`outcome'.dta", ///
      keepusing(p) nogen
    rename p pred_area

    * AUC from A-SD strata
    quietly roctab `outcome' pred_asd, nograph
    local AUC_asd  = r(area)

    * AUC from areas only
    quietly roctab `outcome' pred_area, nograph
    local AUC_area = r(area)

    di "`outcome':"
    di "  AUC (A-SD strata) = " %5.3f `AUC_asd'  " (" %5.1f (`AUC_asd'*100)  "%)"
    di "  AUC (area only)   = " %5.3f `AUC_area' " (" %5.1f (`AUC_area'*100) "%)"

    * Threshold-based classification
    gen high_risk = (pred_asd >= `thresh_`outcome'')
    quietly count if `outcome'==1 & high_risk==1
    local TP = r(N)
    quietly count if `outcome'==0 & high_risk==1
    local FP = r(N)
    quietly count if `outcome'==1 & high_risk==0
    local FN = r(N)
    quietly count if `outcome'==0 & high_risk==0
    local TN = r(N)
    local Ss = `TP' / (`TP' + `FN')
    local Sp = `TN' / (`TN' + `FP')
    di "  TP=" `TP' "  FP=" `FP' "  FN=" `FN' "  TN=" `TN'
    di "  Sensitivity = " %5.1f (`Ss'*100) "%"
    di "  Specificity = " %5.1f (`Sp'*100) "%"
    drop pred_asd pred_area high_risk
  }
```

### Step 9: Figures

**PURPOSE: Reproduce Figure 4 of the paper and draw the deviations of Step 5 from the crude area proportion.** Figure 4 shows the observed proportion of every one of the 300 A-SD strata with its Wilson 95% confidence interval, grouped by sociodemographic stratum and ordered by proportion within each group, coloured by area income, with a vertical line at the population prevalence. The second figure shows the deviation of every A-SD stratum from the crude proportion of its own area, the deviation tabulated in S2 Text, with the Wilson-shift confidence interval; a filled marker indicates that the interval excludes zero and an open marker that it includes zero. Fig 5 of the paper draws instead the deviation from the composition-standardised level of the area, Δstratum, which averages zero within every area; it is drawn by fig45_prevalence_and_deviations.py in S6 File. Within an area the two deviations differ by the composition gap of the area, so the ordering of the strata is the same. The figures are numbered here to match the paper; Figures 1 to 3 of the paper are conceptual illustrations and are not produced from data. The same two figures are produced automatically by the graph option of the smaihda command.

```
  * ------------------------------------------------------------------
  * Value labels. Without them the axis shows 1..12 instead of the
  * descriptive stratum names used in the paper. Run once on the dataset.
  * ------------------------------------------------------------------
  label define sdlab  1 "W, 35-44, Low"    2 "W, 35-44, High"  ///
                      3 "W, 45-54, Low"    4 "W, 45-54, High"  ///
                      5 "W, 55-64, Low"    6 "W, 55-64, High"  ///
                      7 "M, 35-44, Low"    8 "M, 35-44, High"  ///
                      9 "M, 45-54, Low"   10 "M, 45-54, High"  ///
                     11 "M, 55-64, Low"   12 "M, 55-64, High", replace

  * ----- Figure 2: observed proportion of every A-SD stratum -----
  foreach outcome in psycmed private {

    * threshold taken from the data, not written by hand
    use "records-from-table.dta", clear
    quietly summarize `outcome'
    local thresh = r(mean)*100

    use "asd_strata_`outcome'.dta", clear
    label values strata_sociodem_id sdlab
    quietly replace p  = p*100
    quietly replace lo = lo*100
    quietly replace hi = hi*100

    * one row per sociodemographic stratum; the 25 areas fan out within the
    * row, ordered by proportion, which is what makes 300 intervals readable
    bysort strata_sociodem_id (p): gen long rank = _n
    by strata_sociodem_id: gen long k = _N
    gen double y_pos = strata_sociodem_id
    replace y_pos = strata_sociodem_id + 0.72*((rank-1)/(k-1) - 0.5) if k > 1

    twoway ///
      (rcap lo hi y_pos if area_income==0, horizontal ///
        lcolor("174 199 232") lwidth(vthin)) ///
      (rcap lo hi y_pos if area_income==1, horizontal ///
        lcolor("255 200 150") lwidth(vthin)) ///
      (scatter y_pos p if area_income==0, ///
        mcolor("31 119 180") msymbol(circle) msize(vsmall)) ///
      (scatter y_pos p if area_income==1, ///
        mcolor("255 127 14") msymbol(circle) msize(vsmall)) ///
      , ///
      yscale(reverse) ///
      ylabel(1(1)12, valuelabel angle(0) labsize(vsmall)) ///
      ytitle("") xtitle("Prevalence (%)") ///
      xline(`thresh', lpattern(dash) lcolor(black)) ///
      legend(order(3 "Poor area" 4 "Rich area") pos(4) ring(0) col(1) ///
             size(vsmall) region(lcolor(gs12))) ///
      title("`outcome'", size(medsmall)) ///
      graphregion(color(white))
    graph export "figure2_`outcome'.png", replace width(1600)
  }

  * ----- Figure 3: deviation of every A-SD stratum from its area -----
  foreach outcome in psycmed private {
    use "dev_asd_`outcome'.dta", clear
    label values strata_sociodem_id sdlab
    quietly replace dev    = dev*100
    quietly replace dev_lo = dev_lo*100
    quietly replace dev_hi = dev_hi*100

    bysort strata_sociodem_id (dev): gen long rank = _n
    by strata_sociodem_id: gen long k = _N
    gen double y_pos = strata_sociodem_id
    replace y_pos = strata_sociodem_id + 0.72*((rank-1)/(k-1) - 0.5) if k > 1

    twoway ///
      (rcap dev_lo dev_hi y_pos if area_income==0, horizontal ///
        lcolor("174 199 232") lwidth(vthin)) ///
      (rcap dev_lo dev_hi y_pos if area_income==1, horizontal ///
        lcolor("255 200 150") lwidth(vthin)) ///
      (scatter y_pos dev if sig==1 & area_income==0, ///
        msymbol(circle) mcolor("31 119 180") msize(vsmall)) ///
      (scatter y_pos dev if sig==0 & area_income==0, ///
        msymbol(circle_hollow) mlcolor("31 119 180") msize(vsmall)) ///
      (scatter y_pos dev if sig==1 & area_income==1, ///
        msymbol(circle) mcolor("255 127 14") msize(vsmall)) ///
      (scatter y_pos dev if sig==0 & area_income==1, ///
        msymbol(circle_hollow) mlcolor("255 127 14") msize(vsmall)) ///
      , ///
      yscale(reverse) ///
      ylabel(1(1)12, valuelabel angle(0) labsize(vsmall)) ///
      xline(0, lpattern(dash) lcolor(black)) ///
      ytitle("") ///
      xtitle("{&Delta}{subscript:A,SD} (percentage points)") ///
      legend(order(3 "Poor area, 95% CI excludes 0" ///
                   5 "Rich area, 95% CI excludes 0" ///
                   4 "95% CI includes 0") ///
             pos(4) ring(0) col(1) size(vsmall) region(lcolor(gs12))) ///
      title("`outcome'", size(medsmall)) ///
      graphregion(color(white))
    graph export "figure3_`outcome'.png", replace width(1600)
  }
```

### Step 10: Within-Context Bootstrap for CCI, VPC and coAUC

**PURPOSE:** Estimate 95% confidence intervals for CCI, VPC and coAUC using a non-parametric within-context bootstrap. The areas and the strata are held fixed and the resampling unit is the individual within each A-SD cell. For a binary outcome, resampling the individuals of a cell with replacement is the same as drawing its number of cases from a binomial distribution with the size and the observed proportion of the cell. In each iteration the cases of every cell are drawn in this way and CCI, VPC and coAUC are recomputed from the counts. The interval is the 2.5th and 97.5th percentiles of the B = 2000 bootstrap distribution, recentred on the estimate and truncated to the admissible range of the measure. No distributional assumptions are required beyond those of the observed data.

The bootstrap is the natural uncertainty quantification for measures derived from combinations of stratum proportions across multiple levels, since simple analytical formulas are not available for these quantities. The command works on the table of cells, and the code below does the same step by step. In each replicate one line, rbinomial(), redraws the cases of every cell, with the cells sorted by area and stratum, so the result does not depend on the order of the rows.

```
  use "records-from-table.dta", clear
  local B    = 2000
  local seed = 2026

  * AUC from a table of cells: every case and every non-case of a cell
  * counted once, ties at one half. It is the value roctab returns on
  * the individual records.
  capture program drop auc_cells
  program define auc_cells, rclass
    args score cases noncases
    tempvar cum gsum term
    preserve
    sort `score'
    gen double `cum' = sum(`noncases')
    by `score': egen double `gsum' = total(`noncases')
    by `score': replace `cum' = `cum'[_N]
    gen double `term' = `cases'*(`cum' - `gsum') + 0.5*`cases'*`gsum'
    quietly sum `cases', meanonly
    local tc = r(sum)
    quietly sum `noncases', meanonly
    local tn = r(sum)
    quietly sum `term', meanonly
    return scalar auc = r(sum) / (`tc' * `tn')
    restore
  end

  * The six measures of this step from a table of cells with the area,
  * the size n of each cell and its number of cases.
  capture program drop cell_measures
  program define cell_measures, rclass
    args ev
    tempvar p non na eva pa at vA vSD vI
    gen double `p'   = `ev' / n
    gen double `non' = n - `ev'
    quietly sum n, meanonly
    local N = r(sum)
    quietly sum `ev', meanonly
    local grand = r(sum) / `N'
    bysort area: egen double `na' = total(n)
    by area: egen double `eva' = total(`ev')
    gen double `pa' = `eva' / `na'
    by area: gen byte `at' = (_n == 1)
    gen double `vA'  = `na' * (`pa' - `grand')^2 if `at'
    gen double `vSD' = n * (`p' - `pa')^2
    gen double `vI'  = n * `p' * (1 - `p')
    quietly sum `vA', meanonly
    local V_A = r(sum) / `N'
    quietly sum `vSD', meanonly
    local V_SD = r(sum) / `N'
    quietly sum `vI', meanonly
    local V_I = r(sum) / `N'
    local Vtot = `V_A' + `V_SD' + `V_I'
    return scalar CCI     = `V_A' / (`V_A' + `V_SD')
    return scalar VPC_A   = `V_A' / `Vtot'
    return scalar VPC_SD  = `V_SD' / `Vtot'
    return scalar ICC_ASD = (`V_A' + `V_SD') / `Vtot'
    auc_cells `p' `ev' `non'
    return scalar AUC_asd = r(auc)
    auc_cells `pa' `ev' `non'
    return scalar AUC_area = r(auc)
  end

  foreach outcome in psycmed private {
    di "Bootstrap for `outcome' (B=`B')..."
    preserve

    * ---------------------------------------------------------------
    * 1. The table of cells, one row per A-SD cell, and the point
    *    estimates on the observed counts. They centre the intervals.
    * ---------------------------------------------------------------
    quietly collapse (count) n = `outcome' (sum) ev = `outcome', ///
      by(area strata_sociodem_id)
    gen double p0   = ev / n
    gen double ev_b = ev
    quietly cell_measures ev
    local e1 = r(CCI)
    local e2 = r(VPC_A)
    local e3 = r(VPC_SD)
    local e4 = r(ICC_ASD)
    local e5 = r(AUC_asd)
    local e6 = r(AUC_area)
    matrix est = (`e1', `e2', `e3', `e4', `e5', `e6')

    * ---------------------------------------------------------------
    * 2. Replicates. The contexts stay fixed. The cases of every cell
    *    are redrawn from Binomial(n, p) of the cell, which is the same
    *    as resampling its individuals with replacement. A cell with
    *    p = 0 or p = 1 keeps its count. The cells are sorted by area
    *    and stratum before every draw, so the result does not depend
    *    on the order of the rows.
    *    Columns: CCI, VPC_A, VPC_SD, ICC_ASD, AUC_asd, AUC_area
    * ---------------------------------------------------------------
    matrix boot_res = J(`B', 6, .)
    set seed `seed'
    forval b = 1/`B' {
      sort area strata_sociodem_id
      quietly replace ev_b = rbinomial(n, p0) if p0 > 0 & p0 < 1
      quietly cell_measures ev_b
      matrix boot_res[`b',1] = r(CCI)
      matrix boot_res[`b',2] = r(VPC_A)
      matrix boot_res[`b',3] = r(VPC_SD)
      matrix boot_res[`b',4] = r(ICC_ASD)
      matrix boot_res[`b',5] = r(AUC_asd)
      matrix boot_res[`b',6] = r(AUC_area)
    }
    restore

    * ---------------------------------------------------------------
    * 3. Percentile intervals, recentred on the estimate and truncated.
    *    A replicate carries two doses of binomial noise against one in
    *    the observed proportions, so every dispersion-based measure is
    *    centred above the estimate by one dose while its spread is the
    *    sampling spread.
    * ---------------------------------------------------------------
    preserve
    clear
    svmat boot_res, names(col)
    rename c1 CCI
    rename c2 VPC_A
    rename c3 VPC_SD
    rename c4 ICC_ASD
    rename c5 AUC_asd
    rename c6 AUC_area

    di "=== 95% CI for `outcome' (recentred on the estimate) ==="
    local names "CCI VPC_A VPC_SD ICC_ASD AUC_asd AUC_area"
    forval j = 1/6 {
      local v : word `j' of `names'
      quietly _pctile `v', percentiles(2.5 97.5)
      local q_lo = r(r1)
      local q_hi = r(r2)
      quietly sum `v', meanonly
      local mrep = r(mean)
      local e  = est[1,`j']
      local lo = max(`e' + (`q_lo' - `mrep'), 0)
      local hi = min(`e' + (`q_hi' - `mrep'), 1)
      di "  `v': " %5.2f (`lo'*100) "% to " %5.2f (`hi'*100) "%"
    }
    save "bootstrap_`outcome'.dta", replace
    restore
  }
```

### Step 11: Comparison with random-effects MAIHDA (compare option)

**PURPOSE:** Run the smaihda command with the compare option to fit a standard random-effects MAIHDA via melogit on the same A-SD strata used by the simple-means analysis. The compare option reports nine comparative measures spanning the three components of the MAIHDA framework: five for specific contextual effects (SCE), one for general contextual effects (GCE), and three for discriminatory accuracy (DA). These measures allow the user to evaluate empirically, for any given dataset, the consequences of replacing the observed stratum proportions with their shrunken random-effects counterparts.

Random-effects MAIHDA pulls stratum-specific predictions toward the grand mean by an amount that scales inversely with stratum size. The smallest strata, which often represent the most marginalised subpopulations, are shrunk the most, attenuating the very inequalities the analysis is designed to reveal. The compare option quantifies this attenuation directly on the predictions, on the variance partition, and on the discriminatory accuracy.

The main paper reports six of the nine measures in Table 8 (range compression, maximum absolute attenuation with stratum size n, VPC at A-SD on the probability scale, coAUC at A-SD, and sensitivity and specificity at the population-prevalence threshold), chosen to characterise each MAIHDA component concisely. All nine are reported in Table C at the end of this step.

The two methods are placed on the same probability scale before the VPC is compared. The simple-means VPC is the observed variance decomposition of Step 7. The random-effects VPC is obtained by simulation from the fitted model (Method B of Goldstein, Browne and Rasbash 2002): residuals are drawn from the fitted normal distribution, each is converted to a probability, and the coefficient is formed from the variance of these probabilities Goldstein, Browne and Rasbash set out four procedures. Method A linearises the model with a first-order Taylor expansion. Method B is the simulation just described. Method C treats the binary response as if it were Normal. Method D is the latent variable approach, in which the observed 0/1 response is taken to arise from an underlying continuous variable, and it is Method D that yields the familiar latent-scale variance partition with the level-1 variance fixed at pi-squared over three. We use Method B rather than Method D for the comparison. The authors themselves note that Method B involves no approximation and is simple and fast to compute, and it places both estimators on the probability scale, which is the scale on which the inequality is described.over the sum of that variance and the mean Bernoulli variance, that is v2/(v2+v1)

.

Confidence intervals. The simple-means estimates retain the 95% confidence intervals computed in Step 10. The random-effects estimates produced by compare are single point estimates without bootstrap intervals.

```
  * ============================================================
  * (a) Standard invocation of the compare option, on the table
  * ============================================================
  import delimited using "dataset-smaihda_stratum_table.csv", clear

  smaihda cases if outcome == "psycmed", area(area) ///
    sdstratum(strata_sociodem_id) areacov(area_income) n(n) compare

  smaihda cases if outcome == "private", area(area) ///
    sdstratum(strata_sociodem_id) areacov(area_income) n(n) compare
```

The comparative output of compare appears in the Stata console below the standard smaihda output for each outcome, organised in three sub-blocks: per-stratum predictions, probability-scale variance partition, and discriminatory accuracy.

```
* ============================================================
* (b) What compare does internally: the same logic in plain Stata,
*     equivalent to the implementation inside smaihda.ado (S1 File)
* ============================================================
use "records-from-table.dta", clear

foreach outcome in psycmed private {

  * Population prevalence as classification threshold
  quietly sum `outcome'
  local thresh = r(mean)
  local grand  = r(mean)
  local N      = r(N)

  * A-SD stratum identifier
  egen asd_id = group(area strata_sociodem_id)

  * (i) Fit random-effects MAIHDA on A-SD strata
  melogit `outcome' || asd_id:

  * (ii) RE VPC at A-SD on the PROBABILITY scale, by simulation (Method B)
  *      Goldstein, Browne and Rasbash (2002)
  *      Robust extraction of the RE standard deviation across Stata versions.
  *      Stata 17+ stores the variance directly (var(_cons)); older Stata
  *      stores the log-SD as lns1_1_1. We use estat icc as primary method.
  local b0     = _b[`outcome':_cons]
  local _pi2_3 = (c(pi)^2) / 3
  capture quietly estat icc
  if _rc == 0 {
      local _icc2 = r(icc2)
      if `_icc2' < 1 {
          local sd_re = sqrt(`_icc2' * `_pi2_3' / (1 - `_icc2'))
      }
      else {
          local sd_re = 0
      }
  }
  else {
      capture local sd_re = exp([lns1_1_1]_cons)
      if _rc local sd_re = 0
  }
  preserve
    clear
    set obs 100000
    set seed 90125
    gen u  = rnormal(0, `sd_re')
    gen pi = invlogit(`b0' + u)
    gen vi = pi*(1-pi)
    quietly sum pi
    local v2 = r(Var)
    quietly sum vi
    local v1 = r(mean)
    local VPC_re = `v2' / (`v2' + `v1')
  restore

  * (ii-bis) SM VPC at A-SD on the probability scale (observed decomposition)
  preserve
    collapse (count) n_area=`outcome' (sum) ev_area=`outcome', by(area)
    gen p_area     = ev_area / n_area
    gen va_contrib = n_area * (p_area - `grand')^2
    quietly sum va_contrib
    local V_A = r(sum) / `N'
  restore
  preserve
    collapse (count) n_asd=`outcome' (sum) ev_asd=`outcome', by(asd_id area)
    gen p_asd = ev_asd / n_asd
    bysort area: egen n_ar = total(n_asd)
    bysort area: egen ev_ar = total(ev_asd)
    gen p_ar = ev_ar / n_ar
    gen vsd_contrib = n_asd * (p_asd - p_ar)^2
    quietly sum vsd_contrib
    local V_SD = r(sum) / `N'
    gen vi_contrib = n_asd * p_asd * (1 - p_asd)
    quietly sum vi_contrib
    local V_I = r(sum) / `N'
  restore
  local VPC_sm = (`V_A' + `V_SD') / (`V_A' + `V_SD' + `V_I')

  * (iii) Predictions: shrunken (RE, probability scale) and observed (SM)
  predict p_re, mu
  bysort asd_id: egen p_sm = mean(`outcome')

  * (iv) AUC at A-SD level, both sides
  roctab `outcome' p_re, nograph
  local AUC_re = r(area)
  roctab `outcome' p_sm, nograph
  local AUC_sm = r(area)

  * (v) Sensitivity and specificity at population-prevalence threshold
  gen byte high_risk_re = (p_re >= `thresh')
  gen byte high_risk_sm = (p_sm >= `thresh')
  quietly sum high_risk_re if `outcome' == 1
  local Sens_re = r(mean)
  quietly sum high_risk_sm if `outcome' == 1
  local Sens_sm = r(mean)
  quietly sum high_risk_re if `outcome' == 0
  local Spec_re = 1 - r(mean)
  quietly sum high_risk_sm if `outcome' == 0
  local Spec_sm = 1 - r(mean)

  * (vi) Per-stratum predictions and attenuation summaries
  preserve
    bysort asd_id: keep if _n == 1
    quietly sum p_sm
    local SM_range = r(max) - r(min)
    local SM_sd    = r(sd)
    quietly sum p_re
    local RE_range = r(max) - r(min)
    local RE_sd    = r(sd)
    local Range_compr = 100 * `RE_range' / `SM_range'
    gen att = abs(p_sm - p_re)
    quietly sum att
    local Mean_att = r(mean)
    local Max_att  = r(max)
  restore

  di "==================== `outcome' ===================="
  di "SCE:"
  di "  Range compression (RE/SM)   = " %5.1f `Range_compr' "%"
  di "  Mean absolute attenuation   = " %6.4f `Mean_att'
  di "  Maximum absolute attenuation= " %6.4f `Max_att'
  di "GCE:"
  di "  VPC at A-SD (SM, prob scale)= " %5.2f (`VPC_sm'*100) "%"
  di "  VPC at A-SD (RE, prob scale)= " %5.2f (`VPC_re'*100) "%"
  di "DA:"
  di "  AUC at A-SD (SM)            = " %5.3f `AUC_sm'
  di "  AUC at A-SD (RE)            = " %5.3f `AUC_re'
  di "  Sensitivity (SM / RE)       = " %5.3f `Sens_sm' " / " %5.3f `Sens_re'
  di "  Specificity (SM / RE)       = " %5.3f `Spec_sm' " / " %5.3f `Spec_re'

  drop asd_id p_re p_sm high_risk_re high_risk_sm
  }
```

**Table C.** The nine measures of the *compare* output, simple-means (S) and random-effects (RE), Malmö 2006. The main paper (Table 8) reports six of them; the remaining three are given here in full.

| # | Measure (component) | Psychotropic medication | Private GP choice |
| --- | --- | --- | --- |
| 1 | Range of stratum predictions, S (SCE) | 41.98 pp (9.43% to 51.41%) | 66.59 pp (3.52% to 70.10%) |
|  | Range of stratum predictions, RE (SCE) | 34.88 pp (13.32% to 48.20%) | 62.78 pp (4.60% to 67.38%) |
| 2 | Range compression (RE/S, %) (SCE) | 83.09 | 94.28 |
| 3 | SD of stratum predictions, S (SCE) | 8.85 pp | 14.17 pp |
|  | SD of stratum predictions, RE (SCE) | 7.23 pp | 13.34 pp |
| 4 | Mean absolute attenuation (SCE) | 1.32 pp | 0.72 pp |
| 5 | Maximum absolute attenuation (stratum size) (SCE) | 6.44 pp (n = 75) | 4.14 pp (n = 44) |
| 6 | VPC at A-SD, probability scale, standardised, uncorrected (%): S / RE (GCE) | 4.06 / 3.28 | 11.44 / 9.76 |
|  | VPC at A-SD, probability scale, standardised, corrected for binomial noise (%): S / RE (GCE) | 3.30 / 3.28 | 10.72 / 9.76 |
| 7 | coAUC at A-SD (%): S / RE (DA) | 62.7 / 62.7 | 71.8 / 71.8 |
| 8 | Sensitivity at the population-prevalence threshold (%): S / RE (DA) | 59.7 / 57.9 | 66.5 / 65.5 |
| 9 | Specificity at the population-prevalence threshold (%): S / RE (DA) | 58.3 / 60.1 | 65.7 / 66.6 |

Notes. pp, percentage points. The S values are computed from the observed stratum proportions and are reproduced independently by the Python program of S5 Text. All RE values, the compression factor and the attenuation measures are from the compare option of the *smaihda* command. Standard deviations follow Stata’s sample convention (n − 1); the population formula gives 8.83 and 14.15 pp.

When the compare option is used, the command contrasts the random-effects VPC against both standardised simple-means VPCs, uncorrected and corrected. VPC*, the standardised VPC, represents the general contextual effect independent of population composition, and the RE-MAIHDA tau^2 treats strata as units, so it is the natural comparator. The uncorrected form is the observed reality, which in a total-population register is not a sample estimate. The corrected form puts the simple-means VPC on the same noise-discounted footing as the random-effects tau^2.
