## Supplementary material for "Describing health inequalities without distortion: Simple-Means MAIHDA vs Random-Effects MAIHDA": S1 File. The smaihda Stata package: smaihda.ado

```
*! version 1.9.0   06oct2026
*! 06oct2026 (v1.9.0): coAUC*EAR, the area-level contextual AUC
*!   standardised for size and composition. The area-level coAUC* (auc_area_std)
*!   gives every area the same weight but scores it by its crude proportion, which
*!   carries the composition of the area. coAUC*EAR scores every area by its
*!   composition-standardised prevalence, the EAR, the plain mean of the
*!   proportions of its cells with n >= minn, and counts every area once. It is the
*!   coAUC with every cell given the same size, read at the area level, and the
*!   discrimination-scale counterpart of the area VPC*, which is also computed from
*!   the EAR. It has its own within-context bootstrap interval; the random stream is
*!   unchanged, so no other interval moves. The output labels follow the terminology
*!   of the papers: coAUC (crude contextual AUC, discriminatory accuracy), coAUC*
*!   (standardised contextual AUC, general contextual effect) and coAUC*EAR. Nothing
*!   else changes.
*!   Stored results: r(AUC_area_ear), r(AUC_area_ear_lo), r(AUC_area_ear_hi).
*! version 1.8.1   06oct2026
*! 06oct2026 (v1.8.1): noise term of the standardised between-area
*!   component. The area mean of the S_a cells of an area is compared with the grand
*!   mean p*, which is itself the mean of the same K cells, so the expected binomial
*!   noise of V_A* is the sum over areas of (1 - S_a/K)*mean_a(nz)/K, not
*!   sum_a mean_a(nz)/K. The old term ignored that p* is estimated from the same
*!   cells and removed about 1/A of the noise too much (A = number of areas). The
*!   same reasoning gave the factor (1 - 1/S_a) of the within-area component in
*!   v1.5.1, and with one cell per area (S_a = 1) the new term is (1 - 1/K)*mean(nz),
*!   the term of the two-level designs. Verified by binomial simulation in balanced
*!   and unbalanced designs. Only V_A_c and what is computed from it move (corrected
*!   area VPC*, corrected A-SD VPC*, corrected CCI, and the CCI/G/CMI shares); the
*!   identity is unchanged, and no crude or uncorrected measure, AUC or
*!   classification changes.
*! version 1.8.0   26sep2026
*! 26sep2026 (v1.8.0): the command reads a stratum table.
*!   With the new option n(), the data in memory are a table with one row per
*!   stratum: the outcome variable holds the number of cases and n() the number
*!   of individuals of each row. Use -if- to select one outcome when the table
*!   holds several. The command expands the table to individual records inside
*!   -preserve- and runs as before, so the results are those of the records by
*!   construction and the data in memory do not change. Without n() it reads
*!   individual records, as before. The analysis is now the program
*!   _smaihda_run; smaihda is a wrapper that handles n(). The output banner
*!   prints an "Input" line when the data are a table. The Python and R
*!   implementations, also version 1.8.0, read the same table.
*! version 1.7.0   26sep2026
*! 26sep2026 (v1.7.0): the within-context bootstrap works on the table of cells.
*!   The contexts stay fixed, as since v1.6.0. In each replicate the number of
*!   cases of every A-SD cell is drawn from Binomial(n, p) of the cell, which
*!   is the same experiment as resampling its individuals with replacement,
*!   and every measure is recomputed from the counts of the cells by the new
*!   helper _smaihda_compute_cells, with the formulas of _smaihda_compute.
*!   This is the mechanism of the Python and R implementations. The cells are
*!   sorted by area and stratum before every draw, so the interval no longer
*!   depends on the order of the rows: the individual records and the stratum
*!   table expanded with -expand- give the same limits. It is also faster.
*!   The size-standardised area AUC of a replicate is formed from the cells
*!   with n >= minn(), as its point estimate is; the replicate used every cell
*!   before. Nothing changes where no cell is below minn().
*!   No point estimate changes. The limits move within Monte Carlo error.
*!   Stata, Python and R draw different random numbers, so their limits agree
*!   within Monte Carlo error, not digit for digit.
*!   The output banner printed v1.5.2; it now prints v1.7.0.
*! version 1.6.2   24sep2026  EXECUTED IN STATA by Raquel Perez-Vicente, 26sep2026
*! 24sep2026 (v1.6.2): the size-standardised AUC now obeys minn().
*!   One rule, no exceptions: a measure that counts cells applies minn(), a
*!   measure that counts individuals uses them all. AUC_asd_s and AUC_area_s
*!   count cells, so they now use the cells with n >= minn, as the standardised
*!   partition and the CCI/G/CMI shares already did. AUC_asd, AUC_area and the
*!   threshold classification count individuals and keep every cell, because
*!   excluding a cell there would delete real people from a statement about the
*!   population.
*!   Nothing changes on the Malmo dataset (smallest cell 35). It matters in
*!   sparse intersectional designs, which is where MAIHDA is most used: on a
*!   secondary ESS design with 16 cells below ten individuals, the standardised
*!   AUC falls from 81.41% to 80.08% while the crude AUC moves by 0.03 points.
*! version 1.6.1   21sep2026  executed in Stata as part of 1.6.2
*! 22sep2026 (v1.6.1, patch): the global macro _smaihda_flag is renamed smaihda_flag
*!   (Raquel Perez-Vicente, from her Stata run of v1.6.0). In Stata a global whose name
*!   begins with an underscore is really a local ([P] macro), so the flag set inside
*!   _smaihda_show_row never reached the main program and the "(!)" footnote was never
*!   printed. No computation changes. Her run also confirmed the compare block of v1.6.0
*!   in Stata: standardised VPC, psycmed 4.057 / 3.293 vs RE 3.277 and private
*!   11.440 / 10.720 vs RE 9.764, identical to the Python and R twins.
*! 21sep2026 (v1.6.1): the size-standardised AUC.
*!   Two new measures, AUC_asd_s and AUC_area_s, displayed as "AUC std size".
*!   Every cell contributes the same total weight (cases = p, non-cases = 1-p),
*!   so the result is the AUC that would be observed if all cells were of equal
*!   size, exactly as the standardised VPC counts every cell once. The ordinary
*!   AUC counts pairs of individuals, so a cell of 3000 people contributes a
*!   million times more pairs than a cell of 3 and the value depends on how the
*!   population happens to be spread over the cells.
*!   It is the same kind of object as an age-standardised rate: real arithmetic
*!   on a hypothetical population of equal-sized cells, and it is read as such.
*!   It needs no continuity correction, since only the ordering of the
*!   proportions is used and a cell with no events has a well defined rank.
*!   Computed by the new helper _smaihda_auc_eq, with ties at one half as in
*!   -roctab-, and carried through the within-context bootstrap like every
*!   other measure (boot_res now has 15 columns).
*!   Nothing else changes. No existing VPC, CCI, AUC, classification, share or
*!   interval moves. Expected values on the Malmo dataset from the Python and R
*!   twins v1.6.2 (B = 2000, seed 2026), to be confirmed in Stata:
*!     psycmed  AUC 62.71%; AUC std size, strata 62.97%; area 52.97% / 53.22%
*!     private  AUC 71.77%; AUC std size, strata 71.90%; area 67.90% / 68.38%
*! version 1.6.0   20sep2026  EXECUTED IN STATA by Raquel Perez-Vicente, 22sep2026:
*!   the compare block reproduced the Python and R twins exactly, and her run found
*!   the underscore-global bug patched above. The AUC std size of v1.6.1 and the
*!   minn() rule of v1.6.2 were confirmed with 1.6.2 on 26sep2026.
*! 20sep2026 (v1.6.0): the bootstrap keeps the contexts fixed.
*!   The areas and the strata are the observed set, not a sample of exchangeable
*!   contexts, so resampling whole areas would impose the very assumption that
*!   Simple-Means MAIHDA declines in the estimation. The default bootstrap now
*!   holds the contexts fixed and resamples individuals with replacement inside
*!   each A-SD cell (-bsample, strata()-).
*!   A replicate is drawn from proportions that already carry one dose of
*!   binomial noise, so it carries two, and every measure that depends on the
*!   dispersion of the proportions is centred above the estimate by one dose
*!   while its spread is the sampling spread. The reported interval is therefore
*!   the percentile interval recentred on the estimate and truncated to the
*!   admissible range of the measure, which is also the boundary rule already
*!   applied to the corrected components since v1.4.1.
*!   No point estimate changes. The intervals do, and the area-level ones narrow
*!   appreciably, since the variability of which areas were observed no longer
*!   enters them. Expected values on the Malmo dataset (B = 2000, seed 2026),
*!   from the Python twin v1.6.0, to be confirmed within Monte Carlo error:
*!     psycmed  VPC/ICC_A+SD std corr 3.29% (2.88 to 3.71); CCI 4.1% (1.8 to 6.6);
*!              G 95.9% (90.7 to 100.0); CMI 0.0% (0.0 to 5.0); AUC 62.71%
*!              (62.17 to 63.25)
*!     private  VPC/ICC_A+SD std corr 10.72% (10.08 to 11.37); CCI 63.9% (60.5 to
*!              67.3); G 19.0% (16.7 to 21.4); CMI 17.1% (14.3 to 19.9); AUC
*!              71.77% (71.23 to 72.31)
*! version 1.5.2   15sep2026
*! 09sep2026: output labels renamed. GCCI is now "CCI, crude" (every individual counts once) and
*!   the standardised GCCI is "CCI, uncorr / corr" (every cell counts once). Same quantities, same
*!   r() names (r(GCCI), r(GCCI_s), r(GCCI_c) kept for compatibility). "size-weighted" -> "crude",
*!   "size-standardised" -> "standardised" in the display. Version-history notes below keep the old words.
*! (previous: 1.5.1 11sep2026; 1.5.0 09sep2026; 1.4.1 31aug2026; 1.4.0 16aug2026)
*! Distributed with preprint version V.2 of:
*!   Merlo J, Bashir NZ, Rodriguez-Lopez M, Khalaf K, Oberg J, Perez-Vicente R.
*!   Describing health inequalities without distortion:
*!   Simple-Means MAIHDA vs Random-Effects MAIHDA.
*! smaihda - Simple-means MAIHDA for binary outcomes (two- and three-level designs)
*!
*! Companion to: Merlo J. et al. "Describing health inequalities without
*! distortion: Simple-Means MAIHDA vs Random-Effects MAIHDA." Preprint V.2.
*!
*! Implements Steps 1 to 11 of S1 Text of the companion paper
*! as a single user-callable command.
*!
*! CHANGELOG
*! 1.5.2 (15sep2026): Noise term of the standardised within-area component.
*!      In the three-level design the within-area deviation of a cell is taken
*!      from the mean of the S_a cells of its own area, so its expected binomial
*!      noise is (1 - 1/S_a)*nz per cell, not (1 - 1/K)*nz. nz_SD is now the
*!      mean over cells of (1 - 1/S_a)*nz (S_a = standardised cells in the
*!      area; 12 in Malmo). (1 - 1/K)*mean(nz) is kept for the 2-level designs,
*!      where the deviation is from the grand mean of the K cells. Verified by
*!      binomial simulation. Effect in Malmo: V_SD_c rises by about 1 point^2;
*!      CCI/G/CMI move by at most 0.5 points (private GP 64.3/18.5/17.2 ->
*!      63.9/19.0/17.1); V_I_c falls by the same amount; identity unchanged.
*!      Nothing else changes.
*! 1.5.1 (11sep2026): Figure export only. The PNG width goes from 1600 to
*!      2244 pixels, the width of a full-page figure at 300 dpi, and the
*!      command now also writes a vector EPS beside each PNG. No measure,
*!      no computation and no displayed value changes.
*! 1.5.0 (07sep2026): Sparse-design hardening (includes the 1.4.1 rule)
*!    - Step 4 deviation interval by Wilson shift: the cell's Wilson interval
*!      (Step 1) moved by the area proportion, which is treated as known.
*!      The delta-method interval had zero width at p = 0 or p = 1, so every
*!      degenerate cell was flagged as significantly deviating.
*!    - Step 2 EAR standard error from the Wilson interval width of each cell
*!      (zero under the old formula for degenerate cells).
*!    - New option minn(#), default 10: the size-standardised partition, the
*!      size-standardised GCCI and the CCI / G / CMI shares use the cells with
*!      n >= #. With many tiny cells these measures are dominated by binomial
*!      noise and no noise term identifies the components (simulation,
*!      smaihda_sparse_fix.py). Size-weighted measures, AUC, classification,
*!      Steps 1, 2 and 4 are always computed on all cells. minn(1) reproduces
*!      v1.4.1 exactly. New returns r(K_all), r(K_std), r(N_excl).
*!    - Cell-size diagnostic printed in Step 1: median and minimum n; cells
*!      and individuals with n < 5, < 10, < 30; cells with no events and
*!      with all events. The basis of the stratum-design rule.
*!    - Note printed when the area x stratum grid is incomplete (three-level).
*!    - Flag (!) on any summary row whose percentile interval excludes its
*!      point estimate, with a note on cluster-bootstrap bias.
*!    - Summary table columns moved from 28/45 to 30/47 (label overflow).
*!    - Malmo canonical run: all Step 6 values unchanged; Step 4 significant
*!      deviations 137 -> 125 (psycmed) and 134 -> 122 (private), the values
*!      of the Python reference implementation.
*! 1.4.1 (31aug2026): Boundary rule for sparse information
*!    - Corrected variance components floored at zero: a variance cannot
*!      be negative; a negative corrected value is the measurement error
*!      of a true zero when the binomial-noise estimate exceeds the raw
*!      between component (rare outcomes / very small strata).
*!    - GCCI_c and the CCI/G/CMI shares returned as missing when the
*!      corrected between-stratum variance is zero: with no detectable
*!      clustering beyond noise, the shares of that clustering are not
*!      defined (0/0). Previously a meaningless ratio could be printed.
*!    - Applied identically in _smaihda_compute and the bootstrap
*!      compute. No number changes for any dataset in which the
*!      corrected components are positive (Malmo canonical run
*!      unaffected).
*! 1.4.0 (16aug2026): Contextual decomposition (CCI / G / CMI)
*!    - New: the between-stratum variance is split into three shares that sum
*!      to 100%: CCI (between-area clustering, = r(GCCI_c), the corrected
*!      G (the additive sociodemographic differences, the same in every area),
*!      and CMI, the Contextual Modification of Inequalities, i.e. the area-by-
*!      sociodemographic interaction (how much the area reshapes them).
*!      CMI is noise-corrected: gamma_A,SD is a single-cell residual, so its
*!      variance is inflated by binomial sampling noise; the expected floor
*!      (1-1/A)(1-1/S)*mean(p(1-p)/n) is subtracted (A areas, S SD strata).
*!      Three-level design only; suppressed (missing) in the two-level design.
*!    - New returns: r(CCI), r(Gshare), r(CMI) with _lo/_hi cluster-bootstrap
*!      percentile limits. No existing VPC, GCCI, AUC or compare value changes.
*!    - Default seed is now 2026 (canonical; all CIs in the manuscript use it).
*!      Stata and numpy generate different resamples, so the bootstrap CIs
*!      should match the Python (S5 Text) values approximately, not digit for digit.
*!    - Expected values (Malmo, B=2000, seed=2026, from the Python implementation, S5 Text), for
*!      Merida/Raquel to confirm within Monte Carlo error:
*!      psycmed  CCI~4.1%  G~95.9% CMI~0.0% (CI 0.0-2.9, includes 0);
*!      private  CCI~64.3% G~18.5% CMI~17.2% (CI 9.3-26.2, excludes 0).
*! 1.3.1 (14aug2026): Classification counts and PPV at the threshold
*!    - The threshold classification now reports the four cell counts
*!      (TP, FP, FN, TN), the number of strata flagged high-risk, and the
*!      positive predictive value (PPV), on the simple-means side and,
*!      under COMPare, on the random-effects side. The counts were already
*!      computed internally to form sensitivity and specificity; this
*!      exposes them in the output and the returns.
*!    - New returns: r(ppv), r(n_flag), r(TP), r(FP), r(FN), r(TN), and
*!      their random-effects counterparts r(PPV_RE), r(TP_RE), r(FP_RE),
*!      r(FN_RE), r(TN_RE). Sensitivity, specificity, AUC, VPC, GCCI and
*!      all compare values are unchanged.
*! 1.3.0 (12aug2026): Support for the two-level design
*!    - area() and sdstratum() are now both optional; at least one is
*!      required. The command serves two designs:
*!        three-level  area() + sdstratum()  (A-SD strata within areas; also
*!                     reports the GCCI). The multidimensional design of this
*!                     paper.
*!        two-level    either variable alone (individuals within contexts). A
*!                     single analysis that serves institutional/geographical
*!                     MAIHDA (contexts = areas, hospitals, regions) and
*!                     intersectional MAIHDA (contexts = intersectional strata)
*!                     identically; they differ only in what the contexts mean.
*!    - In the two-level design the GCCI is not defined and is suppressed,
*!      the output collapses to a single VPC/ICC (size-weighted and
*!      size-standardised, uncorrected and corrected), and the cluster
*!      bootstrap resamples the single contextual level (areas in the
*!      geographical design, strata in the intersectional design).
*!    - The binomial-noise correction is made design-aware: a single
*!      between-context term (1 - 1/K)*mean(noise) in the 2-level designs,
*!      the area/stratum split in the three-level design.
*!    - compare works unchanged in all three designs; melogit is fitted on
*!      the single contextual level.
*!    - No three-level result changes.
*! 1.2.0 (11aug2026): Size-standardised variance partition and compare
*!    - New: size-standardised VPC and GCCI, in which every occupied A-SD
*!      stratum counts once (weight 1 per stratum), reported ALONGSIDE the
*!      size-weighted (demographic) VPC, not in its place. Two forms are
*!      shown: uncorrected, and corrected for binomial noise. The
*!      correction reassigns the sampling component from the between-stratum
*!      levels to the individual level, so the identity
*!      V_A* + V_SD* + V_I* = pbar*(1-pbar*) is preserved exactly.
*!      Weighting standard: weight 1 per occupied A-SD stratum. In balanced
*!      designs this coincides with equal weight per area; in unbalanced
*!      designs the two differ and the choice must be declared.
*!    - The size-standardised VPC represents the general contextual effect
*!      (GCE) independent of population composition. The size-weighted VPC,
*!      which tracks the discriminatory accuracy, is retained for reference.
*!    - COMPare now contrasts the random-effects VPC against BOTH
*!      size-standardised simple-means VPCs, uncorrected and corrected.
*!      Rationale: tau^2 of RE-MAIHDA treats strata as units, so the
*!      size-standardised VPC is its natural comparator; the corrected form
*!      sits on the same noise-discounted footing as tau^2, while the
*!      uncorrected form is the observed reality, which in a total-population
*!      register is not a sample estimate.
*!    - New returns: r(ICC_ASD_s), r(ICC_ASD_c), r(GCCI_s), r(GCCI_c) and
*!      their _lo/_hi cluster-bootstrap percentile limits; r(VPC_A_s),
*!      r(VPC_SD_s), r(VPC_A_c), r(VPC_SD_c).
*!      No size-weighted VPC, GCCI, AUC or compare value changes.
*! 1.0.9 (17jun2026): GCCI naming made consistent (and a CI fix)
*!    - The GCCI confidence-interval results are now returned as
*!      r(GCCI_lo) and r(GCCI_hi), matching the point estimate r(GCCI).
*!      They were previously named r(GCI_lo) and r(GCI_hi).
*!    - This also fixes a name mismatch: the cluster bootstrap stored the
*!      GCCI percentile CI in locals GCCI_lo/GCCI_hi, but the display and
*!      the returned scalars read GCI_lo/GCI_hi, which stayed missing. So
*!      the GCCI CI was previously shown and returned as missing. The
*!      display and the returns now read the locals the bootstrap fills.
*!    - The typed option keyword is still gci, kept for back-compatibility;
*!      only the printed label and the returned scalar names use GCCI.
*! 1.0.8 (17jun2026): Return the absolute variance components
*!    - _smaihda_compute now returns V_A, V_SD and V_I, the three
*!      probability-scale variance components whose sum is p(1-p).
*!      Up to 1.0.7a only the ratios (VPC_A, VPC_SD, ICC_ASD) were
*!      returned, so the user had to rebuild the components by hand.
*!      They are now available as r(V_A), r(V_SD) and r(V_I), which
*!      makes the decomposition explicit and the identity
*!      V_A + V_SD + V_I = p(1-p) a one-line check. (Suggested by
*!      N. Bashir.) The command also prints the three components and
*!      the identity check, mirroring S1 Text Step 7. No computed VPC,
*!      GCCI, AUC, or comparison value changes.
*! 1.0.7a (16jun2026): Housekeeping after the probability-scale VPC move
*!    - Removed the now-unused pi2_3 local and its argument to the two
*!      helper programs. The probability-scale VPC does not need a
*!      level-1 latent variance. (pi^2/3 is still used, only inside the
*!      compare block, to recover tau^2 from the latent ICC returned by
*!      estat icc; that local is named _pi2_3 and is unaffected.)
*! 1.0.7 (16jun2026): VPC moved to the probability scale
*!    - The variance partition coefficient is now computed on the
*!      probability scale, as a correlation ratio via the law of total
*!      variance (VA + VSD + VI = p(1-p)), so it sits on the same scale
*!      as GCCI and matches the manuscript and S3 Text. Versions up to 1.0.6
*!      reported the latent logit-scale VPC (logit variance components
*!      with a pi^2/3 level-1 variance), a different quantity.
*!    - The method-of-moments (Taylor) sampling-variance correction was
*!      removed from the descriptive VPC; that estimand (latent process
*!      variance) is treated separately (Box 2).
*!    - In COMPare, the random-effects VPC is now obtained by simulation
*!      on the probability scale (Method B of Goldstein, Browne and
*!      Rasbash 2002): u ~ N(0, tau^2), pi = invlogit(b0 + u), and
*!      VPC = var(pi) / (var(pi) + mean(pi(1-pi))). Previously it used
*!      the latent formula tau^2 / (tau^2 + pi^2/3) (their Method C).
*!      The simulation seed is saved and restored, so there is no side
*!      effect on the user's RNG state.
*!    - Fix applied in both _smaihda_compute and _smaihda_compute_boot;
*!      the bootstrap CIs for VPC_A and VPC_SD are now probability-scale.
*!    - Display label changed from "Latent-scale variance partition" to
*!      "Probability-scale variance partition".
*! 1.0.6 (10jun2026): Version-robust random-effects variance extraction
*!    - The COMPare option now reads the melogit between-stratum variance
*!      with a cascade: first the direct variance parameter var(_cons) in
*!      the level equation (Stata 17+), then the legacy log-SD parameter
*!      lns1_1_1 (older Stata), then a fallback to zero (variance on the
*!      boundary). Version 1.0.5 read only lns1_1_1, which is absent in
*!      Stata 17+, so the random-effects ICC was wrongly reported as zero
*!      on those versions. (Reported by Raquel Perez-Vicente.)
*! 1.0.5 (10jun2026): Boundary-safe random-effects variance extraction
*!    - In the COMPare option, the latent-scale random-effects VPC is now
*!      read from the melogit variance with a capture and a fallback to
*!      zero variance. When the between-stratum variance converges to the
*!      zero boundary, the variance parameter is not stored, and estat icc
*!      then raised "lns1_1_1 not found". The command now treats this case
*!      as VPC = 0 and continues. (Reported by Raquel Perez-Vicente.)
*! 1.0.4 (12may2026): Attenuation measures and display fix
*!    - The COMPare option now reports the empirical attenuation of
*!      stratum-specific predictions: range of SM and RE predictions,
*!      range compression (RE range as % of SM range), mean absolute
*!      attenuation, and max absolute attenuation (with the size of
*!      the stratum where shrinkage was largest). These quantify
*!      shrinkage directly even when AUC is invariant to it (AUC is
*!      invariant under monotonic shrinkage, which is approximately
*!      what occurs with uniform stratum sizes).
*!    - Sensitivity and 1-specificity at the population prevalence
*!      threshold are reported for the RE side as well.
*!    - Linesize is set to 120 columns at the start of the command
*!      and restored at the end, so Step 2 and Step 3 tables render
*!      in a single row regardless of the user's current linesize.
*! 1.0.3 (11may2026): Random-effects MAIHDA comparison
*!    - New option COMPare: when specified, additionally fits a
*!      standard random-effects MAIHDA (melogit on A-SD strata)
*!      and displays its VPC and AUC alongside the simple-means
*!      estimates. Integrates rather than replaces the random-
*!      effects approach: the two estimates should agree on overall
*!      contextual structure but differ in stratum-specific
*!      predictions, where shrinkage matters. The RE comparison
*!      uses a single point estimate (no bootstrap) to keep runtime
*!      manageable; simple-means bootstrap CIs are still shown.
*! 1.0.2 (11may2026): VPC bug fix for sparse A-SD cells
*!    - Haldane-Anscombe continuity correction applied to cell
*!      proportions before logit transformation
*!    - Taylor sampling-variance correction reweighted to match
*!      V_obs weighting (was unweighted mean, now sum/N)
*!    - Fix applied in both _smaihda_compute and _smaihda_compute_boot
*! 1.0.1 (10may2026): Bug fixes
*!    - Option SAVEdir(): macro reference fixed (was `save', should be
*!      `savedir' throughout)
*!    - _smaihda_show_row: replaced invalid `=' calls with precomputed
*!      locals
*!    - ICC bootstrap-CI macros renamed for consistency (ICC_AS_lo /
*!      ICC_AS_hi everywhere)
*!    - Removed dead preserve/restore block in _smaihda_compute
*! 1.0.0 (10may2026): Initial version

* ==================================================================
* smaihda (v1.8.0). Without n() the data are individual records and the
* analysis runs on them. With n() the data in memory are a stratum table,
* one row per stratum, with the number of cases in the outcome variable and
* the number of individuals in n(). The table is expanded to individual
* records inside -preserve-, so the results are those of the records by
* construction and the data in memory do not change. The analysis itself
* is _smaihda_run.
* ==================================================================
program define smaihda, rclass
    version 14.0
    syntax varname(numeric) [if] [in] [, N(varname numeric) *]
    if "`n'" == "" {
        _smaihda_run `varlist' `if' `in', `options'
        return add
        exit
    }
    marksample touse
    markout `touse' `n'
    quietly count if `touse'
    if r(N) == 0 {
        display as error "no observations meet the if/in restrictions"
        exit 2000
    }
    capture assert `n' == int(`n') & `n' >= 0 if `touse'
    if _rc {
        display as error "n(`n') must hold non-negative integers"
        exit 459
    }
    capture assert `varlist' == int(`varlist') & `varlist' >= 0 & `varlist' <= `n' if `touse'
    if _rc {
        display as error "`varlist' must hold integers between 0 and n(`n')"
        exit 459
    }
    preserve
    quietly keep if `touse' & `n' > 0
    tempvar row
    quietly gen long `row' = _n
    quietly expand `n'
    quietly bysort `row': replace `varlist' = (_n <= `varlist')
    * n() is no longer needed; dropping it avoids any clash with the
    * variables the analysis creates (it collapses to a variable named n)
    drop `n'
    global smaihda_input `"stratum table, n(`n') `if'"'
    capture noisily _smaihda_run `varlist', `options'
    local rc = _rc
    global smaihda_input ""
    if `rc' exit `rc'
    return add
    restore
end

program define _smaihda_run, rclass
    version 14.0

    syntax varname(numeric) [if] [in] [,        ///
        AREA(varname numeric)                    ///
        SDstratum(varname numeric)               ///
        AREACOVar(varname numeric)               ///
         THReshold(real -1)                      ///
         Bootstrap(integer 2000)                 ///
         SEED(integer 2026)                        ///
         MINN(integer 10)                        ///
         Components(string)                      ///
         SAVEdir(string)                         ///
         COMPare                                 ///
         GRaph                                    ///
         GRAPHDir(string)                         ///
         NODisplay]

    marksample touse
    local outcome  `varlist'
    global smaihda_flag ""

    * --------------------------------------------------------------
    * Design mode. area() and sdstratum() are both optional, but at
    * least one is required. Three configurations are supported:
    *   three-level    : area() + sdstratum()  (A-SD strata within areas)
    *   geographical   : area() only  (institutional/geographical MAIHDA:
    *                    individuals within areas, hospitals, regions ...)
    *   intersectional : sdstratum() only  (I-MAIHDA: individuals within
    *                    intersectional strata, no geographical level)
    * In the two 2-level modes the GCCI is not defined and is suppressed,
    * and the bootstrap resamples individuals within the single contextual level.
    * --------------------------------------------------------------
    if "`area'" == "" & "`sdstratum'" == "" {
        display as error "specify area() and/or sdstratum()"
        exit 198
    }
    tempvar _onelevel
    if "`area'" == "" {
        quietly gen byte `_onelevel' = 1 if `touse'
        local area       `_onelevel'
        local _mode      "intersectional"
        local _modelabel "Two-level MAIHDA (individuals within contexts)"
        local _bootclus  `sdstratum'
    }
    else if "`sdstratum'" == "" {
        quietly gen byte `_onelevel' = 1 if `touse'
        local sdstratum  `_onelevel'
        local _mode      "geographical"
        local _modelabel "Two-level MAIHDA (individuals within contexts)"
        local _bootclus  `area'
    }
    else {
        local _mode      "three-level"
        local _modelabel "Three-level (A-SD strata within areas)"
        local _bootclus  `area'
    }

    * --------------------------------------------------------------
    * Display setup: widen linesize for table rendering, restore at end
    * --------------------------------------------------------------
    local _smaihda_orig_linesize = c(linesize)
    capture quietly set linesize 120

    * --------------------------------------------------------------
    * 1. Input validation
    * --------------------------------------------------------------
    quietly summarize `outcome' if `touse'
    if r(N) == 0 {
        display as error "no observations meet the if/in restrictions"
        exit 2000
    }
    if r(min) < 0 | r(max) > 1 {
        display as error "outcome `outcome' must be binary (0/1)"
        exit 198
    }
    local pop_prev = r(mean)
    local N_total  = r(N)

    if `threshold' < 0 local threshold = `pop_prev'

    * --------------------------------------------------------------
    * 2. Component selection
    * --------------------------------------------------------------
    if "`components'" == "" local components "all"
    local do_props 0
    local do_std   0
    local do_ard   0
    local do_dev   0
    local do_gci   0
    local do_vpc   0
    local do_auc   0
    if strpos("`components'", "all") | strpos("`components'", "props") local do_props 1
    if strpos("`components'", "all") | strpos("`components'", "std")   local do_std   1
    if strpos("`components'", "all") | strpos("`components'", "ard")   local do_ard   1
    if strpos("`components'", "all") | strpos("`components'", "dev")   local do_dev   1
    if strpos("`components'", "all") | strpos("`components'", "gci")   local do_gci   1
    if strpos("`components'", "all") | strpos("`components'", "vpc")   local do_vpc   1
    if strpos("`components'", "all") | strpos("`components'", "auc")   local do_auc   1
    * GCCI is only defined in the three-level design; suppress otherwise.
    if "`_mode'" != "three-level" local do_gci 0
    local do_boot = (`bootstrap' > 0) & (`do_gci' | `do_vpc' | `do_auc')

    if `do_ard' & "`areacovar'" == "" {
        local do_ard 0
    }

    * --------------------------------------------------------------
    * 3. Counts and global stratum identifier
    * --------------------------------------------------------------
    quietly levelsof `area' if `touse', local(area_levels)
    local n_areas : word count `area_levels'

    quietly levelsof `sdstratum' if `touse', local(sd_levels)
    local n_sd : word count `sd_levels'

    tempvar asdstratum
    quietly egen `asdstratum' = group(`area' `sdstratum') if `touse'
    quietly levelsof `asdstratum' if `touse', local(asd_levels)
    local n_asd : word count `asd_levels'

    if "`savedir'" != "" {
        capture mkdir "`savedir'"
    }

    * --------------------------------------------------------------
    * 4. Display banner
    * --------------------------------------------------------------
    if "`nodisplay'" == "" {
        display _newline as text "{hline 70}"
        display as text "Simple-means MAIHDA (smaihda v1.9.0)"
        display as text "{hline 70}"
        display as text "Design:                 " as result "`_modelabel'"
        display as text "Outcome:                " as result "`outcome'"
        if `"$smaihda_input"' != "" {
            display as text "Input:                  " as result `"$smaihda_input"'
        }
        if "`_mode'" == "three-level" {
            display as text "Area variable:          " as result "`area' " ///
                as text "(" as result `n_areas' as text " areas)"
            display as text "SD stratum variable:    " as result "`sdstratum' " ///
                as text "(" as result `n_sd' as text " strata)"
        }
        else if "`_mode'" == "geographical" {
            display as text "Context variable:       " as result "`area' " ///
                as text "(" as result `n_areas' as text " contexts)"
        }
        else {
            display as text "Context variable:       " as result "`sdstratum' " ///
                as text "(" as result `n_sd' as text " contexts)"
        }
        if "`areacovar'" != "" {
            display as text "Area covariate:         " as result "`areacovar'"
        }
        display as text "Total individuals:      " as result `N_total'
        if "`_mode'" == "three-level" {
            display as text "A-SD strata:            " as result `n_asd'
        }
        display as text "Population prevalence:  " as result %5.3f `pop_prev'
        display as text "Threshold:              " as result %5.3f `threshold'
        if `do_boot' {
            display as text "Bootstrap:              " as result ///
                "B = `bootstrap', seed = `seed', contexts fixed, binomial draw per cell"
        }
        display as text "{hline 70}"
    }

    * --------------------------------------------------------------
    * 5. PRELIMINARIES: per-stratum aggregate datasets in tempfiles
    * --------------------------------------------------------------
    tempfile sd_props area_props asd_props
    tempfile area_std

    * 5a. SD strata proportions with Wilson CI
    preserve
        quietly keep if `touse'
        quietly collapse (count) n=`outcome' (sum) events=`outcome', ///
            by(`sdstratum')
        quietly _smaihda_wilson n events
        rename _wp p
        rename _wlo lo
        rename _whi hi
        gen byte high_risk = (lo > `threshold')
        quietly save "`sd_props'", replace
    restore

    * 5b. Area proportions with Wilson CI
    preserve
        quietly keep if `touse'
        if "`areacovar'" != "" {
            quietly collapse (count) n=`outcome' (sum) events=`outcome' ///
                (first) `areacovar', by(`area')
        }
        else {
            quietly collapse (count) n=`outcome' (sum) events=`outcome', ///
                by(`area')
        }
        quietly _smaihda_wilson n events
        rename _wp p
        rename _wlo lo
        rename _whi hi
        gen byte high_risk = (lo > `threshold')
        quietly save "`area_props'", replace
    restore

    * 5c. A-SD strata proportions with Wilson CI
    preserve
        quietly keep if `touse'
        if "`areacovar'" != "" {
            quietly collapse (count) n=`outcome' (sum) events=`outcome' ///
                (first) `areacovar', by(`asdstratum' `area' `sdstratum')
        }
        else {
            quietly collapse (count) n=`outcome' (sum) events=`outcome', ///
                by(`asdstratum' `area' `sdstratum')
        }
        quietly _smaihda_wilson n events
        rename _wp p
        rename _wlo lo
        rename _whi hi
        gen byte high_risk = (lo > `threshold')
        quietly save "`asd_props'", replace
    restore

    * --------------------------------------------------------------
    * 6. Display: Wilson CI proportions
    * --------------------------------------------------------------
    if `do_props' & "`nodisplay'" == "" {
        display _newline as text "{hline 70}"
        display as text "STEP 1: Crude proportions with Wilson 95% CI"
        display as text "{hline 70}"

        preserve
            quietly use "`sd_props'", clear
            display as text _newline "Sociodemographic strata (k=`n_sd'):"
            list `sdstratum' n events p lo hi high_risk, noobs sep(0) abbrev(15)
        restore

        preserve
            quietly use "`area_props'", clear
            display as text _newline "Areas (k=`n_areas'):"
            local _show "`area' n events p lo hi high_risk"
            if "`areacovar'" != "" local _show "`area' `areacovar' n events p lo hi high_risk"
            list `_show', noobs sep(0) abbrev(15)
        restore

        preserve
            quietly use "`asd_props'", clear
            quietly count
            local n_asd_actual = r(N)
            display as text _newline ///
                "A-SD strata: `n_asd_actual' rows (full listing suppressed; saved if -savedir- given)"
            quietly summarize p
            display as text "  proportion range: " as result %5.3f r(min) ///
                as text " to " as result %5.3f r(max)
            quietly count if high_risk == 1
            display as text "  strata above threshold: " as result r(N) ///
                as text " of " as result `n_asd_actual'
            * v1.5.0: cell-size diagnostic (basis of the stratum-design rule)
            quietly summarize n, detail
            local _Ntot = r(sum)
            display as text "  cell-size diagnostic: median n = " as result %4.0f r(p50) ///
                as text ", min n = " as result %4.0f r(min)
            foreach thr in 5 10 30 {
                quietly count if n < `thr'
                local _kc = r(N)
                quietly summarize n if n < `thr'
                local _ni = cond(r(N) > 0, r(sum), 0)
                local _pk = 100 * `_kc' / `n_asd_actual'
                local _pi = 100 * `_ni' / `_Ntot'
                display as text "    n < `thr'" _col(24) ": " as result `_kc' as text " cells (" ///
                    as result %4.1f `_pk' as text "%), " as result `_ni' as text " individuals (" ///
                    as result %4.1f `_pi' as text "%)"
            }
            foreach cond in "events == 0" "events == n" {
                quietly count if `cond'
                local _kc = r(N)
                quietly summarize n if `cond'
                local _ni = cond(r(N) > 0, r(sum), 0)
                local _pk = 100 * `_kc' / `n_asd_actual'
                local _pi = 100 * `_ni' / `_Ntot'
                local _lab = cond("`cond'" == "events == 0", "no events (p = 0)", "all events (p = 1)")
                display as text "    `_lab'" _col(24) ": " as result `_kc' as text " cells (" ///
                    as result %4.1f `_pk' as text "%), " as result `_ni' as text " individuals (" ///
                    as result %4.1f `_pi' as text "%)"
            }
            * v1.5.0: note when the area x stratum grid is incomplete
            if "`_mode'" == "three-level" {
                quietly levelsof `area', local(_gal)
                local _gnA : word count `_gal'
                quietly levelsof `sdstratum', local(_gsl)
                local _gnS : word count `_gsl'
                local _grid = `_gnA' * `_gnS'
                if `n_asd_actual' < `_grid' {
                    local _gempty = `_grid' - `n_asd_actual'
                    display as text "  Note: " as result `n_asd_actual' as text " of " ///
                        as result `_grid' as text " area-by-stratum cells are occupied (" ///
                        as result `_gempty' as text " empty)."
                    display as text "        EAR standardisation averages each area over the strata it contains;"
                    display as text "        areas with different strata are standardised to different populations."
                }
            }
        restore
    }

    if "`savedir'" != "" {
        copy "`sd_props'"   "`savedir'/sd_strata.dta", replace
        copy "`area_props'" "`savedir'/area_props.dta", replace
        copy "`asd_props'"  "`savedir'/asd_props.dta", replace
    }

    * --------------------------------------------------------------
    * 7. EAR-standardised area proportions
    * --------------------------------------------------------------
    if `do_std' {
        preserve
            quietly use "`asd_props'", clear
            * EAR = unweighted mean of SD strata within each area
            quietly bysort `area': egen p_std    = mean(p)
            * Variance propagation: SE = sqrt(sum(p*(1-p)/n)/k^2)
            * v1.5.0: variance from the Wilson interval width (never zero)
            quietly gen var_stratum = ((hi - lo) / (2*1.96))^2
            quietly bysort `area': egen sum_var = sum(var_stratum)
            quietly bysort `area': gen k = _N
            quietly gen se_std = sqrt(sum_var / (k^2))
            quietly gen p_std_lo = p_std - 1.96 * se_std
            quietly gen p_std_hi = p_std + 1.96 * se_std
            quietly bysort `area': keep if _n == 1
            if "`areacovar'" != "" {
                keep `area' `areacovar' p_std se_std p_std_lo p_std_hi k
            }
            else {
                keep `area' p_std se_std p_std_lo p_std_hi k
            }
            quietly save "`area_std'", replace

            if "`nodisplay'" == "" {
                display _newline as text "{hline 70}"
                display as text "STEP 2: EAR-standardised area proportions"
                display as text "{hline 70}"
                format p_std p_std_lo p_std_hi %6.3f
                list, noobs sep(0) abbrev(15)
            }

            if "`savedir'" != "" {
                copy "`area_std'" "`savedir'/area_std.dta", replace
            }
        restore
    }

    * --------------------------------------------------------------
    * 8. ARD and RR (crude and EAR-adjusted) by areacovar
    * --------------------------------------------------------------
    local ARD_c   = .
    local ARD_clo = .
    local ARD_chi = .
    local RR_c    = .
    local RR_clo  = .
    local RR_chi  = .
    local ARD_a   = .
    local ARD_alo = .
    local ARD_ahi = .
    local RR_a    = .
    local RR_alo  = .
    local RR_ahi  = .

    if `do_ard' {
        if "`nodisplay'" == "" {
            display _newline as text "{hline 70}"
            display as text "STEP 3: Area covariate effect (ARD and RR)"
            display as text "by `areacovar'"
            display as text "{hline 70}"
        }

        * --- Crude (raw area proportions) ---
        preserve
            quietly use "`area_props'", clear
            quietly bysort `areacovar': egen mean_p = mean(p)
            quietly gen var_p = p * (1 - p) / n
            quietly bysort `areacovar': egen sum_var = sum(var_p)
            quietly bysort `areacovar': gen k = _N
            quietly gen se_mean = sqrt(sum_var / (k^2))
            quietly bysort `areacovar': keep if _n == 1
            quietly sort `areacovar'

            * areacovar = 0 (reference) is row 1, areacovar = 1 is row 2
            local p0   = mean_p[1]
            local p1   = mean_p[2]
            local se0  = se_mean[1]
            local se1  = se_mean[2]
            local ARD_c   = (`p0' - `p1') * 100
            local seARD   = sqrt(`se0'^2 + `se1'^2)
            local ARD_clo = `ARD_c' - 1.96 * `seARD' * 100
            local ARD_chi = `ARD_c' + 1.96 * `seARD' * 100
            if `p1' > 0 {
                local RR_c   = `p0' / `p1'
                local seLRR  = sqrt(`se0'^2/`p0'^2 + `se1'^2/`p1'^2)
                local RR_clo = `RR_c' * exp(-1.96 * `seLRR')
                local RR_chi = `RR_c' * exp( 1.96 * `seLRR')
            }
        restore

        if "`nodisplay'" == "" {
            display as text "  Crude:"
            display as text "    ARD = " as result %6.2f `ARD_c'  ///
                as text " pp (95% CI " as result %6.2f `ARD_clo' ///
                as text " to " as result %6.2f `ARD_chi' as text ")"
            display as text "    RR  = " as result %6.3f `RR_c' ///
                as text "    (95% CI " as result %6.3f `RR_clo' ///
                as text " to " as result %6.3f `RR_chi' as text ")"
        }

        * --- EAR-adjusted ---
        if `do_std' {
            preserve
                quietly use "`area_std'", clear
                quietly bysort `areacovar': egen mean_pstd = mean(p_std)
                quietly gen var_pstd = se_std^2
                quietly bysort `areacovar': egen sum_var = sum(var_pstd)
                quietly bysort `areacovar': gen ka = _N
                quietly gen se_mean = sqrt(sum_var / (ka^2))
                quietly bysort `areacovar': keep if _n == 1
                quietly sort `areacovar'

                local q0   = mean_pstd[1]
                local q1   = mean_pstd[2]
                local sq0  = se_mean[1]
                local sq1  = se_mean[2]
                local ARD_a   = (`q0' - `q1') * 100
                local seARDa  = sqrt(`sq0'^2 + `sq1'^2)
                local ARD_alo = `ARD_a' - 1.96 * `seARDa' * 100
                local ARD_ahi = `ARD_a' + 1.96 * `seARDa' * 100
                if `q1' > 0 {
                    local RR_a   = `q0' / `q1'
                    local seLRRa = sqrt(`sq0'^2/`q0'^2 + `sq1'^2/`q1'^2)
                    local RR_alo = `RR_a' * exp(-1.96 * `seLRRa')
                    local RR_ahi = `RR_a' * exp( 1.96 * `seLRRa')
                }
            restore

            if "`nodisplay'" == "" {
                display as text "  EAR-adjusted:"
                display as text "    ARD = " as result %6.2f `ARD_a'  ///
                    as text " pp (95% CI " as result %6.2f `ARD_alo' ///
                    as text " to " as result %6.2f `ARD_ahi' as text ")"
                display as text "    RR  = " as result %6.3f `RR_a' ///
                    as text "    (95% CI " as result %6.3f `RR_alo' ///
                    as text " to " as result %6.3f `RR_ahi' as text ")"
            }
        }
    }

    * --------------------------------------------------------------
    * 9. Dev_A,SD with Wilson-shift CI
    * --------------------------------------------------------------
    tempfile dev_data
    if `do_dev' {
        preserve
            quietly use "`asd_props'", clear
            rename n n_asd
            rename events ev_asd
            rename p p_asd
            local _keepvars "`asdstratum' `area' `sdstratum' n_asd ev_asd p_asd lo hi"
            if "`areacovar'" != "" local _keepvars "`_keepvars' `areacovar'"
            keep `_keepvars'
            quietly merge m:1 `area' using "`area_props'", ///
                keepusing(n p) nogen
            rename n n_area
            rename p p_area

            rename lo lo_asd
            rename hi hi_asd
            quietly gen dev = p_asd - p_area
            * v1.5.0 Wilson shift: the cell's Wilson interval moved by the area
            * proportion, which is treated as known. Never degenerates at p = 0 or 1.
            quietly gen dev_lo = lo_asd - p_area
            quietly gen dev_hi = hi_asd - p_area
            quietly gen se_dev = (dev_hi - dev_lo) / (2*1.96)
            quietly gen byte sig = (dev_lo > 0 | dev_hi < 0)
            quietly save "`dev_data'", replace

            if "`nodisplay'" == "" {
                display _newline as text "{hline 70}"
                display as text "STEP 4: Deviation Dev_A,SD with Wilson-shift 95% CI"
                display as text "{hline 70}"
                quietly count
                display as text "  Total A-SD strata: " as result r(N)
                quietly count if sig == 1
                display as text "  Significantly deviating: " as result r(N) ///
                    as text " (" as result %5.1f r(N)/_N*100 as text "%)"
                quietly summarize dev
                display as text "  Dev range: " as result %6.3f r(min) ///
                    as text " to " as result %6.3f r(max)
            }

            if "`savedir'" != "" {
                copy "`dev_data'" "`savedir'/dev_asd.dta", replace
            }
        restore
    }

    * --------------------------------------------------------------
    * 9b. Figures (option -graph-)
    * --------------------------------------------------------------
    if "`graph'" != "" {
        * a sentinel keeps the positional arguments aligned when the user has
        * not supplied an area covariate or a graph directory
        local _acv "`areacovar'"
        local _gdr "`graphdir'"
        if "`_acv'" == "" local _acv "."
        if "`_gdr'" == "" local _gdr "."
        _smaihda_graphs "`asd_props'" "`dev_data'" "`area'" "`sdstratum'" ///
            "`_acv'" `threshold' "`outcome'" "`_gdr'"
    }

    * --------------------------------------------------------------
    * 10. GCCI, VPC, AUC: compute on observed data
    * --------------------------------------------------------------
    local GCCI = .
    local VPC_A = .
    local VPC_SD = .
    local ICC_AS = .
    local V_A = .
    local V_SD = .
    local V_I = .
    local ICC_ASD_s = .
    local ICC_ASD_c = .
    local GCCI_s = .
    local GCCI_c = .
    local VPC_A_s = .
    local VPC_SD_s = .
    local VPC_A_c = .
    local VPC_SD_c = .
    local ICC_ASD_s_lo = .
    local ICC_ASD_s_hi = .
    local ICC_ASD_c_lo = .
    local ICC_ASD_c_hi = .
    local GCCI_s_lo = .
    local GCCI_s_hi = .
    local GCCI_c_lo = .
    local GCCI_c_hi = .
    local CCI = .
    local Gshare = .
    local CMI = .
    local CCI_lo = .
    local CCI_hi = .
    local CMI_lo = .
    local CMI_hi = .
    local Gshare_lo = .
    local Gshare_hi = .
    local AUC_asd = .
    local AUC_area = .
    local AUC_asd_s = .
    local AUC_area_s = .
    local AUC_area_ear = .
    local Sens = .
    local Spec = .
    local PPV  = .
    local n_flag = .
    local TP = .
    local FP = .
    local FN = .
    local TN = .

    if `do_gci' | `do_vpc' | `do_auc' {
        _smaihda_compute "`outcome'" "`area'" "`asdstratum'" ///
            "`asd_props'" "`area_props'" `do_gci' `do_vpc' `do_auc' `touse' "`_mode'" "`sdstratum'" `minn'
        local GCCI     = r(GCCI)
        local VPC_A   = r(VPC_A)
        local VPC_SD  = r(VPC_SD)
        local ICC_AS  = r(ICC_ASD)
        local V_A     = r(V_A)
        local V_SD    = r(V_SD)
        local V_I     = r(V_I)
        local ICC_ASD_s = r(ICC_ASD_s)
        local ICC_ASD_c = r(ICC_ASD_c)
        local GCCI_s    = r(GCCI_s)
        local GCCI_c    = r(GCCI_c)
        local VPC_A_s   = r(VPC_A_s)
        local VPC_SD_s  = r(VPC_SD_s)
        local VPC_A_c   = r(VPC_A_c)
        local VPC_SD_c  = r(VPC_SD_c)
        local CCI       = r(CCI)
        local Gshare    = r(Gshare)
        local CMI       = r(CMI)
        local K_all     = r(K_all)
        local K_std     = r(K_std)
        local N_excl    = r(N_excl)
        local AUC_asd  = r(AUC_asd)
        local AUC_area = r(AUC_area)
        local AUC_asd_s  = r(AUC_asd_s)
        local AUC_area_s = r(AUC_area_s)
        local AUC_area_ear = r(AUC_area_ear)
    }

    * Threshold-based sensitivity / specificity
    if `do_auc' {
        preserve
            quietly keep if `touse'
            quietly merge m:1 `asdstratum' using "`asd_props'", ///
                keepusing(p) nogen keep(match)
            quietly gen byte hr = (p >= `threshold')
            quietly count if `outcome'==1 & hr==1
            local TP = r(N)
            quietly count if `outcome'==0 & hr==1
            local FP = r(N)
            quietly count if `outcome'==1 & hr==0
            local FN = r(N)
            quietly count if `outcome'==0 & hr==0
            local TN = r(N)
            if `TP' + `FN' > 0 local Sens = `TP' / (`TP' + `FN')
            if `TN' + `FP' > 0 local Spec = `TN' / (`TN' + `FP')
            if `TP' + `FP' > 0 local PPV  = `TP' / (`TP' + `FP')
            tempvar _onestr
            quietly bysort `asdstratum': gen byte `_onestr' = (_n==1)
            quietly count if `_onestr'==1 & hr==1
            local n_flag = r(N)
        restore
    }

    * --------------------------------------------------------------
    * 11. Within-context bootstrap
    * --------------------------------------------------------------
    local GCCI_lo  = .
    local GCCI_hi  = .
    local VPC_A_lo  = .
    local VPC_A_hi  = .
    local VPC_SD_lo = .
    local VPC_SD_hi = .
    local ICC_AS_lo  = .
    local ICC_AS_hi  = .
    local AUC_asd_lo = .
    local AUC_asd_hi = .
    local AUC_area_lo = .
    local AUC_area_hi = .
    local AUC_asd_s_lo = .
    local AUC_asd_s_hi = .
    local AUC_area_s_lo = .
    local AUC_area_s_hi = .
    local AUC_area_ear_lo = .
    local AUC_area_ear_hi = .

    if `do_boot' {
        if "`nodisplay'" == "" {
            display _newline as text "{hline 70}"
            display as text "STEP 5: Within-context bootstrap (B = `bootstrap', seed = `seed')"
            display as text "Contexts fixed; the cases of each cell redrawn from Binomial(n, p). Please wait..."
            display as text "{hline 70}"
        }

        tempname boot_res
        matrix `boot_res' = J(`bootstrap', 16, .)

        * v1.7.0: the bootstrap works on the table of cells (asd_props), the
        * table from which the point estimates are computed. The contexts are
        * the observed set and are held fixed. In each replicate the number of
        * cases of every A-SD cell is drawn from Binomial(n, p) of the cell,
        * the same experiment as resampling its individuals with replacement.
        * A cell with p = 0 or p = 1 has no sampling variation and keeps its
        * count. The cells are sorted by area and stratum before every draw,
        * so the interval does not depend on the order of the rows.
        preserve
            quietly use "`asd_props'", clear
            quietly keep `area' `sdstratum' n events
            quietly gen double _p0  = events / n
            quietly gen double _evb = events
            set seed `seed'
            local progress_step = max(1, floor(`bootstrap' / 10))
            forval b = 1/`bootstrap' {
                sort `area' `sdstratum'
                quietly replace _evb = rbinomial(n, _p0) if _p0 > 0 & _p0 < 1
                quietly _smaihda_compute_cells `area' `sdstratum' n _evb "`_mode'" `minn'
                matrix `boot_res'[`b', 1]  = r(GCCI)
                matrix `boot_res'[`b', 2]  = r(VPC_A)
                matrix `boot_res'[`b', 3]  = r(VPC_SD)
                matrix `boot_res'[`b', 4]  = r(ICC_ASD)
                matrix `boot_res'[`b', 5]  = r(AUC_asd)
                matrix `boot_res'[`b', 6]  = r(AUC_area)
                matrix `boot_res'[`b', 7]  = r(ICC_ASD_s)
                matrix `boot_res'[`b', 8]  = r(ICC_ASD_c)
                matrix `boot_res'[`b', 9]  = r(GCCI_s)
                matrix `boot_res'[`b', 10] = r(GCCI_c)
                matrix `boot_res'[`b', 11] = r(CMI)
                matrix `boot_res'[`b', 12] = r(CCI)
                matrix `boot_res'[`b', 13] = r(Gshare)
                matrix `boot_res'[`b', 14] = r(AUC_asd_s)
                matrix `boot_res'[`b', 15] = r(AUC_area_s)
                matrix `boot_res'[`b', 16] = r(AUC_area_ear)
                if "`nodisplay'" == "" & mod(`b', `progress_step') == 0 {
                    display as text "  ... " as result `b' as text " / `bootstrap' replicates"
                }
            }
        restore

        * Compute percentile CIs
        preserve
            clear
            quietly svmat `boot_res', names(col)
            rename c1 GCCI_b
            rename c2 VPC_A_b
            rename c3 VPC_SD_b
            rename c4 ICC_AS_b
            rename c5 AUC_asd_b
            rename c6 AUC_area_b
            rename c7  ICC_ASD_s_b
            rename c8  ICC_ASD_c_b
            rename c9  GCCI_s_b
            rename c10 GCCI_c_b
            rename c11 CMI_b
            rename c12 CCI_b
            rename c13 Gshare_b
            rename c14 AUC_asd_s_b
            rename c15 AUC_area_s_b
            rename c16 AUC_area_ear_b

            foreach v in GCCI VPC_A VPC_SD ICC_AS AUC_asd AUC_area ///
                         AUC_asd_s AUC_area_s AUC_area_ear ///
                         ICC_ASD_s ICC_ASD_c GCCI_s GCCI_c CMI CCI Gshare {
                quietly _pctile `v'_b, percentiles(2.5 97.5)
                local _q_lo = r(r1)
                local _q_hi = r(r2)
                quietly summarize `v'_b, meanonly
                local _mrep = r(mean)
                * v1.6.0: a replicate carries two doses of binomial noise against
                * one in the observed proportions, so every dispersion-based
                * measure is centred above the estimate by one dose. The interval
                * is the percentile interval recentred on the estimate and
                * truncated to the admissible range (all measures are proportions).
                if missing(``v'') | missing(`_mrep') {
                    local `v'_lo = .
                    local `v'_hi = .
                }
                else {
                    local `v'_lo = max(``v'' + (`_q_lo' - `_mrep'), 0)
                    local `v'_hi = min(``v'' + (`_q_hi' - `_mrep'), 1)
                }
            }

            if "`savedir'" != "" {
                quietly save "`savedir'/bootstrap.dta", replace
            }
        restore
    }

    * --------------------------------------------------------------
    * 12. Display GCCI / VPC / AUC summary
    * --------------------------------------------------------------
    if "`nodisplay'" == "" & (`do_gci' | `do_vpc' | `do_auc') {
        display _newline as text "{hline 70}"
        display as text "STEP 6: Summary statistics (CCI, VPC, AUC)"
        display as text "{hline 70}"
        display as text "{col 5}Quantity{col 28}Estimate{col 45}95% CI"
        display as text "{hline 70}"

        if `do_gci' {
            local _e = `GCCI'    * 100
            local _l = `GCCI_lo' * 100
            local _h = `GCCI_hi' * 100
            _smaihda_show_row "CCI, crude (%)" `_e' `_l' `_h'
        }
        if `do_vpc' {
            if "`_mode'" == "three-level" local _sfx " area+SD"
            else                          local _sfx ""

            if "`_mode'" == "three-level" {
                local _e = `VPC_A'    * 100
                local _l = `VPC_A_lo' * 100
                local _h = `VPC_A_hi' * 100
                _smaihda_show_row "VPC area (%)" `_e' `_l' `_h'

                local _e = `VPC_SD'    * 100
                local _l = `VPC_SD_lo' * 100
                local _h = `VPC_SD_hi' * 100
                _smaihda_show_row "VPC SD strata (%)" `_e' `_l' `_h'
            }

            local _e = `ICC_AS'    * 100
            local _l = `ICC_AS_lo' * 100
            local _h = `ICC_AS_hi' * 100
            _smaihda_show_row "VPC/ICC`_sfx' (%)" `_e' `_l' `_h'

            if `K_std' < `K_all' {
                local _kx = `K_all' - `K_std'
                display as text _col(5) "Standardised measures use " as result `K_std' ///
                    as text " of " as result `K_all' as text " cells (n >= `minn'); " ///
                    as result `_kx' as text " cells and " as result %9.0f `N_excl' ///
                    as text " individuals excluded."
            }
            display as text _col(5) "Standardised (weight 1 per context):"
            local _e = `ICC_ASD_s'    * 100
            local _l = `ICC_ASD_s_lo' * 100
            local _h = `ICC_ASD_s_hi' * 100
            _smaihda_show_row "  VPC/ICC`_sfx', uncorr (%)" `_e' `_l' `_h'

            local _e = `ICC_ASD_c'    * 100
            local _l = `ICC_ASD_c_lo' * 100
            local _h = `ICC_ASD_c_hi' * 100
            _smaihda_show_row "  VPC/ICC`_sfx', corr (%)" `_e' `_l' `_h'

            if "`_mode'" == "three-level" {
                local _e = `GCCI_s'    * 100
                local _l = `GCCI_s_lo' * 100
                local _h = `GCCI_s_hi' * 100
                _smaihda_show_row "  CCI, uncorr (%)" `_e' `_l' `_h'

                local _e = `GCCI_c'    * 100
                local _l = `GCCI_c_lo' * 100
                local _h = `GCCI_c_hi' * 100
                _smaihda_show_row "  CCI, corr (%)" `_e' `_l' `_h'

                display as text _col(5) "Contextual decomposition (share of between-stratum var.):"
                local _e = `CCI'    * 100
                local _l = `CCI_lo' * 100
                local _h = `CCI_hi' * 100
                _smaihda_show_row "  CCI clustering (%)" `_e' `_l' `_h'
                local _e = `Gshare'    * 100
                local _l = `Gshare_lo' * 100
                local _h = `Gshare_hi' * 100
                _smaihda_show_row "  G additive (%)" `_e' `_l' `_h'
                local _e = `CMI'    * 100
                local _l = `CMI_lo' * 100
                local _h = `CMI_hi' * 100
                _smaihda_show_row "  CMI modification (%)" `_e' `_l' `_h'
            }
        }
        if `do_auc' {
            _smaihda_show_row "coAUC A-SD strata"   `AUC_asd'   `AUC_asd_lo'   `AUC_asd_hi'
            _smaihda_show_row "coAUC area"          `AUC_area'  `AUC_area_lo'  `AUC_area_hi'
            _smaihda_show_row "coAUC* A-SD strata"  `AUC_asd_s'  `AUC_asd_s_lo'  `AUC_asd_s_hi'
            _smaihda_show_row "coAUC* area (size)"  `AUC_area_s' `AUC_area_s_lo' `AUC_area_s_hi'
            _smaihda_show_row "coAUC*EAR area"      `AUC_area_ear' `AUC_area_ear_lo' `AUC_area_ear_hi'
            local _sens_pct = `Sens' * 100
            local _spec_pct = `Spec' * 100
            display as text _col(5) "Sensitivity (%) " _col(28) ///
                as result %6.1f `_sens_pct'
            display as text _col(5) "Specificity (%) " _col(28) ///
                as result %6.1f `_spec_pct'
            local _ppv_pct = `PPV' * 100
            display as text _col(5) "PPV (%) " _col(28) ///
                as result %6.1f `_ppv_pct'
            display as text _col(5) "Strata flagged high-risk" _col(50) ///
                as result %8.0f `n_flag'
            display as text _col(5) "True positives (cases in flagged)" _col(50) ///
                as result %8.0f `TP'
            display as text _col(5) "False positives (non-cases flagged)" _col(50) ///
                as result %8.0f `FP'
            display as text _col(5) "False negatives (cases not flagged)" _col(50) ///
                as result %8.0f `FN'
            display as text _col(5) "True negatives (non-cases not flagged)" _col(50) ///
                as result %8.0f `TN'
        }
        display as text "{hline 70}"
        if "$smaihda_flag" == "1" {
            display as text "(!) point estimate outside its percentile interval; report the point"
            display as text "    estimate and state the limitation."
        }
        macro drop smaihda_flag
    }

    * --------------------------------------------------------------
    * 12a. Variance partition components (probability scale)
    *      Mirrors S1 Text Step 7: V_A + V_SD + V_I = p_pop(1 - p_pop).
    *      Absolute components; the ratios above are these over the total.
    * --------------------------------------------------------------
    if "`nodisplay'" == "" & `do_vpc' & `V_A' < . {
        local V_tot   = `V_A' + `V_SD' + `V_I'
        local V_check = `pop_prev' * (1 - `pop_prev')
        display _newline as text "Variance partition (probability scale)"
        display as text "{hline 70}"
        display as text _col(5) "V_A  (between areas)"        _col(42) as result %9.6f `V_A'
        display as text _col(5) "V_SD (strata within areas)" _col(42) as result %9.6f `V_SD'
        display as text _col(5) "V_I  (within strata)"       _col(42) as result %9.6f `V_I'
        display as text _col(5) "V_A + V_SD + V_I"           _col(42) as result %9.6f `V_tot'
        display as text _col(5) "p(1-p)  (identity check)"   _col(42) as result %9.6f `V_check'
        display as text "{hline 70}"
    }

    * --------------------------------------------------------------
    * 12b. Compare with random-effects MAIHDA (optional)
    * --------------------------------------------------------------
    local VPC_re      = .
    local AUC_re      = .
    local Sens_re     = .
    local Spec_re     = .
    local PPV_re      = .
    local TP_re       = .
    local FP_re       = .
    local FN_re       = .
    local TN_re       = .
    local SM_min      = .
    local SM_max      = .
    local SM_range    = .
    local SM_sd       = .
    local RE_min      = .
    local RE_max      = .
    local RE_range    = .
    local RE_sd       = .
    local RE_compress = .
    local atten_mean  = .
    local atten_max   = .
    local atten_max_n = .

    if "`compare'" != "" {
        if "`nodisplay'" == "" {
            display _newline as text "{hline 70}"
            display as text "Comparison: simple-means vs random-effects MAIHDA"
            display as text "{hline 70}"
            display as text "  Fitting melogit on A-SD strata. Please wait..."
        }

        tempvar asd_id_re
        quietly egen `asd_id_re' = group(`area' `sdstratum') if `touse'

        capture noisily quietly melogit `outcome' if `touse' || `asd_id_re':
        if _rc != 0 {
            if "`nodisplay'" == "" {
                display as error "  melogit did not converge; comparison skipped."
            }
        }
        else {
            * Variance partition from melogit (latent logit scale).
            * Robust across Stata versions and at the zero-variance boundary.
            * Stata 17+ stores the variance directly as var(_cons) in the
            * equation named after the random-effects level variable, which
            * here is the tempvar `asd_id_re'. Older Stata stores the log-SD
            * as lns1_1_1. If neither parameter is present, the variance has
            * converged to the zero boundary and we treat it as zero.
            tempname _re_var _re_lnsd
            local _pi2_3 = (c(pi)^2) / 3
            * Primary: estat icc — robust across all Stata versions
            capture quietly estat icc
            if _rc == 0 {
                * r(icc2) = ICC at level 2 (between strata)
                * Recover tau^2 from ICC: ICC = tau2/(tau2+pi2/3)
                * => tau2 = ICC*pi2/3 / (1-ICC)
                local _icc2 = r(icc2)
                if `_icc2' < 1 {
                    scalar `_re_var' = `_icc2' * `_pi2_3' / (1 - `_icc2')
                }
                else {
                    scalar `_re_var' = 0
                }
            }
            else {
                * Fallback 1: Stata 17+ stores variance as var(_cons)
                capture scalar `_re_var' = _b[`asd_id_re':var(_cons)]
                if _rc {
                    * Fallback 2: older Stata stores log-SD as lns1_1_1
                    capture scalar `_re_lnsd' = _b[lns1_1_1:_cons]
                    if _rc {
                        * Zero-variance boundary
                        scalar `_re_var' = 0
                    }
                    else {
                        scalar `_re_var' = exp(2 * `_re_lnsd')
                    }
                }
            }
            * RE VPC on the PROBABILITY scale by simulation (Method B,
            * Goldstein, Browne and Rasbash 2002), so it matches the
            * simple-means probability-scale VPC. Reuses `_re_var' (the
            * between-stratum variance) extracted by the cascade above.
            * The RNG seed is saved and restored so the command has no
            * side effect on the user's random-number state.
            local _b0    = _b[`outcome':_cons]
            tempname _sd_re
            scalar `_sd_re' = sqrt(`_re_var')
            local _seed0 = c(seed)
            preserve
                quietly clear
                quietly set obs 100000
                set seed 90125
                tempvar _u _pis _vis
                quietly gen double `_u'   = rnormal(0, `_sd_re')
                quietly gen double `_pis' = invlogit(`_b0' + `_u')
                quietly gen double `_vis' = `_pis' * (1 - `_pis')
                quietly summarize `_pis'
                local _v2 = r(Var)
                quietly summarize `_vis'
                local _v1 = r(mean)
            restore
            set seed `_seed0'
            local VPC_re = `_v2' / (`_v2' + `_v1')

            * Shrunken predicted probabilities at the individual level
            tempvar p_re p_sm_indiv
            quietly predict `p_re' if `touse'

            * Build per-individual simple-means prediction (observed
            * stratum proportion expanded to all individuals in the stratum)
            quietly bysort `asd_id_re': egen `p_sm_indiv' = ///
                mean(`outcome') if `touse'

            * AUC from shrunken predictions (per-individual, but equal
            * within stratum)
            quietly roctab `outcome' `p_re' if `touse', nograph
            local AUC_re = r(area)

            * Sensitivity and 1-specificity at the population-prevalence
            * threshold, using RE-shrunken stratum predictions
            tempvar high_risk_re cls_re cls_neg_re
            quietly gen byte `high_risk_re' = (`p_re' >= `threshold') ///
                if `touse'
            quietly gen byte `cls_re' = ///
                (`high_risk_re' == 1 & `outcome' == 1) if `touse'
            quietly summarize `cls_re' if `touse' & `outcome' == 1, meanonly
            local Sens_re = r(mean)
            quietly gen byte `cls_neg_re' = ///
                (`high_risk_re' == 0 & `outcome' == 0) if `touse'
            quietly summarize `cls_neg_re' if `touse' & `outcome' == 0, meanonly
            local Spec_re = r(mean)
            quietly count if `touse' & `high_risk_re'==1 & `outcome'==1
            local TP_re = r(N)
            quietly count if `touse' & `high_risk_re'==1 & `outcome'==0
            local FP_re = r(N)
            quietly count if `touse' & `high_risk_re'==0 & `outcome'==1
            local FN_re = r(N)
            quietly count if `touse' & `high_risk_re'==0 & `outcome'==0
            local TN_re = r(N)
            if `TP_re' + `FP_re' > 0 local PPV_re = `TP_re' / (`TP_re' + `FP_re')

            * Attenuation measures: collapse to one row per A-SD stratum,
            * compute summaries of SM and RE per-stratum predictions
            preserve
                quietly keep if `touse'
                quietly bysort `asd_id_re': keep if _n == 1
                quietly count
                local _nstrat = r(N)

                quietly summarize `p_sm_indiv'
                local SM_min   = r(min)
                local SM_max   = r(max)
                local SM_range = `SM_max' - `SM_min'
                local SM_sd    = r(sd)

                quietly summarize `p_re'
                local RE_min   = r(min)
                local RE_max   = r(max)
                local RE_range = `RE_max' - `RE_min'
                local RE_sd    = r(sd)

                if `SM_range' > 0 {
                    local RE_compress = (`RE_range' / `SM_range') * 100
                }

                * Per-stratum absolute attenuation
                tempvar _atten
                quietly gen `_atten' = abs(`p_sm_indiv' - `p_re')
                quietly summarize `_atten'
                local atten_mean = r(mean)
                local atten_max  = r(max)
            restore

            * n_j of the max-attenuation stratum.
            * Within each A-SD stratum, both p_sm_indiv and p_re are
            * constant (one prediction per stratum, expanded to individuals),
            * so |p_sm - p_re| is constant within stratum. We can therefore
            * count individuals whose attenuation matches the max directly
            * on the original (uncollapsed) data.
            quietly count if `touse' & ///
                abs(abs(`p_sm_indiv' - `p_re') - `atten_max') < 1e-10
            local atten_max_n = r(N)

            if "`nodisplay'" == "" {
                display _newline as text "Per-stratum predictions (A-SD level):"
                display as text _col(3) "Quantity" _col(40) ///
                    "Simple-means" _col(60) "Random-effects"
                display as text "{hline 70}"
                local _sm_min = `SM_min' * 100
                local _sm_max = `SM_max' * 100
                local _sm_rng = `SM_range' * 100
                local _sm_sd  = `SM_sd'  * 100
                local _re_min = `RE_min' * 100
                local _re_max = `RE_max' * 100
                local _re_rng = `RE_range' * 100
                local _re_sd  = `RE_sd'  * 100
                local _atten_mean_pp = `atten_mean' * 100
                local _atten_max_pp  = `atten_max'  * 100
                display as text _col(3) "Min predicted prevalence (%)" ///
                    _col(40) as result %8.2f `_sm_min' ///
                    _col(60) %8.2f `_re_min'
                display as text _col(3) "Max predicted prevalence (%)" ///
                    _col(40) as result %8.2f `_sm_max' ///
                    _col(60) %8.2f `_re_max'
                display as text _col(3) "Range of predictions (pp)" ///
                    _col(40) as result %8.2f `_sm_rng' ///
                    _col(60) %8.2f `_re_rng'
                if !missing(`RE_compress') {
                    display as text _col(3) "Range compression (RE/SM, %)" ///
                        _col(60) as result %8.2f `RE_compress'
                }
                display as text _col(3) "SD of predictions (pp)" ///
                    _col(40) as result %8.2f `_sm_sd' ///
                    _col(60) %8.2f `_re_sd'
                display as text _col(3) "Mean abs. attenuation (pp)" ///
                    _col(60) as result %8.2f `_atten_mean_pp'
                display as text _col(3) "Max abs. attenuation (pp)" ///
                    _col(60) as result %8.2f `_atten_max_pp' ///
                    as text "   (n=" as result `atten_max_n' as text ")"

                display _newline as text ///
                    "General contextual effect (probability-scale VPC):"
                display as text "{hline 70}"
                if !missing(`VPC_re') {
                    local _re_icc = `VPC_re'    * 100
                    local _sm_s   = `ICC_ASD_s' * 100
                    local _sm_c   = `ICC_ASD_c' * 100
                    display as text _col(3) "VPC/ICC area+SD (%)" ///
                        _col(40) "Simple-means" _col(60) "Random-effects"
                    display as text _col(5) "standardised, uncorr" ///
                        _col(40) as result %8.3f `_sm_s' ///
                        _col(60) %8.3f `_re_icc'
                    display as text _col(5) "standardised, corr" ///
                        _col(40) as result %8.3f `_sm_c' ///
                        _col(60) %8.3f `_re_icc'
                    display as text ///
                        "  GCE is compared on the standardised VPC (weight 1"
                    display as text ///
                        "  per stratum), the composition-independent measure. The"
                    display as text ///
                        "  RE tau^2 treats strata as units, matching this weighting."
                    display as text ///
                        "  Uncorrected is the observed reality; corrected puts the"
                    display as text ///
                        "  simple-means VPC on the same noise-discounted footing as"
                    display as text ///
                        "  the random-effects tau^2."
                }

                display _newline as text "Discriminatory accuracy and classification:"
                display as text "{hline 70}"
                display as text _col(3) "AUC A-SD strata" _col(40) ///
                    as result %8.3f `AUC_asd' _col(60) %8.3f `AUC_re'
                local _sens_sm = `Sens'   * 100
                local _spec_sm = `Spec'   * 100
                local _sens_re = `Sens_re' * 100
                local _spec_re = `Spec_re' * 100
                display as text _col(3) "Sensitivity (%)" _col(40) ///
                    as result %8.1f `_sens_sm' _col(60) %8.1f `_sens_re'
                display as text _col(3) "Specificity (%)" _col(40) ///
                    as result %8.1f `_spec_sm' _col(60) %8.1f `_spec_re'
                local _ppv_sm = `PPV'    * 100
                local _ppv_re = `PPV_re' * 100
                display as text _col(3) "PPV (%)" _col(40) ///
                    as result %8.1f `_ppv_sm' _col(60) %8.1f `_ppv_re'
                display as text _col(3) "TP (cases flagged)" _col(40) ///
                    as result %8.0f `TP' _col(60) %8.0f `TP_re'
                display as text _col(3) "FP (non-cases flagged)" _col(40) ///
                    as result %8.0f `FP' _col(60) %8.0f `FP_re'
                display as text _col(3) "FN (cases not flagged)" _col(40) ///
                    as result %8.0f `FN' _col(60) %8.0f `FN_re'
                display as text _col(3) "TN (non-cases not flagged)" _col(40) ///
                    as result %8.0f `TN' _col(60) %8.0f `TN_re'

                display as text "{hline 70}"
                display as text "  Simple-means uses observed stratum proportions;"
                display as text "  random-effects uses shrunken (empirical Bayes) predictions."
                display as text "  Range, SD and attenuation quantify shrinkage directly."
                display as text "  AUC is invariant under monotonic shrinkage; coincidence"
                display as text "  with simple-means AUC indicates uniform-enough stratum sizes"
                display as text "  for the ranking to be preserved. In intersectional designs"
                display as text "  with highly variable stratum sizes, AUC of the RE side"
                display as text "  typically falls below AUC of simple-means."
            }
        }
    }

    * --------------------------------------------------------------
    * 13. Return values
    * --------------------------------------------------------------
    return scalar N             = `N_total'
    return scalar n_areas       = `n_areas'
    return scalar n_sd          = `n_sd'
    return scalar n_asd         = `n_asd'
    return scalar pop_prev      = `pop_prev'
    return scalar threshold     = `threshold'

    return scalar GCCI           = `GCCI'
    return scalar GCCI_lo       = `GCCI_lo'
    return scalar GCCI_hi       = `GCCI_hi'

    return scalar VPC_A         = `VPC_A'
    return scalar VPC_A_lo      = `VPC_A_lo'
    return scalar VPC_A_hi      = `VPC_A_hi'

    return scalar VPC_SD        = `VPC_SD'
    return scalar VPC_SD_lo     = `VPC_SD_lo'
    return scalar VPC_SD_hi     = `VPC_SD_hi'

    return scalar ICC_ASD       = `ICC_AS'
    return scalar ICC_ASD_lo    = `ICC_AS_lo'
    return scalar ICC_ASD_hi    = `ICC_AS_hi'

    return scalar V_A           = `V_A'
    return scalar V_SD          = `V_SD'
    return scalar V_I           = `V_I'

    return scalar ICC_ASD_s     = `ICC_ASD_s'
    return scalar ICC_ASD_s_lo  = `ICC_ASD_s_lo'
    return scalar ICC_ASD_s_hi  = `ICC_ASD_s_hi'
    return scalar ICC_ASD_c     = `ICC_ASD_c'
    return scalar ICC_ASD_c_lo  = `ICC_ASD_c_lo'
    return scalar ICC_ASD_c_hi  = `ICC_ASD_c_hi'
    return scalar GCCI_s        = `GCCI_s'
    return scalar GCCI_s_lo     = `GCCI_s_lo'
    return scalar GCCI_s_hi     = `GCCI_s_hi'
    return scalar GCCI_c        = `GCCI_c'
    return scalar GCCI_c_lo     = `GCCI_c_lo'
    return scalar GCCI_c_hi     = `GCCI_c_hi'
    return scalar CCI           = `CCI'
    return scalar CCI_lo        = `CCI_lo'
    return scalar CCI_hi        = `CCI_hi'
    return scalar Gshare        = `Gshare'
    return scalar Gshare_lo     = `Gshare_lo'
    return scalar Gshare_hi     = `Gshare_hi'
    return scalar CMI           = `CMI'
    return scalar CMI_lo        = `CMI_lo'
    return scalar CMI_hi        = `CMI_hi'
    return scalar VPC_A_s        = `VPC_A_s'
    return scalar VPC_SD_s       = `VPC_SD_s'
    return scalar VPC_A_c        = `VPC_A_c'
    return scalar VPC_SD_c       = `VPC_SD_c'

    return scalar AUC_asd       = `AUC_asd'
    return scalar AUC_asd_lo    = `AUC_asd_lo'
    return scalar AUC_asd_hi    = `AUC_asd_hi'

    return scalar AUC_asd_s     = `AUC_asd_s'
    return scalar AUC_asd_s_lo  = `AUC_asd_s_lo'
    return scalar AUC_asd_s_hi  = `AUC_asd_s_hi'
    return scalar AUC_area_s    = `AUC_area_s'
    return scalar AUC_area_s_lo = `AUC_area_s_lo'
    return scalar AUC_area_s_hi = `AUC_area_s_hi'
    return scalar AUC_area_ear    = `AUC_area_ear'
    return scalar AUC_area_ear_lo = `AUC_area_ear_lo'
    return scalar AUC_area_ear_hi = `AUC_area_ear_hi'
    return scalar AUC_area      = `AUC_area'
    return scalar AUC_area_lo   = `AUC_area_lo'
    return scalar AUC_area_hi   = `AUC_area_hi'

    return scalar sensitivity   = `Sens'
    return scalar specificity   = `Spec'
    return scalar ppv           = `PPV'
    return scalar n_flag        = `n_flag'
    return scalar TP            = `TP'
    return scalar FP            = `FP'
    return scalar FN            = `FN'
    return scalar TN            = `TN'

    return scalar VPC_RE        = `VPC_re'
    return scalar AUC_RE        = `AUC_re'
    return scalar Sens_RE       = `Sens_re'
    return scalar Spec_RE       = `Spec_re'
    return scalar PPV_RE        = `PPV_re'
    return scalar TP_RE         = `TP_re'
    return scalar FP_RE         = `FP_re'
    return scalar FN_RE         = `FN_re'
    return scalar TN_RE         = `TN_re'

    return scalar SM_range      = `SM_range'
    return scalar SM_sd         = `SM_sd'
    return scalar RE_range      = `RE_range'
    return scalar RE_sd         = `RE_sd'
    return scalar RE_compress   = `RE_compress'
    return scalar atten_mean    = `atten_mean'
    return scalar atten_max     = `atten_max'
    return scalar atten_max_n   = `atten_max_n'

    return scalar ARD_crude     = `ARD_c'
    return scalar ARD_crude_lo  = `ARD_clo'
    return scalar ARD_crude_hi  = `ARD_chi'
    return scalar RR_crude      = `RR_c'
    return scalar RR_crude_lo   = `RR_clo'
    return scalar RR_crude_hi   = `RR_chi'

    return scalar ARD_adj       = `ARD_a'
    return scalar ARD_adj_lo    = `ARD_alo'
    return scalar ARD_adj_hi    = `ARD_ahi'
    return scalar RR_adj        = `RR_a'
    return scalar RR_adj_lo     = `RR_alo'
    return scalar RR_adj_hi     = `RR_ahi'

    return local cmd "smaihda"
    return local outcome "`outcome'"
    return local area "`area'"
    return local sdstratum "`sdstratum'"

    * Restore the user's original linesize
    capture quietly set linesize `_smaihda_orig_linesize'
end

* ==================================================================
* Helper: Wilson 95% CI computed in-dataset
* Adds variables _wp, _wlo, _whi to the working dataset
* Requires variables `n' and `events' to exist
* ==================================================================
program define _smaihda_wilson
    args nvar evar
    quietly gen double _wp = `evar' / `nvar'
    quietly gen double _wden = 1 + (1.96)^2 / `nvar'
    quietly gen double _wctr = (_wp + (1.96)^2/(2*`nvar')) / _wden
    quietly gen double _wmrg = (1.96/_wden) * ///
        sqrt(_wp*(1-_wp)/`nvar' + (1.96)^2/(4*`nvar'^2))
    quietly gen double _wlo = _wctr - _wmrg
    quietly gen double _whi = _wctr + _wmrg
    quietly drop _wden _wctr _wmrg
end

* ==================================================================
* Helper: GCCI, VPC, AUC on the observed sample
* Reads asd_props and area_props tempfiles; works on the active dataset
* ==================================================================
program define _smaihda_compute, rclass
    args outcome area_var asd_var asd_props area_props ///
        do_gci do_vpc do_auc touse cmode sd_var minn

    quietly summarize `outcome' if `touse'
    local grand = r(mean)
    local N_b   = r(N)

    * GCCI: V_between and V_within
    if `do_gci' {
        preserve
            quietly use "`area_props'", clear
            quietly gen vb = n * (p - `grand')^2
            quietly summarize vb
            local V_between = r(sum) / `N_b'
        restore

        preserve
            quietly use "`asd_props'", clear
            rename p p_asd
            rename n n_asd
            quietly merge m:1 `area_var' using "`area_props'", ///
                keepusing(p) keep(match) nogen
            rename p p_area
            quietly gen dev = p_asd - p_area
            quietly gen vw = n_asd * dev^2
            quietly summarize vw
            local V_within = r(sum) / `N_b'
        restore

        local GCCI = `V_between' / (`V_between' + `V_within')
        return scalar GCCI = `GCCI'
    }
    else {
        return scalar GCCI = .
    }

    * VPC on the PROBABILITY scale (correlation ratio; law of total
    * variance VA + VSD + VI = p(1-p)). Same scale as GCCI. The latent
    * logit-scale version and its method-of-moments (Taylor) noise
    * correction were removed in v1.0.7; that estimand is treated
    * separately (Box 2).
    if `do_vpc' {
        * Between-area component V_A
        preserve
            quietly use "`area_props'", clear
            quietly gen va = n * (p - `grand')^2
            quietly summarize va
            local V_A = r(sum) / `N_b'
        restore

        * Between A-SD-within-area component V_SD and within component V_I
        preserve
            quietly use "`asd_props'", clear
            rename p p_asd
            rename n n_asd
            quietly merge m:1 `area_var' using "`area_props'", ///
                keepusing(p) keep(match) nogen
            rename p p_area
            quietly gen vsd = n_asd * (p_asd - p_area)^2
            quietly summarize vsd
            local V_SD = r(sum) / `N_b'
            quietly gen vi = n_asd * p_asd * (1 - p_asd)
            quietly summarize vi
            local V_I = r(sum) / `N_b'
        restore

        local denom   = `V_A' + `V_SD' + `V_I'
        local VPC_A   = `V_A'  / `denom'
        local VPC_SD  = `V_SD' / `denom'
        local ICC_AS  = (`V_A' + `V_SD') / `denom'

        return scalar VPC_A = `VPC_A'
        return scalar VPC_SD = `VPC_SD'
        return scalar ICC_ASD = `ICC_AS'

        return scalar V_A  = `V_A'
        return scalar V_SD = `V_SD'
        return scalar V_I  = `V_I'

        * ----- Size-standardised variance partition (weight 1 per A-SD -----
        * stratum). p*_a is the unweighted mean of stratum proportions in an
        * area; pbar* the unweighted grand mean over all A-SD strata. Two
        * forms are returned: uncorrected, and corrected for binomial noise
        * (reassigned to V_I*, so the identity is preserved).
        preserve
            quietly use "`asd_props'", clear
            rename p p_asd
            rename n n_asd
            * v1.5.0: size-standardised measures on cells with n >= minn
            quietly count
            local K_all = r(N)
            quietly summarize n_asd if n_asd < `minn'
            local N_excl = cond(r(N) > 0, r(sum), 0)
            quietly keep if n_asd >= `minn'
            quietly count
            local Kstd = r(N)
            quietly bysort `area_var': egen p_a_star = mean(p_asd)
            quietly summarize p_asd
            local pbar_s = r(mean)
            quietly gen double _vas  = (p_a_star - `pbar_s')^2
            quietly summarize _vas
            local V_A_s = r(sum) / `Kstd'
            quietly gen double _vsds = (p_asd - p_a_star)^2
            quietly summarize _vsds
            local V_SD_s = r(sum) / `Kstd'
            quietly gen double _vis  = p_asd * (1 - p_asd)
            quietly summarize _vis
            local V_I_s = r(sum) / `Kstd'
            quietly gen double _nz   = p_asd * (1 - p_asd) / n_asd
            quietly summarize _nz
            local nz_SD = (1 - 1/`Kstd') * r(sum) / `Kstd'
            * v1.5.2: three-level design. The within-area deviation is taken from
            * the mean of the S_a cells of its own area, so its expected noise is
            * (1 - 1/S_a)*nz per cell. (1 - 1/K)*mean(nz) stays for the 2-level
            * designs, where the deviation is from the grand mean of the K cells.
            if "`cmode'" == "three-level" {
                quietly bysort `area_var': gen double _Sa = _N
                quietly gen double _nzS = (1 - 1/_Sa) * _nz
                quietly summarize _nzS
                local nz_SD = r(sum) / `Kstd'
            }
            quietly bysort `area_var': egen _nzam = mean(_nz)
            quietly bysort `area_var': gen _first = _n == 1
            * v1.8.1: (1 - S_a/K), because p* is the mean of the same K cells
            quietly bysort `area_var': gen double _SaA = _N
            quietly gen double _nzamw = (1 - _SaA/`Kstd') * _nzam
            quietly summarize _nzamw if _first
            local nz_A = r(sum) / `Kstd'
            * ----- Three-way split of V_SD: additive (G) and modification (CMI) -----
            * delta_SD = average within-area deviation of a sociodemographic stratum
            * across areas; gamma = its area-specific departure (the modification).
            local V_grad = .
            local V_CMI  = .
            local meannz = .
            local nA = .
            local nS = .
            if "`cmode'" == "three-level" {
                quietly summarize _nz
                local meannz = r(mean)
                quietly gen double _dstar = p_asd - p_a_star
                quietly bysort `sd_var': egen double _delta = mean(_dstar)
                quietly gen double _gamma = _dstar - _delta
                quietly gen double _vg = _delta^2
                quietly summarize _vg
                local V_grad = r(sum) / `Kstd'
                quietly gen double _vc = _gamma^2
                quietly summarize _vc
                local V_CMI = r(sum) / `Kstd'
                quietly levelsof `area_var', local(_al)
                local nA : word count `_al'
                quietly levelsof `sd_var', local(_sl)
                local nS : word count `_sl'
            }
        restore
        local den_s     = `pbar_s' * (1 - `pbar_s')
        local VPC_A_s   = `V_A_s'  / `den_s'
        local VPC_SD_s  = `V_SD_s' / `den_s'
        local ICC_ASD_s = (`V_A_s' + `V_SD_s') / `den_s'
        local GCCI_s    = `V_A_s' / (`V_A_s' + `V_SD_s')
        if "`cmode'" == "three-level" {
            local V_A_c  = `V_A_s'  - `nz_A'
            local V_SD_c = `V_SD_s' - `nz_SD'
        }
        else {
            * 2-level: a single between component. nz_SD equals
            * (1 - 1/K)*mean(nz), the between-context sampling noise. Subtract
            * it from the between component (area in geographical, stratum in
            * intersectional); the area/stratum split does not apply.
            local V_A_c  = `V_A_s'
            local V_SD_c = `V_SD_s'
            if "`cmode'" == "geographical" local V_A_c  = `V_A_s'  - `nz_SD'
            else                           local V_SD_c = `V_SD_s' - `nz_SD'
        }
        * v1.4.1: a variance component cannot be negative. When the binomial
        * noise estimate exceeds the raw between component (very sparse
        * information), the corrected component is floored at zero.
        local V_A_c  = max(`V_A_c',  0)
        local V_SD_c = max(`V_SD_c', 0)
        local VPC_A_c   = `V_A_c'  / `den_s'
        local VPC_SD_c  = `V_SD_c' / `den_s'
        local ICC_ASD_c = (`V_A_c' + `V_SD_c') / `den_s'
        * v1.4.1: GCCI_c is undefined when no contextual clustering is
        * detectable beyond binomial noise.
        if (`V_A_c' + `V_SD_c') > 0 local GCCI_c = `V_A_c' / (`V_A_c' + `V_SD_c')
        else                        local GCCI_c = .
        * ----- Contextual decomposition: CCI (level) / G (additive) / CMI -----
        * (three shares of the between-stratum variance; CMI noise-corrected).
        local CCI    = .
        local Gshare = .
        local CMI    = .
        if "`cmode'" == "three-level" {
            local floor_cmi = (1 - 1/`nA')*(1 - 1/`nS')*`meannz'
            local V_CMI_c = max(`V_CMI' - `floor_cmi', 0)
            if `V_CMI_c' > `V_SD_c' local V_CMI_c = `V_SD_c'
            local _tot = `V_A_c' + `V_SD_c'
            * v1.4.1: shares undefined when the corrected between variance is zero
            if `_tot' > 0 {
                local CCI    = `V_A_c' / `_tot'
                local CMI    = `V_CMI_c' / `_tot'
                local Gshare = (`V_SD_c' - `V_CMI_c') / `_tot'
            }
        }
        return scalar CCI    = `CCI'
        return scalar Gshare = `Gshare'
        return scalar CMI    = `CMI'
        return scalar VPC_A_s   = `VPC_A_s'
        return scalar VPC_SD_s  = `VPC_SD_s'
        return scalar ICC_ASD_s = `ICC_ASD_s'
        return scalar GCCI_s    = `GCCI_s'
        return scalar VPC_A_c   = `VPC_A_c'
        return scalar VPC_SD_c  = `VPC_SD_c'
        return scalar ICC_ASD_c = `ICC_ASD_c'
        return scalar GCCI_c    = `GCCI_c'
        return scalar K_all     = `K_all'
        return scalar K_std     = `Kstd'
        return scalar N_excl    = `N_excl'
    }
    else {
        return scalar VPC_A = .
        return scalar VPC_SD = .
        return scalar ICC_ASD = .

        return scalar V_A  = .
        return scalar V_SD = .
        return scalar V_I  = .

        return scalar VPC_A_s   = .
        return scalar VPC_SD_s  = .
        return scalar ICC_ASD_s = .
        return scalar GCCI_s    = .
        return scalar VPC_A_c   = .
        return scalar VPC_SD_c  = .
        return scalar ICC_ASD_c = .
        return scalar GCCI_c    = .
    }

    * AUC
    if `do_auc' {
        preserve
            quietly keep if `touse'
            quietly merge m:1 `asd_var' using "`asd_props'", ///
                keepusing(p) nogen keep(match)
            rename p pred_asd
            quietly merge m:1 `area_var' using "`area_props'", ///
                keepusing(p) nogen keep(match)
            rename p pred_area

            quietly roctab `outcome' pred_asd, nograph
            local AUC_asd = r(area)
            quietly roctab `outcome' pred_area, nograph
            local AUC_area = r(area)
        restore

        * v1.6.1: the same two AUC with every cell counted once
        * v1.6.2: cells below minn() are excluded, as in the standardised partition
        preserve
            quietly use "`asd_props'", clear
            quietly gen byte _big = (n >= `minn')
            _smaihda_auc_eq p _big
            local AUC_asd_s = r(auc)
        restore
        preserve
            quietly use "`asd_props'", clear
            quietly keep if n >= `minn'
            quietly collapse (sum) n events, by(`area_var')
            quietly gen double p = events / n
            _smaihda_auc_eq p
            local AUC_area_s = r(auc)
        restore

        * v1.9.0: coAUC*EAR, every area counted once and scored by its EAR,
        * the plain mean of the proportions of its cells with n >= minn
        preserve
            quietly use "`asd_props'", clear
            quietly keep if n >= `minn'
            quietly collapse (mean) p, by(`area_var')
            _smaihda_auc_eq p
            local AUC_area_ear = r(auc)
        restore

        return scalar AUC_asd = `AUC_asd'
        return scalar AUC_area = `AUC_area'
        return scalar AUC_asd_s = `AUC_asd_s'
        return scalar AUC_area_s = `AUC_area_s'
        return scalar AUC_area_ear = `AUC_area_ear'
    }
    else {
        return scalar AUC_asd = .
        return scalar AUC_area = .
        return scalar AUC_asd_s = .
        return scalar AUC_area_s = .
        return scalar AUC_area_ear = .
    }
end

* ==================================================================
* Helper (v1.7.0): GCCI, VPC, CCI/G/CMI and AUC from a table of cells
* ------------------------------------------------------------------
* The active dataset has one row per A-SD cell with its area, its
* sociodemographic stratum, its number of individuals and its number of
* cases. Every measure is computed from the two counts of each cell, with
* the formulas that _smaihda_compute applies to the observed cells. Used
* by the within-context bootstrap, which redraws the cases of each cell.
* args: area_var sd_var n_var ev_var cmode minn
* ==================================================================
program define _smaihda_compute_cells, rclass
    args area_var sd_var n_var ev_var cmode minn

    tempvar p non n_a ev_a p_a at vb vw vi
    quietly gen double `p'   = `ev_var' / `n_var'
    quietly gen double `non' = `n_var' - `ev_var'
    quietly summarize `n_var', meanonly
    local N_b = r(sum)
    quietly summarize `ev_var', meanonly
    local grand = r(sum) / `N_b'

    * --- Crude components: every individual counts once ---
    quietly bysort `area_var': egen double `n_a'  = total(`n_var')
    quietly by `area_var': egen double `ev_a' = total(`ev_var')
    quietly gen double `p_a' = `ev_a' / `n_a'
    quietly by `area_var': gen byte `at' = (_n == 1)
    quietly gen double `vb' = `n_a' * (`p_a' - `grand')^2 if `at'
    quietly summarize `vb', meanonly
    local V_between = r(sum) / `N_b'
    quietly gen double `vw' = `n_var' * (`p' - `p_a')^2
    quietly summarize `vw', meanonly
    local V_within = r(sum) / `N_b'
    quietly gen double `vi' = `n_var' * `p' * (1 - `p')
    quietly summarize `vi', meanonly
    local V_I = r(sum) / `N_b'

    return scalar GCCI = `V_between' / (`V_between' + `V_within')
    local denom = `V_between' + `V_within' + `V_I'
    return scalar VPC_A   = `V_between' / `denom'
    return scalar VPC_SD  = `V_within'  / `denom'
    return scalar ICC_ASD = (`V_between' + `V_within') / `denom'

    * --- Standardised components: every cell with n >= minn counts once ---
    tempvar big p_as vas vsds nz Sa nzS nz_am ats
    quietly gen byte `big' = (`n_var' >= `minn')
    quietly count if `big'
    local Kstd = r(N)
    quietly summarize `p' if `big', meanonly
    local pbar_s = r(mean)
    quietly bysort `area_var': egen double `p_as' = mean(cond(`big', `p', .))
    quietly gen double `vas'  = (`p_as' - `pbar_s')^2 if `big'
    quietly summarize `vas', meanonly
    local V_A_s = r(sum) / `Kstd'
    quietly gen double `vsds' = (`p' - `p_as')^2 if `big'
    quietly summarize `vsds', meanonly
    local V_SD_s = r(sum) / `Kstd'
    quietly gen double `nz' = `p' * (1 - `p') / `n_var' if `big'
    quietly summarize `nz', meanonly
    local nz_SD = (1 - 1/`Kstd') * r(sum) / `Kstd'
    * three-level design: per-cell factor (1 - 1/S_a), S_a = cells of the area
    if "`cmode'" == "three-level" {
        quietly by `area_var': egen double `Sa' = total(`big')
        quietly gen double `nzS' = (1 - 1/`Sa') * `nz' if `big'
        quietly summarize `nzS', meanonly
        local nz_SD = r(sum) / `Kstd'
    }
    quietly by `area_var': egen double `nz_am' = mean(`nz')
    quietly by `area_var': gen byte `ats' = (_n == 1) & !missing(`nz_am')
    * v1.8.1: (1 - S_a/K), because p* is the mean of the same K cells
    tempvar SaA nz_amw
    quietly by `area_var': egen double `SaA' = total(`big')
    quietly gen double `nz_amw' = (1 - `SaA'/`Kstd') * `nz_am'
    quietly summarize `nz_amw' if `ats', meanonly
    local nz_A = r(sum) / `Kstd'
    local den_s = `pbar_s' * (1 - `pbar_s')
    if "`cmode'" == "three-level" {
        local V_A_c  = `V_A_s'  - `nz_A'
        local V_SD_c = `V_SD_s' - `nz_SD'
    }
    else {
        local V_A_c  = `V_A_s'
        local V_SD_c = `V_SD_s'
        if "`cmode'" == "geographical" local V_A_c  = `V_A_s'  - `nz_SD'
        else                           local V_SD_c = `V_SD_s' - `nz_SD'
    }
    * boundary rule (v1.4.1)
    local V_A_c  = max(`V_A_c',  0)
    local V_SD_c = max(`V_SD_c', 0)
    return scalar ICC_ASD_s = (`V_A_s' + `V_SD_s') / `den_s'
    return scalar ICC_ASD_c = (`V_A_c' + `V_SD_c') / `den_s'
    return scalar GCCI_s    = `V_A_s' / (`V_A_s' + `V_SD_s')
    if (`V_A_c' + `V_SD_c') > 0 return scalar GCCI_c = `V_A_c' / (`V_A_c' + `V_SD_c')
    else                        return scalar GCCI_c = .

    * --- Contextual decomposition: CCI / G / CMI (three-level design) ---
    if "`cmode'" == "three-level" {
        tempvar dstar delta gamma vc sdtag
        quietly gen double `dstar' = `p' - `p_as' if `big'
        quietly bysort `sd_var': egen double `delta' = mean(`dstar')
        quietly gen double `gamma' = `dstar' - `delta' if `big'
        quietly summarize `nz', meanonly
        local meannz = r(mean)
        quietly gen double `vc' = `gamma'^2 if `big'
        quietly summarize `vc', meanonly
        local V_CMI = r(sum) / `Kstd'
        quietly count if `ats'
        local nA = r(N)
        quietly egen byte `sdtag' = tag(`sd_var') if `big'
        quietly count if `sdtag' == 1
        local nS = r(N)
        local floor_cmi = (1 - 1/`nA')*(1 - 1/`nS')*`meannz'
        local V_CMI_c = max(`V_CMI' - `floor_cmi', 0)
        if `V_CMI_c' > `V_SD_c' local V_CMI_c = `V_SD_c'
        local _tot = `V_A_c' + `V_SD_c'
        if `_tot' > 0 {
            return scalar CCI    = `V_A_c' / `_tot'
            return scalar Gshare = (`V_SD_c' - `V_CMI_c') / `_tot'
            return scalar CMI    = `V_CMI_c' / `_tot'
        }
        else {
            return scalar CCI    = .
            return scalar Gshare = .
            return scalar CMI    = .
        }
    }
    else {
        return scalar CCI    = .
        return scalar Gshare = .
        return scalar CMI    = .
    }

    * --- AUC ---
    * Crude: every individual counts once, the cases and non-cases of each
    * cell carrying the proportion of the cell, or of its area.
    _smaihda_auc_cells `p' `ev_var' `non'
    return scalar AUC_asd = r(auc)
    _smaihda_auc_cells `p_a' `ev_var' `non'
    return scalar AUC_area = r(auc)
    * Size-standardised: every cell with n >= minn counts once
    _smaihda_auc_eq `p' `big'
    return scalar AUC_asd_s = r(auc)
    * ... and every area counts once, its proportion formed from those cells
    tempvar nba eba pba tba
    quietly bysort `area_var': egen double `nba' = total(cond(`big', `n_var', 0))
    quietly by `area_var': egen double `eba' = total(cond(`big', `ev_var', 0))
    quietly gen double `pba' = `eba' / `nba' if `nba' > 0
    quietly by `area_var': gen byte `tba' = (_n == 1) & (`nba' > 0)
    _smaihda_auc_eq `pba' `tba'
    return scalar AUC_area_s = r(auc)
    * v1.9.0: coAUC*EAR, every area counted once and scored by its EAR
    tempvar pea
    quietly by `area_var': egen double `pea' = mean(cond(`big', `p', .))
    _smaihda_auc_eq `pea' `tba'
    return scalar AUC_area_ear = r(auc)
end

* ==================================================================
* Helper (v1.7.0): AUC from a table of cells
* ------------------------------------------------------------------
* The Mann-Whitney form of the AUC with every case and every non-case of
* each cell counted once, which is the value -roctab- returns on the
* individual records. Ties are counted at one half, as in roctab.
* args: pvar (the score of the cell), cavar (cases), novar (non-cases)
* ==================================================================
program define _smaihda_auc_cells, rclass
    args pvar cavar novar
    tempname auc
    scalar `auc' = .
    preserve
        quietly keep if !missing(`pvar')
        quietly count
        if r(N) >= 1 {
            tempvar cum gsum term
            sort `pvar'
            quietly gen double `cum' = sum(`novar')
            quietly by `pvar': egen double `gsum' = total(`novar')
            quietly by `pvar': replace `cum' = `cum'[_N]
            quietly gen double `term' = `cavar'*(`cum' - `gsum') + 0.5*`cavar'*`gsum'
            quietly summarize `cavar', meanonly
            local _tc = r(sum)
            quietly summarize `novar', meanonly
            local _tn = r(sum)
            quietly summarize `term', meanonly
            local _num = r(sum)
            if `_tc' > 0 & `_tn' > 0 scalar `auc' = `_num'/(`_tc'*`_tn')
        }
    restore
    return scalar auc = `auc'
end

* ==================================================================
* Helper (v1.6.1): size-standardised AUC from cell proportions
* ------------------------------------------------------------------
* Every cell contributes the same total weight (cases = p, non-cases
* = 1 - p), so the result is the AUC that would be observed if all
* cells were of equal size. The ordinary AUC counts pairs of
* individuals, so a cell of 3000 people contributes a million times
* more pairs than a cell of 3 and the value depends on how the
* population happens to be spread over the cells. Weighting every
* individual by 1/n of its own cell removes that, exactly as the
* standardised VPC counts every cell once. It is the same kind of
* object as an age-standardised rate: real arithmetic on a
* hypothetical population of equal-sized cells. No continuity
* correction is needed, since only the ordering of the proportions
* is used and a cell with no events has a well defined rank.
* Ties are counted at one half, as in roctab.
* args: pvar (the cell proportion), tagvar (optional 0/1 selector)
* ==================================================================
program define _smaihda_auc_eq, rclass
    args pvar tagvar
    tempname auc
    scalar `auc' = .
    preserve
        if "`tagvar'" != "" quietly keep if `tagvar'
        quietly keep if !missing(`pvar')
        quietly count
        if r(N) >= 2 {
            tempvar ca no cum gsum below term
            quietly gen double `ca' = `pvar'
            quietly gen double `no' = 1 - `pvar'
            sort `pvar'
            quietly gen double `cum' = sum(`no')
            quietly by `pvar': gen double `gsum' = sum(`no')
            quietly by `pvar': replace `gsum' = `gsum'[_N]
            quietly by `pvar': replace `cum'  = `cum'[_N]
            quietly gen double `below' = `cum' - `gsum'
            quietly gen double `term' = `ca'*`below' + 0.5*`ca'*`gsum'
            quietly summarize `ca', meanonly
            local _tc = r(sum)
            quietly summarize `no', meanonly
            local _tn = r(sum)
            quietly summarize `term', meanonly
            local _num = r(sum)
            if `_tc' > 0 & `_tn' > 0 scalar `auc' = `_num'/(`_tc'*`_tn')
        }
    restore
    return scalar auc = `auc'
end

* ==================================================================
* Helper: display one row of the summary table
* ==================================================================
program define _smaihda_show_row
    args label est lo hi
    if missing(`lo') | missing(`hi') {
        display as text _col(5) "`label'" _col(30) as result %6.3f `est'
    }
    else {
        * v1.5.0: flag a point estimate outside its own percentile interval
        local _flag ""
        if `est' < `lo' | `est' > `hi' {
            local _flag "  (!)"
            global smaihda_flag "1"
        }
        display as text _col(5) "`label'" _col(30) as result %6.3f `est' ///
            _col(47) as result "(" %6.3f `lo' ", " %6.3f `hi' ")" as text "`_flag'"
    }
end

* ==================================================================
* Helper: figures
* Reproduces Figure 2 (prevalence of all A-SD strata) and Figure 3
* (deviations Dev_A,SD) of the companion paper.
* Called from -smaihda- when the -graph- option is specified.
*   args: 1 asd file  2 dev file  3 area  4 sdstratum  5 areacovar
*         6 threshold (proportion)  7 outcome name  8 graphdir
* ==================================================================
program define _smaihda_graphs
    args asdfile devfile area sdstratum areacovar thresh outcome gdir

    * "." is the sentinel used by the caller for an omitted argument
    if "`areacovar'" == "." local areacovar ""
    if "`gdir'"      == "." local gdir ""

    local thr100 = `thresh'*100

    * ---------- Figure 2: prevalence of the A-SD strata ----------
    preserve
        quietly use "`asdfile'", clear
        quietly replace p  = p*100
        quietly replace lo = lo*100
        quietly replace hi = hi*100

        * rank within sociodemographic stratum, so each group fans out
        tempvar rk kk yy
        quietly bysort `sdstratum' (p): gen long `rk' = _n
        quietly by `sdstratum': gen long `kk' = _N
        quietly gen double `yy' = `sdstratum'
        quietly replace `yy' = `sdstratum' + 0.72*((`rk'-1)/(`kk'-1) - 0.5) ///
            if `kk' > 1

        * descriptive axis labels if the user has attached a value label
        local ylab ""
        local vlab : value label `sdstratum'
        quietly levelsof `sdstratum', local(slevels)
        foreach s of local slevels {
            if "`vlab'" != "" {
                local txt : label `vlab' `s'
                local ylab `ylab' `s' "`txt'"
            }
            else {
                local ylab `ylab' `s' "`s'"
            }
        }

        if "`areacovar'" != "" {
            twoway ///
              (rcap lo hi `yy' if `areacovar'==0, horizontal                  ///
                    lcolor("174 199 232") lwidth(vthin))                    ///
              (rcap lo hi `yy' if `areacovar'==1, horizontal                  ///
                    lcolor("255 200 150") lwidth(vthin))                    ///
              (scatter `yy' p if `areacovar'==0,                              ///
                    mcolor("31 119 180") msymbol(circle) msize(vsmall))     ///
              (scatter `yy' p if `areacovar'==1,                              ///
                    mcolor("255 127 14") msymbol(circle) msize(vsmall))     ///
              ,                                                             ///
              yscale(reverse) ylabel(`ylab', angle(0) labsize(vsmall))      ///
              ytitle("") xtitle("Prevalence (%)")                           ///
              xline(`thr100', lpattern(dash) lcolor(black))                 ///
              legend(order(3 "Poor area" 4 "Rich area") pos(4) ring(0)      ///
                     col(1) size(vsmall) region(lcolor(gs12)))              ///
              title("`outcome'", size(medsmall))                            ///
              graphregion(color(white)) plotregion(margin(medium))          ///
              name(smaihda_prev, replace)
        }
        else {
            twoway ///
              (rcap lo hi `yy', horizontal lcolor(gs12) lwidth(vthin))        ///
              (scatter `yy' p, mcolor("31 119 180") msymbol(circle)           ///
                    msize(vsmall))                                          ///
              ,                                                             ///
              yscale(reverse) ylabel(`ylab', angle(0) labsize(vsmall))      ///
              ytitle("") xtitle("Prevalence (%)")                           ///
              xline(`thr100', lpattern(dash) lcolor(black))                 ///
              legend(off) title("`outcome'", size(medsmall))                ///
              graphregion(color(white)) plotregion(margin(medium))          ///
              name(smaihda_prev, replace)
        }
        if "`gdir'" != "" {
            quietly graph export "`gdir'/smaihda_prevalence_`outcome'.png", ///
                replace width(2244)
            capture quietly graph export ///
                "`gdir'/smaihda_prevalence_`outcome'.eps", replace
        }
    restore

    * ---------- Figure 3: deviations Dev_A,SD ----------
    capture confirm file "`devfile'"
    if _rc {
        display as text ///
          "  (deviation figure skipped: run without -components()- to obtain it)"
        exit
    }

    preserve
        quietly use "`devfile'", clear
        quietly replace dev    = dev*100
        quietly replace dev_lo = dev_lo*100
        quietly replace dev_hi = dev_hi*100

        tempvar rk kk yy
        quietly bysort `sdstratum' (dev): gen long `rk' = _n
        quietly by `sdstratum': gen long `kk' = _N
        quietly gen double `yy' = `sdstratum'
        quietly replace `yy' = `sdstratum' + 0.72*((`rk'-1)/(`kk'-1) - 0.5) ///
            if `kk' > 1

        local ylab ""
        local vlab : value label `sdstratum'
        quietly levelsof `sdstratum', local(slevels)
        foreach s of local slevels {
            if "`vlab'" != "" {
                local txt : label `vlab' `s'
                local ylab `ylab' `s' "`txt'"
            }
            else {
                local ylab `ylab' `s' "`s'"
            }
        }

        local hasacv = 0
        capture confirm variable `areacovar'
        if !_rc & "`areacovar'" != "" local hasacv = 1

        if `hasacv' {
            twoway ///
              (rcap dev_lo dev_hi `yy' if `areacovar'==0, horizontal          ///
                    lcolor("174 199 232") lwidth(vthin))                    ///
              (rcap dev_lo dev_hi `yy' if `areacovar'==1, horizontal          ///
                    lcolor("255 200 150") lwidth(vthin))                    ///
              (scatter `yy' dev if sig==1 & `areacovar'==0,                   ///
                    msymbol(circle) mcolor("31 119 180") msize(vsmall))     ///
              (scatter `yy' dev if sig==0 & `areacovar'==0,                   ///
                    msymbol(circle_hollow) mlcolor("31 119 180")            ///
                    msize(vsmall))                                          ///
              (scatter `yy' dev if sig==1 & `areacovar'==1,                   ///
                    msymbol(circle) mcolor("255 127 14") msize(vsmall))     ///
              (scatter `yy' dev if sig==0 & `areacovar'==1,                   ///
                    msymbol(circle_hollow) mlcolor("255 127 14")            ///
                    msize(vsmall))                                          ///
              ,                                                             ///
              yscale(reverse) ylabel(`ylab', angle(0) labsize(vsmall))      ///
              xline(0, lpattern(dash) lcolor(black))                        ///
              ytitle("")                                                    ///
              xtitle("{&Delta}{subscript:A,SD} (percentage points)")        ///
              legend(order(3 "Poor area, 95% CI excludes 0"                 ///
                           5 "Rich area, 95% CI excludes 0"                 ///
                           4 "95% CI includes 0")                           ///
                     pos(4) ring(0) col(1) size(vsmall)                     ///
                     region(lcolor(gs12)))                                  ///
              title("`outcome'", size(medsmall))                            ///
              graphregion(color(white)) plotregion(margin(medium))          ///
              name(smaihda_dev, replace)
        }
        else {
            twoway ///
              (rcap dev_lo dev_hi `yy', horizontal                           ///
                    lcolor("174 199 232") lwidth(vthin))                    ///
              (scatter `yy' dev if sig==1, msymbol(circle)                    ///
                    mcolor("31 119 180") msize(vsmall))                     ///
              (scatter `yy' dev if sig==0, msymbol(circle_hollow)             ///
                    mlcolor("31 119 180") msize(vsmall))                    ///
              ,                                                             ///
              yscale(reverse) ylabel(`ylab', angle(0) labsize(vsmall))      ///
              xline(0, lpattern(dash) lcolor(black))                        ///
              ytitle("")                                                    ///
              xtitle("{&Delta}{subscript:A,SD} (percentage points)")        ///
              legend(order(2 "95% CI excludes 0" 3 "95% CI includes 0")     ///
                     pos(4) ring(0) col(1) size(vsmall))                    ///
              title("`outcome'", size(medsmall))                            ///
              graphregion(color(white)) plotregion(margin(medium))          ///
              name(smaihda_dev, replace)
        }
        if "`gdir'" != "" {
            quietly graph export "`gdir'/smaihda_deviations_`outcome'.png", ///
                replace width(2244)
            capture quietly graph export ///
                "`gdir'/smaihda_deviations_`outcome'.eps", replace
        }
    restore
end
```
