## Supplementary material for "Describing health inequalities without distortion: Simple-Means MAIHDA vs Random-Effects MAIHDA": S1 File. The smaihda Stata package: smaihda_v1.9.0_test.do

```
* smaihda v1.9.0. Test on the Malmo stratum table (6 October 2026)
*
* Run from this folder, S1_File_smaihda_package, with the folder
* S5_File_stratum_table beside it, as in the supporting information.
*
* The command runs twice on each outcome:
*   (1) on the stratum table of S5 File, read directly with the option n();
*   (2) on the same table expanded by hand to individual records with
*       -expand-, one outcome at a time, without n().
* The two runs must give the same results, the bootstrap limits included:
* the table and the records carry the same information, and the bootstrap
* draws the cases of each cell in a fixed order of the cells, whatever the
* order of the rows. The do-file stops with an error if a result differs.
* The random-effects results of -compare- are compared with a tolerance of
* 1e-5, because -melogit- sums over the records in a different order.
*
* Expected values (B = 2000, seed 2026). The point estimates are those of
* the paper and of the Python and R implementations (S5 and S6 Text). The
* bootstrap limits agree with theirs within Monte Carlo error, because each
* program draws its own random numbers. Python limits, for reference:
*   psycmed  VPC/ICC A+SD std corr 3.30% (2.88 to 3.71); CCI 4.1% (1.9 to
*            6.6); G 95.9% (90.7 to 100.0); CMI 0.0% (0.0 to 5.0);
*            AUC 62.71% (62.17 to 63.25); AUC area 52.97% (52.40 to 53.54);
*            AUC std size, strata 62.97%; area 53.22%; coAUC*EAR area
*            52.86% (52.21 to 53.51)
*   private  VPC/ICC A+SD std corr 10.72% (10.08 to 11.37); CCI 63.9% (60.5
*            to 67.3); G 19.0% (16.7 to 21.4); CMI 17.1% (14.3 to 19.9);
*            AUC 71.77% (71.23 to 72.31); AUC area 67.90% (67.33 to 68.49);
*            AUC std size, strata 71.90%; area 68.38%; coAUC*EAR area
*            67.46% (66.84 to 68.09)

clear all
set more off
log using "smaihda_v1.9.0_test.log", replace text
local table "../S5_File_stratum_table/dataset-smaihda_stratum_table.csv"

* Results compared between the two runs
local sm "N GCCI GCCI_lo GCCI_hi VPC_A VPC_A_lo VPC_A_hi VPC_SD VPC_SD_lo VPC_SD_hi"
local sm "`sm' ICC_ASD ICC_ASD_lo ICC_ASD_hi ICC_ASD_s ICC_ASD_s_lo ICC_ASD_s_hi"
local sm "`sm' ICC_ASD_c ICC_ASD_c_lo ICC_ASD_c_hi GCCI_s GCCI_s_lo GCCI_s_hi"
local sm "`sm' GCCI_c GCCI_c_lo GCCI_c_hi CCI CCI_lo CCI_hi Gshare Gshare_lo Gshare_hi"
local sm "`sm' CMI CMI_lo CMI_hi AUC_asd AUC_asd_lo AUC_asd_hi AUC_area AUC_area_lo"
local sm "`sm' AUC_area_hi AUC_asd_s AUC_asd_s_lo AUC_asd_s_hi AUC_area_s AUC_area_s_lo"
local sm "`sm' AUC_area_s_hi AUC_area_ear AUC_area_ear_lo AUC_area_ear_hi sensitivity specificity ppv ARD_crude ARD_crude_lo"
local sm "`sm' ARD_crude_hi RR_crude ARD_adj ARD_adj_lo ARD_adj_hi RR_adj"
local re "VPC_RE AUC_RE Sens_RE Spec_RE RE_compress atten_max atten_max_n"

* (1) The stratum table, read directly
import delimited using "`table'", clear
foreach y in psycmed private {
    smaihda cases if outcome == "`y'", area(area) sdstratum(strata_sociodem_id) ///
        areacovar(area_income) n(n) compare
    foreach r in `sm' `re' {
        scalar A_`y'_`r' = r(`r')
    }
}

* (2) The same table, expanded by hand to individual records
foreach y in psycmed private {
    import delimited using "`table'", clear
    quietly keep if outcome == "`y'"
    quietly expand n
    sort area strata_sociodem_id
    quietly by area strata_sociodem_id: gen byte `y' = (_n <= cases)
    drop outcome n cases
    smaihda `y', area(area) sdstratum(strata_sociodem_id) areacovar(area_income) compare
    local nbad = 0
    foreach r in `sm' `re' {
        scalar B_`y'_`r' = r(`r')
        local tol = cond(strpos(" `re' ", " `r' "), 1e-5, 1e-9)
        local d = cond(missing(scalar(A_`y'_`r')) & missing(scalar(B_`y'_`r')), 0, ///
            reldif(scalar(A_`y'_`r'), scalar(B_`y'_`r')))
        if missing(`d') | `d' > `tol' {
            display as error "`y' `r': table " scalar(A_`y'_`r') ", records " scalar(B_`y'_`r')
            local nbad = `nbad' + 1
        }
    }
    display as text "`y': " as result `nbad' as text " results differ between the table and the records"
    assert `nbad' == 0
}
display as text "All compared results are the same from the table and from the records."
log close
```
