## Supplementary material for "Describing health inequalities without distortion: Simple-Means MAIHDA vs Random-Effects MAIHDA": S2 Text. Supplementary tables

Supplementary Information S2 Text. Tables

**Index of tables**

- Table A. Dev_A,SD with 95% CIs — Psychotropic medication use (300 A-SD strata).
- Table B. Dev_A,SD with 95% CIs — Private GP choice (300 A-SD strata).
- Table C. Geographical spread of Dev_A,SD across the 25 areas, by stratum — Psychotropic.
- Table D. Geographical spread of Dev_A,SD across the 25 areas, by stratum — Private GP.

Legend for Tables Table A and Table B (Dev_A,SD):

Source: Malmö 2006 dataset (N = 43,291).

Two outcomes:

- Psychotropic medication use (overall prevalence 26.0%)

- Private GP choice (overall prevalence 22.1%)

Stratification: 25 geographical areas crossed with 12 sociodemographic strata = 300 A-SD strata (each row = one A-SD stratum).

Sociodemographic strata are defined by sex (0 = women, 1 = men), age band (1 = 35-44, 2 = 45-54, 3 = 55-64) and income (0 = low, 1 = high).

Columns:

- area: area identifier (1 to 25)
- SD: sociodemographic stratum identifier (1 to 12)
- sex / age / income: defining characteristics of the SD stratum, coded as above
- n_ASD: number of individuals in the A-SD stratum
- events: number of individuals with the outcome in the A-SD stratum
- p_ASD (%): observed proportion in the A-SD stratum
- n_area: number of individuals in the area (across all SD strata)
- p_area (%): crude proportion in the area (across all SD strata)
- Dev_A,SD (pp): deviation = p_ASD minus p_area, in percentage points
- SE (pp): (95% CI high − 95% CI low) / 3.92, the width of the Wilson-shift interval expressed as a standard error
- 95% CI low / high (pp): the Wilson 95% interval of the A-SD stratum proportion shifted by p_area, which is treated as known because it rests on the whole area. The interval is not symmetric around Dev_A,SD.
- Significant: 'Yes' if the 95% CI excludes zero, else 'No'

Significant rows are highlighted in light yellow.

Computation reproduces the deviation step of the smaihda command, which is STEP 4 of its output and Step 5 of S1 Text. A delta-method interval, dev ± 1.96 SE, has zero width for a stratum in which everyone or no one has the outcome; the Wilson shift never degenerates. In Malmö no stratum is at 0% or 100%, and the minimum stratum size is 35. The deviation is significant in 125 strata for psychotropic medication use and in 122 for private GP choice. Fig 5 of the main paper draws the deviations from the composition-standardised level of the area instead, which average zero within every area (Section 2.2 of the main paper). Within an area the two deviations differ by the composition gap of the area, on average 0.6 percentage points for psychotropic medication use and 1.2 for private GP choice, so the ordering of the strata within an area is the same.

Reference: the bootstrap used for the aggregate measures elsewhere in the paper follows Davison and Hinkley (1997); here a Wilson interval suffices, because each deviation involves only one small proportion and one large one.

**Table A. Dev_A,SD with 95% CIs — Psychotropic medication use (300 A-SD strata).**

**Overall prevalence: 25.99% | N = 43,291 across 300 A-SD strata Significantly deviating strata (95% CI does not cross zero): 125 of 300 (41.7%)**

| **Area** | **SD** | **Sex** | **Age** | **Income** | **n_ASD** | **Events** | **p_ASD (%)** | **n_area** | **p_area (%)** | **Dev_A,SD (pp)** | **SE (pp)** | **95% CI low (pp)** | **95% CI high (pp)** | **Significant** |
| --- | --- | --- | --- | --- | --- | --- | --- | --- | --- | --- | --- | --- | --- | --- |
| 1 | 1 | 0 | 1 | 0 | 79 | 27 | 34.18 | 1186 | 24.96 | 9.22 | 5.22 | -0.29 | 20.19 | No |
| 1 | 2 | 0 | 1 | 1 | 167 | 40 | 23.95 | 1186 | 24.96 | -1.01 | 3.28 | -6.85 | 6.01 | No |
| 1 | 3 | 0 | 2 | 0 | 66 | 23 | 34.85 | 1186 | 24.96 | 9.89 | 5.72 | -0.48 | 21.93 | No |
| 1 | 4 | 0 | 2 | 1 | 163 | 43 | 26.38 | 1186 | 24.96 | 1.42 | 3.42 | -4.74 | 8.68 | No |
| 1 | 5 | 0 | 3 | 0 | 74 | 37 | 50.00 | 1186 | 24.96 | 25.04 | 5.67 | 13.93 | 36.15 | Yes |
| 1 | 6 | 0 | 3 | 1 | 138 | 45 | 32.61 | 1186 | 24.96 | 7.65 | 3.94 | 0.39 | 15.85 | Yes |
| 1 | 7 | 1 | 1 | 0 | 51 | 5 | 9.80 | 1186 | 24.96 | -15.15 | 4.26 | -20.70 | -3.98 | Yes |
| 1 | 8 | 1 | 1 | 1 | 98 | 15 | 15.31 | 1186 | 24.96 | -9.65 | 3.63 | -15.46 | -1.23 | Yes |
| 1 | 9 | 1 | 2 | 0 | 38 | 8 | 21.05 | 1186 | 24.96 | -3.91 | 6.45 | -13.88 | 11.39 | No |
| 1 | 10 | 1 | 2 | 1 | 127 | 17 | 13.39 | 1186 | 24.96 | -11.57 | 3.03 | -16.43 | -4.56 | Yes |
| 1 | 11 | 1 | 3 | 0 | 57 | 21 | 36.84 | 1186 | 24.96 | 11.88 | 6.20 | 0.57 | 24.86 | Yes |
| 1 | 12 | 1 | 3 | 1 | 128 | 15 | 11.72 | 1186 | 24.96 | -13.24 | 2.86 | -17.73 | -6.52 | Yes |
| 2 | 1 | 0 | 1 | 0 | 163 | 44 | 26.99 | 1772 | 25.56 | 1.43 | 3.45 | -4.80 | 8.72 | No |
| 2 | 2 | 0 | 1 | 1 | 182 | 37 | 20.33 | 1772 | 25.56 | -5.23 | 2.97 | -10.44 | 1.20 | No |
| 2 | 3 | 0 | 2 | 0 | 129 | 49 | 37.98 | 1772 | 25.56 | 12.42 | 4.21 | 4.51 | 21.03 | Yes |
| 2 | 4 | 0 | 2 | 1 | 178 | 53 | 29.78 | 1772 | 25.56 | 4.21 | 3.40 | -2.02 | 11.30 | No |
| 2 | 5 | 0 | 3 | 0 | 174 | 73 | 41.95 | 1772 | 25.56 | 16.39 | 3.70 | 9.31 | 23.82 | Yes |
| 2 | 6 | 0 | 3 | 1 | 191 | 48 | 25.13 | 1772 | 25.56 | -0.43 | 3.12 | -6.05 | 6.17 | No |
| 2 | 7 | 1 | 1 | 0 | 112 | 23 | 20.54 | 1772 | 25.56 | -5.03 | 3.79 | -11.47 | 3.37 | No |
| 2 | 8 | 1 | 1 | 1 | 134 | 18 | 13.43 | 1772 | 25.56 | -12.13 | 2.95 | -16.90 | -5.33 | Yes |
| 2 | 9 | 1 | 2 | 0 | 94 | 27 | 28.72 | 1772 | 25.56 | 3.16 | 4.59 | -5.01 | 13.00 | No |
| 2 | 10 | 1 | 2 | 1 | 131 | 19 | 14.50 | 1772 | 25.56 | -11.06 | 3.08 | -16.08 | -4.02 | Yes |
| 2 | 11 | 1 | 3 | 0 | 115 | 30 | 26.09 | 1772 | 25.56 | 0.52 | 4.05 | -6.64 | 9.23 | No |
| 2 | 12 | 1 | 3 | 1 | 169 | 32 | 18.93 | 1772 | 25.56 | -6.63 | 3.00 | -11.82 | -0.06 | Yes |
| 3 | 1 | 0 | 1 | 0 | 294 | 88 | 29.93 | 2633 | 26.66 | 3.27 | 2.66 | -1.68 | 8.74 | No |
| 3 | 2 | 0 | 1 | 1 | 286 | 58 | 20.28 | 2633 | 26.66 | -6.38 | 2.37 | -10.63 | -1.34 | Yes |
| 3 | 3 | 0 | 2 | 0 | 238 | 88 | 36.97 | 2633 | 26.66 | 10.31 | 3.11 | 4.43 | 16.61 | Yes |
| 3 | 4 | 0 | 2 | 1 | 246 | 70 | 28.46 | 2633 | 26.66 | 1.79 | 2.86 | -3.48 | 7.73 | No |
| 3 | 5 | 0 | 3 | 0 | 255 | 97 | 38.04 | 2633 | 26.66 | 11.38 | 3.02 | 5.64 | 17.47 | Yes |
| 3 | 6 | 0 | 3 | 1 | 214 | 72 | 33.64 | 2633 | 26.66 | 6.98 | 3.20 | 0.99 | 13.55 | Yes |
| 3 | 7 | 1 | 1 | 0 | 222 | 48 | 21.62 | 2633 | 26.66 | -5.04 | 2.75 | -9.95 | 0.83 | No |
| 3 | 8 | 1 | 1 | 1 | 197 | 27 | 13.71 | 2633 | 26.66 | -12.96 | 2.45 | -17.07 | -7.46 | Yes |

| **Area** | **SD** | **Sex** | **Age** | **Income** | **n_ASD** | **Events** | **p_ASD (%)** | **n_area** | **p_area (%)** | **Dev_A,SD (pp)** | **SE (pp)** | **95% CI low (pp)** | **95% CI high (pp)** | **Significant** |
| --- | --- | --- | --- | --- | --- | --- | --- | --- | --- | --- | --- | --- | --- | --- |
| 3 | 9 | 1 | 2 | 0 | 169 | 54 | 31.95 | 2633 | 26.66 | 5.29 | 3.55 | -1.27 | 12.66 | No |
| 3 | 10 | 1 | 2 | 1 | 175 | 30 | 17.14 | 2633 | 26.66 | -9.52 | 2.84 | -14.38 | -3.24 | Yes |
| 3 | 11 | 1 | 3 | 0 | 164 | 43 | 26.22 | 2633 | 26.66 | -0.44 | 3.41 | -6.57 | 6.78 | No |
| 3 | 12 | 1 | 3 | 1 | 173 | 27 | 15.61 | 2633 | 26.66 | -11.05 | 2.76 | -15.71 | -4.91 | Yes |
| 4 | 1 | 0 | 1 | 0 | 189 | 62 | 32.80 | 1994 | 28.03 | 4.77 | 3.39 | -1.52 | 11.75 | No |
| 4 | 2 | 0 | 1 | 1 | 230 | 56 | 24.35 | 1994 | 28.03 | -3.69 | 2.81 | -8.78 | 2.25 | No |
| 4 | 3 | 0 | 2 | 0 | 175 | 70 | 40.00 | 1994 | 28.03 | 11.97 | 3.66 | 5.00 | 19.36 | Yes |
| 4 | 4 | 0 | 2 | 1 | 196 | 62 | 31.63 | 1994 | 28.03 | 3.60 | 3.29 | -2.51 | 10.41 | No |
| 4 | 5 | 0 | 3 | 0 | 174 | 74 | 42.53 | 1994 | 28.03 | 14.49 | 3.71 | 7.39 | 21.92 | Yes |
| 4 | 6 | 0 | 3 | 1 | 147 | 42 | 28.57 | 1994 | 28.03 | 0.54 | 3.69 | -6.15 | 8.31 | No |
| 4 | 7 | 1 | 1 | 0 | 142 | 29 | 20.42 | 1994 | 28.03 | -7.61 | 3.36 | -13.42 | -0.24 | Yes |
| 4 | 8 | 1 | 1 | 1 | 172 | 25 | 14.53 | 1994 | 28.03 | -13.50 | 2.69 | -17.99 | -7.46 | Yes |
| 4 | 9 | 1 | 2 | 0 | 124 | 42 | 33.87 | 1994 | 28.03 | 5.84 | 4.19 | -1.90 | 14.54 | No |
| 4 | 10 | 1 | 2 | 1 | 181 | 40 | 22.10 | 1994 | 28.03 | -5.93 | 3.07 | -11.36 | 0.65 | No |
| 4 | 11 | 1 | 3 | 0 | 118 | 33 | 27.97 | 1994 | 28.03 | -0.07 | 4.08 | -7.37 | 8.63 | No |
| 4 | 12 | 1 | 3 | 1 | 146 | 24 | 16.44 | 1994 | 28.03 | -11.60 | 3.06 | -16.73 | -4.74 | Yes |
| 5 | 1 | 0 | 1 | 0 | 95 | 25 | 26.32 | 1665 | 25.53 | 0.79 | 4.45 | -7.02 | 10.44 | No |
| 5 | 2 | 0 | 1 | 1 | 208 | 44 | 21.15 | 1665 | 25.53 | -4.37 | 2.82 | -9.37 | 1.68 | No |
| 5 | 3 | 0 | 2 | 0 | 97 | 37 | 38.14 | 1665 | 25.53 | 12.62 | 4.84 | 3.58 | 22.56 | Yes |
| 5 | 4 | 0 | 2 | 1 | 218 | 54 | 24.77 | 1665 | 25.53 | -0.75 | 2.91 | -6.02 | 5.38 | No |
| 5 | 5 | 0 | 3 | 0 | 121 | 49 | 40.50 | 1665 | 25.53 | 14.97 | 4.40 | 6.65 | 23.88 | Yes |
| 5 | 6 | 0 | 3 | 1 | 203 | 66 | 32.51 | 1665 | 25.53 | 6.99 | 3.26 | 0.92 | 13.70 | Yes |
| 5 | 7 | 1 | 1 | 0 | 90 | 23 | 25.56 | 1665 | 25.53 | 0.03 | 4.53 | -7.85 | 9.91 | No |
| 5 | 8 | 1 | 1 | 1 | 138 | 19 | 13.77 | 1665 | 25.53 | -11.76 | 2.94 | -16.53 | -5.02 | Yes |
| 5 | 9 | 1 | 2 | 0 | 76 | 19 | 25.00 | 1665 | 25.53 | -0.53 | 4.88 | -8.90 | 10.25 | No |
| 5 | 10 | 1 | 2 | 1 | 158 | 35 | 22.15 | 1665 | 25.53 | -3.37 | 3.28 | -9.14 | 3.72 | No |
| 5 | 11 | 1 | 3 | 0 | 90 | 24 | 26.67 | 1665 | 25.53 | 1.14 | 4.59 | -6.90 | 11.09 | No |
| 5 | 12 | 1 | 3 | 1 | 171 | 30 | 17.54 | 1665 | 25.53 | -7.98 | 2.90 | -12.95 | -1.59 | Yes |
| 6 | 1 | 0 | 1 | 0 | 91 | 28 | 30.77 | 1266 | 22.35 | 8.42 | 4.76 | -0.13 | 18.52 | No |
| 6 | 2 | 0 | 1 | 1 | 169 | 23 | 13.61 | 1266 | 22.35 | -8.74 | 2.64 | -13.11 | -2.76 | Yes |
| 6 | 3 | 0 | 2 | 0 | 68 | 18 | 26.47 | 1266 | 22.35 | 4.12 | 5.24 | -4.90 | 15.65 | No |
| 6 | 4 | 0 | 2 | 1 | 183 | 50 | 27.32 | 1266 | 22.35 | 4.97 | 3.27 | -0.97 | 11.84 | No |
| 6 | 5 | 0 | 3 | 0 | 82 | 32 | 39.02 | 1266 | 22.35 | 16.67 | 5.27 | 6.83 | 27.49 | Yes |
| 6 | 6 | 0 | 3 | 1 | 144 | 45 | 31.25 | 1266 | 22.35 | 8.90 | 3.82 | 1.90 | 16.87 | Yes |
| 6 | 7 | 1 | 1 | 0 | 66 | 15 | 22.73 | 1266 | 22.35 | 0.37 | 5.07 | -8.07 | 11.82 | No |

| **Area** | **SD** | **Sex** | **Age** | **Income** | **n_ASD** | **Events** | **p_ASD (%)** | **n_area** | **p_area (%)** | **Dev_A,SD (pp)** | **SE (pp)** | **95% CI low (pp)** | **95% CI high (pp)** | **Significant** |
| --- | --- | --- | --- | --- | --- | --- | --- | --- | --- | --- | --- | --- | --- | --- |
| 6 | 8 | 1 | 1 | 1 | 120 | 13 | 10.83 | 1266 | 22.35 | -11.52 | 2.86 | -15.91 | -4.70 | Yes |
| 6 | 9 | 1 | 2 | 0 | 35 | 10 | 28.57 | 1266 | 22.35 | 6.22 | 7.33 | -6.03 | 22.70 | No |
| 6 | 10 | 1 | 2 | 1 | 126 | 18 | 14.29 | 1266 | 22.35 | -8.07 | 3.12 | -13.12 | -0.90 | Yes |
| 6 | 11 | 1 | 3 | 0 | 44 | 9 | 20.45 | 1266 | 22.35 | -1.90 | 5.96 | -11.20 | 12.15 | No |
| 6 | 12 | 1 | 3 | 1 | 138 | 22 | 15.94 | 1266 | 22.35 | -6.41 | 3.11 | -11.58 | 0.61 | No |
| 7 | 1 | 0 | 1 | 0 | 117 | 32 | 27.35 | 1340 | 27.39 | -0.04 | 4.07 | -7.30 | 8.66 | No |
| 7 | 2 | 0 | 1 | 1 | 169 | 30 | 17.75 | 1340 | 27.39 | -9.64 | 2.93 | -14.66 | -3.18 | Yes |
| 7 | 3 | 0 | 2 | 0 | 89 | 35 | 39.33 | 1340 | 27.39 | 11.94 | 5.07 | 2.43 | 22.33 | Yes |
| 7 | 4 | 0 | 2 | 1 | 153 | 37 | 24.18 | 1340 | 27.39 | -3.21 | 3.43 | -9.30 | 4.16 | No |
| 7 | 5 | 0 | 3 | 0 | 100 | 48 | 48.00 | 1340 | 27.39 | 20.61 | 4.90 | 11.08 | 30.30 | Yes |
| 7 | 6 | 0 | 3 | 1 | 115 | 37 | 32.17 | 1340 | 27.39 | 4.79 | 4.30 | -3.06 | 13.78 | No |
| 7 | 7 | 1 | 1 | 0 | 80 | 21 | 26.25 | 1340 | 27.39 | -1.14 | 4.84 | -9.53 | 9.43 | No |
| 7 | 8 | 1 | 1 | 1 | 128 | 21 | 16.41 | 1340 | 27.39 | -10.98 | 3.26 | -16.40 | -3.61 | Yes |
| 7 | 9 | 1 | 2 | 0 | 82 | 28 | 34.15 | 1340 | 27.39 | 6.76 | 5.13 | -2.59 | 17.52 | No |
| 7 | 10 | 1 | 2 | 1 | 134 | 26 | 19.40 | 1340 | 27.39 | -7.99 | 3.40 | -13.79 | -0.48 | Yes |
| 7 | 11 | 1 | 3 | 0 | 73 | 29 | 39.73 | 1340 | 27.39 | 12.34 | 5.59 | 1.90 | 23.80 | Yes |
| 7 | 12 | 1 | 3 | 1 | 100 | 23 | 23.00 | 1340 | 27.39 | -4.39 | 4.16 | -11.54 | 4.77 | No |
| 8 | 1 | 0 | 1 | 0 | 127 | 38 | 29.92 | 1452 | 28.37 | 1.55 | 4.01 | -5.73 | 10.00 | No |
| 8 | 2 | 0 | 1 | 1 | 142 | 29 | 20.42 | 1452 | 28.37 | -7.95 | 3.36 | -13.76 | -0.58 | Yes |
| 8 | 3 | 0 | 2 | 0 | 117 | 49 | 41.88 | 1452 | 28.37 | 13.51 | 4.49 | 4.96 | 22.56 | Yes |
| 8 | 4 | 0 | 2 | 1 | 150 | 44 | 29.33 | 1452 | 28.37 | 0.96 | 3.68 | -5.74 | 8.69 | No |
| 8 | 5 | 0 | 3 | 0 | 138 | 61 | 44.20 | 1452 | 28.37 | 15.83 | 4.17 | 7.81 | 24.16 | Yes |
| 8 | 6 | 0 | 3 | 1 | 115 | 34 | 29.57 | 1452 | 28.37 | 1.19 | 4.20 | -6.38 | 10.08 | No |
| 8 | 7 | 1 | 1 | 0 | 151 | 34 | 22.52 | 1452 | 28.37 | -5.86 | 3.37 | -11.79 | 1.44 | No |
| 8 | 8 | 1 | 1 | 1 | 114 | 24 | 21.05 | 1452 | 28.37 | -7.32 | 3.79 | -13.80 | 1.04 | No |
| 8 | 9 | 1 | 2 | 0 | 106 | 32 | 30.19 | 1452 | 28.37 | 1.81 | 4.39 | -6.11 | 11.12 | No |
| 8 | 10 | 1 | 2 | 1 | 98 | 16 | 16.33 | 1452 | 28.37 | -12.05 | 3.72 | -18.07 | -3.49 | Yes |
| 8 | 11 | 1 | 3 | 0 | 89 | 26 | 29.21 | 1452 | 28.37 | 0.84 | 4.74 | -7.59 | 10.99 | No |
| 8 | 12 | 1 | 3 | 1 | 105 | 25 | 23.81 | 1452 | 28.37 | -4.57 | 4.11 | -11.70 | 4.41 | No |
| 9 | 1 | 0 | 1 | 0 | 198 | 55 | 27.78 | 1957 | 26.57 | 1.21 | 3.16 | -4.56 | 7.82 | No |
| 9 | 2 | 0 | 1 | 1 | 216 | 35 | 16.20 | 1957 | 26.57 | -10.37 | 2.50 | -14.68 | -4.87 | Yes |
| 9 | 3 | 0 | 2 | 0 | 194 | 72 | 37.11 | 1957 | 26.57 | 10.54 | 3.44 | 4.06 | 17.53 | Yes |
| 9 | 4 | 0 | 2 | 1 | 163 | 44 | 26.99 | 1957 | 26.57 | 0.42 | 3.45 | -5.80 | 7.71 | No |
| 9 | 5 | 0 | 3 | 0 | 206 | 89 | 43.20 | 1957 | 26.57 | 16.63 | 3.42 | 10.05 | 23.46 | Yes |
| 9 | 6 | 0 | 3 | 1 | 145 | 41 | 28.28 | 1957 | 26.57 | 1.70 | 3.70 | -4.99 | 9.52 | No |

| **Area** | **SD** | **Sex** | **Age** | **Income** | **n_ASD** | **Events** | **p_ASD (%)** | **n_area** | **p_area (%)** | **Dev_A,SD (pp)** | **SE (pp)** | **95% CI low (pp)** | **95% CI high (pp)** | **Significant** |
| --- | --- | --- | --- | --- | --- | --- | --- | --- | --- | --- | --- | --- | --- | --- |
| 9 | 7 | 1 | 1 | 0 | 178 | 36 | 20.22 | 1957 | 26.57 | -6.35 | 3.00 | -11.59 | 0.15 | No |
| 9 | 8 | 1 | 1 | 1 | 161 | 24 | 14.91 | 1957 | 26.57 | -11.66 | 2.81 | -16.34 | -5.35 | Yes |
| 9 | 9 | 1 | 2 | 0 | 140 | 42 | 30.00 | 1957 | 26.57 | 3.43 | 3.83 | -3.55 | 11.47 | No |
| 9 | 10 | 1 | 2 | 1 | 116 | 23 | 19.83 | 1957 | 26.57 | -6.74 | 3.68 | -12.98 | 1.43 | No |
| 9 | 11 | 1 | 3 | 0 | 117 | 32 | 27.35 | 1957 | 26.57 | 0.78 | 4.07 | -6.48 | 9.48 | No |
| 9 | 12 | 1 | 3 | 1 | 123 | 27 | 21.95 | 1957 | 26.57 | -4.62 | 3.70 | -11.02 | 3.48 | No |
| 10 | 1 | 0 | 1 | 0 | 182 | 43 | 23.63 | 2166 | 27.42 | -3.80 | 3.13 | -9.38 | 2.88 | No |
| 10 | 2 | 0 | 1 | 1 | 235 | 52 | 22.13 | 2166 | 27.42 | -5.30 | 2.70 | -10.13 | 0.44 | No |
| 10 | 3 | 0 | 2 | 0 | 197 | 71 | 36.04 | 2166 | 27.42 | 8.62 | 3.39 | 2.24 | 15.53 | Yes |
| 10 | 4 | 0 | 2 | 1 | 207 | 64 | 30.92 | 2166 | 27.42 | 3.49 | 3.19 | -2.41 | 10.09 | No |
| 10 | 5 | 0 | 3 | 0 | 201 | 85 | 42.29 | 2166 | 27.42 | 14.86 | 3.45 | 8.24 | 21.78 | Yes |
| 10 | 6 | 0 | 3 | 1 | 179 | 58 | 32.40 | 2166 | 27.42 | 4.98 | 3.47 | -1.45 | 12.14 | No |
| 10 | 7 | 1 | 1 | 0 | 164 | 36 | 21.95 | 2166 | 27.42 | -5.47 | 3.21 | -11.13 | 1.46 | No |
| 10 | 8 | 1 | 1 | 1 | 168 | 27 | 16.07 | 2166 | 27.42 | -11.35 | 2.83 | -16.14 | -5.05 | Yes |
| 10 | 9 | 1 | 2 | 0 | 134 | 37 | 27.61 | 2166 | 27.42 | 0.19 | 3.82 | -6.68 | 8.30 | No |
| 10 | 10 | 1 | 2 | 1 | 172 | 36 | 20.93 | 2166 | 27.42 | -6.49 | 3.08 | -11.90 | 0.19 | No |
| 10 | 11 | 1 | 3 | 0 | 147 | 40 | 27.21 | 2166 | 27.42 | -0.21 | 3.64 | -6.76 | 7.49 | No |
| 10 | 12 | 1 | 3 | 1 | 180 | 45 | 25.00 | 2166 | 27.42 | -2.42 | 3.20 | -8.18 | 4.38 | No |
| 11 | 1 | 0 | 1 | 0 | 108 | 27 | 25.00 | 1521 | 22.62 | 2.38 | 4.12 | -4.83 | 11.31 | No |
| 11 | 2 | 0 | 1 | 1 | 217 | 41 | 18.89 | 1521 | 22.62 | -3.72 | 2.65 | -8.37 | 2.01 | No |
| 11 | 3 | 0 | 2 | 0 | 86 | 28 | 32.56 | 1521 | 22.62 | 9.94 | 4.96 | 0.97 | 20.41 | Yes |
| 11 | 4 | 0 | 2 | 1 | 220 | 50 | 22.73 | 1521 | 22.62 | 0.11 | 2.81 | -4.93 | 6.09 | No |
| 11 | 5 | 0 | 3 | 0 | 107 | 34 | 31.78 | 1521 | 22.62 | 9.16 | 4.43 | 1.10 | 18.48 | Yes |
| 11 | 6 | 0 | 3 | 1 | 162 | 52 | 32.10 | 1521 | 22.62 | 9.48 | 3.63 | 2.78 | 17.01 | Yes |
| 11 | 7 | 1 | 1 | 0 | 75 | 18 | 24.00 | 1521 | 22.62 | 1.38 | 4.85 | -6.86 | 12.16 | No |
| 11 | 8 | 1 | 1 | 1 | 152 | 17 | 11.18 | 1521 | 22.62 | -11.43 | 2.57 | -15.52 | -5.44 | Yes |
| 11 | 9 | 1 | 2 | 0 | 49 | 10 | 20.41 | 1521 | 22.62 | -2.21 | 5.65 | -11.13 | 11.02 | No |
| 11 | 10 | 1 | 2 | 1 | 140 | 25 | 17.86 | 1521 | 22.62 | -4.76 | 3.22 | -10.22 | 2.42 | No |
| 11 | 11 | 1 | 3 | 0 | 67 | 18 | 26.87 | 1521 | 22.62 | 4.25 | 5.31 | -4.89 | 15.90 | No |
| 11 | 12 | 1 | 3 | 1 | 138 | 24 | 17.39 | 1521 | 22.62 | -5.23 | 3.21 | -10.64 | 1.96 | No |
| 12 | 1 | 0 | 1 | 0 | 213 | 55 | 25.82 | 2161 | 25.78 | 0.05 | 2.98 | -5.37 | 6.32 | No |
| 12 | 2 | 0 | 1 | 1 | 202 | 38 | 18.81 | 2161 | 25.78 | -6.96 | 2.74 | -11.75 | -1.01 | Yes |
| 12 | 3 | 0 | 2 | 0 | 173 | 63 | 36.42 | 2161 | 25.78 | 10.64 | 3.62 | 3.84 | 18.03 | Yes |
| 12 | 4 | 0 | 2 | 1 | 230 | 54 | 23.48 | 2161 | 25.78 | -2.30 | 2.78 | -7.31 | 3.59 | No |
| 12 | 5 | 0 | 3 | 0 | 216 | 87 | 40.28 | 2161 | 25.78 | 14.50 | 3.31 | 8.19 | 21.16 | Yes |

| **Area** | **SD** | **Sex** | **Age** | **Income** | **n_ASD** | **Events** | **p_ASD (%)** | **n_area** | **p_area (%)** | **Dev_A,SD (pp)** | **SE (pp)** | **95% CI low (pp)** | **95% CI high (pp)** | **Significant** |
| --- | --- | --- | --- | --- | --- | --- | --- | --- | --- | --- | --- | --- | --- | --- |
| 12 | 6 | 0 | 3 | 1 | 197 | 53 | 26.90 | 2161 | 25.78 | 1.13 | 3.14 | -4.58 | 7.72 | No |
| 12 | 7 | 1 | 1 | 0 | 160 | 31 | 19.38 | 2161 | 25.78 | -6.40 | 3.11 | -11.78 | 0.41 | No |
| 12 | 8 | 1 | 1 | 1 | 171 | 26 | 15.20 | 2161 | 25.78 | -10.57 | 2.74 | -15.18 | -4.43 | Yes |
| 12 | 9 | 1 | 2 | 0 | 114 | 28 | 24.56 | 2161 | 25.78 | -1.21 | 3.99 | -8.20 | 7.43 | No |
| 12 | 10 | 1 | 2 | 1 | 155 | 38 | 24.52 | 2161 | 25.78 | -1.26 | 3.43 | -7.36 | 6.08 | No |
| 12 | 11 | 1 | 3 | 0 | 151 | 45 | 29.80 | 2161 | 25.78 | 4.03 | 3.68 | -2.69 | 11.75 | No |
| 12 | 12 | 1 | 3 | 1 | 179 | 39 | 21.79 | 2161 | 25.78 | -3.99 | 3.07 | -9.41 | 2.62 | No |
| 13 | 1 | 0 | 1 | 0 | 111 | 27 | 24.32 | 1682 | 26.40 | -2.07 | 4.03 | -9.11 | 6.68 | No |
| 13 | 2 | 0 | 1 | 1 | 195 | 47 | 24.10 | 1682 | 26.40 | -2.29 | 3.04 | -7.76 | 4.17 | No |
| 13 | 3 | 0 | 2 | 0 | 116 | 44 | 37.93 | 1682 | 26.40 | 11.53 | 4.44 | 3.22 | 20.62 | Yes |
| 13 | 4 | 0 | 2 | 1 | 187 | 59 | 31.55 | 1682 | 26.40 | 5.15 | 3.37 | -1.08 | 12.13 | No |
| 13 | 5 | 0 | 3 | 0 | 156 | 63 | 40.38 | 1682 | 26.40 | 13.99 | 3.88 | 6.61 | 21.83 | Yes |
| 13 | 6 | 0 | 3 | 1 | 206 | 60 | 29.13 | 1682 | 26.40 | 2.73 | 3.14 | -3.05 | 9.27 | No |
| 13 | 7 | 1 | 1 | 0 | 96 | 20 | 20.83 | 1682 | 26.40 | -5.56 | 4.10 | -12.49 | 3.60 | No |
| 13 | 8 | 1 | 1 | 1 | 142 | 15 | 10.56 | 1682 | 26.40 | -15.83 | 2.60 | -19.89 | -9.70 | Yes |
| 13 | 9 | 1 | 2 | 0 | 64 | 22 | 34.38 | 1682 | 26.40 | 7.98 | 5.78 | -2.47 | 20.20 | No |
| 13 | 10 | 1 | 2 | 1 | 146 | 25 | 17.12 | 1682 | 26.40 | -9.27 | 3.11 | -14.52 | -2.34 | Yes |
| 13 | 11 | 1 | 3 | 0 | 78 | 21 | 26.92 | 1682 | 26.40 | 0.53 | 4.93 | -8.06 | 11.28 | No |
| 13 | 12 | 1 | 3 | 1 | 185 | 41 | 22.16 | 1682 | 26.40 | -4.23 | 3.04 | -9.62 | 2.28 | No |
| 14 | 1 | 0 | 1 | 0 | 200 | 46 | 23.00 | 2252 | 26.87 | -3.87 | 2.96 | -9.16 | 2.44 | No |
| 14 | 2 | 0 | 1 | 1 | 299 | 74 | 24.75 | 2252 | 26.87 | -2.12 | 2.49 | -6.67 | 3.08 | No |
| 14 | 3 | 0 | 2 | 0 | 153 | 58 | 37.91 | 2252 | 26.87 | 11.04 | 3.88 | 3.74 | 18.94 | Yes |
| 14 | 4 | 0 | 2 | 1 | 229 | 62 | 27.07 | 2252 | 26.87 | 0.21 | 2.92 | -5.13 | 6.31 | No |
| 14 | 5 | 0 | 3 | 0 | 177 | 74 | 41.81 | 2252 | 26.87 | 14.94 | 3.67 | 7.93 | 22.31 | Yes |
| 14 | 6 | 0 | 3 | 1 | 173 | 48 | 27.75 | 2252 | 26.87 | 0.88 | 3.38 | -5.25 | 7.98 | No |
| 14 | 7 | 1 | 1 | 0 | 204 | 50 | 24.51 | 2252 | 26.87 | -2.36 | 2.99 | -7.75 | 3.98 | No |
| 14 | 8 | 1 | 1 | 1 | 167 | 28 | 16.77 | 2252 | 26.87 | -10.10 | 2.88 | -15.00 | -3.70 | Yes |
| 14 | 9 | 1 | 2 | 0 | 139 | 44 | 31.65 | 2252 | 26.87 | 4.79 | 3.90 | -2.36 | 12.93 | No |
| 14 | 10 | 1 | 2 | 1 | 219 | 44 | 20.09 | 2252 | 26.87 | -6.77 | 2.70 | -11.54 | -0.97 | Yes |
| 14 | 11 | 1 | 3 | 0 | 133 | 46 | 34.59 | 2252 | 26.87 | 7.72 | 4.07 | 0.17 | 16.14 | Yes |
| 14 | 12 | 1 | 3 | 1 | 159 | 31 | 19.50 | 2252 | 26.87 | -7.37 | 3.13 | -12.78 | -0.52 | Yes |
| 15 | 1 | 0 | 1 | 0 | 153 | 37 | 24.18 | 1652 | 23.00 | 1.18 | 3.43 | -4.92 | 8.54 | No |
| 15 | 2 | 0 | 1 | 1 | 171 | 37 | 21.64 | 1652 | 23.00 | -1.36 | 3.13 | -6.88 | 5.39 | No |
| 15 | 3 | 0 | 2 | 0 | 102 | 37 | 36.27 | 1652 | 23.00 | 13.27 | 4.68 | 4.60 | 22.94 | Yes |
| 15 | 4 | 0 | 2 | 1 | 162 | 42 | 25.93 | 1652 | 23.00 | 2.92 | 3.41 | -3.21 | 10.17 | No |

| **Area** | **SD** | **Sex** | **Age** | **Income** | **n_ASD** | **Events** | **p_ASD (%)** | **n_area** | **p_area (%)** | **Dev_A,SD (pp)** | **SE (pp)** | **95% CI low (pp)** | **95% CI high (pp)** | **Significant** |
| --- | --- | --- | --- | --- | --- | --- | --- | --- | --- | --- | --- | --- | --- | --- |
| 15 | 5 | 0 | 3 | 0 | 163 | 50 | 30.67 | 1652 | 23.00 | 7.67 | 3.58 | 1.11 | 15.13 | Yes |
| 15 | 6 | 0 | 3 | 1 | 194 | 42 | 21.65 | 1652 | 23.00 | -1.35 | 2.94 | -6.57 | 4.96 | No |
| 15 | 7 | 1 | 1 | 0 | 115 | 23 | 20.00 | 1652 | 23.00 | -3.00 | 3.70 | -9.29 | 5.22 | No |
| 15 | 8 | 1 | 1 | 1 | 135 | 15 | 11.11 | 1652 | 23.00 | -11.89 | 2.72 | -16.15 | -5.48 | Yes |
| 15 | 9 | 1 | 2 | 0 | 85 | 25 | 29.41 | 1652 | 23.00 | 6.41 | 4.86 | -2.22 | 16.82 | No |
| 15 | 10 | 1 | 2 | 1 | 120 | 17 | 14.17 | 1652 | 23.00 | -8.84 | 3.18 | -13.97 | -1.48 | Yes |
| 15 | 11 | 1 | 3 | 0 | 97 | 24 | 24.74 | 1652 | 23.00 | 1.74 | 4.33 | -5.78 | 11.18 | No |
| 15 | 12 | 1 | 3 | 1 | 155 | 31 | 20.00 | 1652 | 23.00 | -3.00 | 3.20 | -8.54 | 3.99 | No |
| 16 | 1 | 0 | 1 | 0 | 163 | 52 | 31.90 | 1989 | 26.65 | 5.26 | 3.61 | -1.41 | 12.76 | No |
| 16 | 2 | 0 | 1 | 1 | 202 | 45 | 22.28 | 1989 | 26.65 | -4.37 | 2.91 | -9.56 | 1.86 | No |
| 16 | 3 | 0 | 2 | 0 | 132 | 41 | 31.06 | 1989 | 26.65 | 4.41 | 3.98 | -2.85 | 12.75 | No |
| 16 | 4 | 0 | 2 | 1 | 189 | 52 | 27.51 | 1989 | 26.65 | 0.87 | 3.22 | -5.00 | 7.63 | No |
| 16 | 5 | 0 | 3 | 0 | 226 | 84 | 37.17 | 1989 | 26.65 | 10.52 | 3.19 | 4.48 | 16.99 | Yes |
| 16 | 6 | 0 | 3 | 1 | 218 | 69 | 31.65 | 1989 | 26.65 | 5.00 | 3.13 | -0.81 | 11.45 | No |
| 16 | 7 | 1 | 1 | 0 | 162 | 40 | 24.69 | 1989 | 26.65 | -1.96 | 3.36 | -7.96 | 5.22 | No |
| 16 | 8 | 1 | 1 | 1 | 115 | 16 | 13.91 | 1989 | 26.65 | -12.73 | 3.23 | -17.90 | -5.24 | Yes |
| 16 | 9 | 1 | 2 | 0 | 127 | 27 | 21.26 | 1989 | 26.65 | -5.39 | 3.60 | -11.60 | 2.52 | No |
| 16 | 10 | 1 | 2 | 1 | 156 | 29 | 18.59 | 1989 | 26.65 | -8.06 | 3.10 | -13.38 | -1.22 | Yes |
| 16 | 11 | 1 | 3 | 0 | 117 | 37 | 31.62 | 1989 | 26.65 | 4.98 | 4.24 | -2.75 | 13.87 | No |
| 16 | 12 | 1 | 3 | 1 | 182 | 38 | 20.88 | 1989 | 26.65 | -5.77 | 3.00 | -11.04 | 0.71 | No |
| 17 | 1 | 0 | 1 | 0 | 236 | 69 | 29.24 | 2348 | 26.75 | 2.49 | 2.94 | -2.94 | 8.59 | No |
| 17 | 2 | 0 | 1 | 1 | 240 | 45 | 18.75 | 2348 | 26.75 | -8.00 | 2.51 | -12.43 | -2.58 | Yes |
| 17 | 3 | 0 | 2 | 0 | 210 | 82 | 39.05 | 2348 | 26.75 | 12.30 | 3.34 | 5.96 | 19.04 | Yes |
| 17 | 4 | 0 | 2 | 1 | 245 | 67 | 27.35 | 2348 | 26.75 | 0.60 | 2.83 | -4.60 | 6.50 | No |
| 17 | 5 | 0 | 3 | 0 | 210 | 101 | 48.10 | 2348 | 26.75 | 21.35 | 3.42 | 14.69 | 28.08 | Yes |
| 17 | 6 | 0 | 3 | 1 | 167 | 46 | 27.54 | 2348 | 26.75 | 0.80 | 3.43 | -5.41 | 8.02 | No |
| 17 | 7 | 1 | 1 | 0 | 203 | 38 | 18.72 | 2348 | 26.75 | -8.03 | 2.73 | -12.79 | -2.10 | Yes |
| 17 | 8 | 1 | 1 | 1 | 182 | 27 | 14.84 | 2348 | 26.75 | -11.91 | 2.63 | -16.35 | -6.02 | Yes |
| 17 | 9 | 1 | 2 | 0 | 180 | 48 | 26.67 | 2348 | 26.75 | -0.08 | 3.27 | -6.00 | 6.82 | No |
| 17 | 10 | 1 | 2 | 1 | 176 | 34 | 19.32 | 2348 | 26.75 | -7.43 | 2.96 | -12.58 | -0.97 | Yes |
| 17 | 11 | 1 | 3 | 0 | 163 | 42 | 25.77 | 2348 | 26.75 | -0.98 | 3.40 | -7.08 | 6.24 | No |
| 17 | 12 | 1 | 3 | 1 | 136 | 29 | 21.32 | 2348 | 26.75 | -5.42 | 3.49 | -11.47 | 2.20 | No |
| 18 | 1 | 0 | 1 | 0 | 128 | 33 | 25.78 | 1726 | 25.26 | 0.52 | 3.83 | -6.27 | 8.73 | No |
| 18 | 2 | 0 | 1 | 1 | 205 | 31 | 15.12 | 1726 | 25.26 | -10.14 | 2.50 | -14.40 | -4.60 | Yes |
| 18 | 3 | 0 | 2 | 0 | 113 | 43 | 38.05 | 1726 | 25.26 | 12.79 | 4.50 | 4.37 | 22.00 | Yes |

| **Area** | **SD** | **Sex** | **Age** | **Income** | **n_ASD** | **Events** | **p_ASD (%)** | **n_area** | **p_area (%)** | **Dev_A,SD (pp)** | **SE (pp)** | **95% CI low (pp)** | **95% CI high (pp)** | **Significant** |
| --- | --- | --- | --- | --- | --- | --- | --- | --- | --- | --- | --- | --- | --- | --- |
| 18 | 4 | 0 | 2 | 1 | 212 | 57 | 26.89 | 1726 | 25.26 | 1.63 | 3.03 | -3.89 | 7.97 | No |
| 18 | 5 | 0 | 3 | 0 | 128 | 62 | 48.44 | 1726 | 25.26 | 23.18 | 4.35 | 14.69 | 31.75 | Yes |
| 18 | 6 | 0 | 3 | 1 | 207 | 69 | 33.33 | 1726 | 25.26 | 8.07 | 3.25 | 2.01 | 14.75 | Yes |
| 18 | 7 | 1 | 1 | 0 | 110 | 28 | 25.45 | 1726 | 25.26 | 0.19 | 4.10 | -7.02 | 9.07 | No |
| 18 | 8 | 1 | 1 | 1 | 118 | 12 | 10.17 | 1726 | 25.26 | -15.09 | 2.81 | -19.35 | -8.32 | Yes |
| 18 | 9 | 1 | 2 | 0 | 85 | 22 | 25.88 | 1726 | 25.26 | 0.62 | 4.68 | -7.50 | 10.83 | No |
| 18 | 10 | 1 | 2 | 1 | 144 | 17 | 11.81 | 1726 | 25.26 | -13.46 | 2.70 | -17.76 | -7.17 | Yes |
| 18 | 11 | 1 | 3 | 0 | 86 | 17 | 19.77 | 1726 | 25.26 | -5.49 | 4.25 | -12.54 | 4.14 | No |
| 18 | 12 | 1 | 3 | 1 | 190 | 45 | 23.68 | 1726 | 25.26 | -1.58 | 3.07 | -7.06 | 4.95 | No |
| 19 | 1 | 0 | 1 | 0 | 86 | 25 | 29.07 | 1217 | 22.93 | 6.14 | 4.81 | -2.39 | 16.47 | No |
| 19 | 2 | 0 | 1 | 1 | 157 | 26 | 16.56 | 1217 | 22.93 | -6.36 | 2.96 | -11.37 | 0.23 | No |
| 19 | 3 | 0 | 2 | 0 | 70 | 26 | 37.14 | 1217 | 22.93 | 14.22 | 5.63 | 3.85 | 25.93 | Yes |
| 19 | 4 | 0 | 2 | 1 | 117 | 27 | 23.08 | 1217 | 22.93 | 0.15 | 3.86 | -6.55 | 8.57 | No |
| 19 | 5 | 0 | 3 | 0 | 103 | 42 | 40.78 | 1217 | 22.93 | 17.85 | 4.76 | 8.86 | 27.51 | Yes |
| 19 | 6 | 0 | 3 | 1 | 160 | 54 | 33.75 | 1217 | 22.93 | 10.82 | 3.70 | 3.96 | 18.46 | Yes |
| 19 | 7 | 1 | 1 | 0 | 70 | 12 | 17.14 | 1217 | 22.93 | -5.78 | 4.47 | -12.84 | 4.69 | No |
| 19 | 8 | 1 | 1 | 1 | 117 | 13 | 11.11 | 1217 | 22.93 | -11.81 | 2.93 | -16.32 | -4.84 | Yes |
| 19 | 9 | 1 | 2 | 0 | 71 | 16 | 22.54 | 1217 | 22.93 | -0.39 | 4.88 | -8.55 | 10.59 | No |
| 19 | 10 | 1 | 2 | 1 | 83 | 10 | 12.05 | 1217 | 22.93 | -10.88 | 3.60 | -16.25 | -2.15 | Yes |
| 19 | 11 | 1 | 3 | 0 | 70 | 12 | 17.14 | 1217 | 22.93 | -5.78 | 4.47 | -12.84 | 4.69 | No |
| 19 | 12 | 1 | 3 | 1 | 113 | 16 | 14.16 | 1217 | 22.93 | -8.77 | 3.28 | -14.02 | -1.16 | Yes |
| 20 | 1 | 0 | 1 | 0 | 178 | 33 | 18.54 | 1796 | 24.28 | -5.74 | 2.90 | -10.76 | 0.62 | No |
| 20 | 2 | 0 | 1 | 1 | 202 | 47 | 23.27 | 1796 | 24.28 | -1.01 | 2.96 | -6.30 | 5.28 | No |
| 20 | 3 | 0 | 2 | 0 | 152 | 55 | 36.18 | 1796 | 24.28 | 11.91 | 3.85 | 4.70 | 19.80 | Yes |
| 20 | 4 | 0 | 2 | 1 | 159 | 39 | 24.53 | 1796 | 24.28 | 0.25 | 3.39 | -5.78 | 7.49 | No |
| 20 | 5 | 0 | 3 | 0 | 144 | 56 | 38.89 | 1796 | 24.28 | 14.61 | 4.01 | 7.04 | 22.77 | Yes |
| 20 | 6 | 0 | 3 | 1 | 141 | 41 | 29.08 | 1796 | 24.28 | 4.80 | 3.78 | -2.06 | 12.77 | No |
| 20 | 7 | 1 | 1 | 0 | 148 | 33 | 22.30 | 1796 | 24.28 | -1.98 | 3.40 | -7.94 | 5.38 | No |
| 20 | 8 | 1 | 1 | 1 | 168 | 24 | 14.29 | 1796 | 24.28 | -9.99 | 2.70 | -14.48 | -3.90 | Yes |
| 20 | 9 | 1 | 2 | 0 | 147 | 37 | 25.17 | 1796 | 24.28 | 0.89 | 3.55 | -5.43 | 8.48 | No |
| 20 | 10 | 1 | 2 | 1 | 130 | 18 | 13.85 | 1796 | 24.28 | -10.43 | 3.03 | -15.34 | -3.45 | Yes |
| 20 | 11 | 1 | 3 | 0 | 114 | 30 | 26.32 | 1796 | 24.28 | 2.04 | 4.08 | -5.18 | 10.80 | No |
| 20 | 12 | 1 | 3 | 1 | 113 | 23 | 20.35 | 1796 | 24.28 | -3.92 | 3.76 | -10.31 | 4.42 | No |
| 21 | 1 | 0 | 1 | 0 | 193 | 64 | 33.16 | 1842 | 30.40 | 2.76 | 3.36 | -3.50 | 9.67 | No |
| 21 | 2 | 0 | 1 | 1 | 158 | 37 | 23.42 | 1842 | 30.40 | -6.98 | 3.34 | -12.91 | 0.20 | No |

| **Area** | **SD** | **Sex** | **Age** | **Income** | **n_ASD** | **Events** | **p_ASD (%)** | **n_area** | **p_area (%)** | **Dev_A,SD (pp)** | **SE (pp)** | **95% CI low (pp)** | **95% CI high (pp)** | **Significant** |
| --- | --- | --- | --- | --- | --- | --- | --- | --- | --- | --- | --- | --- | --- | --- |
| 21 | 3 | 0 | 2 | 0 | 177 | 91 | 51.41 | 1842 | 30.40 | 21.01 | 3.72 | 13.70 | 28.27 | Yes |
| 21 | 4 | 0 | 2 | 1 | 163 | 42 | 25.77 | 1842 | 30.40 | -4.63 | 3.40 | -10.74 | 2.58 | No |
| 21 | 5 | 0 | 3 | 0 | 180 | 84 | 46.67 | 1842 | 30.40 | 16.26 | 3.68 | 9.12 | 23.55 | Yes |
| 21 | 6 | 0 | 3 | 1 | 108 | 42 | 38.89 | 1842 | 30.40 | 8.49 | 4.61 | -0.17 | 17.91 | No |
| 21 | 7 | 1 | 1 | 0 | 199 | 46 | 23.12 | 1842 | 30.40 | -7.29 | 2.97 | -12.60 | -0.95 | Yes |
| 21 | 8 | 1 | 1 | 1 | 134 | 21 | 15.67 | 1842 | 30.40 | -14.73 | 3.13 | -19.92 | -7.63 | Yes |
| 21 | 9 | 1 | 2 | 0 | 148 | 46 | 31.08 | 1842 | 30.40 | 0.68 | 3.76 | -6.22 | 8.54 | No |
| 21 | 10 | 1 | 2 | 1 | 120 | 21 | 17.50 | 1842 | 30.40 | -12.90 | 3.45 | -18.66 | -5.13 | Yes |
| 21 | 11 | 1 | 3 | 0 | 156 | 51 | 32.69 | 1842 | 30.40 | 2.29 | 3.72 | -4.58 | 9.99 | No |
| 21 | 12 | 1 | 3 | 1 | 106 | 15 | 14.15 | 1842 | 30.40 | -16.25 | 3.39 | -21.63 | -8.36 | Yes |
| 22 | 1 | 0 | 1 | 0 | 111 | 30 | 27.03 | 1293 | 22.89 | 4.13 | 4.16 | -3.26 | 13.06 | No |
| 22 | 2 | 0 | 1 | 1 | 172 | 33 | 19.19 | 1293 | 22.89 | -3.71 | 2.99 | -8.89 | 2.83 | No |
| 22 | 3 | 0 | 2 | 0 | 66 | 20 | 30.30 | 1293 | 22.89 | 7.41 | 5.53 | -2.34 | 19.33 | No |
| 22 | 4 | 0 | 2 | 1 | 162 | 43 | 26.54 | 1293 | 22.89 | 3.65 | 3.44 | -2.55 | 10.94 | No |
| 22 | 5 | 0 | 3 | 0 | 96 | 33 | 34.38 | 1293 | 22.89 | 11.48 | 4.76 | 2.75 | 21.42 | Yes |
| 22 | 6 | 0 | 3 | 1 | 107 | 33 | 30.84 | 1293 | 22.89 | 7.95 | 4.40 | -0.01 | 17.24 | No |
| 22 | 7 | 1 | 1 | 0 | 98 | 19 | 19.39 | 1293 | 22.89 | -3.50 | 3.96 | -10.11 | 5.41 | No |
| 22 | 8 | 1 | 1 | 1 | 106 | 10 | 9.43 | 1293 | 22.89 | -13.46 | 2.88 | -17.69 | -6.39 | Yes |
| 22 | 9 | 1 | 2 | 0 | 70 | 24 | 34.29 | 1293 | 22.89 | 11.39 | 5.54 | 1.35 | 23.07 | Yes |
| 22 | 10 | 1 | 2 | 1 | 131 | 15 | 11.45 | 1293 | 22.89 | -11.44 | 2.80 | -15.83 | -4.86 | Yes |
| 22 | 11 | 1 | 3 | 0 | 71 | 22 | 30.99 | 1293 | 22.89 | 8.09 | 5.37 | -1.45 | 19.59 | No |
| 22 | 12 | 1 | 3 | 1 | 103 | 14 | 13.59 | 1293 | 22.89 | -9.30 | 3.38 | -14.62 | -1.36 | Yes |
| 23 | 1 | 0 | 1 | 0 | 183 | 53 | 28.96 | 1691 | 27.62 | 1.34 | 3.33 | -4.74 | 8.30 | No |
| 23 | 2 | 0 | 1 | 1 | 146 | 30 | 20.55 | 1691 | 27.62 | -7.07 | 3.32 | -12.83 | 0.20 | No |
| 23 | 3 | 0 | 2 | 0 | 170 | 75 | 44.12 | 1691 | 27.62 | 16.50 | 3.77 | 9.25 | 24.01 | Yes |
| 23 | 4 | 0 | 2 | 1 | 153 | 43 | 28.10 | 1691 | 27.62 | 0.49 | 3.60 | -6.03 | 8.08 | No |
| 23 | 5 | 0 | 3 | 0 | 175 | 79 | 45.14 | 1691 | 27.62 | 17.53 | 3.72 | 10.34 | 24.92 | Yes |
| 23 | 6 | 0 | 3 | 1 | 133 | 36 | 27.07 | 1691 | 27.62 | -0.55 | 3.81 | -7.38 | 7.57 | No |
| 23 | 7 | 1 | 1 | 0 | 158 | 30 | 18.99 | 1691 | 27.62 | -8.63 | 3.11 | -13.98 | -1.81 | Yes |
| 23 | 8 | 1 | 1 | 1 | 100 | 11 | 11.00 | 1691 | 27.62 | -16.62 | 3.16 | -21.36 | -8.99 | Yes |
| 23 | 9 | 1 | 2 | 0 | 138 | 36 | 26.09 | 1691 | 27.62 | -1.53 | 3.70 | -8.14 | 6.37 | No |
| 23 | 10 | 1 | 2 | 1 | 115 | 19 | 16.52 | 1691 | 27.62 | -11.10 | 3.45 | -16.78 | -3.25 | Yes |
| 23 | 11 | 1 | 3 | 0 | 112 | 34 | 30.36 | 1691 | 27.62 | 2.74 | 4.28 | -5.01 | 11.79 | No |
| 23 | 12 | 1 | 3 | 1 | 108 | 21 | 19.44 | 1691 | 27.62 | -8.17 | 3.78 | -14.53 | 0.29 | No |
| 24 | 1 | 0 | 1 | 0 | 95 | 33 | 34.74 | 1266 | 28.83 | 5.91 | 4.80 | -2.91 | 15.90 | No |

| **Area** | **SD** | **Sex** | **Age** | **Income** | **n_ASD** | **Events** | **p_ASD (%)** | **n_area** | **p_area (%)** | **Dev_A,SD (pp)** | **SE (pp)** | **95% CI low (pp)** | **95% CI high (pp)** | **Significant** |
| --- | --- | --- | --- | --- | --- | --- | --- | --- | --- | --- | --- | --- | --- | --- |
| 24 | 2 | 0 | 1 | 1 | 140 | 26 | 18.57 | 1266 | 28.83 | -10.26 | 3.27 | -15.83 | -3.01 | Yes |
| 24 | 3 | 0 | 2 | 0 | 75 | 38 | 50.67 | 1266 | 28.83 | 21.84 | 5.63 | 10.77 | 32.84 | Yes |
| 24 | 4 | 0 | 2 | 1 | 123 | 28 | 22.76 | 1266 | 28.83 | -6.07 | 3.75 | -12.59 | 2.10 | No |
| 24 | 5 | 0 | 3 | 0 | 94 | 40 | 42.55 | 1266 | 28.83 | 13.72 | 5.00 | 4.21 | 23.82 | Yes |
| 24 | 6 | 0 | 3 | 1 | 125 | 52 | 41.60 | 1266 | 28.83 | 12.77 | 4.34 | 4.50 | 21.53 | Yes |
| 24 | 7 | 1 | 1 | 0 | 113 | 27 | 23.89 | 1266 | 28.83 | -4.94 | 3.97 | -11.86 | 3.70 | No |
| 24 | 8 | 1 | 1 | 1 | 110 | 11 | 10.00 | 1266 | 28.83 | -18.83 | 2.89 | -23.16 | -11.81 | Yes |
| 24 | 9 | 1 | 2 | 0 | 105 | 38 | 36.19 | 1266 | 28.83 | 7.36 | 4.61 | -1.19 | 16.89 | No |
| 24 | 10 | 1 | 2 | 1 | 105 | 22 | 20.95 | 1266 | 28.83 | -7.88 | 3.94 | -14.57 | 0.86 | No |
| 24 | 11 | 1 | 3 | 0 | 84 | 29 | 34.52 | 1266 | 28.83 | 5.69 | 5.08 | -3.60 | 16.34 | No |
| 24 | 12 | 1 | 3 | 1 | 97 | 21 | 21.65 | 1266 | 28.83 | -7.18 | 4.14 | -14.21 | 2.01 | No |
| 25 | 1 | 0 | 1 | 0 | 111 | 20 | 18.02 | 1424 | 21.98 | -3.96 | 3.63 | -10.00 | 4.22 | No |
| 25 | 2 | 0 | 1 | 1 | 180 | 25 | 13.89 | 1424 | 21.98 | -8.09 | 2.58 | -12.39 | -2.28 | Yes |
| 25 | 3 | 0 | 2 | 0 | 77 | 25 | 32.47 | 1424 | 21.98 | 10.49 | 5.23 | 1.08 | 21.56 | Yes |
| 25 | 4 | 0 | 2 | 1 | 184 | 46 | 25.00 | 1424 | 21.98 | 3.02 | 3.17 | -2.68 | 9.74 | No |
| 25 | 5 | 0 | 3 | 0 | 108 | 35 | 32.41 | 1424 | 21.98 | 10.43 | 4.44 | 2.34 | 19.73 | Yes |
| 25 | 6 | 0 | 3 | 1 | 152 | 50 | 32.89 | 1424 | 21.98 | 10.91 | 3.77 | 3.95 | 18.72 | Yes |
| 25 | 7 | 1 | 1 | 0 | 78 | 24 | 30.77 | 1424 | 21.98 | 8.79 | 5.12 | -0.35 | 19.73 | No |
| 25 | 8 | 1 | 1 | 1 | 130 | 15 | 11.54 | 1424 | 21.98 | -10.44 | 2.82 | -14.86 | -3.81 | Yes |
| 25 | 9 | 1 | 2 | 0 | 56 | 16 | 28.57 | 1424 | 21.98 | 6.59 | 5.88 | -3.56 | 19.50 | No |
| 25 | 10 | 1 | 2 | 1 | 137 | 17 | 12.41 | 1424 | 21.98 | -9.57 | 2.83 | -14.09 | -3.01 | Yes |
| 25 | 11 | 1 | 3 | 0 | 67 | 14 | 20.90 | 1424 | 21.98 | -1.08 | 4.90 | -9.10 | 10.09 | No |
| 25 | 12 | 1 | 3 | 1 | 144 | 26 | 18.06 | 1424 | 21.98 | -3.92 | 3.19 | -9.35 | 3.16 | No |

**Table B. Dev_A,SD with 95% CIs — Private GP choice (300 A-SD strata).**

**Overall prevalence: 22.10% | N = 43,291 across 300 A-SD strata Significantly deviating strata (95% CI does not cross zero): 122 of 300 (40.7%)**

| **Area** | **SD** | **Sex** | **Age** | **Income** | **n_ASD** | **Events** | **p_ASD (%)** | **n_area** | **p_area (%)** | **Dev_A,SD (pp)** | **SE (pp)** | **95% CI low (pp)** | **95% CI high (pp)** | **Significant** |
| --- | --- | --- | --- | --- | --- | --- | --- | --- | --- | --- | --- | --- | --- | --- |
| 1 | 1 | 0 | 1 | 0 | 79 | 17 | 21.52 | 1186 | 17.79 | 3.73 | 4.57 | -3.90 | 14.00 | No |
| 1 | 2 | 0 | 1 | 1 | 167 | 24 | 14.37 | 1186 | 17.79 | -3.42 | 2.71 | -7.94 | 2.70 | No |
| 1 | 3 | 0 | 2 | 0 | 66 | 19 | 28.79 | 1186 | 17.79 | 11.00 | 5.45 | 1.48 | 22.85 | Yes |
| 1 | 4 | 0 | 2 | 1 | 163 | 19 | 11.66 | 1186 | 17.79 | -6.13 | 2.52 | -10.20 | -0.30 | Yes |
| 1 | 5 | 0 | 3 | 0 | 74 | 21 | 28.38 | 1186 | 17.79 | 10.59 | 5.14 | 1.58 | 21.73 | Yes |
| 1 | 6 | 0 | 3 | 1 | 138 | 13 | 9.42 | 1186 | 17.79 | -8.37 | 2.52 | -12.20 | -2.34 | Yes |
| 1 | 7 | 1 | 1 | 0 | 51 | 8 | 15.69 | 1186 | 17.79 | -2.10 | 5.06 | -9.62 | 10.22 | No |
| 1 | 8 | 1 | 1 | 1 | 98 | 17 | 17.35 | 1186 | 17.79 | -0.44 | 3.80 | -6.67 | 8.24 | No |
| 1 | 9 | 1 | 2 | 0 | 38 | 13 | 34.21 | 1186 | 17.79 | 16.42 | 7.37 | 3.42 | 32.32 | Yes |
| 1 | 10 | 1 | 2 | 1 | 127 | 22 | 17.32 | 1186 | 17.79 | -0.47 | 3.34 | -6.06 | 7.05 | No |
| 1 | 11 | 1 | 3 | 0 | 57 | 17 | 29.82 | 1186 | 17.79 | 12.03 | 5.90 | 1.74 | 24.87 | Yes |
| 1 | 12 | 1 | 3 | 1 | 128 | 21 | 16.41 | 1186 | 17.79 | -1.38 | 3.26 | -6.80 | 5.99 | No |
| 2 | 1 | 0 | 1 | 0 | 163 | 14 | 8.59 | 1772 | 9.31 | -0.72 | 2.22 | -4.13 | 4.59 | No |
| 2 | 2 | 0 | 1 | 1 | 182 | 21 | 11.54 | 1772 | 9.31 | 2.23 | 2.38 | -1.64 | 7.68 | No |
| 2 | 3 | 0 | 2 | 0 | 129 | 13 | 10.08 | 1772 | 9.31 | 0.77 | 2.68 | -3.33 | 7.17 | No |
| 2 | 4 | 0 | 2 | 1 | 178 | 19 | 10.67 | 1772 | 9.31 | 1.36 | 2.33 | -2.37 | 6.76 | No |
| 2 | 5 | 0 | 3 | 0 | 174 | 14 | 8.05 | 1772 | 9.31 | -1.27 | 2.09 | -4.46 | 3.74 | No |
| 2 | 6 | 0 | 3 | 1 | 191 | 17 | 8.90 | 1772 | 9.31 | -0.41 | 2.08 | -3.68 | 4.48 | No |
| 2 | 7 | 1 | 1 | 0 | 112 | 7 | 6.25 | 1772 | 9.31 | -3.06 | 2.37 | -6.25 | 3.03 | No |
| 2 | 8 | 1 | 1 | 1 | 134 | 9 | 6.72 | 1772 | 9.31 | -2.60 | 2.22 | -5.74 | 2.96 | No |
| 2 | 9 | 1 | 2 | 0 | 94 | 9 | 9.57 | 1772 | 9.31 | 0.26 | 3.08 | -4.19 | 7.89 | No |
| 2 | 10 | 1 | 2 | 1 | 131 | 14 | 10.69 | 1772 | 9.31 | 1.38 | 2.72 | -2.84 | 7.83 | No |
| 2 | 11 | 1 | 3 | 0 | 115 | 5 | 4.35 | 1772 | 9.31 | -4.96 | 2.02 | -7.44 | 0.46 | No |
| 2 | 12 | 1 | 3 | 1 | 169 | 23 | 13.61 | 1772 | 9.31 | 4.30 | 2.64 | -0.07 | 10.28 | No |
| 3 | 1 | 0 | 1 | 0 | 294 | 47 | 15.99 | 2633 | 16.26 | -0.27 | 2.14 | -4.02 | 4.36 | No |
| 3 | 2 | 0 | 1 | 1 | 286 | 32 | 11.19 | 2633 | 16.26 | -5.07 | 1.87 | -8.22 | -0.89 | Yes |
| 3 | 3 | 0 | 2 | 0 | 238 | 39 | 16.39 | 2633 | 16.26 | 0.13 | 2.40 | -4.03 | 5.36 | No |
| 3 | 4 | 0 | 2 | 1 | 246 | 41 | 16.67 | 2633 | 16.26 | 0.41 | 2.37 | -3.73 | 5.57 | No |
| 3 | 5 | 0 | 3 | 0 | 255 | 60 | 23.53 | 2633 | 16.26 | 7.27 | 2.64 | 2.48 | 12.85 | Yes |
| 3 | 6 | 0 | 3 | 1 | 214 | 42 | 19.63 | 2633 | 16.26 | 3.37 | 2.70 | -1.39 | 9.21 | No |
| 3 | 7 | 1 | 1 | 0 | 222 | 30 | 13.51 | 2633 | 16.26 | -2.74 | 2.30 | -6.62 | 2.38 | No |
| 3 | 8 | 1 | 1 | 1 | 197 | 28 | 14.21 | 2633 | 16.26 | -2.04 | 2.49 | -6.24 | 3.52 | No |

| **Area** | **SD** | **Sex** | **Age** | **Income** | **n_ASD** | **Events** | **p_ASD (%)** | **n_area** | **p_area (%)** | **Dev_A,SD (pp)** | **SE (pp)** | **95% CI low (pp)** | **95% CI high (pp)** | **Significant** |
| --- | --- | --- | --- | --- | --- | --- | --- | --- | --- | --- | --- | --- | --- | --- |
| 3 | 9 | 1 | 2 | 0 | 169 | 22 | 13.02 | 2633 | 16.26 | -3.24 | 2.59 | -7.50 | 2.67 | No |
| 3 | 10 | 1 | 2 | 1 | 175 | 22 | 12.57 | 2633 | 16.26 | -3.68 | 2.51 | -7.80 | 2.05 | No |
| 3 | 11 | 1 | 3 | 0 | 164 | 25 | 15.24 | 2633 | 16.26 | -1.01 | 2.80 | -5.71 | 5.28 | No |
| 3 | 12 | 1 | 3 | 1 | 173 | 40 | 23.12 | 2633 | 16.26 | 6.87 | 3.18 | 1.21 | 13.69 | Yes |
| 4 | 1 | 0 | 1 | 0 | 189 | 49 | 25.93 | 1994 | 35.36 | -9.43 | 3.17 | -15.15 | -2.75 | Yes |
| 4 | 2 | 0 | 1 | 1 | 230 | 78 | 33.91 | 1994 | 35.36 | -1.44 | 3.10 | -7.25 | 4.89 | No |
| 4 | 3 | 0 | 2 | 0 | 175 | 45 | 25.71 | 1994 | 35.36 | -9.64 | 3.28 | -15.55 | -2.69 | Yes |
| 4 | 4 | 0 | 2 | 1 | 196 | 79 | 40.31 | 1994 | 35.36 | 4.95 | 3.47 | -1.67 | 11.94 | No |
| 4 | 5 | 0 | 3 | 0 | 174 | 60 | 34.48 | 1994 | 35.36 | -0.87 | 3.57 | -7.53 | 6.46 | No |
| 4 | 6 | 0 | 3 | 1 | 147 | 77 | 52.38 | 1994 | 35.36 | 17.02 | 4.07 | 8.99 | 24.93 | Yes |
| 4 | 7 | 1 | 1 | 0 | 142 | 34 | 23.94 | 1994 | 35.36 | -11.41 | 3.55 | -17.69 | -3.77 | Yes |
| 4 | 8 | 1 | 1 | 1 | 172 | 60 | 34.88 | 1994 | 35.36 | -0.47 | 3.60 | -7.19 | 6.91 | No |
| 4 | 9 | 1 | 2 | 0 | 124 | 37 | 29.84 | 1994 | 35.36 | -5.52 | 4.06 | -12.87 | 3.04 | No |
| 4 | 10 | 1 | 2 | 1 | 181 | 66 | 36.46 | 1994 | 35.36 | 1.11 | 3.54 | -5.56 | 8.33 | No |
| 4 | 11 | 1 | 3 | 0 | 118 | 46 | 38.98 | 1994 | 35.36 | 3.63 | 4.42 | -4.69 | 12.64 | No |
| 4 | 12 | 1 | 3 | 1 | 146 | 74 | 50.68 | 1994 | 35.36 | 15.33 | 4.08 | 7.31 | 23.32 | Yes |
| 5 | 1 | 0 | 1 | 0 | 95 | 11 | 11.58 | 1665 | 29.19 | -17.61 | 3.31 | -22.60 | -9.63 | Yes |
| 5 | 2 | 0 | 1 | 1 | 208 | 73 | 35.10 | 1665 | 29.19 | 5.91 | 3.28 | -0.26 | 12.61 | No |
| 5 | 3 | 0 | 2 | 0 | 97 | 12 | 12.37 | 1665 | 29.19 | -16.82 | 3.36 | -21.97 | -8.80 | Yes |
| 5 | 4 | 0 | 2 | 1 | 218 | 91 | 41.74 | 1665 | 29.19 | 12.55 | 3.31 | 6.21 | 19.19 | Yes |
| 5 | 5 | 0 | 3 | 0 | 121 | 23 | 19.01 | 1665 | 29.19 | -10.18 | 3.55 | -16.18 | -2.28 | Yes |
| 5 | 6 | 0 | 3 | 1 | 203 | 68 | 33.50 | 1665 | 29.19 | 4.31 | 3.29 | -1.82 | 11.05 | No |
| 5 | 7 | 1 | 1 | 0 | 90 | 4 | 4.44 | 1665 | 29.19 | -24.74 | 2.33 | -27.45 | -18.31 | Yes |
| 5 | 8 | 1 | 1 | 1 | 138 | 47 | 34.06 | 1665 | 29.19 | 4.87 | 3.99 | -2.51 | 13.11 | No |
| 5 | 9 | 1 | 2 | 0 | 76 | 9 | 11.84 | 1665 | 29.19 | -17.35 | 3.74 | -22.83 | -8.19 | Yes |
| 5 | 10 | 1 | 2 | 1 | 158 | 69 | 43.67 | 1665 | 29.19 | 14.48 | 3.90 | 6.99 | 22.27 | Yes |
| 5 | 11 | 1 | 3 | 0 | 90 | 11 | 12.22 | 1665 | 29.19 | -16.97 | 3.47 | -22.23 | -8.62 | Yes |
| 5 | 12 | 1 | 3 | 1 | 171 | 68 | 39.77 | 1665 | 29.19 | 10.58 | 3.70 | 3.54 | 18.06 | Yes |
| 6 | 1 | 0 | 1 | 0 | 91 | 33 | 36.26 | 1266 | 47.24 | -10.97 | 4.94 | -20.11 | -0.72 | Yes |
| 6 | 2 | 0 | 1 | 1 | 169 | 94 | 55.62 | 1266 | 47.24 | 8.39 | 3.78 | 0.85 | 15.67 | Yes |
| 6 | 3 | 0 | 2 | 0 | 68 | 25 | 36.76 | 1266 | 47.24 | -10.47 | 5.70 | -20.94 | 1.41 | No |
| 6 | 4 | 0 | 2 | 1 | 183 | 96 | 52.46 | 1266 | 47.24 | 5.22 | 3.65 | -1.99 | 12.33 | No |
| 6 | 5 | 0 | 3 | 0 | 82 | 34 | 41.46 | 1266 | 47.24 | -5.77 | 5.32 | -15.82 | 5.04 | No |
| 6 | 6 | 0 | 3 | 1 | 144 | 72 | 50.00 | 1266 | 47.24 | 2.76 | 4.11 | -5.30 | 10.82 | No |
| 6 | 7 | 1 | 1 | 0 | 66 | 12 | 18.18 | 1266 | 47.24 | -29.05 | 4.70 | -36.52 | -18.09 | Yes |

| **Area** | **SD** | **Sex** | **Age** | **Income** | **n_ASD** | **Events** | **p_ASD (%)** | **n_area** | **p_area (%)** | **Dev_A,SD (pp)** | **SE (pp)** | **95% CI low (pp)** | **95% CI high (pp)** | **Significant** |
| --- | --- | --- | --- | --- | --- | --- | --- | --- | --- | --- | --- | --- | --- | --- |
| 6 | 8 | 1 | 1 | 1 | 120 | 65 | 54.17 | 1266 | 47.24 | 6.93 | 4.48 | -1.97 | 15.58 | No |
| 6 | 9 | 1 | 2 | 0 | 35 | 9 | 25.71 | 1266 | 47.24 | -21.52 | 7.12 | -33.07 | -5.17 | Yes |
| 6 | 10 | 1 | 2 | 1 | 126 | 69 | 54.76 | 1266 | 47.24 | 7.53 | 4.37 | -1.18 | 15.95 | No |
| 6 | 11 | 1 | 3 | 0 | 44 | 21 | 47.73 | 1266 | 47.24 | 0.49 | 7.22 | -13.48 | 14.83 | No |
| 6 | 12 | 1 | 3 | 1 | 138 | 68 | 49.28 | 1266 | 47.24 | 2.04 | 4.20 | -6.17 | 10.29 | No |
| 7 | 1 | 0 | 1 | 0 | 117 | 30 | 25.64 | 1340 | 21.72 | 3.92 | 3.99 | -3.12 | 12.52 | No |
| 7 | 2 | 0 | 1 | 1 | 169 | 30 | 17.75 | 1340 | 21.72 | -3.96 | 2.93 | -8.99 | 2.49 | No |
| 7 | 3 | 0 | 2 | 0 | 89 | 20 | 22.47 | 1340 | 21.72 | 0.76 | 4.37 | -6.67 | 10.46 | No |
| 7 | 4 | 0 | 2 | 1 | 153 | 26 | 16.99 | 1340 | 21.72 | -4.72 | 3.03 | -9.85 | 2.02 | No |
| 7 | 5 | 0 | 3 | 0 | 100 | 19 | 19.00 | 1340 | 21.72 | -2.72 | 3.89 | -9.20 | 6.06 | No |
| 7 | 6 | 0 | 3 | 1 | 115 | 42 | 36.52 | 1340 | 21.72 | 14.81 | 4.42 | 6.57 | 23.91 | Yes |
| 7 | 7 | 1 | 1 | 0 | 80 | 10 | 12.50 | 1340 | 21.72 | -9.22 | 3.72 | -14.78 | -0.21 | Yes |
| 7 | 8 | 1 | 1 | 1 | 128 | 27 | 21.09 | 1340 | 21.72 | -0.62 | 3.58 | -6.80 | 7.23 | No |
| 7 | 9 | 1 | 2 | 0 | 82 | 19 | 23.17 | 1340 | 21.72 | 1.45 | 4.59 | -6.35 | 11.66 | No |
| 7 | 10 | 1 | 2 | 1 | 134 | 25 | 18.66 | 1340 | 21.72 | -3.06 | 3.35 | -8.75 | 4.38 | No |
| 7 | 11 | 1 | 3 | 0 | 73 | 15 | 20.55 | 1340 | 21.72 | -1.17 | 4.67 | -8.85 | 9.46 | No |
| 7 | 12 | 1 | 3 | 1 | 100 | 28 | 28.00 | 1340 | 21.72 | 6.28 | 4.43 | -1.58 | 15.77 | No |
| 8 | 1 | 0 | 1 | 0 | 127 | 28 | 22.05 | 1452 | 23.14 | -1.09 | 3.65 | -7.42 | 6.88 | No |
| 8 | 2 | 0 | 1 | 1 | 142 | 37 | 26.06 | 1452 | 23.14 | 2.92 | 3.65 | -3.61 | 10.70 | No |
| 8 | 3 | 0 | 2 | 0 | 117 | 21 | 17.95 | 1452 | 23.14 | -5.19 | 3.53 | -11.09 | 2.74 | No |
| 8 | 4 | 0 | 2 | 1 | 150 | 39 | 26.00 | 1452 | 23.14 | 2.86 | 3.55 | -3.50 | 10.42 | No |
| 8 | 5 | 0 | 3 | 0 | 138 | 23 | 16.67 | 1452 | 23.14 | -6.47 | 3.16 | -11.77 | 0.63 | No |
| 8 | 6 | 0 | 3 | 1 | 115 | 35 | 30.43 | 1452 | 23.14 | 7.29 | 4.23 | -0.37 | 16.22 | No |
| 8 | 7 | 1 | 1 | 0 | 151 | 19 | 12.58 | 1452 | 23.14 | -10.56 | 2.71 | -14.94 | -4.32 | Yes |
| 8 | 8 | 1 | 1 | 1 | 114 | 26 | 22.81 | 1452 | 23.14 | -0.33 | 3.89 | -7.07 | 8.18 | No |
| 8 | 9 | 1 | 2 | 0 | 106 | 25 | 23.58 | 1452 | 23.14 | 0.44 | 4.08 | -6.62 | 9.36 | No |
| 8 | 10 | 1 | 2 | 1 | 98 | 29 | 29.59 | 1452 | 23.14 | 6.45 | 4.54 | -1.68 | 16.12 | No |
| 8 | 11 | 1 | 3 | 0 | 89 | 20 | 22.47 | 1452 | 23.14 | -0.67 | 4.37 | -8.10 | 9.04 | No |
| 8 | 12 | 1 | 3 | 1 | 105 | 34 | 32.38 | 1452 | 23.14 | 9.24 | 4.50 | 1.05 | 18.68 | Yes |
| 9 | 1 | 0 | 1 | 0 | 198 | 16 | 8.08 | 1957 | 11.04 | -2.96 | 1.96 | -6.00 | 1.69 | No |
| 9 | 2 | 0 | 1 | 1 | 216 | 22 | 10.19 | 1957 | 11.04 | -0.85 | 2.07 | -4.21 | 3.90 | No |
| 9 | 3 | 0 | 2 | 0 | 194 | 18 | 9.28 | 1957 | 11.04 | -1.76 | 2.10 | -5.09 | 3.15 | No |
| 9 | 4 | 0 | 2 | 1 | 163 | 21 | 12.88 | 1957 | 11.04 | 1.85 | 2.63 | -2.45 | 7.86 | No |
| 9 | 5 | 0 | 3 | 0 | 206 | 23 | 11.17 | 1957 | 11.04 | 0.13 | 2.20 | -3.48 | 5.16 | No |
| 9 | 6 | 0 | 3 | 1 | 145 | 23 | 15.86 | 1957 | 11.04 | 4.82 | 3.03 | -0.23 | 11.64 | No |

| **Area** | **SD** | **Sex** | **Age** | **Income** | **n_ASD** | **Events** | **p_ASD (%)** | **n_area** | **p_area (%)** | **Dev_A,SD (pp)** | **SE (pp)** | **95% CI low (pp)** | **95% CI high (pp)** | **Significant** |
| --- | --- | --- | --- | --- | --- | --- | --- | --- | --- | --- | --- | --- | --- | --- |
| 9 | 7 | 1 | 1 | 0 | 178 | 12 | 6.74 | 1957 | 11.04 | -4.30 | 1.92 | -7.14 | 0.38 | No |
| 9 | 8 | 1 | 1 | 1 | 161 | 23 | 14.29 | 1957 | 11.04 | 3.25 | 2.76 | -1.33 | 9.49 | No |
| 9 | 9 | 1 | 2 | 0 | 140 | 8 | 5.71 | 1957 | 11.04 | -5.32 | 2.03 | -8.11 | -0.17 | Yes |
| 9 | 10 | 1 | 2 | 1 | 116 | 19 | 16.38 | 1957 | 11.04 | 5.34 | 3.43 | -0.29 | 13.13 | No |
| 9 | 11 | 1 | 3 | 0 | 117 | 9 | 7.69 | 1957 | 11.04 | -3.34 | 2.52 | -6.94 | 2.94 | No |
| 9 | 12 | 1 | 3 | 1 | 123 | 22 | 17.89 | 1957 | 11.04 | 6.85 | 3.44 | 1.08 | 14.56 | Yes |
| 10 | 1 | 0 | 1 | 0 | 182 | 17 | 9.34 | 2166 | 12.93 | -3.59 | 2.18 | -7.01 | 1.52 | No |
| 10 | 2 | 0 | 1 | 1 | 235 | 32 | 13.62 | 2166 | 12.93 | 0.69 | 2.24 | -3.11 | 5.66 | No |
| 10 | 3 | 0 | 2 | 0 | 197 | 21 | 10.66 | 2166 | 12.93 | -2.27 | 2.21 | -5.85 | 2.82 | No |
| 10 | 4 | 0 | 2 | 1 | 207 | 30 | 14.49 | 2166 | 12.93 | 1.57 | 2.45 | -2.58 | 7.01 | No |
| 10 | 5 | 0 | 3 | 0 | 201 | 26 | 12.94 | 2166 | 12.93 | 0.01 | 2.37 | -3.94 | 5.35 | No |
| 10 | 6 | 0 | 3 | 1 | 179 | 40 | 22.35 | 2166 | 12.93 | 9.42 | 3.09 | 3.93 | 16.07 | Yes |
| 10 | 7 | 1 | 1 | 0 | 164 | 10 | 6.10 | 2166 | 12.93 | -6.83 | 1.92 | -9.58 | -2.07 | Yes |
| 10 | 8 | 1 | 1 | 1 | 168 | 30 | 17.86 | 2166 | 12.93 | 4.93 | 2.94 | -0.12 | 11.42 | No |
| 10 | 9 | 1 | 2 | 0 | 134 | 9 | 6.72 | 2166 | 12.93 | -6.21 | 2.22 | -9.35 | -0.66 | Yes |
| 10 | 10 | 1 | 2 | 1 | 172 | 19 | 11.05 | 2166 | 12.93 | -1.88 | 2.40 | -5.74 | 3.68 | No |
| 10 | 11 | 1 | 3 | 0 | 147 | 9 | 6.12 | 2166 | 12.93 | -6.80 | 2.03 | -9.67 | -1.70 | Yes |
| 10 | 12 | 1 | 3 | 1 | 180 | 37 | 20.56 | 2166 | 12.93 | 7.63 | 3.00 | 2.37 | 14.12 | Yes |
| 11 | 1 | 0 | 1 | 0 | 108 | 33 | 30.56 | 1521 | 42.87 | -12.31 | 4.37 | -20.21 | -3.08 | Yes |
| 11 | 2 | 0 | 1 | 1 | 217 | 106 | 48.85 | 1521 | 42.87 | 5.98 | 3.36 | -0.59 | 12.59 | No |
| 11 | 3 | 0 | 2 | 0 | 86 | 31 | 36.05 | 1521 | 42.87 | -6.82 | 5.07 | -16.17 | 3.72 | No |
| 11 | 4 | 0 | 2 | 1 | 220 | 85 | 38.64 | 1521 | 42.87 | -4.23 | 3.26 | -10.42 | 2.35 | No |
| 11 | 5 | 0 | 3 | 0 | 107 | 40 | 37.38 | 1521 | 42.87 | -5.48 | 4.60 | -14.06 | 3.97 | No |
| 11 | 6 | 0 | 3 | 1 | 162 | 90 | 55.56 | 1521 | 42.87 | 12.69 | 3.86 | 5.00 | 20.12 | Yes |
| 11 | 7 | 1 | 1 | 0 | 75 | 19 | 25.33 | 1521 | 42.87 | -17.53 | 4.94 | -26.01 | -6.66 | Yes |
| 11 | 8 | 1 | 1 | 1 | 152 | 84 | 55.26 | 1521 | 42.87 | 12.40 | 3.98 | 4.46 | 20.07 | Yes |
| 11 | 9 | 1 | 2 | 0 | 49 | 13 | 26.53 | 1521 | 42.87 | -16.34 | 6.14 | -26.66 | -2.60 | Yes |
| 11 | 10 | 1 | 2 | 1 | 140 | 55 | 39.29 | 1521 | 42.87 | -3.58 | 4.07 | -11.28 | 4.69 | No |
| 11 | 11 | 1 | 3 | 0 | 67 | 24 | 35.82 | 1521 | 42.87 | -7.05 | 5.71 | -17.47 | 4.92 | No |
| 11 | 12 | 1 | 3 | 1 | 138 | 72 | 52.17 | 1521 | 42.87 | 9.31 | 4.19 | 1.03 | 17.47 | Yes |
| 12 | 1 | 0 | 1 | 0 | 213 | 22 | 10.33 | 2161 | 11.99 | -1.66 | 2.10 | -5.06 | 3.16 | No |
| 12 | 2 | 0 | 1 | 1 | 202 | 24 | 11.88 | 2161 | 11.99 | -0.10 | 2.28 | -3.87 | 5.08 | No |
| 12 | 3 | 0 | 2 | 0 | 173 | 31 | 17.92 | 2161 | 11.99 | 5.93 | 2.91 | 0.94 | 12.33 | Yes |
| 12 | 4 | 0 | 2 | 1 | 230 | 35 | 15.22 | 2161 | 11.99 | 3.23 | 2.37 | -0.84 | 8.44 | No |
| 12 | 5 | 0 | 3 | 0 | 216 | 18 | 8.33 | 2161 | 11.99 | -3.65 | 1.90 | -6.65 | 0.80 | No |

| **Area** | **SD** | **Sex** | **Age** | **Income** | **n_ASD** | **Events** | **p_ASD (%)** | **n_area** | **p_area (%)** | **Dev_A,SD (pp)** | **SE (pp)** | **95% CI low (pp)** | **95% CI high (pp)** | **Significant** |
| --- | --- | --- | --- | --- | --- | --- | --- | --- | --- | --- | --- | --- | --- | --- |
| 12 | 6 | 0 | 3 | 1 | 197 | 29 | 14.72 | 2161 | 11.99 | 2.74 | 2.52 | -1.54 | 8.36 | No |
| 12 | 7 | 1 | 1 | 0 | 160 | 14 | 8.75 | 2161 | 11.99 | -3.24 | 2.26 | -6.70 | 2.17 | No |
| 12 | 8 | 1 | 1 | 1 | 171 | 13 | 7.60 | 2161 | 11.99 | -4.38 | 2.06 | -7.49 | 0.59 | No |
| 12 | 9 | 1 | 2 | 0 | 114 | 9 | 7.89 | 2161 | 11.99 | -4.09 | 2.58 | -7.78 | 2.34 | No |
| 12 | 10 | 1 | 2 | 1 | 155 | 25 | 16.13 | 2161 | 11.99 | 4.14 | 2.95 | -0.82 | 10.74 | No |
| 12 | 11 | 1 | 3 | 0 | 151 | 12 | 7.95 | 2161 | 11.99 | -4.04 | 2.24 | -7.38 | 1.39 | No |
| 12 | 12 | 1 | 3 | 1 | 179 | 27 | 15.08 | 2161 | 11.99 | 3.10 | 2.67 | -1.41 | 9.07 | No |
| 13 | 1 | 0 | 1 | 0 | 111 | 30 | 27.03 | 1682 | 28.54 | -1.51 | 4.16 | -8.90 | 7.42 | No |
| 13 | 2 | 0 | 1 | 1 | 195 | 54 | 27.69 | 1682 | 28.54 | -0.85 | 3.18 | -6.65 | 5.82 | No |
| 13 | 3 | 0 | 2 | 0 | 116 | 30 | 25.86 | 1682 | 28.54 | -2.68 | 4.02 | -9.78 | 5.98 | No |
| 13 | 4 | 0 | 2 | 1 | 187 | 56 | 29.95 | 1682 | 28.54 | 1.41 | 3.32 | -4.70 | 8.32 | No |
| 13 | 5 | 0 | 3 | 0 | 156 | 37 | 23.72 | 1682 | 28.54 | -4.82 | 3.38 | -10.81 | 2.44 | No |
| 13 | 6 | 0 | 3 | 1 | 206 | 48 | 23.30 | 1682 | 28.54 | -5.24 | 2.93 | -10.49 | 0.99 | No |
| 13 | 7 | 1 | 1 | 0 | 96 | 31 | 32.29 | 1682 | 28.54 | 3.75 | 4.69 | -4.76 | 13.63 | No |
| 13 | 8 | 1 | 1 | 1 | 142 | 49 | 34.51 | 1682 | 28.54 | 5.97 | 3.94 | -1.35 | 14.10 | No |
| 13 | 9 | 1 | 2 | 0 | 64 | 14 | 21.88 | 1682 | 28.54 | -6.66 | 5.08 | -15.04 | 4.90 | No |
| 13 | 10 | 1 | 2 | 1 | 146 | 51 | 34.93 | 1682 | 28.54 | 6.39 | 3.90 | -0.86 | 14.42 | No |
| 13 | 11 | 1 | 3 | 0 | 78 | 19 | 24.36 | 1682 | 28.54 | -4.18 | 4.78 | -12.35 | 6.40 | No |
| 13 | 12 | 1 | 3 | 1 | 185 | 61 | 32.97 | 1682 | 28.54 | 4.44 | 3.43 | -1.93 | 11.50 | No |
| 14 | 1 | 0 | 1 | 0 | 200 | 23 | 11.50 | 2252 | 18.47 | -6.97 | 2.26 | -10.69 | -1.81 | Yes |
| 14 | 2 | 0 | 1 | 1 | 299 | 65 | 21.74 | 2252 | 18.47 | 3.27 | 2.38 | -1.03 | 8.28 | No |
| 14 | 3 | 0 | 2 | 0 | 153 | 15 | 9.80 | 2252 | 18.47 | -8.67 | 2.43 | -12.44 | -2.93 | Yes |
| 14 | 4 | 0 | 2 | 1 | 229 | 60 | 26.20 | 2252 | 18.47 | 7.73 | 2.89 | 2.46 | 13.78 | Yes |
| 14 | 5 | 0 | 3 | 0 | 177 | 18 | 10.17 | 2252 | 18.47 | -8.30 | 2.29 | -11.94 | -2.97 | Yes |
| 14 | 6 | 0 | 3 | 1 | 173 | 50 | 28.90 | 2252 | 18.47 | 10.43 | 3.42 | 4.19 | 17.58 | Yes |
| 14 | 7 | 1 | 1 | 0 | 204 | 17 | 8.33 | 2252 | 18.47 | -10.14 | 1.96 | -13.20 | -5.53 | Yes |
| 14 | 8 | 1 | 1 | 1 | 167 | 51 | 30.54 | 2252 | 18.47 | 12.07 | 3.53 | 5.58 | 19.42 | Yes |
| 14 | 9 | 1 | 2 | 0 | 139 | 13 | 9.35 | 2252 | 18.47 | -9.12 | 2.50 | -12.93 | -3.13 | Yes |
| 14 | 10 | 1 | 2 | 1 | 219 | 42 | 19.18 | 2252 | 18.47 | 0.71 | 2.65 | -3.96 | 6.43 | No |
| 14 | 11 | 1 | 3 | 0 | 133 | 20 | 15.04 | 2252 | 18.47 | -3.43 | 3.10 | -8.52 | 3.62 | No |
| 14 | 12 | 1 | 3 | 1 | 159 | 42 | 26.42 | 2252 | 18.47 | 7.94 | 3.47 | 1.70 | 15.29 | Yes |
| 15 | 1 | 0 | 1 | 0 | 153 | 13 | 8.50 | 1652 | 22.46 | -13.96 | 2.29 | -17.43 | -8.46 | Yes |
| 15 | 2 | 0 | 1 | 1 | 171 | 37 | 21.64 | 1652 | 22.46 | -0.82 | 3.13 | -6.33 | 5.94 | No |
| 15 | 3 | 0 | 2 | 0 | 102 | 15 | 14.71 | 1652 | 22.46 | -7.75 | 3.50 | -13.34 | 0.40 | No |
| 15 | 4 | 0 | 2 | 1 | 162 | 41 | 25.31 | 1652 | 22.46 | 2.85 | 3.39 | -3.22 | 10.06 | No |

| **Area** | **SD** | **Sex** | **Age** | **Income** | **n_ASD** | **Events** | **p_ASD (%)** | **n_area** | **p_area (%)** | **Dev_A,SD (pp)** | **SE (pp)** | **95% CI low (pp)** | **95% CI high (pp)** | **Significant** |
| --- | --- | --- | --- | --- | --- | --- | --- | --- | --- | --- | --- | --- | --- | --- |
| 15 | 5 | 0 | 3 | 0 | 163 | 33 | 20.25 | 1652 | 22.46 | -2.21 | 3.13 | -7.66 | 4.61 | No |
| 15 | 6 | 0 | 3 | 1 | 194 | 71 | 36.60 | 1652 | 22.46 | 14.14 | 3.43 | 7.68 | 21.12 | Yes |
| 15 | 7 | 1 | 1 | 0 | 115 | 12 | 10.43 | 1652 | 22.46 | -12.02 | 2.88 | -16.39 | -5.10 | Yes |
| 15 | 8 | 1 | 1 | 1 | 135 | 42 | 31.11 | 1652 | 22.46 | 8.65 | 3.94 | 1.46 | 16.89 | Yes |
| 15 | 9 | 1 | 2 | 0 | 85 | 8 | 9.41 | 1652 | 22.46 | -13.05 | 3.22 | -17.61 | -4.97 | Yes |
| 15 | 10 | 1 | 2 | 1 | 120 | 33 | 27.50 | 1652 | 22.46 | 5.04 | 4.03 | -2.15 | 13.64 | No |
| 15 | 11 | 1 | 3 | 0 | 97 | 9 | 9.28 | 1652 | 22.46 | -13.18 | 3.00 | -17.50 | -5.76 | Yes |
| 15 | 12 | 1 | 3 | 1 | 155 | 57 | 36.77 | 1652 | 22.46 | 14.32 | 3.83 | 7.13 | 22.14 | Yes |
| 16 | 1 | 0 | 1 | 0 | 163 | 10 | 6.13 | 1989 | 11.21 | -5.08 | 1.93 | -7.85 | -0.29 | Yes |
| 16 | 2 | 0 | 1 | 1 | 202 | 21 | 10.40 | 1989 | 11.21 | -0.82 | 2.16 | -4.31 | 4.16 | No |
| 16 | 3 | 0 | 2 | 0 | 132 | 8 | 6.06 | 1989 | 11.21 | -5.15 | 2.14 | -8.11 | 0.29 | No |
| 16 | 4 | 0 | 2 | 1 | 189 | 35 | 18.52 | 1989 | 11.21 | 7.31 | 2.82 | 2.42 | 13.45 | Yes |
| 16 | 5 | 0 | 3 | 0 | 226 | 15 | 6.64 | 1989 | 11.21 | -4.57 | 1.68 | -7.15 | -0.55 | Yes |
| 16 | 6 | 0 | 3 | 1 | 218 | 39 | 17.89 | 1989 | 11.21 | 6.68 | 2.59 | 2.16 | 12.31 | Yes |
| 16 | 7 | 1 | 1 | 0 | 162 | 10 | 6.17 | 1989 | 11.21 | -5.04 | 1.94 | -7.82 | -0.22 | Yes |
| 16 | 8 | 1 | 1 | 1 | 115 | 15 | 13.04 | 1989 | 11.21 | 1.83 | 3.15 | -3.15 | 9.20 | No |
| 16 | 9 | 1 | 2 | 0 | 127 | 9 | 7.09 | 1989 | 11.21 | -4.13 | 2.33 | -7.44 | 1.71 | No |
| 16 | 10 | 1 | 2 | 1 | 156 | 27 | 17.31 | 1989 | 11.21 | 6.10 | 3.02 | 0.96 | 12.80 | Yes |
| 16 | 11 | 1 | 3 | 0 | 117 | 8 | 6.84 | 1989 | 11.21 | -4.37 | 2.40 | -7.71 | 1.70 | No |
| 16 | 12 | 1 | 3 | 1 | 182 | 26 | 14.29 | 1989 | 11.21 | 3.07 | 2.59 | -1.27 | 8.90 | No |
| 17 | 1 | 0 | 1 | 0 | 236 | 53 | 22.46 | 2348 | 22.91 | -0.46 | 2.70 | -5.31 | 5.29 | No |
| 17 | 2 | 0 | 1 | 1 | 240 | 77 | 32.08 | 2348 | 22.91 | 9.17 | 2.99 | 3.59 | 15.32 | Yes |
| 17 | 3 | 0 | 2 | 0 | 210 | 31 | 14.76 | 2348 | 22.91 | -8.15 | 2.45 | -12.31 | -2.72 | Yes |
| 17 | 4 | 0 | 2 | 1 | 245 | 65 | 26.53 | 2348 | 22.91 | 3.62 | 2.80 | -1.52 | 9.48 | No |
| 17 | 5 | 0 | 3 | 0 | 210 | 43 | 20.48 | 2348 | 22.91 | -2.44 | 2.77 | -7.34 | 3.53 | No |
| 17 | 6 | 0 | 3 | 1 | 167 | 42 | 25.15 | 2348 | 22.91 | 2.24 | 3.33 | -3.73 | 9.33 | No |
| 17 | 7 | 1 | 1 | 0 | 203 | 30 | 14.78 | 2348 | 22.91 | -8.13 | 2.49 | -12.36 | -2.60 | Yes |
| 17 | 8 | 1 | 1 | 1 | 182 | 58 | 31.87 | 2348 | 22.91 | 8.96 | 3.42 | 2.62 | 16.04 | Yes |
| 17 | 9 | 1 | 2 | 0 | 180 | 29 | 16.11 | 2348 | 22.91 | -6.80 | 2.74 | -11.46 | -0.73 | Yes |
| 17 | 10 | 1 | 2 | 1 | 176 | 50 | 28.41 | 2348 | 22.91 | 5.50 | 3.37 | -0.65 | 12.56 | No |
| 17 | 11 | 1 | 3 | 0 | 163 | 27 | 16.56 | 2348 | 22.91 | -6.35 | 2.90 | -11.27 | 0.11 | No |
| 17 | 12 | 1 | 3 | 1 | 136 | 33 | 24.26 | 2348 | 22.91 | 1.35 | 3.64 | -5.08 | 9.20 | No |
| 18 | 1 | 0 | 1 | 0 | 128 | 11 | 8.59 | 1726 | 22.94 | -14.35 | 2.52 | -18.08 | -8.21 | Yes |
| 18 | 2 | 0 | 1 | 1 | 205 | 53 | 25.85 | 1726 | 22.94 | 2.91 | 3.04 | -2.60 | 9.31 | No |
| 18 | 3 | 0 | 2 | 0 | 113 | 15 | 13.27 | 1726 | 22.94 | -9.67 | 3.20 | -14.73 | -2.19 | Yes |

| **Area** | **SD** | **Sex** | **Age** | **Income** | **n_ASD** | **Events** | **p_ASD (%)** | **n_area** | **p_area (%)** | **Dev_A,SD (pp)** | **SE (pp)** | **95% CI low (pp)** | **95% CI high (pp)** | **Significant** |
| --- | --- | --- | --- | --- | --- | --- | --- | --- | --- | --- | --- | --- | --- | --- |
| 18 | 4 | 0 | 2 | 1 | 212 | 53 | 25.00 | 1726 | 22.94 | 2.06 | 2.96 | -3.29 | 8.30 | No |
| 18 | 5 | 0 | 3 | 0 | 128 | 9 | 7.03 | 1726 | 22.94 | -15.91 | 2.32 | -19.20 | -10.12 | Yes |
| 18 | 6 | 0 | 3 | 1 | 207 | 74 | 35.75 | 1726 | 22.94 | 12.81 | 3.30 | 6.59 | 19.54 | Yes |
| 18 | 7 | 1 | 1 | 0 | 110 | 11 | 10.00 | 1726 | 22.94 | -12.94 | 2.89 | -17.27 | -5.92 | Yes |
| 18 | 8 | 1 | 1 | 1 | 118 | 34 | 28.81 | 1726 | 22.94 | 5.87 | 4.12 | -1.53 | 14.61 | No |
| 18 | 9 | 1 | 2 | 0 | 85 | 12 | 14.12 | 1726 | 22.94 | -8.83 | 3.78 | -14.68 | 0.13 | No |
| 18 | 10 | 1 | 2 | 1 | 144 | 46 | 31.94 | 1726 | 22.94 | 9.00 | 3.84 | 1.94 | 17.00 | Yes |
| 18 | 11 | 1 | 3 | 0 | 86 | 9 | 10.47 | 1726 | 22.94 | -12.48 | 3.34 | -17.34 | -4.24 | Yes |
| 18 | 12 | 1 | 3 | 1 | 190 | 69 | 36.32 | 1726 | 22.94 | 13.37 | 3.46 | 6.87 | 20.42 | Yes |
| 19 | 1 | 0 | 1 | 0 | 86 | 21 | 24.42 | 1217 | 41.33 | -16.91 | 4.57 | -24.77 | -6.87 | Yes |
| 19 | 2 | 0 | 1 | 1 | 157 | 84 | 53.50 | 1217 | 41.33 | 12.17 | 3.93 | 4.38 | 19.80 | Yes |
| 19 | 3 | 0 | 2 | 0 | 70 | 14 | 20.00 | 1217 | 41.33 | -21.33 | 4.72 | -29.03 | -10.51 | Yes |
| 19 | 4 | 0 | 2 | 1 | 117 | 64 | 54.70 | 1217 | 41.33 | 13.37 | 4.53 | 4.34 | 22.10 | Yes |
| 19 | 5 | 0 | 3 | 0 | 103 | 22 | 21.36 | 1217 | 41.33 | -19.97 | 4.00 | -26.78 | -11.10 | Yes |
| 19 | 6 | 0 | 3 | 1 | 160 | 77 | 48.12 | 1217 | 41.33 | 6.79 | 3.90 | -0.81 | 14.49 | No |
| 19 | 7 | 1 | 1 | 0 | 70 | 18 | 25.71 | 1217 | 41.33 | -15.62 | 5.13 | -24.40 | -4.30 | Yes |
| 19 | 8 | 1 | 1 | 1 | 117 | 57 | 48.72 | 1217 | 41.33 | 7.39 | 4.55 | -1.48 | 16.34 | No |
| 19 | 9 | 1 | 2 | 0 | 71 | 16 | 22.54 | 1217 | 41.33 | -18.80 | 4.88 | -26.96 | -7.82 | Yes |
| 19 | 10 | 1 | 2 | 1 | 83 | 46 | 55.42 | 1217 | 41.33 | 14.09 | 5.34 | 3.39 | 24.31 | Yes |
| 19 | 11 | 1 | 3 | 0 | 70 | 20 | 28.57 | 1217 | 41.33 | -12.76 | 5.29 | -22.01 | -1.28 | Yes |
| 19 | 12 | 1 | 3 | 1 | 113 | 64 | 56.64 | 1217 | 41.33 | 15.31 | 4.59 | 6.10 | 24.08 | Yes |
| 20 | 1 | 0 | 1 | 0 | 178 | 12 | 6.74 | 1796 | 14.76 | -8.01 | 1.92 | -10.86 | -3.34 | Yes |
| 20 | 2 | 0 | 1 | 1 | 202 | 58 | 28.71 | 1796 | 14.76 | 13.96 | 3.16 | 8.16 | 20.55 | Yes |
| 20 | 3 | 0 | 2 | 0 | 152 | 17 | 11.18 | 1796 | 14.76 | -3.57 | 2.57 | -7.65 | 2.43 | No |
| 20 | 4 | 0 | 2 | 1 | 159 | 26 | 16.35 | 1796 | 14.76 | 1.60 | 2.93 | -3.34 | 8.13 | No |
| 20 | 5 | 0 | 3 | 0 | 144 | 13 | 9.03 | 1796 | 14.76 | -5.73 | 2.42 | -9.40 | 0.08 | No |
| 20 | 6 | 0 | 3 | 1 | 141 | 20 | 14.18 | 1796 | 14.76 | -0.57 | 2.94 | -5.38 | 6.14 | No |
| 20 | 7 | 1 | 1 | 0 | 148 | 10 | 6.76 | 1796 | 14.76 | -8.00 | 2.11 | -11.04 | -2.76 | Yes |
| 20 | 8 | 1 | 1 | 1 | 168 | 44 | 26.19 | 1796 | 14.76 | 11.44 | 3.36 | 5.37 | 18.56 | Yes |
| 20 | 9 | 1 | 2 | 0 | 147 | 9 | 6.12 | 1796 | 14.76 | -8.63 | 2.03 | -11.50 | -3.53 | Yes |
| 20 | 10 | 1 | 2 | 1 | 130 | 24 | 18.46 | 1796 | 14.76 | 3.71 | 3.39 | -2.02 | 11.25 | No |
| 20 | 11 | 1 | 3 | 0 | 114 | 12 | 10.53 | 1796 | 14.76 | -4.23 | 2.90 | -8.63 | 2.75 | No |
| 20 | 12 | 1 | 3 | 1 | 113 | 20 | 17.70 | 1796 | 14.76 | 2.94 | 3.57 | -3.00 | 11.01 | No |
| 21 | 1 | 0 | 1 | 0 | 193 | 13 | 6.74 | 1842 | 8.25 | -1.52 | 1.84 | -4.27 | 2.93 | No |
| 21 | 2 | 0 | 1 | 1 | 158 | 19 | 12.03 | 1842 | 8.25 | 3.77 | 2.60 | -0.42 | 9.77 | No |

| **Area** | **SD** | **Sex** | **Age** | **Income** | **n_ASD** | **Events** | **p_ASD (%)** | **n_area** | **p_area (%)** | **Dev_A,SD (pp)** | **SE (pp)** | **95% CI low (pp)** | **95% CI high (pp)** | **Significant** |
| --- | --- | --- | --- | --- | --- | --- | --- | --- | --- | --- | --- | --- | --- | --- |
| 21 | 3 | 0 | 2 | 0 | 177 | 15 | 8.47 | 1842 | 8.25 | 0.22 | 2.12 | -3.05 | 5.26 | No |
| 21 | 4 | 0 | 2 | 1 | 163 | 11 | 6.75 | 1842 | 8.25 | -1.50 | 2.01 | -4.44 | 3.43 | No |
| 21 | 5 | 0 | 3 | 0 | 180 | 14 | 7.78 | 1842 | 8.25 | -0.47 | 2.03 | -3.56 | 4.38 | No |
| 21 | 6 | 0 | 3 | 1 | 108 | 19 | 17.59 | 1842 | 8.25 | 9.34 | 3.64 | 3.31 | 17.60 | Yes |
| 21 | 7 | 1 | 1 | 0 | 199 | 7 | 3.52 | 1842 | 8.25 | -4.73 | 1.37 | -6.54 | -1.17 | Yes |
| 21 | 8 | 1 | 1 | 1 | 134 | 14 | 10.45 | 1842 | 8.25 | 2.20 | 2.67 | -1.93 | 8.52 | No |
| 21 | 9 | 1 | 2 | 0 | 148 | 8 | 5.41 | 1842 | 8.25 | -2.85 | 1.92 | -5.49 | 2.05 | No |
| 21 | 10 | 1 | 2 | 1 | 120 | 10 | 8.33 | 1842 | 8.25 | 0.08 | 2.57 | -3.66 | 6.41 | No |
| 21 | 11 | 1 | 3 | 0 | 156 | 12 | 7.69 | 1842 | 8.25 | -0.56 | 2.17 | -3.80 | 4.71 | No |
| 21 | 12 | 1 | 3 | 1 | 106 | 10 | 9.43 | 1842 | 8.25 | 1.18 | 2.88 | -3.05 | 8.25 | No |
| 22 | 1 | 0 | 1 | 0 | 111 | 13 | 11.71 | 1293 | 17.63 | -5.92 | 3.07 | -10.66 | 1.38 | No |
| 22 | 2 | 0 | 1 | 1 | 172 | 42 | 24.42 | 1293 | 17.63 | 6.79 | 3.25 | 0.97 | 13.72 | Yes |
| 22 | 3 | 0 | 2 | 0 | 66 | 7 | 10.61 | 1293 | 17.63 | -7.03 | 3.85 | -12.40 | 2.68 | No |
| 22 | 4 | 0 | 2 | 1 | 162 | 40 | 24.69 | 1293 | 17.63 | 7.06 | 3.36 | 1.06 | 14.23 | Yes |
| 22 | 5 | 0 | 3 | 0 | 96 | 8 | 8.33 | 1293 | 17.63 | -9.30 | 2.88 | -13.35 | -2.04 | Yes |
| 22 | 6 | 0 | 3 | 1 | 107 | 20 | 18.69 | 1293 | 17.63 | 1.06 | 3.74 | -5.20 | 9.48 | No |
| 22 | 7 | 1 | 1 | 0 | 98 | 5 | 5.10 | 1293 | 17.63 | -12.53 | 2.35 | -15.43 | -6.24 | Yes |
| 22 | 8 | 1 | 1 | 1 | 106 | 30 | 28.30 | 1293 | 17.63 | 10.67 | 4.32 | 2.97 | 19.89 | Yes |
| 22 | 9 | 1 | 2 | 0 | 70 | 6 | 8.57 | 1293 | 17.63 | -9.06 | 3.44 | -13.65 | -0.17 | Yes |
| 22 | 10 | 1 | 2 | 1 | 131 | 34 | 25.95 | 1293 | 17.63 | 8.32 | 3.79 | 1.57 | 16.44 | Yes |
| 22 | 11 | 1 | 3 | 0 | 71 | 4 | 5.63 | 1293 | 17.63 | -12.00 | 2.91 | -15.42 | -4.02 | Yes |
| 22 | 12 | 1 | 3 | 1 | 103 | 19 | 18.45 | 1293 | 17.63 | 0.81 | 3.80 | -5.49 | 9.39 | No |
| 23 | 1 | 0 | 1 | 0 | 183 | 25 | 13.66 | 1691 | 13.31 | 0.36 | 2.54 | -3.88 | 6.08 | No |
| 23 | 2 | 0 | 1 | 1 | 146 | 20 | 13.70 | 1691 | 13.31 | 0.39 | 2.85 | -4.26 | 6.91 | No |
| 23 | 3 | 0 | 2 | 0 | 170 | 28 | 16.47 | 1691 | 13.31 | 3.16 | 2.84 | -1.66 | 9.47 | No |
| 23 | 4 | 0 | 2 | 1 | 153 | 22 | 14.38 | 1691 | 13.31 | 1.07 | 2.84 | -3.61 | 7.51 | No |
| 23 | 5 | 0 | 3 | 0 | 175 | 22 | 12.57 | 1691 | 13.31 | -0.73 | 2.51 | -4.86 | 4.99 | No |
| 23 | 6 | 0 | 3 | 1 | 133 | 17 | 12.78 | 1691 | 13.31 | -0.52 | 2.90 | -5.17 | 6.21 | No |
| 23 | 7 | 1 | 1 | 0 | 158 | 13 | 8.23 | 1691 | 13.31 | -5.08 | 2.22 | -8.43 | 0.26 | No |
| 23 | 8 | 1 | 1 | 1 | 100 | 18 | 18.00 | 1691 | 13.31 | 4.69 | 3.82 | -1.61 | 13.36 | No |
| 23 | 9 | 1 | 2 | 0 | 138 | 16 | 11.59 | 1691 | 13.31 | -1.71 | 2.74 | -6.04 | 4.70 | No |
| 23 | 10 | 1 | 2 | 1 | 115 | 19 | 16.52 | 1691 | 13.31 | 3.22 | 3.45 | -2.47 | 11.06 | No |
| 23 | 11 | 1 | 3 | 0 | 112 | 12 | 10.71 | 1691 | 13.31 | -2.59 | 2.95 | -7.07 | 4.49 | No |
| 23 | 12 | 1 | 3 | 1 | 108 | 13 | 12.04 | 1691 | 13.31 | -1.27 | 3.15 | -6.13 | 6.21 | No |
| 24 | 1 | 0 | 1 | 0 | 95 | 24 | 25.26 | 1266 | 46.52 | -21.26 | 4.40 | -28.92 | -11.68 | Yes |

| **Area** | **SD** | **Sex** | **Age** | **Income** | **n_ASD** | **Events** | **p_ASD (%)** | **n_area** | **p_area (%)** | **Dev_A,SD (pp)** | **SE (pp)** | **95% CI low (pp)** | **95% CI high (pp)** | **Significant** |
| --- | --- | --- | --- | --- | --- | --- | --- | --- | --- | --- | --- | --- | --- | --- |
| 24 | 2 | 0 | 1 | 1 | 140 | 84 | 60.00 | 1266 | 46.52 | 13.48 | 4.09 | 5.20 | 21.22 | Yes |
| 24 | 3 | 0 | 2 | 0 | 75 | 16 | 21.33 | 1266 | 46.52 | -25.19 | 4.67 | -32.94 | -14.64 | Yes |
| 24 | 4 | 0 | 2 | 1 | 123 | 86 | 69.92 | 1266 | 46.52 | 23.39 | 4.08 | 14.79 | 30.79 | Yes |
| 24 | 5 | 0 | 3 | 0 | 94 | 33 | 35.11 | 1266 | 46.52 | -11.42 | 4.83 | -20.31 | -1.36 | Yes |
| 24 | 6 | 0 | 3 | 1 | 125 | 81 | 64.80 | 1266 | 46.52 | 18.28 | 4.21 | 9.58 | 26.09 | Yes |
| 24 | 7 | 1 | 1 | 0 | 113 | 18 | 15.93 | 1266 | 46.52 | -30.60 | 3.43 | -36.20 | -22.75 | Yes |
| 24 | 8 | 1 | 1 | 1 | 110 | 68 | 61.82 | 1266 | 46.52 | 15.29 | 4.56 | 5.96 | 23.83 | Yes |
| 24 | 9 | 1 | 2 | 0 | 105 | 16 | 15.24 | 1266 | 46.52 | -31.29 | 3.50 | -36.92 | -23.20 | Yes |
| 24 | 10 | 1 | 2 | 1 | 105 | 71 | 67.62 | 1266 | 46.52 | 21.09 | 4.50 | 11.66 | 29.29 | Yes |
| 24 | 11 | 1 | 3 | 0 | 84 | 24 | 28.57 | 1266 | 46.52 | -17.95 | 4.84 | -26.51 | -7.52 | Yes |
| 24 | 12 | 1 | 3 | 1 | 97 | 68 | 70.10 | 1266 | 46.52 | 23.58 | 4.58 | 13.84 | 31.78 | Yes |
| 25 | 1 | 0 | 1 | 0 | 111 | 26 | 23.42 | 1424 | 38.97 | -15.55 | 3.98 | -22.46 | -6.87 | Yes |
| 25 | 2 | 0 | 1 | 1 | 180 | 81 | 45.00 | 1424 | 38.97 | 6.03 | 3.67 | -1.06 | 13.32 | No |
| 25 | 3 | 0 | 2 | 0 | 77 | 32 | 41.56 | 1424 | 38.97 | 2.58 | 5.48 | -7.77 | 13.74 | No |
| 25 | 4 | 0 | 2 | 1 | 184 | 72 | 39.13 | 1424 | 38.97 | 0.16 | 3.56 | -6.60 | 7.36 | No |
| 25 | 5 | 0 | 3 | 0 | 108 | 31 | 28.70 | 1424 | 38.97 | -10.27 | 4.29 | -17.96 | -1.12 | Yes |
| 25 | 6 | 0 | 3 | 1 | 152 | 75 | 49.34 | 1424 | 38.97 | 10.37 | 4.00 | 2.53 | 18.23 | Yes |
| 25 | 7 | 1 | 1 | 0 | 78 | 16 | 20.51 | 1424 | 38.97 | -18.46 | 4.52 | -25.94 | -8.22 | Yes |
| 25 | 8 | 1 | 1 | 1 | 130 | 50 | 38.46 | 1424 | 38.97 | -0.51 | 4.21 | -8.43 | 8.07 | No |
| 25 | 9 | 1 | 2 | 0 | 56 | 18 | 32.14 | 1424 | 38.97 | -6.83 | 6.07 | -17.57 | 6.20 | No |
| 25 | 10 | 1 | 2 | 1 | 137 | 57 | 41.61 | 1424 | 38.97 | 2.63 | 4.15 | -5.28 | 11.00 | No |
| 25 | 11 | 1 | 3 | 0 | 67 | 23 | 34.33 | 1424 | 38.97 | -4.65 | 5.66 | -14.89 | 7.29 | No |
| 25 | 12 | 1 | 3 | 1 | 144 | 74 | 51.39 | 1424 | 38.97 | 12.41 | 4.11 | 4.32 | 20.43 | Yes |

**Table C.** Geographical spread of the stratum deviations (Dev_A,SD) across the 25 areas, by sociodemographic stratum — Psychotropic medication use.

For each of the 12 sociodemographic strata, its deviation Dev_A,SD (the stratum prevalence minus the crude prevalence of its own area, in percentage points) is summarised across the 25 areas. Columns: Mean (unweighted mean across the 25 areas), Min and Max (smallest and largest deviation), Range (Max − Min), Poor and Rich (mean deviation in poor and in rich areas), Rich − Poor (their difference). Stratum labels: W = women, M = men; age band in years; Low / High = income below / above the Malmö median. A wide range indicates strong geographical modification of the stratum position; a small Rich − Poor difference indicates that area income explains little of it.

| Stratum | Mean | Min | Max | Range | Poor | Rich | Rich-Poor |
| --- | --- | --- | --- | --- | --- | --- | --- |
| W 35-44 Low | 1.7 | -5.7 | 9.2 | 15.0 | 1.2 | 2.3 | 1.1 |
| W 45-54 Low | 11.8 | 4.1 | 21.8 | 17.7 | 12.7 | 10.9 | -1.8 |
| W 55-64 Low | 15.4 | 7.7 | 25.0 | 17.4 | 15.1 | 15.6 | 0.5 |
| W 35-44 High | -5.8 | -10.4 | -1.0 | 9.4 | -6.2 | -5.4 | 0.8 |
| W 45-54 High | 0.9 | -6.1 | 5.2 | 11.2 | -0.0 | 1.9 | 1.9 |
| W 55-64 High | 5.0 | -1.4 | 12.8 | 14.1 | 3.7 | 6.4 | 2.7 |
| M 35-44 Low | -4.0 | -15.2 | 8.8 | 23.9 | -5.5 | -2.4 | 3.1 |
| M 45-54 Low | 2.6 | -5.4 | 11.4 | 16.8 | 1.7 | 3.5 | 1.8 |
| M 55-64 Low | 2.2 | -5.8 | 12.3 | 18.1 | 2.3 | 2.2 | -0.1 |
| M 35-44 High | -12.3 | -18.8 | -7.3 | 11.5 | -12.5 | -12.2 | 0.3 |
| M 45-54 High | -8.7 | -13.5 | -1.3 | 12.2 | -8.2 | -9.2 | -1.0 |
| M 55-64 High | -6.7 | -16.3 | -1.6 | 14.7 | -7.1 | -6.2 | 0.9 |

Table D. Geographical spread of the stratum deviations (Dev_A,SD) across the 25 areas, by sociodemographic stratum — Private GP choice. Columns and stratum labels as in Table C.

| Stratum | Mean | Min | Max | Range | Poor | Rich | Rich-Poor |
| --- | --- | --- | --- | --- | --- | --- | --- |
| W 35-44 Low | -6.6 | -21.3 | 3.9 | 25.2 | -4.8 | -8.5 | -3.7 |
| W 45-54 Low | -5.1 | -25.2 | 11.0 | 36.2 | -4.6 | -5.6 | -1.0 |
| W 55-64 Low | -4.6 | -20.0 | 10.6 | 30.6 | -2.9 | -6.4 | -3.5 |
| W 35-44 High | 3.2 | -5.1 | 14.0 | 19.0 | 3.0 | 3.4 | 0.4 |
| W 45-54 High | 3.6 | -6.1 | 23.4 | 29.5 | 4.5 | 2.6 | -1.9 |
| W 55-64 High | 6.3 | -8.4 | 18.3 | 26.6 | 7.0 | 5.5 | -1.5 |
| M 35-44 Low | -10.6 | -30.6 | 3.8 | 34.3 | -8.5 | -12.8 | -4.3 |
| M 45-54 Low | -7.5 | -31.3 | 16.4 | 47.7 | -6.8 | -8.4 | -1.6 |
| M 55-64 Low | -5.1 | -18.0 | 12.0 | 30.0 | -4.0 | -6.4 | -2.4 |
| M 35-44 High | 4.6 | -4.4 | 15.3 | 19.7 | 4.4 | 4.9 | 0.5 |
| M 45-54 High | 4.5 | -3.7 | 21.1 | 24.8 | 4.0 | 5.1 | 1.1 |
| M 55-64 High | 7.2 | -1.4 | 23.6 | 25.0 | 6.8 | 7.6 | 0.8 |
