## Supplementary material for "Describing health inequalities without distortion: Simple-Means MAIHDA vs Random-Effects MAIHDA": S3 Text. The compare measures

### Conceptual guide to the comparative measures reported by the *compare* option of *smaihda*

### 1. Why compare simple means and random effects

Multilevel Analysis of Individual Heterogeneity and Discriminatory Accuracy (MAIHDA) is most often implemented through random-effects multilevel logistic regression with random intercepts at the level of the sociodemographic-by-context strata. Under this approach, each stratum receives a shrunken predicted probability: an empirical Bayes point estimate that pulls the observed stratum proportion toward the grand mean of the data.

Shrinkage is the deliberate consequence of treating strata as exchangeable realisations of a common distribution. Its magnitude scales inversely with stratum size: the smaller the stratum, the more its observed proportion is pulled towards the grand mean, because a small stratum carries less information about its own risk. It is easiest to see for a continuous outcome, where the random-effects prediction for stratum j is a reliability-weighted average of the stratum’s own observed mean and the grand mean (Leckie et al. 2025):

m̂ⱼ = wⱼ ȳⱼ + (1 − wⱼ) μ

where μ is the grand mean of the strata and the weight is the reliability of the observed mean as an estimate of the stratum’s true mean. The weight is the ratio of the true-mean variance to the observed-mean variance, and it runs from 0 to 1. Note that μ is not the population mean. It is a mean of the stratum means in which each stratum is weighted by its own reliability, so it lies between the population mean and a mean in which every stratum counts equally, and it approaches the latter as the strata grow large:

wⱼ = σ²ᵤ / (σ²ᵤ + vⱼ)

with σ²ᵤ the between-stratum variance and vⱼ the sampling variance of the stratum’s own mean. For a continuous outcome vⱼ = σ²ₑ/nⱼ, the within-stratum individual variance divided by the stratum size. The amount the model removes from a stratum is (1 − wⱼ) times its distance from the grand mean. A large stratum has a small vⱼ, a weight near one, and little shrinkage. A small stratum has a large vⱼ, a weight near zero, and strong shrinkage towards the grand mean.

The outcomes in this paper are binary, so the same logic operates on the logit scale, and predictions are read back as probabilities. The structure of the weight is unchanged. Only the sampling variance changes. For a binary outcome the observed stratum logit has sampling variance vⱼ = 1/(nⱼ pⱼ(1 − pⱼ)), where pⱼ is the observed prevalence. This is again an individual variance divided by n, now with the binomial information pⱼ(1 − pⱼ) carried by each individual.

The logistic model has no closed-form weight, because the stratum effects are predicted by integrating over a non-linear likelihood, and several approximations exist. We fit the random-effects model with melogit, which estimates by adaptive quadrature and predicts the stratum effects by empirical Bayes. The shrinkage it applies is governed by the information the binary data carry about each stratum, which is nⱼ pⱼ(1 − pⱼ). The reliability-weight expression above, with vⱼ = 1/(nⱼ pⱼ(1 − pⱼ)), is the closed-form approximation to this behaviour. Section 2 sets out the point and the interval in full.

One clarification helps readers familiar with the variance-partition work of Goldstein, Browne and Rasbash. The level-1 logistic variance π²/3 does not enter the shrinkage factor. That quantity belongs to the latent-scale variance partition, a separate convention used for the VPC, and it is taken up there in Section 5. The shrinkage of the per-stratum predictions depends on the binomial information, not on π²/3.

In intersectional designs, the strata with the fewest individuals often represent the most marginalised subpopulations: rare combinations of low income, low education, particular age groups and particular residential areas. The reliability-weighted logic, applied to such designs, attenuates precisely the predictions about the groups whose specific risks the analysis was intended to make visible. The simple-means approach reports the observed proportion in each stratum without shrinkage, and so does not exhibit this attenuation.

Whether the attenuation introduced by shrinkage is a feature or a defect of the random-effects approach is the central question that the comparison is designed to answer empirically. The *compare* option of the *smaihda* command fits a standard random-effects MAIHDA via *melogit* on the same A-SD strata used by the simple-means analysis, and reports nine quantities that characterise the difference between the two methods. These quantities do not arbitrate between methods, but they make the practical consequences of choosing one over the other visible and quantifiable for any given dataset.

The concepts introduced here are taken up again, with full detail, in the sections that follow. This section gives the intuition but sections that follow set out the same ideas formally, with the point estimate, the interval and a worked example.

### 2. How shrinkage works: the weight, the point, and the interval

**2.1 What shrinkage is**

Random-effects estimation does not return a stratum’s own summary. For each stratum it returns a posterior estimate that blends the stratum’s own data with the distribution of all strata. Because the model treats the strata as exchangeable draws from a common normal distribution, it pulls every stratum’s estimate towards the grand mean. The pull is gentle for strata that carry a lot of information and strong for strata that carry little. This pull is shrinkage.

Two quantities govern its strength: the between-stratum variance σ²ᵤ, which says how far apart the strata really sit, and the amount of data in the stratum, which says how much its own mean can be trusted. Everything below makes that statement precise.

**2.2 The reliability weight**

Write the grand mean as μ, a stratum’s own observed mean as ȳⱼ, the between-stratum variance as σ²ᵤ, and the sampling variance of the stratum’s own mean as vⱼ. For a continuous outcome vⱼ = σ²ₑ/nⱼ, where σ²ₑ is the within-stratum variance and nⱼ the stratum size. For a binary outcome the same role is played, on the logit scale, by

vⱼ = 1 / (nⱼ pⱼ(1 − pⱼ))

The reliability weight is

wⱼ = σ²ᵤ/(σ²ᵤ + vⱼ) = σ²ᵤ/(σ²ᵤ + σ²ₑ/nⱼ)

It is the share of the stratum’s apparent departure that the model keeps as signal. When σ²ᵤ is large or nⱼ is large, vⱼ is small beside σ²ᵤ, wⱼ approaches one, and the stratum’s own data dominate. When σ²ᵤ is small or nⱼ is small, wⱼ approaches zero, and the grand mean dominates. The complement (1 − wⱼ) is the share the model treats as noise and removes.

The between-stratum variance σ²ᵤ is also what the GCE rests on, because the GCE is reported as the VPC, which is a function of σ²ᵤ. So, a small GCE and a strong shrinkage are two readings of the same small σ²ᵤ.

**2.3 Shrinkage and the specific contextual effect**

The shrunken estimate is the precision-weighted posterior mean,

m̂ⱼ = μ + wⱼ(ȳⱼ − μ)

Under simple-means the specific contextual effect of the stratum is its full departure from the grand mean,

(ȳⱼ − μ).

Under random effects that departure becomes

wⱼ(ȳⱼ − μ), attenuated by the factor (1 − wⱼ).

The attenuation has a direction. It always pulls towards μ, the point of no inequality. So, shrinkage does not scatter the estimate around the truth. It moves it systematically towards the centre. A small stratum that genuinely sits far from the average is reported closer to the average than it is. Reading the confidence interval correctly does not put the point back where the data placed it.

**2.4 Shrinkage and the interval**

The posterior variance of the shrunken estimate is

Var(m̂ⱼ) = σ²ᵤ(1 − wⱼ) = 1/(1/σ²ᵤ + nⱼ/σ²ₑ)

so the credible interval half-width is proportional to √(σ²ᵤ(1 − wⱼ)). The two limits mirror the point. When nⱼ is large, wⱼ approaches one, the variance approaches vⱼ = σ²ₑ/nⱼ, and the interval matches the simple-means interval. When nⱼ is small and σ²ᵤ is small, wⱼ approaches zero, the variance approaches σ²ᵤ, and the interval collapses to the narrow between-stratum scale.

The single fact that matters is this. The same weight wⱼ governs the point in Section 2.3 and the interval here. For a small stratum sitting on a small σ²ᵤ, the point and the interval drawn in around are pulled to the grand mean at the same time. The interval is narrow not because the stratum’s own data are informative, but because the model has lent it precision borrowed from the other strata. The result is a tight interval around a displaced point. The estimate looks both attenuated and certain.

Simple means behave in the opposite way. The estimate is ȳⱼ, unbiased for the stratum’s own proportion, and its interval half-width is √(σ²ₑ/nⱼ), or the Wilson interval for a proportion. For a small stratum this interval is wide. The width is not a defect. It is an honest disclosure of how little information the stratum’s own data carry.

**2.5 A worked binary example**

The outcomes in this paper are binary, so the mechanics operate on the logit scale, and the predictions are read back as probabilities. Take a population prevalence of 0.10, so the grand mean on the logit scale is logit(0.10) = −2.20. Consider a small stratum of 25 individuals with five cases, an observed prevalence of 0.20, or logit(0.20) = −1.39. Its departure from the grand mean is 0.81 on the logit scale, and the sampling variance of that logit estimate is vⱼ = 1/(25 × 0.20 × 0.80) = 0.25.

Suppose the between-stratum variance is σ²ᵤ = 0.10, a small GCE. The weight is wⱼ = 0.10/(0.10 + 0.25) = 0.29. The shrunken departure is 0.29 × 0.81 = 0.23, so the shrunken logit is −1.97 and the shrunken prevalence is 0.12. The observed 0.20 has been pulled most of the way back to the grand mean of 0.10.

The intervals complete the picture. Simple means report 0.20 with a Wilson 95% interval of about 0.09 to 0.39, wide because 25 individuals carry little information. Random effects report 0.12 with a credible interval of about 0.08 to 0.19. The random-effects interval is narrower than the simple-means one, and it excludes the observed 0.20 entirely. The same small σ²ᵤ that produced a small GCE has pulled the point down and tightened the interval around it. The stratum’s real elevation is now invisible, and the narrow interval lends the attenuated figure an unwarranted confidence.

*Worked contrast: the same formula in two worlds.* The reliability weight w = σ²ᵤ/(σ²ᵤ + σ²ₑ/n) is one formula, but its consequences depend entirely on where it is applied. Consider it in its legitimate home first. A patient. Systolic blood pressure in a clinic population: between-patient standard deviation 15 mmHg, within-patient standard deviation 10 mmHg, so the intraclass correlation is 225/325 = 0.69, a typical value for a stable physiological quantity. A patient whose three measurements average 160 mmHg, against a clinic mean of 130, receives weight w = 225/(225 + 100/3) = 0.87. The empirical Bayes prediction is 130 + 0.87 × 30 = 156 mmHg, with a credible interval of roughly 146 to 167. Of a 30 mmHg deviation, 87% survives three measurements. The patient remains above the 140 mmHg threshold; the value is smoothed, not hidden. Here exchangeability has a warrant: the measurements fluctuate around a homeostatic set point that exists, so the latent mean has a referent and the fluctuation around it is noise. A stratum. Now the intersectional stratum above: overall prevalence 10%, a stratum of 25 people with an observed prevalence of 20%, double the average, and a between-stratum variance of 0.10 on the logit scale, a VPC of about 3%, at the low end of the 3% to 15% range typical of published MAIHDA studies. The weight is w = 0.29. The prediction returns 12.3%: of the 10 percentage-point excess, 2.3 points survive twenty-five people. The credible interval, 7.7% to 19.1%, excludes the observed value. If a monitoring programme flagged strata above 15%, this stratum, whose members experience the outcome at twice the average rate, would vanish from the list. Here exchangeability has no warrant: there is no social set point around which the stratum fluctuates; the 20% is not noise around a latent essence, it is the phenomenon. The contrast is the point. The patient keeps 87% of a real deviation with three measurements; the stratum keeps 23% of a real excess with twenty-five people. Same formula, opposite consequences, because the ICC is high where the exchangeability assumption is mechanistically defensible and the VPC is low where it is not. The machinery is gentle where it is warranted and severe where it is not, which is why a method that is standard in clinical repeated measures becomes a distortion in descriptive social epidemiology.

**2.6 Credible intervals and bootstrap intervals**

The interval attached to a shrunken estimate is a credible interval. There are two routes to it, and they differ only in detail.

The analytic empirical-Bayes interval is the one written in Section 2.4. It is centred on the shrunken point m̂ⱼ with half-width proportional to √(σ²ᵤ(1 − wⱼ)). It conditions on the fitted model and on a single estimate of σ²ᵤ.

The Markov chain Monte Carlo interval comes from a fully Bayesian fit that places priors on σ²ᵤ and on the stratum effects and samples the posterior directly. It gives the same picture, a point pulled towards the grand mean and an interval conditioned on the assumed distribution of strata. The one difference is that the chains also carry the uncertainty in σ²ᵤ itself, which the analytic version fixes at a point. So, the chain intervals are usually a little wider than the analytic ones. They remain narrower than the stratum’s own data-based interval for a small stratum and still centred on a shrunken point.

The bootstrap and the analytic frequentist intervals are a different family. The non-parametric bootstrap resamples the individuals inside each cell and recomputes the quantity. The Wilson interval and the delta method work from the stratum’s own counts. None of these uses a prior or the between-stratum variance. They are centred on the unbiased observed estimate, and they say what the stratum’s own data support, no more and no less.

The divide is therefore interpretive, not technical. A credible interval, analytic or from chains, answers a conditional question: given the model and the assumed distribution of strata, what is the latent stratum effect. A bootstrap or Wilson interval answers a direct question: what does this stratum’s own data support. For a small stratum the two can disagree sharply, the credible interval narrow around a shrunken point, the data-based interval wide around the observed one. That disagreement is not an error in either. It is the signature of shrinkage, and it is what the comparison is built to reveal.

For descriptive inequality on near-complete population data the question the paper asks is the direct one. The observed stratum proportions are the realised inequality, and the data-based interval is the faithful statement of their uncertainty. The credible interval remains the right tool when the target is the latent effect under an explicit exchangeability assumption. The *compare* option reports both, so the applied researcher can see, stratum by stratum, how far the two accounts diverge.

### 3. Framework: nine measures across three MAIHDA components

The nine measures produced by *compare* span the three components of the MAIHDA framework:

| Component | Measures | Count |
| --- | --- | --- |
| Specific contextual effects (SCE) | Range of stratum predictions (SM, RE), range compression, SD of stratum predictions (SM, RE), mean absolute attenuation, maximum absolute attenuation with stratum size n | 5 |
| General contextual effects (GCE) | Variance partition coefficient (VPC) at the A-SD stratum level on the probability scale | 1 |
| Discriminatory accuracy (DA) | Contextual area under the ROC curve (coAUC) at the A-SD level, sensitivity at the population-prevalence threshold, specificity at the population-prevalence threshold | 3 |

The five SCE measures reflect that shrinkage acts first on the per-stratum predictions, so the comparison records both the summary amount of compression and the worst-case displacement. The single GCE measure and the three DA measures serve as summary-level checks that the two methods capture the same overall amount of contextual structuring and the same ranking of strata by risk.

The main paper reports six of these nine measures (Section 3.4, Table 8): range compression and maximum absolute attenuation for SCE, the VPC for GCE, and the coAUC, sensitivity and specificity for DA. The remaining three measures are described in the present document and reported in full in Table C in S1 Text.

### 4. Description of each measure

Each measure is presented in three components: a formal definition, a description of the calculation procedure in non-technical terms, and the interpretation.

#### Specific contextual effects (SCE)

##### **1. Range of stratum predictions (SM and RE)**

*Definition.* The difference between the highest-risk and the lowest-risk A-SD stratum on the predicted prevalence scale, computed separately under each method.

*Calculation.* For each method, identify the A-SD stratum with the highest predicted prevalence and the one with the lowest, and take the difference.

*Interpretation.* The range is the simplest visual summary of how far apart a method places the strata on the prevalence scale. A wide range means the method preserves marked between-stratum differences; a narrow range means the strata are crowded together near the grand mean. Because the random-effects model shrinks extreme strata toward the grand mean, the random-effects range is typically smaller than the simple-means range. The gap between the two ranges is the most direct visual expression of how much of the empirical inequality has been suppressed by the model.

##### **2. Range compression (RE / SM, %)**

*Definition.* The random-effects range expressed as a percentage of the simple-means range.

*Calculation.* Divide RE_range by SM_range and multiply by 100.

*Interpretation.* A value of 100% means that the random-effects model preserves the full range of inequality that the observed proportions reveal. A value of 50% means that the random-effects model has cut the inequality range in half. Lower values reflect stronger shrinkage. The complement of this measure (100 minus the reported value) is the share of the empirical inequality range that has been hidden from view.

##### **3. Standard deviation of stratum predictions (SM and RE)**

*Definition.* The standard deviation of stratum-specific predicted prevalences across the A-SD strata, computed separately under each method.

*Calculation.* Treat each A-SD stratum as a single observation with its predicted prevalence and compute the standard deviation across strata.

*Interpretation.* A complementary measure of dispersion that is less sensitive to extremes than the range. A smaller RE_SD relative to SM_SD indicates that shrinkage has compressed not only the extreme strata but also the central body of the stratum distribution.

##### **4. Mean absolute attenuation**

*Definition.* The average size, across all A-SD strata, of the absolute difference between the simple-means prediction and the random-effects prediction.

*Calculation.* For each stratum, take the absolute difference between the observed proportion (simple means) and the shrunken predicted probability (random effects). Average these absolute differences across strata.

*Interpretation.* This summarises how far the typical stratum prediction has been moved by shrinkage, expressed in percentage points. A mean absolute attenuation of 1.32 pp (psychotropic medication use, Malmö), for example, means that on average the random-effects prediction sits that many percentage points away from the value the observed proportion would assign. Because shrinkage always pulls toward the grand mean, this is a direct measure of how much epidemiological information about the typical stratum has been displaced.

##### **5. Maximum absolute attenuation with stratum size n**

*Definition.* The largest absolute difference between the simple-means and random-effects predictions across all A-SD strata, reported together with the size n of the stratum where it occurs.

*Calculation.* Identify the A-SD stratum with the largest absolute difference between observed proportion and random-effects predicted probability. Report both the magnitude of the attenuation and the number of individuals in that stratum.

*Interpretation.* This identifies the stratum whose prediction has been moved the most by the random-effects model. The displacement depends on two factors: the stratum size, through the reliability weight, and the distance of its observed proportion from the grand mean. The absolute attenuation is (1 - w_j) times that distance, so the largest displacement falls on a stratum that is both small, so w_j is low, and far from the mean. This is why n is reported with the value. A finding such as 6.44 pp in a stratum of n = 75 (psychotropic medication use, Malmö) expresses how much epidemiological information about a small, extreme group has been suppressed by the model. In intersectional designs the smallest strata often represent the most marginalised subpopulations, so this shrinkage falls preferentially on the groups whose specific risks the analysis is intended to make visible.

#### General contextual effects (GCE)

##### **6. VPC at the A-SD stratum level on the probability scale**

*Definition.* The share of the total individual variance, on the probability scale, that lies between the A-SD strata. It is the variance-partition coefficient computed so that the simple-means and random-effects values rest on the same footing.

*Calculation.* The general contextual effect is a property of the contextual boundaries, independent of how many individuals fall on each side, so the simple-means side is reported as VPC*, in which all occupied A-SD strata are given the same size. The between-stratum component is the variance of the observed stratum proportions with weight one per stratum, and the total is p*(1−p*) at the standardised mean. Two forms are reported: uncorrected, and corrected for binomial noise, which reassigns the sampling component of each stratum from the between-stratum level to the individual level so that the identity is preserved. The crude VPC of the main text remains available, but the standardised version is the one compared here, because it measures the general contextual effect independently of composition and because the random-effects τ² treats strata as units, which is the same equal weighting.

For the random-effects side, fit melogit on the A-SD strata with a random intercept, then place the coefficient on the probability scale by simulation, following Method B of Goldstein, Browne and Rasbash (Goldstein, Browne, and Rasbash 2002). Draw a large number of stratum residuals from the fitted normal distribution N(0, σ²ᵤ). Convert each to a predicted probability π through the inverse logit. Compute v2, the variance of these probabilities, which is the between-stratum component, and v1, the mean of π(1−π) across the draws, which is the within-stratum component. The coefficient is

VPC_RE = v2 / ( v2 + v1 ).

The denominator v2 + v1 is the total individual variance on the probability scale, so the coefficient is the between-stratum share of that total. It is therefore comparable, term for term, with the simple-means value.

*Interpretation.* Both methods are expressed on the same probability scale, so their values are directly comparable. The simple-means side is shown in two forms. The uncorrected standardised VPC is the observed reality, which in a total-population register is not a sample estimate. The corrected form removes the binomial sampling component and therefore sits on the same noise-discounted footing as the random-effects τ², which is the fairer comparison of contextual structuring; in a design with many small strata this correction is decisive, whereas in a balanced design it barely moves the value. Close agreement between the random-effects VPC and the corrected standardised VPC indicates that the two methods capture the same general contextual effect, even when their per-stratum predictions diverge in magnitude. A difference reflects the distributional assumption of the random-effects model, the normal prior on the stratum effects, not a difference of scale. The probability-scale partition is distinct from the latent-scale partition, which fixes the individual variance at π²/3, and the two need not agree. This scale question is taken up in Section 5.

*Boundary behaviour with sparse information.* The corrected components are estimated by subtraction, so sampling error can carry them slightly below zero when the true between-stratum variance is close to the binomial noise. A variance cannot be negative. A negative corrected value is the measurement error of a true zero, and the command floors the corrected components at zero. When the corrected between-stratum variance is zero, the corrected CCI and the CCI, G and CMI shares are not defined, because they are shares of a quantity that is absent. The command reports them as missing and prints that no contextual clustering is detectable beyond binomial noise.

*Sparse designs.* Two further safeguards protect designs in which many strata hold few individuals. First, the interval of the deviation of a stratum from its area (STEP 4) is the Wilson interval of the stratum shifted by the area proportion. A delta-method interval has zero width when everyone, or no one, in the stratum has the outcome, so that every such stratum would be declared significantly deviating whatever its size. Second, the standardised partition and the CCI/G/CMI shares are computed on strata with at least minn() individuals, default 10. Where many strata hold fewer than five individuals, these measures are dominated by binomial sampling noise. A noise term under-corrects in strata at 0% or 100%, and a stronger noise term over-corrects, so that no noise term recovers the components; a minimum stratum size does. The command also prints a cell-size diagnostic and a note when the area-by-stratum grid is incomplete. None of this affects the Malmö analysis, whose smallest stratum has 35 individuals.

What follows describes what to expect when the command is run on data with a rare outcome or very small strata, that is, with few expected cases per stratum. The specific contextual effects are always delivered: every stratum keeps its observed proportion and its Wilson interval. The intervals are wide, and that width is the answer, imprecision made visible rather than removed. The crude VPC remains valid and is typically small. The corrected standardised VPC may reach its floor at zero, with the message above, while the uncorrected form stays positive because it still contains the binomial noise. The gap between the two forms is itself informative: it is the share of the visible dispersion that the data cannot distinguish from noise.

On the random-effects side, *melogit* typically either fails to converge, in which case the command says so and skips the comparison, or converges to the zero-variance boundary. In the second case the variance parameter is close to zero, every stratum prediction collapses towards the grand mean, and the *compare* block shows a strong range compression. This is case B2 of the interpretative rule: the random-effects model finds no inequality, but compression alone could have produced that finding, so an absence of inequality cannot be concluded.

The reading is the same on both sides. The data do not contain enough information to support the chosen stratification. Report the observed proportions with their intervals, state their imprecision plainly, and reconsider whether the stratification fits the size of the dataset. Neither method can supply information the data do not contain. The difference is visibility: the simple-means output declares the shortage, while a random-effects model at the zero-variance boundary can look like a secure finding of no inequality.

#### Discriminatory accuracy (DA)

##### **7. coAUC at the A-SD stratum level**

*Definition.* The area under the ROC curve when individuals are scored by the predicted risk of their A-SD stratum.

*Calculation.* Assign each individual the stratum-specific predicted risk produced by the method (observed proportion for simple means, shrunken predicted probability for random effects). Compute the area under the ROC curve as the probability that a randomly chosen individual with the outcome receives a higher stratum score than a randomly chosen individual without the outcome.

*Interpretation.* The coAUC depends only on the ranking of strata by predicted risk, not on the absolute magnitude of the predictions. When stratum sizes are sufficiently uniform, random-effects shrinkage is approximately monotonic and the ranking of strata is preserved, so the two methods produce identical or near-identical coAUCs. In intersectional designs with stratum sizes that differ substantially, differential shrinkage can reorder some strata and lower the random-effects coAUC below the simple-means coAUC. Close agreement in coAUC therefore indicates that the random-effects model has not reordered the strata, regardless of the magnitude of per-stratum attenuation.

##### **8. Sensitivity at the population-prevalence threshold**

*Definition.* The proportion of individuals with the outcome who belong to A-SD strata whose predicted risk equals or exceeds the population prevalence.

*Calculation.* Set the classification threshold equal to the sample population prevalence of the outcome. Flag each A-SD stratum as high-risk if its predicted prevalence is at or above the threshold. Among individuals with the outcome, compute the share who fall into high-risk strata.

*Interpretation.* Sensitivity quantifies how well a targeted strategy directed at high-risk strata would capture the cases. A high sensitivity means most cases are concentrated in the strata identified as high-risk; a low sensitivity means many cases lie outside the high-risk strata and would be missed by a targeted approach. Comparing sensitivity between methods shows whether shrinkage has moved enough stratum predictions across the threshold to change which strata are flagged as high-risk, and therefore how many cases the same targeted strategy would reach in practice.

##### **9. Specificity at the population-prevalence threshold**

*Definition.* The proportion of individuals without the outcome who belong to A-SD strata whose predicted risk lies below the population prevalence.

*Calculation.* At the same threshold used for sensitivity, flag each A-SD stratum as low-risk if its predicted prevalence is below the population prevalence. Among individuals without the outcome, compute the share who fall into low-risk strata.

*Interpretation.* Specificity quantifies how well a targeted strategy correctly avoids non-cases. A high specificity means non-cases are concentrated in the strata identified as low-risk; a low specificity means many non-cases would be unnecessarily included in a targeted intervention. The trade-off between sensitivity and specificity at the population-prevalence threshold is the empirical expression of Rose’s prevention paradox at the A-SD stratum level.

### 5. The choice of scales: a worked example

The scales used by *compare* reflect different rationales for different families of measures. The per-stratum predictions, the attenuation summaries, and the sensitivity and specificity at the population-prevalence threshold are reported on the proportion scale because that is the scale on which public health interpretation operates. The variance partition coefficient is also reported on the probability scale, so that both methods sit on a single, directly interpretable footing. The coAUC requires no scale choice, since it depends only on the ranking of strata.

The choice of the proportion scale for the per-stratum attenuation is consequential and worth illustrating numerically. Reliability-weighted shrinkage operates on the latent logit scale: each stratum’s deviation from the grand mean is multiplied by its shrinkage factor λ_j. Reporting attenuation on the logit scale would show only this operation. Reporting it on the proportion scale captures both this operation and the non-linear back-transformation through the inverse logit, which amplifies attenuation in strata whose prevalence is near 50% and reduces it for strata at the extremes of the prevalence distribution.

A numerical example clarifies the difference. Suppose the grand mean on the logit scale is β₀ = −1.0, which corresponds to a population prevalence of approximately 27%. Consider two strata that are each shrunk by 0.5 logit units toward the grand mean. The first stratum has an empirical logit of −3.0, corresponding to a prevalence of 4.7%, and the second has an empirical logit of +1.0, corresponding to a prevalence of 73.1%.

| Stratum | Empirical logit | Empirical prevalence | Post-shrinkage logit | Post-shrinkage prevalence | Attenuation in pp |
| --- | --- | --- | --- | --- | --- |
| 1 | −3.0 | 4.7% | −2.5 | 7.6% | 2.9 |
| 2 | +1.0 | 73.1% | +0.5 | 62.2% | 10.9 |

The same 0.5 units of logit shrinkage produce 2.9 percentage points of attenuation in the low-prevalence stratum and 10.9 percentage points in the high-prevalence stratum, a factor of nearly four. On the logit scale, the two strata would appear to have been moved identically; on the proportion scale, the difference is large. The proportion scale is the scale on which the consequences of shrinkage become legible to the reader of an epidemiological paper, because that is the scale on which prevalence is interpreted and on which public health decisions are framed.

The variance partition coefficient is reported on the probability scale. A binary outcome has a total individual variance of p(1−p), and the partition expresses how much of this total lies between A-SD strata. The simple-means side reads this directly from the observed decomposition; the random-effects side is placed on the same scale by simulation from the fitted model (Goldstein, Browne and Rasbash 2002). Goldstein and colleagues show that the probability-scale partition and the latent logit partition are different quantities: the latent version fixes the individual variance at π²/3, whereas the probability version uses the actual Bernoulli variance, and the two need not agree. We report the probability-scale partition because it is the scale on which the rest of the comparison operates and because it places both methods on a common, directly interpretable footing. The probability-scale partition does depend on the population prevalence, so for comparison across outcomes or studies it should be read together with that prevalence.

### 6. How to read the *compare* output

The *compare* option produces its output in the Stata console in three blocks, mirroring the three MAIHDA components:

- **Per-stratum predictions and attenuation (proportion scale)** reports the five SCE measures: minimum and maximum predicted prevalence under each method, range and range compression, SD of stratum predictions under each method, mean and maximum absolute attenuation, and the size n of the stratum at the maximum attenuation.
- **General contextual effect (probability-scale VPC)** reports the random-effects VPC against the simple-means VPC* in two forms, uncorrected and corrected for binomial noise. VPC* measures the general contextual effect independently of composition; the corrected form is the one directly comparable with the random-effects τ².
- **Discriminatory accuracy** reports the coAUC under both methods, and the sensitivity and specificity at the population-prevalence threshold.

The simple-means estimates retain the 95% confidence intervals reported in Sections 3.1 to 3.3 of the main paper. The random-effects estimates produced by *compare* are single point estimates without bootstrap intervals, since refitting *melogit* within each of 2,000 bootstrap replicates would impose a runtime cost without changing the conceptual basis of the comparison.

The full Stata syntax for invoking *compare* and the equivalent step-by-step implementation in plain Stata are documented in Step 11 of S1 Text.

Goldstein, H, W Browne, and J Rasbash. 2002. ‘Partitioning Variation in Multilevel Models’, *Understanding Statistics*, 1: 223–31.

Leckie, George, Andrew Bell, Juan Merlo, SV Subramanian, and Clare Evans. 2025. ‘The Statistical Advantages of Multilevel Analysis of Individual Heterogeneity and Discriminatory Accuracy for Estimating Intersectional Inequalities’, *SOCIOLOGICAL METHODS & RESEARCH*, 0: 00491241251385123.
