## Supplementary material for "Describing health inequalities without distortion: Simple-Means MAIHDA vs Random-Effects MAIHDA": S4 Text. Reanalysis of published data (opioid misuse)

**A reproducible demonstration of the *compare* diagnostic: prescription opioid misuse in the United States**

**S4.1 Purpose**

This Supplementary Information illustrates the *compare* diagnostic on a published and fully reproducible example. The aim is didactic. We show, on real data, how the random-effects assumption changes the stratum-specific picture that Simple-Means MAIHDA (S-MAIHDA) reports from the same data.

We chose this example for three reasons. First, the data are public, because the analysis rests on the public-use file of the 2015 National Survey on Drug Use and Health. Second, the per-stratum results are already in print, so any reader can reproduce the comparison directly from the published table. Third, the strata differ widely in size and the outcome is uncommon, which is the setting in which shrinkage is expected to act, and a useful contrast to the large and reasonably uniform strata of the Malmo application in the main text.

The example comes from earlier work by our own group. We revisit it here in a constructive spirit, to demonstrate the diagnostic, and not to revise the conclusions of the original study.

**S4.2 The study and the data**

Persmark and colleagues studied prescription opioid misuse in the adult population of the United States. They built 72 intersectional strata from the combination of gender, race or ethnicity, income, and age, and nested 43,409 individuals within them. The outcome was self-reported prescription opioid misuse.

The strata differ widely in size, from 50 to 3,030 individuals. Fourteen strata contain fewer than 100 individuals. The overall prevalence of the outcome is 6.08%, so this is an uncommon outcome.

We reanalyse the empty model, that is, the model with an intercept and a random effect for the strata and no covariates. The published table reports, for every stratum, the observed percentage with the outcome and the predicted absolute risk from the random-effects model. These two columns are all we need. We analyse the data without survey weights, because the purpose here is to illustrate a statistical property and not to produce weighted national estimates.

**S4.3 The two estimators, side by side**

Simple-Means MAIHDA (S-MAIHDA) estimates each stratum by its observed proportion. This is the unbiased maximum-likelihood estimate for that stratum, computed from its own individuals. Random-Effects MAIHDA (RE-MAIHDA) replaces the observed proportion with a precision-weighted prediction that is pulled toward the population average. The pull is stronger for the smaller strata, which carry less information.

The published table gives both quantities for every stratum. The observed percentage is the S-MAIHDA estimate. The predicted absolute risk is the RE-MAIHDA estimate. The comparison below is therefore a direct reading of the two columns, with no further modelling required. The full table of 72 strata is reproduced at the end of this document as Table S4-D.

**S4.4 Result of the *compare* diagnostic**

**Table S4-A. The *compare* diagnostic on the probability scale, S-MAIHDA against RE-MAIHDA.**

| **Quantity** | **Simple-Means MAIHDA** | **Random-Effects MAIHDA** |
| --- | --- | --- |
| Between-stratum VPC (probability scale) | 1.45% | 1.21% |
| Area under the ROC curve | 64.1% | 64% |
| Range of stratum estimates (lowest to highest) | 16 pp | 11.2 pp |
| High-risk strata at the prevalence threshold | 28 of 72 | 25 of 72 |
| Sensitivity at that threshold | 74.2% | 65.4% |
| Specificity at that threshold | 46.2% | 55% |

*pp, percentage points. The between-stratum variance on the latent (logit) scale is tau-squared = 0.258, which corresponds to a latent-scale VPC of 7.3%. The probability-scale VPC is much lower because the outcome is uncommon.*

On the aggregate measures the two methods agree. The variance partition coefficient is small under both, 1.45% for S-MAIHDA and 1.21% for RE-MAIHDA. A small probability-scale value is expected for an uncommon outcome, because the individual Bernoulli variance is large relative to the between-stratum variance. The area under the ROC curve is almost identical, 64.1% against 64.0%. A researcher who reported only these aggregate numbers would see no difference between the two methods.

**The crude and the standardised VPC.** The variance partition above gives each stratum its number of individuals. Because this design has 72 strata of very unequal size, from 50 to 3,030 individuals, it is worth asking what the VPC looks like when all strata are given the same size, since the general contextual effect is defined independently of composition. Table S4-B reports the crude VPC alongside VPC*, the standardised VPC, in the two forms produced by the smaihda command: uncorrected, and corrected for binomial noise.

**Table S4-B. Crude and standardised between-stratum VPC on the probability scale, with 95% within-context bootstrap confidence intervals (B = 2000, seed 2026): the 72 strata are held fixed and the individuals are resampled inside each stratum, and the percentile interval is recentred on the estimate.**

| VPC | Estimate | 95% CI |
| --- | --- | --- |
| Crude | 1.45% | 1.24 to 1.69 |
| Standardised, uncorrected | 1.80% | 1.25 to 2.68 |
| Standardised, corrected | 1.33% | 0.80 to 2.20 |

Giving all strata the same size raises the VPC from 1.45% to 1.80%, because the small strata with extreme proportions, such as the 16% stratum of 50 individuals, now count as much as the large ones. But at these sizes each observed proportion carries a large binomial sampling variance, and when all strata are given the same size that variance enters the between-stratum component with full force. The correction, which reassigns the sampling component to the individual level, brings the coefficient down to 1.33%, close to the crude value and below the uncorrected one. The uncorrected figure is not wrong. In a total-population register it is the observed reality. But it is inflated by sampling noise, and the correction separates the noise from the between-stratum signal. The intervals show the same thing from the other side. Holding the strata fixed and resampling the individuals inside them gives a narrow interval for the crude coefficient, 1.24 to 1.69, because it is dominated by the large strata, and a wide one for the standardised coefficients, because under equal weighting the small strata carry their full binomial variability into the estimate.

This is the practical lesson of the example. In the balanced Malmö design of the main text the noise correction barely moved the VPC, because the strata are large. In a typical intersectional design such as this one, with many small strata, the correction is decisive. Reporting both forms, as the command does, lets the reader see the observed value and the noise-discounted value side by side.

The aggregate agreement, however, conceals a redistribution at the level of the individual strata. RE-MAIHDA compresses the spread of the stratum estimates. The observed proportions span 16.0 percentage points from the lowest stratum to the highest. The random-effects predictions span only 11.2 points. The random-effects model retains 70% of the observed spread and removes 30% of it.

**S4.5 Where the two methods diverge: the small, high-risk strata**

The compression is not uniform. It falls on the small strata. Among strata with fewer than 100 individuals the average attenuation is 1.71 percentage points. Among the larger strata it is 0.41 points. The largest strata are barely touched. The three largest, each above 2,600 individuals, are displaced by 0.04 points or less.

The single most attenuated stratum is informative. It is the stratum of African-American women, of high income, aged 18 to 29, with 50 individuals. Its observed prevalence of prescription opioid misuse is 16.0%, the highest of any stratum. RE-MAIHDA pulls this estimate down to 10.2%, an attenuation of 5.76 percentage points. The Wilson 95% interval for the observed proportion is wide, from 8.3% to 28.5%, which is the honest reflection of a small sample. S-MAIHDA reports the unbiased estimate of 16.0% together with this wide interval. It states what was observed and is explicit about the imprecision. RE-MAIHDA reports a narrower and more confident value of 10.2% that sits closer to the population average.

Shrinkage acts in both directions. Small strata with no observed cases are pulled up toward the average, and small strata with high observed risk are pulled down. In each case the distinctive value of the small stratum is moved toward the centre. The strata below show this pattern, with the largest strata included for contrast.

**Table S4-C. The most attenuated strata, with the largest strata shown for contrast.**

| **Stratum (race, gender, income, age)** | **N** | **Observed %** | **RE predicted %** | **Attenuation (pp)** |
| --- | --- | --- | --- | --- |
| Afr. American, female, high, 18–29 | 50 | 16.0 | 10.2 | 5.76 |
| Other REM, female, high, 50+ | 65 | 0.0 | 3.1 | 3.15 |
| Other REM, male, med, 50+ | 103 | 0.0 | 2.6 | 2.62 |
| Other REM, male, high, 50+ | 74 | 1.4 | 3.5 | 2.15 |
| Afr. American, female, high, 50+ | 72 | 1.4 | 3.5 | 2.13 |
| White, female, medium, 30-49 | 3030 | 4.5 | 4.5 | 0.00 |
| White, male, medium, 30-49 | 2615 | 6.1 | 6.1 | 0.04 |

*Observed %, the S-MAIHDA estimate. RE predicted %, the RE-MAIHDA estimate. Attenuation, the absolute difference in percentage points. Other REM, other racial or ethnic minorities, following the original study.*

**S4.6 Interpretation**

This example carries a specific lesson, and it connects to the purpose of the original study. That study set out to reveal high-risk intersections that dominant narratives overlook, and it named young, high-income African-American women as one such group. RE-MAIHDA, applied to the same data, attenuates exactly this group more than any other, and moves its estimate from 16.0% toward the population average. The distributional assumption that smooths the estimates also softens the very signal the analysis was designed to surface.

This is not a defect in the original study, which reported the random-effects predictions as is conventional. It is a property of the estimator, and it is the property the *compare* diagnostic is built to expose.

The practical consequence is visible in the discriminatory-accuracy summary. At a threshold set to the population prevalence, S-MAIHDA flags 28 strata as higher than average and RE-MAIHDA flags 25. The sensitivity of the simple-means classification is 74.2%, against 65.4% for the random-effects classification. The shrinkage that pulls the small high-risk strata toward the average moves some of them below the threshold, so they are no longer flagged. For an exercise whose aim is to identify groups for attention, this matters.

The contrast with the Malmo application in the main text is instructive. In Malmo the strata are large and reasonably uniform in size, so shrinkage acts gently, and the aggregate measures and the rankings are largely preserved; the divergence there is mild and appears only at the level of individual strata. Here the strata differ widely in size and the outcome is uncommon, so shrinkage acts strongly on the small tail, and that tail is where the marginalised, high-risk groups sit. The two applications together show the range of the diagnostic. It reports close agreement when the random-effects assumption is benign, and it reports a clear divergence when the assumption reshapes the result.

This is what the *compare* diagnostic delivers. It does not declare one method correct. It makes the footprint of the distributional assumption visible on the analyst’s own data, stratum by stratum, so that the choice between reporting the observed inequality and reporting the shrunken estimate is made knowingly.

**S4.7 Reproducibility**

The analysis uses the per-stratum data published by Persmark and colleagues, which derive from the public-use file of the 2015 National Survey on Drug Use and Health. The full table of 72 strata is reproduced below as Table S4-D. The same table is also provided as S4 File, a comma-separated file with the number of individuals and the number of cases of each stratum, so that the analysis can be re-run without transcribing the table. The observed percentages are the S-MAIHDA estimates and the predicted absolute risks are the RE-MAIHDA estimates from the empty model. The data are analysed without survey weights, because the purpose is to illustrate a statistical property and not to produce weighted national estimates. The variance partition coefficients are computed on the probability scale by the law of total variance, as in the main text. Confidence intervals for the observed proportions are Wilson intervals. The area under the ROC curve and the sensitivity and specificity at the population prevalence are computed as described in the main text and in S1 Text. One caution for readers replicating from Table S4-D: the classification threshold is the unrounded population prevalence (6.082%). One large stratum (White men, medium income, aged 30 to 49) has a random-effects prediction that rounds to 6.08% but lies just below the threshold. Using the rounded values of the table flips this stratum to the flagged side and yields 26 flagged strata with a sensitivity of 71.5%, instead of the 25 and 65.4% reported in Table S4-A.

**Table S4-D. The 72 intersectional strata: observed prevalence (S-MAIHDA) and predicted absolute risk (RE-MAIHDA), empty model.**

| **Gender** | **Race / ethnicity** | **Income** | **Age** | **N** | **Observed %** | **RE %** |
| --- | --- | --- | --- | --- | --- | --- |
| Male | White | High | 18–29 | 686 | 9.33 | 8.94 |
| Female | White | High | 18–29 | 567 | 6.70 | 6.50 |
| Male | White | High | 30–49 | 821 | 6.09 | 5.99 |
| Female | White | High | 30–49 | 790 | 4.18 | 4.26 |
| Male | White | High | 50+ | 1084 | 1.94 | 2.23 |
| Female | White | High | 50+ | 1113 | 1.44 | 1.81 |
| Male | White | Med | 18–29 | 2061 | 9.22 | 9.07 |
| Female | White | Med | 18–29 | 2206 | 6.84 | 6.78 |
| Male | White | Med | 30–49 | 2615 | 6.12 | 6.08 |
| Female | White | Med | 30–49 | 3030 | 4.49 | 4.49 |
| Male | White | Med | 50+ | 1231 | 2.36 | 2.57 |
| Female | White | Med | 50+ | 1407 | 1.92 | 2.16 |
| Male | White | Low | 18–29 | 1975 | 13.27 | 13.04 |
| Female | White | Low | 18–29 | 2523 | 8.84 | 8.73 |
| Male | White | Low | 30–49 | 953 | 9.86 | 9.56 |
| Female | White | Low | 30–49 | 1422 | 8.16 | 8.01 |
| Male | White | Low | 50+ | 566 | 4.42 | 4.48 |
| Female | White | Low | 50+ | 900 | 2.33 | 2.63 |
| Male | Afr. American | High | 18–29 | 69 | 8.70 | 6.93 |
| Female | Afr. American | High | 18–29 | 50 | 16.00 | 10.24 |
| Male | Afr. American | High | 30–49 | 101 | 1.98 | 3.50 |
| Female | Afr. American | High | 30–49 | 106 | 1.89 | 3.44 |
| Male | Afr. American | High | 50+ | 71 | 2.82 | 4.10 |
| Female | Afr. American | High | 50+ | 72 | 1.39 | 3.52 |
| Male | Afr. American | Med | 18–29 | 355 | 8.45 | 7.88 |
| Female | Afr. American | Med | 18–29 | 395 | 4.81 | 4.79 |
| Male | Afr. American | Med | 30–49 | 303 | 5.94 | 5.75 |
| Female | Afr. American | Med | 30–49 | 378 | 4.23 | 4.37 |
| Male | Afr. American | Med | 50+ | 156 | 2.56 | 3.47 |
| Female | Afr. American | Med | 50+ | 190 | 2.63 | 3.44 |
| Male | Afr. American | Low | 18–29 | 704 | 8.24 | 7.99 |
| Female | Afr. American | Low | 18–29 | 1010 | 4.55 | 4.58 |
| Male | Afr. American | Low | 30–49 | 361 | 6.09 | 5.89 |
| Female | Afr. American | Low | 30–49 | 692 | 3.90 | 4.02 |
| Male | Afr. American | Low | 50+ | 174 | 5.75 | 5.53 |
| Female | Afr. American | Low | 50+ | 290 | 3.79 | 4.06 |
| Male | Hispanic | High | 18–29 | 85 | 7.06 | 6.15 |
| Female | Hispanic | High | 18–29 | 73 | 9.59 | 7.49 |
| Male | Hispanic | High | 30–49 | 98 | 4.08 | 4.55 |
| Female | Hispanic | High | 30–49 | 83 | 4.82 | 4.98 |
| Male | Hispanic | High | 50+ | 63 | 3.17 | 4.33 |
| Female | Hispanic | High | 50+ | 58 | 3.45 | 4.49 |
| Male | Hispanic | Med | 18–29 | 615 | 8.62 | 8.25 |
| Female | Hispanic | Med | 18–29 | 632 | 4.91 | 4.93 |
| Male | Hispanic | Med | 30–49 | 482 | 4.15 | 4.29 |
| Female | Hispanic | Med | 30–49 | 546 | 2.93 | 3.23 |
| Male | Hispanic | Med | 50+ | 134 | 3.73 | 4.27 |
| Female | Hispanic | Med | 50+ | 169 | 2.96 | 3.71 |
| Male | Hispanic | Low | 18–29 | 1094 | 7.68 | 7.52 |
| Female | Hispanic | Low | 18–29 | 1364 | 7.48 | 7.35 |
| Male | Hispanic | Low | 30–49 | 686 | 6.41 | 6.28 |
| Female | Hispanic | Low | 30–49 | 998 | 4.51 | 4.56 |
| Male | Hispanic | Low | 50+ | 180 | 5.56 | 5.39 |
| Female | Hispanic | Low | 50+ | 252 | 3.97 | 4.21 |
| Male | Other REM | High | 18–29 | 88 | 9.09 | 7.33 |
| Female | Other REM | High | 18–29 | 84 | 4.76 | 4.93 |
| Male | Other REM | High | 30–49 | 106 | 2.83 | 3.89 |
| Female | Other REM | High | 30–49 | 115 | 2.61 | 3.73 |
| Male | Other REM | High | 50+ | 74 | 1.35 | 3.50 |
| Female | Other REM | High | 50+ | 65 | 0.00 | 3.15 |
| Male | Other REM | Med | 18–29 | 357 | 6.72 | 6.43 |
| Female | Other REM | Med | 18–29 | 424 | 7.78 | 7.38 |
| Male | Other REM | Med | 30–49 | 402 | 4.23 | 4.38 |
| Female | Other REM | Med | 30–49 | 476 | 4.20 | 4.32 |
| Male | Other REM | Med | 50+ | 103 | 0.00 | 2.62 |
| Female | Other REM | Med | 50+ | 130 | 0.77 | 2.74 |
| Male | Other REM | Low | 18–29 | 548 | 8.21 | 7.84 |
| Female | Other REM | Low | 18–29 | 615 | 6.99 | 6.78 |
| Male | Other REM | Low | 30–49 | 248 | 6.45 | 6.13 |
| Female | Other REM | Low | 30–49 | 314 | 7.01 | 6.61 |
| Male | Other REM | Low | 50+ | 104 | 4.81 | 4.92 |
| Female | Other REM | Low | 50+ | 117 | 1.71 | 3.26 |

*RE %, RE-MAIHDA predicted absolute risk. Other REM, other racial or ethnic minorities, following the original study.*

*Please confirm the complete author list and order against the published article in EndNote before submission.*
