## Supplementary material for "Describing health inequalities without distortion: Simple-Means MAIHDA vs Random-Effects MAIHDA": S5 Text. Python programs

### Simple-Means MAIHDA in Python: a complete, licence-free reproduction

This Supplementary Information reproduces every simple-means measure of the main paper in Python, using only the numpy library. It mirrors the Stata programs of S1 Text and the smaihda command of S1 File (version 1.9.0), and returns the same numbers, including the standardised variance partition (uncorrected and corrected for binomial noise), the contextual decomposition into clustering (CCI), additive differences (G) and modification (CMI) with its noise floor, the threshold classification (sensitivity, specificity, positive predictive value and the TP/FP/FN/TN cell counts), and 95% within-context bootstrap confidence intervals.

The listing below is version 1.9.0 of the script.

We provide it for two reasons. First, a second implementation in an independent language is a direct test of reproducibility: any reader can run it and obtain the published figures from the stratum table (S5 File). Second, Python is free and widely taught, so the material is open to readers without a Stata licence, which suits the didactic aim of this work.

**Scope.** The script covers the simple-means side of the analysis, which is the contribution of the paper and needs no specialised software. The random-effects comparison is fitted with melogit in the Stata command (S1 Text Step 11, S1 File), because a mixed model is best left to a dedicated estimator; the simple-means measures compared against it are the ones reproduced here.

**How to run.** Place the script next to the stratum table dataset-smaihda_stratum_table.csv (S5 File) and run:

python S2_File_smaihda_python.py dataset-smaihda_stratum_table.csv psycmed private

The only dependency is numpy. The script reads the file by name and recognises from its columns whether it holds the stratum table or individual records. The stratum table holds one row per stratum and outcome, with the number of individuals and the number of cases. Individual records are first aggregated to the 300 area-sociodemographic strata, and give the same output. The script prints the measures for both outcomes: the full variance-partition grid (V_A, V_SD and their sum, crude and standardised, uncorrected and corrected), the CCI, the contextual AUC (coAUC) at the A-SD and at the area level, their standardised versions (coAUC*) and the area coAUC*EAR, standardised for size and composition, the threshold classification, and the contextual decomposition (CCI, G, CMI). The printed confidence intervals are the canonical values reported in the paper. The bootstrap uses B = 2,000 replicates with seed 2026, the canonical seed of the paper. Stata and numpy generate different resamples, so the bootstrap confidence intervals of the smaihda command agree with these within Monte Carlo error rather than digit for digit. Optional tokens on the command line change the minimum stratum size of the standardised measures (minn=, default 10), the number of bootstrap replicates (B=) and the seed (seed=).

**Correspondence with S1 Text.** The function per_cell forms the strata, with their number of individuals and of cases, from the individual records or from the stratum table, and orders them by area and stratum (S1 Text Steps 0, 1, 5). decompose computes the weighted and standardised components with the binomial-noise reassignment (S1 Text Steps 6 and 7). coefficients forms the CCI and the VPCs. three_way computes the contextual decomposition CCI / G / CMI with the CMI noise floor (S1 Text Step 7b). deviations gives the deviation of each stratum from its area with the Wilson-shift interval (S1 Text Step 5). auc gives the contextual AUC (coAUC), the discriminatory accuracy, auc_area its area-level version, auc_std and auc_area_std their standardised versions (coAUC*), auc_area_ear the area coAUC*EAR, with every area scored by its composition-standardised prevalence (EAR), and classify the threshold classification (S1 Text Step 8). bootstrap gives the confidence intervals of every measure, the CCI, G and CMI shares included. It holds the contexts fixed and redraws the number of cases of each stratum from a binomial distribution with the observed size and proportion, which is the same as resampling the individuals inside each cell (S1 Text Step 10). bootstrap_cluster, which resamples the 25 areas with replacement, is kept for comparison and does not produce the reported intervals. The Wilson helper gives the interval for a single proportion (S1 Text Step 2).

### The program

```
"""
Simple-Means MAIHDA in Python (v1.9.0, 6 October 2026)
=====================================================================
Reproduces every simple-means measure of the main paper from its stratum
table (S5 File), or from individual records, using only numpy. It mirrors the
R twin (S6 Text) v1.9.0, and the Stata command smaihda v1.9.0 gives the same point
estimates: crude and standardised VPC and CCI (uncorrected
and corrected for binomial noise, with the v1.4.1 boundary rule), AUC at the
stratum and at the area level, threshold classification (sensitivity,
specificity, PPV, TP/FP/FN/TN in individuals), the CCI/G/CMI split, 95%
within-context bootstrap intervals (contexts fixed, individuals resampled).

v1.9.0 (6 October 2026): coAUC*EAR, the area-level contextual AUC
standardised for size and composition. The area-level coAUC* (auc_area_std)
gives every area the same weight but scores it by its crude proportion, which
carries the composition of the area. coAUC*EAR scores every area by its
composition-standardised prevalence, the EAR, the plain mean of the
proportions of its cells with n >= minn, and counts every area once. It is the
coAUC with every cell given the same size, read at the area level, and the
discrimination-scale counterpart of the area VPC*, which is also computed from
the EAR. It has its own within-context bootstrap interval; the random stream is
unchanged, so no other interval moves. The output labels follow the terminology
of the papers: coAUC (crude contextual AUC, discriminatory accuracy), coAUC*
(standardised contextual AUC, general contextual effect) and coAUC*EAR. Nothing
else changes.

v1.8.1 (6 October 2026): noise term of the standardised between-area
component. The area mean of the S_a cells of an area is compared with the grand
mean p*, which is itself the mean of the same K cells, so the expected binomial
noise of V_A* is the sum over areas of (1 - S_a/K)*mean_a(nz)/K, not
sum_a mean_a(nz)/K. The old term ignored that p* is estimated from the same
cells and removed about 1/A of the noise too much (A = number of areas). The
same reasoning gave the factor (1 - 1/S_a) of the within-area component in
v1.5.1, and with one cell per area (S_a = 1) the new term is (1 - 1/K)*mean(nz),
the term of the two-level designs. Verified by binomial simulation in balanced
and unbalanced designs. Only V_A_c and what is computed from it move (corrected
area VPC*, corrected A-SD VPC*, corrected CCI, and the CCI/G/CMI shares); the
identity is unchanged, and no crude or uncorrected measure, AUC or
classification changes.

v1.8.0 (26 September 2026): the engine also reads the stratum table, one
row per stratum and outcome with the columns area, strata_sociodem_id,
strata_global_id, outcome, n and cases; any other column is ignored, and rows
that share a stratum and outcome are summed. Every measure is a function of the
cell counts, so the output is the one obtained from the individual records.
Individual input works as before. The kind of input is recognised from the
columns.

v1.7.0 (25 September 2026): the cells are ordered by area and stratum rather
    than by order of appearance in the file. The bootstrap draws a vector, so the
    position of a cell decided which random number it received, and the interval
    limits therefore depended on how the input rows happened to be sorted. No
    point estimate was ever affected, and in the datasets of the paper, which are
    already sorted by area and stratum, no reported value changes.
v1.6.2 (24 September 2026): the size-standardised AUC now obeys minn, like
every other measure that counts cells rather than individuals. One rule, no
exceptions: a measure that counts cells applies minn, a measure that counts
individuals uses them all. Nothing changes on Malmo (smallest cell 35) or on
the primary ESS design (smallest cell 13); it matters in sparse intersectional
designs, which is where MAIHDA is most used.

v1.6.1 (21 September 2026): the size-standardised AUC, auc_std() and
auc_area_std(), reported as "AUC std by size" with its own within-context
bootstrap interval. Every individual is weighted by 1/n of its own cell, so
every cell contributes the same total weight, exactly as the standardised VPC
counts every cell once. Nothing else changes: no existing VPC, CCI, AUC,
classification or share moves, and the two new measures are additions to the
output, not replacements. See auc_std() for what it is invariant to and for
the caveat that it is read on a hypothetical population of equal-sized cells,
like an age-standardised rate.

v1.6.0 (20 September 2026): the default bootstrap keeps the contexts fixed.
The areas and the strata are the observed set, not a sample of exchangeable
contexts, so the uncertainty of a summary measure is the uncertainty of the
observed proportions themselves. In each replicate the number of events of
every cell is redrawn from Binomial(n, p) of the cell and every measure is
recomputed. A replicate is drawn from proportions that already carry one
dose of binomial noise, so it carries two, and every measure that depends on
the dispersion of the proportions is centred above the estimate by one dose
while its spread is the sampling spread. The reported interval is the
percentile interval recentred on the estimate, truncated to the admissible
range of the measure. Verified on Malmo: with the recentring, every point
estimate lies inside its own interval, and the shares that sit at a boundary
are reported there (CMI of psychotropic medication use 0.0%, 0.0 to 5.0).
The cluster bootstrap that resamples areas remains available as
bootstrap_cluster() for comparison, but it treats the contexts as
exchangeable and is no longer the default.

Earlier, new in v1.5.0:

  - Step 4: deviation of each cell from the crude proportion of its area,
    with a Wilson-shift interval (the cell's Wilson interval moved by the
    area proportion, which is treated as known). The Wald interval of v1.4.x
    had zero width for cells with p = 0 or p = 1.
  - minn (default 10): the size-standardised partition, the standardised
    GCCI and the CCI/G/CMI shares use the cells with n >= minn. Weighted
    measures, AUC and classification always use all cells. minn=1
    reproduces v1.4.1 exactly.
  - A cell-size diagnostic (median and minimum n; cells and individuals
    with n < 5, < 10, < 30; cells with no events and with all events).
  - A flag (!) when a share lies outside its own percentile interval.

v1.5.1 (15 September 2026): noise term of the standardised within-area
component. The within-area deviation of a cell is taken from the mean of
the S_a cells of its own area, so its expected binomial noise is
(1 - 1/S_a)*nz per cell, not (1 - 1/K)*nz. nz_SD is now the mean over
cells of (1 - 1/S_a)*nz (S_a = standardised cells in the area; 12 in
Malmo). Verified by binomial simulation. Effect in Malmo: V_SD_c rises by
about 1 point^2; CCI/G/CMI move by at most 0.5 points (private GP
64.3/18.5/17.2 -> 63.9/19.0/17.1); the identity is unchanged. Nothing else
changes.

Boundary rule (v1.4.1, documented here from v1.6.0): a corrected variance
component is truncated at zero, since the method-of-moments difference
between the observed variance and its expected noise can be negative when
the true component is zero or close to it. A share computed from a truncated
component is reported at the boundary, and an interval that reaches the
boundary is truncated with it.

Input : CSV with columns area, strata_sociodem_id, strata_global_id and one
        0/1 column per outcome (individual records), or the stratum table,
        one row per stratum and outcome with area, strata_sociodem_id,
        strata_global_id, outcome, n and cases (from v1.8.0).
Run   : python S2_File_smaihda_python.py [dataset-smaihda_stratum_table.csv] [outcome ...] [minn=10] [B=2000] [seed=2026]
"""
import csv, math, sys
import numpy as np

CSV = "dataset-smaihda_stratum_table.csv"
OUTCOMES = ["psycmed", "private"]
SEED, B, MINN = 2026, 2000, 10

# ---- command line ----------------------------------------------------
_args = sys.argv[1:]
for a in [x for x in _args if "=" in x]:
    k, v = a.split("=", 1)
    if k == "minn": MINN = int(v)
    elif k == "B": B = int(v)
    elif k == "seed": SEED = int(v)
_pos = [x for x in _args if "=" not in x]
if _pos: CSV = _pos[0]
if len(_pos) > 1: OUTCOMES = _pos[1:]

# ---- intervals -------------------------------------------------------
def wilson(k, n, z=1.96):
    if n == 0: return (float('nan'), float('nan'))
    p = k / n; d = 1 + z*z/n
    c = (p + z*z/(2*n)) / d
    h = z*math.sqrt(p*(1-p)/n + z*z/(4*n*n)) / d
    return (c-h, c+h)

def wilson_vec(ev, n, z=1.96):
    p = ev / n; d = 1 + z*z/n
    c = (p + z*z/(2*n)) / d
    h = z*np.sqrt(p*(1-p)/n + z*z/(4*n*n)) / d
    return c-h, c+h

# ---- data ------------------------------------------------------------
def load(csv_path):
    return list(csv.DictReader(open(csv_path)))

def is_table(rows):
    """v1.8.0: True when the rows are a stratum table, not individual records."""
    return bool(rows) and all(k in rows[0] for k in ("outcome", "n", "cases"))

def _cell_key(r):
    return r.get("strata_global_id") or "%s|%s" % (r["area"], r["strata_sociodem_id"])

def table_n(rows):
    """Individuals behind a stratum table: in each stratum, the largest n over
    the outcomes, since the outcomes differ only by their missing values."""
    tot = {}
    for r in rows:
        k = (_cell_key(r), r["outcome"])
        tot[k] = tot.get(k, 0) + int(float(r["n"]))
    best = {}
    for (k, _o), v in tot.items():
        best[k] = max(best.get(k, 0), v)
    return sum(best.values())

def per_cell(rows, outcome):
    """One row per A-SD cell, ordered by area and stratum: area, sd, n, ev, p."""
    agg = {}
    if is_table(rows):
        # v1.8.0: the stratum table. Rows sharing a stratum and outcome are summed.
        for r in rows:
            if r["outcome"] != outcome: continue
            nn = int(float(r["n"]))
            if nn <= 0: continue
            k = _cell_key(r)
            if k not in agg: agg[k] = [r["area"], r["strata_sociodem_id"], 0, 0]
            agg[k][2] += nn; agg[k][3] += int(float(r["cases"]))
    else:
        for r in rows:
            if r[outcome] in ("", "NA"): continue
            k = r["strata_global_id"]
            if k not in agg: agg[k] = [r["area"], r["strata_sociodem_id"], 0, 0]
            agg[k][2] += 1; agg[k][3] += int(r[outcome])
    # v1.7.0: the cells are ordered by area and stratum, not by order of
    # appearance in the file. The bootstrap draws a vector, so the position of a
    # cell decides which random number it receives, and the interval limits
    # would otherwise depend on how the input rows happen to be sorted. Ordering
    # makes the interval a function of the data alone. No point estimate depends
    # on the order, and in the datasets of the paper, which are already sorted,
    # no reported value changes.
    def _key(k):
        a, d = agg[k][0], agg[k][1]
        try:
            return (0, float(a), float(d), "")
        except (TypeError, ValueError):
            return (1, 0.0, 0.0, "%s|%s" % (a, d))
    order = sorted(agg, key=_key)
    area = np.array([agg[k][0] for k in order]); sd = np.array([agg[k][1] for k in order])
    n = np.array([agg[k][2] for k in order], float); ev = np.array([agg[k][3] for k in order], float)
    return area, sd, n, ev, ev / n

# ---- variance decomposition -------------------------------------------
def decompose(area, n, p, minn=None):
    """Weighted components on all cells; size-standardised components on the
    cells with n >= minn (v1.5.0)."""
    if minn is None: minn = MINN
    N = n.sum(); K_all = len(p)
    areas = np.unique(area)
    p_a_w = {}; n_a = {}
    for a in areas:
        m = area == a
        n_a[a] = n[m].sum(); p_a_w[a] = (n[m]*p[m]).sum()/n_a[a]
    grand = (n*p).sum()/N
    V_A = sum(n_a[a]*(p_a_w[a]-grand)**2 for a in areas)/N
    V_SD = sum(n[i]*(p[i]-p_a_w[area[i]])**2 for i in range(K_all))/N
    V_I = (n*p*(1-p)).sum()/N
    # size-standardised block on n >= minn
    keep = n >= minn
    a_s, n_s, p_s = area[keep], n[keep], p[keep]
    K = len(p_s); areas_s = np.unique(a_s)
    p_a_s = {a: p_s[a_s == a].mean() for a in areas_s}
    pbar_s = p_s.mean()
    V_A_s = sum((p_a_s[a_s[i]]-pbar_s)**2 for i in range(K))/K
    V_SD_s = sum((p_s[i]-p_a_s[a_s[i]])**2 for i in range(K))/K
    V_I_s = (p_s*(1-p_s)).mean()
    nz = p_s*(1-p_s)/n_s
    # v1.5.1: per-cell factor (1 - 1/S_a), S_a = standardised cells in the cell's area
    S_a = {a: int((a_s == a).sum()) for a in areas_s}
    nz_SD = float(np.mean([(1 - 1/S_a[a_s[i]])*nz[i] for i in range(K)]))
    # v1.8.1: (1 - S_a/K), because p* is the mean of the same K cells
    nz_A = sum((1 - S_a[a]/K)*nz[a_s == a].mean() for a in areas_s)/K
    # v1.4.1 boundary rule: corrected components floored at zero
    V_A_c, V_SD_c = max(V_A_s-nz_A, 0.0), max(V_SD_s-nz_SD, 0.0)
    return dict(grand=grand, V_A=V_A, V_SD=V_SD, V_I=V_I,
                pbar_s=pbar_s, V_A_s=V_A_s, V_SD_s=V_SD_s, V_I_s=V_I_s,
                V_A_c=V_A_c, V_SD_c=V_SD_c,
                K_all=K_all, K_std=K, N_excl=float(n[~keep].sum()))

def coefficients(d):
    den = d["grand"]*(1-d["grand"]); dens = d["pbar_s"]*(1-d["pbar_s"])
    out = {}
    out["VPC_A"]   = d["V_A"]/den
    out["VPC_ASD"] = (d["V_A"]+d["V_SD"])/den
    out["GCCI"]    = d["V_A"]/(d["V_A"]+d["V_SD"])
    out["VPC_SD"]  = d["V_SD"]/den
    out["VPC_ASD_s"] = (d["V_A_s"]+d["V_SD_s"])/dens
    out["GCCI_s"]    = d["V_A_s"]/(d["V_A_s"]+d["V_SD_s"])
    out["VPC_ASD_c"] = (d["V_A_c"]+d["V_SD_c"])/dens
    _denc = d["V_A_c"]+d["V_SD_c"]
    out["GCCI_c"]    = d["V_A_c"]/_denc if _denc > 0 else float("nan")
    out["VPC_A_s"]  = d["V_A_s"]/dens
    out["VPC_SD_s"] = d["V_SD_s"]/dens
    out["VPC_A_c"]  = d["V_A_c"]/dens
    out["VPC_SD_c"] = d["V_SD_c"]/dens
    out["ident_w"] = d["V_A"]+d["V_SD"]+d["V_I"]
    out["ident_s"] = d["V_A_s"]+d["V_SD_s"]+d["V_I_s"]
    out["den_w"], out["den_s"] = den, dens
    return out

# ---- Step 4: deviations with Wilson shift (v1.5.0) ---------------------
def deviations(area, n, ev):
    p = ev / n
    areas = np.unique(area)
    pA_d = {a: ev[area == a].sum() / n[area == a].sum() for a in areas}
    pA = np.array([pA_d[a] for a in area])
    lo_w, hi_w = wilson_vec(ev, n)
    dev = p - pA; lo = lo_w - pA; hi = hi_w - pA
    return dev, lo, hi, (lo > 0) | (hi < 0)

# ---- discrimination and classification --------------------------------
def auc(n, p):
    cases = n*p; non = n*(1-p); tc, tn = cases.sum(), non.sum()
    conc = sum(cases[i]*non[p<p[i]].sum() for i in range(len(p)))
    ties = sum(cases[i]*non[p==p[i]].sum() for i in range(len(p)))
    return (conc+0.5*ties)/(tc*tn)

def auc_area(area, n, p):
    areas = np.unique(area)
    n_a = np.array([n[area==a].sum() for a in areas])
    p_a = np.array([(n[area==a]*p[area==a]).sum()/n[area==a].sum() for a in areas])
    return auc(n_a, p_a)

def auc_std(n, p, minn=None):
    """Size-standardised AUC (v1.6.1): every individual is weighted by 1/n of
    its own cell, so every cell contributes the same total weight and the
    result is the AUC that would be observed if all cells were of equal size.

    The ordinary AUC counts pairs of individuals, so a cell of 3000 people
    contributes a million times more pairs than a cell of 3, and the value
    therefore depends on how the population happens to be distributed across
    the cells rather than on the contextual structure alone. Weighting by 1/n
    removes that dependence, in the same way the standardised VPC counts every
    cell once. It is the same kind of object as an age-standardised rate: real
    arithmetic on a hypothetical population in which the cells are of equal
    size, and it is read as such.

    Two properties, both verified by simulation. It is invariant to the size
    distribution: with the contextual effect held fixed, the ordinary AUC falls
    by 4.7 points as the largest cell grows from 1% to 32% of the population
    while this one does not move. And it is nearly invariant to the prevalence
    of the outcome, because a rank measure never sees the base rate: with the
    latent ICC held fixed and the prevalence moved from 5% to 45%, it moves
    0.04 points at an ICC of 1% and 1.44 points at an ICC of 35%, where the
    probability-scale VPC moves five-fold and 1.8-fold respectively (S7 Text).

    It needs no continuity correction. It uses only the ordering of the
    proportions, so a cell with no events has a perfectly well defined rank.

    v1.6.2: the cells with n < minn are excluded, as in every other measure
    that counts cells rather than individuals. Counting a cell once gives a
    cell of three people the weight of a cell of three thousand, and the
    proportion of the first is almost pure noise, so the rule that already
    governs the standardised partition governs this measure too. The crude AUC
    keeps every cell, because it counts individuals and excluding a cell there
    would delete real people from a statement about the population."""
    if minn is None: minn = MINN
    n = np.asarray(n, float); p = np.asarray(p, float)
    keep = n >= minn
    return auc(np.ones(int(keep.sum())), p[keep])

def auc_area_std(area, n, p, minn=None):
    """The area-level AUC with every area counted once.

    The cells with n < minn are excluded first, for the same reason as in
    auc_std. The area proportion is then formed from the surviving cells of
    the area, weighted by their size, and every area contributes the same
    total weight to the AUC."""
    if minn is None: minn = MINN
    n = np.asarray(n, float); p = np.asarray(p, float)
    keep = n >= minn
    a_, n_, p_ = np.asarray(area)[keep], n[keep], p[keep]
    areas = np.unique(a_)
    p_a = np.array([(n_[a_==a]*p_[a_==a]).sum()/n_[a_==a].sum() for a in areas])
    return auc(np.ones(len(areas)), p_a)

def auc_area_ear(area, n, p, minn=None):
    """coAUC*EAR (v1.9.0): every area counted once and scored by its EAR, the
    plain mean of the proportions of its cells with n >= minn. Standardised for
    size and for composition; the counterpart of the area VPC*."""
    if minn is None: minn = MINN
    n = np.asarray(n, float); p = np.asarray(p, float)
    keep = n >= minn
    a_, p_ = np.asarray(area)[keep], p[keep]
    areas = np.unique(a_)
    ear = np.array([p_[a_ == a].mean() for a in areas])
    return auc(np.ones(len(areas)), ear)

def classify(n, p, threshold):
    hr = p >= threshold
    cases = n*p; non = n*(1-p)
    TP = cases[hr].sum();  FP = non[hr].sum()
    FN = cases[~hr].sum(); TN = non[~hr].sum()
    return dict(threshold=threshold, n_flag=int(hr.sum()), TP=TP, FP=FP, FN=FN, TN=TN,
                sens=TP/(TP+FN) if (TP+FN) > 0 else float('nan'),
                spec=TN/(TN+FP) if (TN+FP) > 0 else float('nan'),
                ppv=TP/(TP+FP) if (TP+FP) > 0 else float('nan'))

# ---- contextual split CCI / G / CMI (cells with n >= minn) -------------
def three_way(area, sd, n, p, minn=None):
    if minn is None: minn = MINN
    keep = n >= minn
    area, sd, n, p = area[keep], sd[keep], n[keep], p[keep]
    areas = np.unique(area); A = len(areas); S = len(np.unique(sd)); K = len(p)
    pa = {a: p[area == a].mean() for a in areas}; m = p.mean()
    paA = np.array([pa[a] for a in area])
    V_A_s = np.mean((paA - m) ** 2)
    dA = p - paA
    V_SD_s = np.mean(dA ** 2)
    delt = {s: dA[sd == s].mean() for s in np.unique(sd)}
    gamma = dA - np.array([delt[s] for s in sd])
    V_int = np.mean(gamma ** 2)
    nz = p * (1 - p) / n; vbar = nz.mean()
    S_a = {a: int((area == a).sum()) for a in areas}           # v1.5.1
    nz_SD = float(np.mean([(1 - 1.0 / S_a[area[i]]) * nz[i] for i in range(K)]))
    nz_A = sum((1 - S_a[a]/K) * nz[area == a].mean() for a in areas) / K   # v1.8.1
    V_A_c  = max(V_A_s  - nz_A, 0.0)
    V_SD_c = max(V_SD_s - nz_SD, 0.0)
    V_CMI_c = min(max(V_int - (1 - 1.0/A) * (1 - 1.0/S) * vbar, 0.0), V_SD_c)
    tot = V_A_c + V_SD_c
    if tot <= 0: return float("nan"), float("nan"), float("nan")
    return 100 * V_A_c / tot, 100 * (V_SD_c - V_CMI_c) / tot, 100 * V_CMI_c / tot

# ---- within-context bootstrap (default from v1.6.0) ---------------------
def bootstrap(area, sd, n, p, seed=None, nboot=None):
    """Contexts fixed, individuals resampled inside each cell.

    The areas and the strata are the observed set, not a sample of
    exchangeable contexts, so the only random element is which individuals
    fell into each cell. In each replicate the number of events of a cell is
    redrawn from Binomial(n, p) of the cell, which is equivalent to
    resampling its individuals with replacement.

    A replicate is drawn from the observed proportions, which already carry
    one dose of binomial noise, so it carries two. Every measure that depends
    on the dispersion of the proportions is therefore centred above the
    estimate, by one dose of noise, while its spread is the sampling spread.
    The reported interval is the percentile interval recentred on the
    estimate, and it is then truncated to the admissible range of the measure
    (the boundary rule).
    """
    seed = SEED if seed is None else seed; nboot = B if nboot is None else nboot
    rng = np.random.default_rng(seed)
    keys = ["VPC_A","VPC_SD","VPC_ASD","GCCI","VPC_A_s","VPC_SD_s","VPC_ASD_s","GCCI_s",
            "VPC_A_c","VPC_SD_c","VPC_ASD_c","GCCI_c","AUC","AUC_area",
            "AUC_s","AUC_area_s","CCI","G","CMI","AUC_area_ear"]
    store = {k: [] for k in keys}
    ni = np.rint(n).astype(int)
    for _ in range(nboot):
        pp = rng.binomial(ni, p) / ni
        c = coefficients(decompose(area, n, pp))
        cci, g, cmi = three_way(area, sd, n, pp)
        for k in keys[:12]: store[k].append(c[k])
        store["AUC"].append(auc(n, pp)); store["AUC_area"].append(auc_area(area, n, pp))
        store["AUC_s"].append(auc_std(n, pp))
        store["AUC_area_s"].append(auc_area_std(area, n, pp))
        store["CCI"].append(cci); store["G"].append(g); store["CMI"].append(cmi)
        store["AUC_area_ear"].append(auc_area_ear(area, n, pp))
    # point estimates on the observed data, for the recentring
    c0 = coefficients(decompose(area, n, p))
    cci0, g0, cmi0 = three_way(area, sd, n, p)
    est = {k: c0[k] for k in keys[:12]}
    est["AUC"] = auc(n, p); est["AUC_area"] = auc_area(area, n, p)
    est["AUC_s"] = auc_std(n, p); est["AUC_area_s"] = auc_area_std(area, n, p)
    est["CCI"], est["G"], est["CMI"] = cci0, g0, cmi0
    est["AUC_area_ear"] = auc_area_ear(area, n, p)
    # shares and the CCI are reported in percent; the rest as proportions
    hi_bound = {k: (100.0 if k in ("CCI","G","CMI") else 1.0) for k in keys}
    out = {}
    for k in keys:
        rep = np.asarray(store[k], float)
        m = np.nanmean(rep)
        if np.isnan(m) or np.isnan(est[k]):
            out[k] = (float("nan"), float("nan")); continue
        lo = est[k] + (np.nanpercentile(rep, 2.5) - m)
        hi = est[k] + (np.nanpercentile(rep, 97.5) - m)
        # boundary rule: an interval that reaches the boundary is truncated with it
        out[k] = (max(lo, 0.0), min(hi, hi_bound[k]))
    return out

# ---- cluster bootstrap (kept for comparison, not the default) -----------
def bootstrap_cluster(area, sd, n, p, seed=None, nboot=None):
    """Areas resampled with replacement. This treats the contexts as
    exchangeable draws from a wider set of contexts, which is the assumption
    Simple-Means MAIHDA declines, so it is no longer the default. It answers
    a different question, the variability expected over other sets of areas
    like these."""
    seed = SEED if seed is None else seed; nboot = B if nboot is None else nboot
    rng = np.random.default_rng(seed)
    areas = np.unique(area)
    keys = ["VPC_A","VPC_SD","VPC_ASD","GCCI","VPC_A_s","VPC_SD_s","VPC_ASD_s","GCCI_s",
            "VPC_A_c","VPC_SD_c","VPC_ASD_c","GCCI_c","AUC","AUC_area",
            "AUC_s","AUC_area_s","CCI","G","CMI","AUC_area_ear"]
    store = {k: [] for k in keys}
    for _ in range(nboot):
        samp = rng.choice(areas, size=len(areas), replace=True)
        idx = np.concatenate([np.where(area==a)[0] for a in samp])
        newarea = np.concatenate([[f"{a}_{j}"]*np.sum(area==a) for j, a in enumerate(samp)])
        aa, nn, pp, ss = newarea, n[idx], p[idx], sd[idx]
        c = coefficients(decompose(aa, nn, pp))
        cci, g, cmi = three_way(aa, ss, nn, pp)
        for k in keys[:12]: store[k].append(c[k])
        store["AUC"].append(auc(nn, pp)); store["AUC_area"].append(auc_area(aa, nn, pp))
        store["AUC_s"].append(auc_std(nn, pp))
        store["AUC_area_s"].append(auc_area_std(aa, nn, pp))
        store["CCI"].append(cci); store["G"].append(g); store["CMI"].append(cmi)
        store["AUC_area_ear"].append(auc_area_ear(aa, nn, pp))
    return {k: (np.nanpercentile(store[k], 2.5), np.nanpercentile(store[k], 97.5)) for k in keys}

# ---- main ---------------------------------------------------------------
def main():
    rows = load(CSV)
    print(f"smaihda Python v1.9.0 | minn = {MINN} | B = {B} | seed = {SEED} | "
          f"within-context bootstrap (contexts fixed)")
    print(f"STEP 1  Data verification: N={table_n(rows) if is_table(rows) else len(rows)}  areas={len({r['area'] for r in rows})}  "
          f"strata={len({r['strata_global_id'] for r in rows})}")
    for out in OUTCOMES:
        area, sd, n, ev, p = per_cell(rows, out)
        d = decompose(area, n, p); c = coefficients(d)
        A = auc(n, p); Aa = auc_area(area, n, p)
        As = auc_std(n, p); Aas = auc_area_std(area, n, p); Aae = auc_area_ear(area, n, p)
        ci = bootstrap(area, sd, n, p)
        print(f"\n===== {out}  (prevalence {d['grand']*100:.1f}%) =====")
        # cell-size diagnostic
        N = n.sum(); K = len(n); grid = len(np.unique(area)) * len(np.unique(sd))
        def pct(k): return f"{int(k.sum())} cells ({100*k.mean():.1f}%), {n[k].sum():.0f} individuals ({100*n[k].sum()/N:.1f}%)"
        print(f"  cell-size diagnostic: {K} occupied of {grid} area x stratum cells ({grid-K} empty); "
              f"median n = {np.median(n):.0f}, min n = {n.min():.0f}")
        print(f"    n < 5   : {pct(n < 5)}")
        print(f"    n < 10  : {pct(n < 10)}")
        print(f"    n < 30  : {pct(n < 30)}")
        print(f"    no events (p = 0)  : {pct(ev == 0)}")
        print(f"    all events (p = 1) : {pct(ev == n)}")
        print(f"  identity weighted : {c['ident_w']:.8f} vs {c['den_w']:.8f}")
        print(f"  identity standard.: {c['ident_s']:.8f} vs {c['den_s']:.8f}")
        dev, lo, hi, sig = deviations(area, n, ev)
        print(f"  STEP 4 deviations (Wilson shift): {int(sig.sum())} of {len(sig)} significant; "
              f"range {dev.min():.3f} to {dev.max():.3f}")
        if d["K_std"] < d["K_all"]:
            print(f"  standardised measures use {d['K_std']} of {d['K_all']} cells (n >= {MINN}); "
                  f"{d['K_all']-d['K_std']} cells and {d['N_excl']:.0f} individuals excluded")
        def line(name, val, key):
            lo_, hi_ = ci[key]; print(f"  {name:26} {val*100:6.2f}%  ({lo_*100:.2f} to {hi_*100:.2f})")
        line("VPC_A, crude",            c["VPC_A"],     "VPC_A")
        line("VPC_SD, crude",           c["VPC_SD"],    "VPC_SD")
        line("VPC/ICC_A+SD, crude",     c["VPC_ASD"],   "VPC_ASD")
        line("CCI, crude",             c["GCCI"],      "GCCI")
        line("VPC_A std, uncorrected",      c["VPC_A_s"],   "VPC_A_s")
        line("VPC_A std, corrected",        c["VPC_A_c"],   "VPC_A_c")
        line("VPC_SD std, uncorrected",     c["VPC_SD_s"],  "VPC_SD_s")
        line("VPC_SD std, corrected",       c["VPC_SD_c"],  "VPC_SD_c")
        line("VPC/ICC_A+SD std, uncorr",    c["VPC_ASD_s"], "VPC_ASD_s")
        line("VPC/ICC_A+SD std, corrected", c["VPC_ASD_c"], "VPC_ASD_c")
        line("CCI, std, uncorrected",       c["GCCI_s"],    "GCCI_s")
        line("CCI, std, corrected",         c["GCCI_c"],    "GCCI_c")
        line("coAUC (A-SD strata)",         A,              "AUC")
        line("coAUC (area level)",          Aa,             "AUC_area")
        line("coAUC* (A-SD strata)",        As,             "AUC_s")
        line("coAUC* (area, size)",         Aas,            "AUC_area_s")
        line("coAUC*EAR (area)",            Aae,            "AUC_area_ear")
        cl = classify(n, p, threshold=d["grand"])
        print(f"  threshold (pop. prevalence) = {cl['threshold']*100:.2f}%")
        print(f"  strata flagged high-risk    = {cl['n_flag']}")
        print(f"  sensitivity / specificity   = {cl['sens']*100:.1f}% / {cl['spec']*100:.1f}%")
        print(f"  positive predictive value   = {cl['ppv']*100:.1f}%")
        print(f"  TP / FP / FN / TN (indiv.)  = {cl['TP']:.0f} / {cl['FP']:.0f} / {cl['FN']:.0f} / {cl['TN']:.0f}")
        cci, g, cmi = three_way(area, sd, n, p)
        print("  contextual decomposition (standardised; CMI noise-corrected):")
        flags = []
        for name, val, key in (("CCI  clustering   ", cci, "CCI"), ("G    additive     ", g, "G"), ("CMI  modification ", cmi, "CMI")):
            lo_, hi_ = ci[key]
            fl = "  (!)" if (not math.isnan(val)) and (val < lo_ or val > hi_) else ""
            if fl: flags.append(key)
            print(f"    {name} = {val:5.1f}%  ({lo_:.1f} to {hi_:.1f}){fl}")
        if flags:
            print("    (!) a point estimate lies outside its own percentile interval")

if __name__ == "__main__":
    main()
```
