## Supplementary material for "Describing health inequalities without distortion: Simple-Means MAIHDA vs Random-Effects MAIHDA": S6 Text. R programs

**Simple-Means MAIHDA in R: an independent reproduction in base R**

This Supplementary Information reproduces every simple-means measure of the main paper in R, using base R only, with no packages. It is a twin of the Python script of S5 Text and of the smaihda command of S1 File (version 1.9.0): the same aggregation to the 300 A-SD strata, the same crude and standardised variance partition (uncorrected and corrected for binomial noise, with the boundary rule), the same contextual decomposition into CCI, G and CMI, the same contextual AUC (coAUC) at the stratum and at the area level, with its standardised versions (coAUC* and the area coAUC*EAR), the same threshold classification in individuals, the deviation of each stratum from its area with the Wilson-shift interval (Step 4), the cell-size diagnostic, and 95% within-context bootstrap intervals, the contexts held fixed and the individuals resampled inside each cell.

Every point estimate agrees with the Python script (S5 Text) and with the smaihda command (S1 File) at every printed figure. The bootstrap intervals agree within Monte Carlo error, since R, numpy and Stata draw different random sequences.

**Input and use.** The script reads the stratum table (dataset-smaihda_stratum_table.csv, S5 File), one row per stratum and outcome with the number of individuals and the number of cases, or individual records, and recognises the kind of input from the columns. It takes on the command line the file, the outcome names, and the optional tokens minn=, B= and seed=. Typing Rscript S3_File_smaihda_R.R dataset-smaihda_stratum_table.csv psycmed private reproduces the analysis of the paper, and individual records in place of the table give the same output. With minn=1 every stratum enters the standardised measures.

Rscript S3_File_smaihda_R.R dataset-smaihda_stratum_table.csv psycmed private

**Listing of S3_File_smaihda_R.R (version 1.9.0)**

\# Simple-Means MAIHDA in R (v1.9.0, 6 October 2026)
\# ==================================================================
\# Twin of the Python implementation (S5 Text). Base R only. Reproduces every
\# simple-means measure of the paper from its stratum table (S5 File) or from
\# individual records: crude and standardised VPC and CCI (uncorrected and corrected
\# for binomial noise, with the v1.4.1 boundary rule), AUC at the stratum and at
\# the area level, threshold classification (sensitivity, specificity, PPV,
\# TP/FP/FN/TN in individuals), the CCI/G/CMI split, all with 95% within-context
\# bootstrap confidence intervals (contexts fixed, individuals resampled), and,
\# new in v1.5.0, Step 4 (deviation of each cell from its area with a
\# Wilson-shift interval) and the minn rule for the size-standardised measures.
\# The Stata command smaihda v1.9.0 gives the same point estimates.
\#
\# v1.9.0 (6 October 2026): coAUC*EAR, the area-level contextual AUC
\# standardised for size and composition. The area-level coAUC* (auc_area_std)
\# gives every area the same weight but scores it by its crude proportion, which
\# carries the composition of the area. coAUC*EAR scores every area by its
\# composition-standardised prevalence, the EAR, the plain mean of the
\# proportions of its cells with n >= minn, and counts every area once. It is the
\# coAUC with every cell given the same size, read at the area level, and the
\# discrimination-scale counterpart of the area VPC*, which is also computed from
\# the EAR. It has its own within-context bootstrap interval; the random stream is
\# unchanged, so no other interval moves. The output labels follow the terminology
\# of the papers: coAUC (crude contextual AUC, discriminatory accuracy), coAUC*
\# (standardised contextual AUC, general contextual effect) and coAUC*EAR. Nothing
\# else changes.
\#
\# v1.8.1 (6 October 2026): noise term of the standardised between-area
\# component. The area mean of the S_a cells of an area is compared with the grand
\# mean p*, which is itself the mean of the same K cells, so the expected binomial
\# noise of V_A* is the sum over areas of (1 - S_a/K)*mean_a(nz)/K, not
\# sum_a mean_a(nz)/K. The old term ignored that p* is estimated from the same
\# cells and removed about 1/A of the noise too much (A = number of areas). The
\# same reasoning gave the factor (1 - 1/S_a) of the within-area component in
\# v1.5.1, and with one cell per area (S_a = 1) the new term is (1 - 1/K)*mean(nz),
\# the term of the two-level designs. Verified by binomial simulation in balanced
\# and unbalanced designs. Only V_A_c and what is computed from it move (corrected
\# area VPC*, corrected A-SD VPC*, corrected CCI, and the CCI/G/CMI shares); the
\# identity is unchanged, and no crude or uncorrected measure, AUC or
\# classification changes.
\#
\# v1.8.0 (26 September 2026): the engine also reads the stratum table, one row
\#	per stratum and outcome with the columns area, strata_sociodem_id,
\#	strata_global_id, outcome, n and cases; other columns are ignored and rows
\#	that share a stratum and outcome are summed. Every measure is a function of
\#	the cell counts, so the output is the one obtained from the individual
\#	records. The kind of input is recognised from the columns.
\# v1.7.0: the cells are ordered by area and stratum rather than by order of
\#	appearance in the file, so the bootstrap interval is a function of the
\#	data alone and not of how the input rows happen to be sorted. No point
\#	estimate is affected.
\# v1.6.2: auc_std and auc_area_std obey minn, like every other measure that
\#	counts cells rather than individuals. v1.6.1 added them.
\#
\# v1.6.0 (20 September 2026): the default bootstrap keeps the contexts fixed.
\#	The areas and the strata are the observed set, not a sample of
\#	exchangeable contexts, so the uncertainty of a summary measure is the
\#	uncertainty of the observed proportions themselves. In each replicate the
\#	number of events of every cell is redrawn from Binomial(n, p) of the
\#	cell. A replicate carries two doses of binomial noise against one in the
\#	observed proportions, so every dispersion-based measure is centred above
\#	the estimate by one dose; the interval is the percentile interval
\#	recentred on the estimate and truncated to the admissible range of the
\#	measure. The cluster bootstrap remains available as bootstrap_cluster().
\#	Mirrors the Python twin v1.8.0 and smaihda.ado v1.8.0, which also draws
\#	the events of each cell from Binomial(n, p), in the same order of cells.
\#
\# v1.5.1 (15 September 2026): noise term of the standardised within-area
\#	component. The within-area deviation of a cell is taken from the mean of
\#	the S_a cells of its own area, so its expected binomial noise is
\#	(1 - 1/S_a)*nz per cell, not (1 - 1/K)*nz. nz_SD is now the mean over
\#	cells of (1 - 1/S_a)*nz. Verified by binomial simulation. Effect in Malmo:
\#	CCI/G/CMI move by at most 0.5 points; identity unchanged. Mirrors
\#	smaihda.ado v1.5.2 and the Python twin v1.5.1.
\#
\# v1.5.0 changes:
\#	- Step 4 deviation interval = the cell's Wilson interval shifted by the
\#	crude area proportion, which is treated as known. The Wald interval of
\#	v1.4.x had zero width for cells with p = 0 or 1.
\#	- minn (default 10): the size-standardised partition, the standardised
\#	GCCI and the CCI/G/CMI shares use the cells with n >= minn. Weighted
\#	measures, AUC and classification always use all cells. minn=1
\#	reproduces v1.4.1 exactly.
\#
\# Point estimates must match the Python implementation to 8 decimals. Bootstrap intervals match
\# approximately (different random streams).
\#
\# Input : CSV with columns area, strata_sociodem_id, strata_global_id and
\#	one 0/1 column per outcome (rows with a missing outcome are dropped),
\#	or the stratum table, one row per stratum and outcome with area,
\#	strata_sociodem_id, strata_global_id, outcome, n and cases (from v1.8.0).
\# Run	: Rscript S3_File_smaihda_R.R dataset-smaihda_stratum_table.csv psycmed private [minn=10] [B=2000] [seed=2026]
\#	(arguments: file, outcome names, optional minn=, B= and seed= tokens)

args <- commandArgs(trailingOnly = TRUE)
opt <- args[grepl("^(minn|B|seed)=", args)]
args <- args[!grepl("^(minn|B|seed)=", args)]
CSV	<- if (length(args) >= 1) args[1] else "dataset-smaihda_stratum_table.csv"
OUTCOMES <- if (length(args) >= 2) args[-1] else c("psycmed", "private")
SEED <- 2026; B <- 2000; MINN <- 10
for (o in opt) {
 kv <- strsplit(o, "=")[[1]]
 if (kv[1] == "minn") MINN <- as.integer(kv[2])
 if (kv[1] == "B")	B	<- as.integer(kv[2])
 if (kv[1] == "seed") SEED <- as.integer(kv[2])
}
if (nzchar(Sys.getenv("SMAIHDA_B"))) B <- as.integer(Sys.getenv("SMAIHDA_B"))

\# ---- intervals ----------------------------------------------------------
wilson <- function(x, n, z = 1.96) {
 p <- x / n
 d <- 1 + z * z / n
 c0 <- (p + z * z / (2 * n)) / d
 h <- z * sqrt(p * (1 - p) / n + z * z / (4 * n * n)) / d
 list(lo = c0 - h, hi = c0 + h)
}

\# ---- aggregation --------------------------------------------------------
is_table <- function(d) all(c("outcome", "n", "cases") %in% names(d))

cell_key <- function(d) {
 if ("strata_global_id" %in% names(d)) as.character(d$strata_global_id)
 else paste(d$area, d$strata_sociodem_id, sep = "|")
}

table_n <- function(d) {
 \# individuals behind the table: per stratum, the largest n over the outcomes
 s <- tapply(as.numeric(d$n), list(cell_key(d), d$outcome), sum)
 sum(apply(s, 1, max, na.rm = TRUE))
}

per_cell <- function(d, outcome) {
 if (is_table(d)) {
 \# v1.8.0: the stratum table; rows sharing a stratum and outcome are summed
 t <- d[d$outcome == outcome & d$n > 0, ]
 key <- cell_key(t)
 first <- !duplicated(key)
 ids <- key[first]
 ka <- suppressWarnings(as.numeric(t$area[first]))
 kd <- suppressWarnings(as.numeric(t$strata_sociodem_id[first]))
 if (any(is.na(ka)) || any(is.na(kd))) {
 o <- order(as.character(t$area[first]), as.character(t$strata_sociodem_id[first]))
 } else {
 o <- order(ka, kd)
 }
 ids <- ids[o]
 m <- match(key, ids)
 n <- as.numeric(rowsum(as.numeric(t$n), m)[, 1])
 ev <- as.numeric(rowsum(as.numeric(t$cases), m)[, 1])
 return(list(area = as.character(t$area[first])[o],
 sd	= as.character(t$strata_sociodem_id[first])[o],
 n = n, ev = ev, p = ev / n))
 }
 \# v1.7.0: one row per A-SD cell, ordered by area and stratum, not by order of
 \# appearance in the file. The bootstrap draws a vector, so the position of a
 \# cell decides which random number it receives; ordering makes the interval a
 \# function of the data alone. Same key as the Python twin.
 d <- d[!is.na(d[[outcome]]), ]
 key <- as.character(d$strata_global_id)
 first <- !duplicated(key)
 ids <- key[first]
 ka <- suppressWarnings(as.numeric(d$area[first]))
 kd <- suppressWarnings(as.numeric(d$strata_sociodem_id[first]))
 if (any(is.na(ka)) || any(is.na(kd))) {
 o <- order(as.character(d$area[first]), as.character(d$strata_sociodem_id[first]))
 } else {
 o <- order(ka, kd)
 }
 ids <- ids[o]
 m <- match(key, ids)
 n <- as.numeric(tabulate(m, nbins = length(ids)))
 ev <- as.numeric(rowsum(as.numeric(d[[outcome]]), m)[, 1])
 list(area = as.character(d$area[first])[o],
 sd	= as.character(d$strata_sociodem_id[first])[o],
 n = n, ev = ev, p = ev / n)
}

\# ---- variance decomposition (weighted on all cells; standardised on n >= minn)
decompose <- function(area, n, p, minn = MINN) {
 N <- sum(n)
 areas <- unique(area)
 n_a	<- tapply(n, area, sum)[areas]
 p_a_w <- (tapply(n * p, area, sum) / tapply(n, area, sum))[areas]
 grand <- sum(n * p) / N
 V_A <- sum(n_a * (p_a_w - grand)^2) / N
 V_SD <- sum(n * (p - p_a_w[area])^2) / N
 V_I <- sum(n * p * (1 - p)) / N
 \# v1.5.0: size-standardised block on the cells with n >= minn
 k <- n >= minn
 K_all <- length(p); K <- sum(k)
 as <- area[k]; ns <- n[k]; ps <- p[k]
 areas_s <- unique(as)
 p_a_s <- tapply(ps, as, mean)[areas_s]
 pbar_s <- mean(ps)
 V_A_s <- sum((p_a_s[as] - pbar_s)^2) / K
 V_SD_s <- sum((ps - p_a_s[as])^2) / K
 V_I_s <- mean(ps * (1 - ps))
 nz <- ps * (1 - ps) / ns
 S_a	<- as.numeric(table(as)[as.character(as)])	# v1.5.1: cells per area
 nz_SD <- mean((1 - 1 / S_a) * nz)
 \# v1.8.1: (1 - S_a/K), because p* is the mean of the same K cells
 S_ar <- tapply(ps, as, length)
 nz_A <- sum((1 - S_ar / K) * tapply(nz, as, mean)) / K
 \# v1.4.1 boundary rule: corrected components floored at zero
 V_A_c <- max(V_A_s - nz_A, 0)
 V_SD_c <- max(V_SD_s - nz_SD, 0)
 list(grand = grand, V_A = V_A, V_SD = V_SD, V_I = V_I,
 pbar_s = pbar_s, V_A_s = V_A_s, V_SD_s = V_SD_s, V_I_s = V_I_s,
 V_A_c = V_A_c, V_SD_c = V_SD_c,
 K_all = K_all, K_std = K, N_excl = sum(n[!k]))
}

coefficients_sm <- function(d) {
 den <- d$grand * (1 - d$grand)
 dens <- d$pbar_s * (1 - d$pbar_s)
 denc <- d$V_A_c + d$V_SD_c
 list(
 VPC_A = d$V_A / den, VPC_SD = d$V_SD / den,
 VPC_ASD = (d$V_A + d$V_SD) / den, GCCI = d$V_A / (d$V_A + d$V_SD),
 VPC_A_s = d$V_A_s / dens, VPC_SD_s = d$V_SD_s / dens,
 VPC_ASD_s = (d$V_A_s + d$V_SD_s) / dens, GCCI_s = d$V_A_s / (d$V_A_s + d$V_SD_s),
 VPC_A_c = d$V_A_c / dens, VPC_SD_c = d$V_SD_c / dens,
 VPC_ASD_c = (d$V_A_c + d$V_SD_c) / dens,
 \# v1.4.1 boundary rule: GCCI_c undefined when no clustering beyond noise
 GCCI_c = if (denc > 0) d$V_A_c / denc else NA_real_,
 ident_w = d$V_A + d$V_SD + d$V_I, ident_s = d$V_A_s + d$V_SD_s + d$V_I_s,
 den_w = den, den_s = dens)
}

\# ---- Step 4: deviation from the crude area proportion, Wilson shift -----
deviations_sm <- function(area, n, ev) {
 p <- ev / n
 n_a <- tapply(n, area, sum); ev_a <- tapply(ev, area, sum)
 pA <- as.numeric((ev_a / n_a)[area])
 w <- wilson(ev, n)
 dev <- p - pA; lo <- w$lo - pA; hi <- w$hi - pA
 list(dev = dev, lo = lo, hi = hi, sig = (lo > 0) | (hi < 0))
}

\# ---- discrimination -----------------------------------------------------
auc_sm <- function(n, p) {
 cases <- n * p; non <- n * (1 - p)
 tc <- sum(cases); tn <- sum(non)
 conc <- 0; ties <- 0
 for (i in seq_along(p)) {
 conc <- conc + cases[i] * sum(non[p < p[i]])
 ties <- ties + cases[i] * sum(non[p == p[i]])
 }
 (conc + 0.5 * ties) / (tc * tn)
}

auc_area <- function(area, n, p) {
 areas <- unique(area)
 n_a <- as.numeric(tapply(n, area, sum)[areas])
 p_a <- as.numeric((tapply(n * p, area, sum) / tapply(n, area, sum))[areas])
 auc_sm(n_a, p_a)
}

\# Size-standardised AUC (v1.6.1). Every individual is weighted by 1/n of its
\# own cell, so every cell contributes the same total weight and the result is
\# the AUC that would be observed if all cells were of equal size, exactly as
\# the standardised VPC counts every cell once. It is the same kind of object
\# as an age-standardised rate: real arithmetic on a hypothetical population.
\# It needs no continuity correction, since it uses only the ordering of the
\# proportions and a cell with no events has a well defined rank.
\# v1.6.2: the cells with n < minn are excluded, as in every other measure that
\# counts cells rather than individuals. One rule, no exceptions: a measure that
\# counts cells applies minn, a measure that counts individuals uses them all.
auc_std <- function(n, p, minn = MINN) {
 keep <- n >= minn
 auc_sm(rep(1, sum(keep)), p[keep])
}

auc_area_std <- function(area, n, p, minn = MINN) {
 keep <- n >= minn
 a <- area[keep]; nn <- n[keep]; pp <- p[keep]
 areas <- unique(a)
 p_a <- as.numeric((tapply(nn * pp, a, sum) / tapply(nn, a, sum))[areas])
 auc_sm(rep(1, length(p_a)), p_a)
}

auc_area_ear <- function(area, n, p, minn = MINN) {
 \# v1.9.0: coAUC*EAR, every area counted once and scored by its EAR, the plain
 \# mean of the proportions of its cells with n >= minn
 keep <- n >= minn
 ear <- tapply(p[keep], area[keep], mean)
 auc_sm(rep(1, length(ear)), as.numeric(ear))
}

classify_sm <- function(n, p, threshold) {
 hr <- p >= threshold
 cases <- n * p; non <- n * (1 - p)
 TP <- sum(cases[hr]); FP <- sum(non[hr])
 FN <- sum(cases[!hr]); TN <- sum(non[!hr])
 list(threshold = threshold, n_flag = sum(hr), TP = TP, FP = FP, FN = FN, TN = TN,
 sens = if (TP + FN > 0) TP / (TP + FN) else NA,
 spec = if (TN + FP > 0) TN / (TN + FP) else NA,
 ppv = if (TP + FP > 0) TP / (TP + FP) else NA)
}

\# ---- contextual split CCI / G / CMI (cells with n >= minn) --------------
three_way <- function(area, sd, n, p, minn = MINN) {
 k <- n >= minn
 area <- area[k]; sd <- sd[k]; n <- n[k]; p <- p[k]
 areas <- unique(area); A <- length(areas); S <- length(unique(sd)); K <- length(p)
 pa <- tapply(p, area, mean); m <- mean(p)
 paA <- as.numeric(pa[area])
 V_A_s <- mean((paA - m)^2)
 dA <- p - paA
 V_SD_s <- mean(dA^2)
 delt <- tapply(dA, sd, mean)
 gamma <- dA - as.numeric(delt[sd])
 V_int <- mean(gamma^2)
 nz <- p * (1 - p) / n; vbar <- mean(nz)
 S_a	<- as.numeric(table(area)[as.character(area)])	# v1.5.1
 nz_SD <- mean((1 - 1 / S_a) * nz)
 nz_A <- sum((1 - tapply(p, area, length) / K) * tapply(nz, area, mean)) / K	# v1.8.1
 V_A_c	<- max(V_A_s - nz_A, 0)
 V_SD_c <- max(V_SD_s - nz_SD, 0)
 V_CMI_c <- min(max(V_int - (1 - 1 / A) * (1 - 1 / S) * vbar, 0), V_SD_c)
 tot <- V_A_c + V_SD_c
 if (tot > 0) c(CCI = 100 * V_A_c / tot, G = 100 * (V_SD_c - V_CMI_c) / tot, CMI = 100 * V_CMI_c / tot)
 else c(CCI = NA_real_, G = NA_real_, CMI = NA_real_)
}

BOOT_KEYS <- c("VPC_A","VPC_SD","VPC_ASD","GCCI","VPC_A_s","VPC_SD_s","VPC_ASD_s","GCCI_s",
 "VPC_A_c","VPC_SD_c","VPC_ASD_c","GCCI_c","AUC","AUC_area",
 "AUC_s","AUC_area_s","CCI","G","CMI","AUC_area_ear")

\# ---- within-context bootstrap (default from v1.6.0) ---------------------
\# The areas and the strata are the observed set, not a sample of exchangeable
\# contexts, so the only random element is which individuals fell into each
\# cell. In each replicate the number of events of a cell is redrawn from
\# Binomial(n, p) of the cell. A replicate is drawn from proportions that
\# already carry one dose of binomial noise, so it carries two, and every
\# measure that depends on the dispersion of the proportions is centred above
\# the estimate by one dose while its spread is the sampling spread. The
\# interval is the percentile interval recentred on the estimate, truncated to
\# the admissible range of the measure (the boundary rule).
bootstrap_sm <- function(area, sd, n, p, seed = SEED, nboot = B) {
 set.seed(seed)
 keys <- BOOT_KEYS
 store <- matrix(NA_real_, nboot, length(keys), dimnames = list(NULL, keys))
 ni <- as.integer(round(n))
 for (b in seq_len(nboot)) {
 pp <- rbinom(length(ni), ni, p) / ni
 cf <- coefficients_sm(decompose(area, n, pp))
 tw <- three_way(area, sd, n, pp)
 store[b, ] <- c(unlist(cf[keys[1:12]]), auc_sm(n, pp), auc_area(area, n, pp),
 auc_std(n, pp), auc_area_std(area, n, pp), tw, auc_area_ear(area, n, pp))
 }
 \# point estimates on the observed data, for the recentring
 cf0 <- coefficients_sm(decompose(area, n, p))
 est <- c(unlist(cf0[keys[1:12]]), auc_sm(n, p), auc_area(area, n, p),
 auc_std(n, p), auc_area_std(area, n, p),
 three_way(area, sd, n, p), auc_area_ear(area, n, p))
 names(est) <- keys
 qs <- apply(store, 2, quantile, probs = c(0.025, 0.975), na.rm = TRUE, type = 7)
 mn <- apply(store, 2, mean, na.rm = TRUE)
 out <- qs
 for (k in keys) {
 top <- if (k %in% c("CCI", "G", "CMI")) 100 else 1
 if (is.na(est[[k]]) || is.na(mn[[k]])) { out[, k] <- NA_real_; next }
 lo <- est[[k]] + (qs[1, k] - mn[[k]])
 hi <- est[[k]] + (qs[2, k] - mn[[k]])
 out[, k] <- c(max(lo, 0), min(hi, top))
 }
 out
}

\# ---- cluster bootstrap (kept for comparison, not the default) -----------
\# Areas resampled with replacement, duplicates relabelled. This treats the
\# contexts as exchangeable draws from a wider set of contexts, which is the
\# assumption Simple-Means MAIHDA declines.
resample_areas <- function(area) {
 areas <- unique(area)
 samp <- sample(areas, length(areas), replace = TRUE)
 idx <- unlist(lapply(samp, function(a) which(area == a)))
 newarea <- unlist(lapply(seq_along(samp),
 function(j) rep(paste0(samp[j], "_", j), sum(area == samp[j]))))
 list(idx = idx, area = newarea)
}

bootstrap_cluster <- function(area, sd, n, p, seed = SEED, nboot = B) {
 set.seed(seed)
 keys <- BOOT_KEYS
 store <- matrix(NA_real_, nboot, length(keys), dimnames = list(NULL, keys))
 for (b in seq_len(nboot)) {
 r <- resample_areas(area)
 aa <- r$area; nn <- n[r$idx]; pp <- p[r$idx]; ss <- sd[r$idx]
 cf <- coefficients_sm(decompose(aa, nn, pp))
 tw <- three_way(aa, ss, nn, pp)
 store[b, ] <- c(unlist(cf[keys[1:12]]), auc_sm(nn, pp), auc_area(aa, nn, pp),
 auc_std(nn, pp), auc_area_std(aa, nn, pp), tw, auc_area_ear(aa, nn, pp))
 }
 apply(store, 2, quantile, probs = c(0.025, 0.975), na.rm = TRUE, type = 7)
}

\# ---- main ---------------------------------------------------------------
main <- function() {
 d <- read.csv(CSV, stringsAsFactors = FALSE)
 cat(sprintf("smaihda R v1.9.0 | minn = %d | B = %d | seed = %d\n", MINN, B, SEED))
 cat(sprintf("STEP 1 Data verification: N=%d areas=%d strata=%d\n",
 if (is_table(d)) as.integer(table_n(d)) else nrow(d),
 length(unique(d$area)), length(unique(d$strata_global_id))))
 for (out in OUTCOMES) {
 cl0 <- per_cell(d, out)
 area <- cl0$area; sd <- cl0$sd; n <- cl0$n; ev <- cl0$ev; p <- cl0$p
 dd <- decompose(area, n, p); cf <- coefficients_sm(dd)
 A <- auc_sm(n, p); Aa <- auc_area(area, n, p)
 As <- auc_std(n, p); Aas <- auc_area_std(area, n, p); Aae <- auc_area_ear(area, n, p)
 ci <- bootstrap_sm(area, sd, n, p)
 cat(sprintf("\n===== %s (prevalence %.1f%%) =====\n", out, dd$grand * 100))
 \# v1.5.0 cell-size diagnostic (the basis of the stratum-design rule)
 N <- sum(n); K <- length(n)
 A_n <- length(unique(area)); S_n <- length(unique(sd)); grid <- A_n * S_n
 pct <- function(k) sprintf("%d cells (%.1f%%), %.0f individuals (%.1f%%)", sum(k), 100 * mean(k), sum(n[k]), 100 * sum(n[k]) / N)
 cat(sprintf(" cell-size diagnostic: %d occupied of %d area x stratum cells (%d empty); median n = %.0f, min n = %.0f\n",
 K, grid, grid - K, median(n), min(n)))
 cat(sprintf("	n < 5	: %s\n", pct(n < 5)))
 cat(sprintf("	n < 10 : %s\n", pct(n < 10)))
 cat(sprintf("	n < 30 : %s\n", pct(n < 30)))
 cat(sprintf("	no events (p = 0) : %s\n", pct(ev == 0)))
 cat(sprintf("	all events (p = 1) : %s\n", pct(ev == n)))
 cat(sprintf(" identity weighted : %.8f vs %.8f\n", cf$ident_w, cf$den_w))
 cat(sprintf(" identity standard.: %.8f vs %.8f\n", cf$ident_s, cf$den_s))
 dv <- deviations_sm(area, n, ev)
 cat(sprintf(" STEP 4 deviations (Wilson shift): %d of %d significant; range %.3f to %.3f\n",
 sum(dv$sig), length(dv$sig), min(dv$dev), max(dv$dev)))
 if (dd$K_std < dd$K_all)
 cat(sprintf(" standardised measures use %d of %d cells (n >= %d); %d cells and %.0f individuals excluded\n",
 dd$K_std, dd$K_all, MINN, dd$K_all - dd$K_std, dd$N_excl))
 line <- function(name, val, key)
 cat(sprintf(" %-26s %6.2f%% (%.2f to %.2f)\n", name, val * 100,
 ci[1, key] * 100, ci[2, key] * 100))
 line("VPC_A, crude", cf$VPC_A, "VPC_A")
 line("VPC_SD, crude", cf$VPC_SD, "VPC_SD")
 line("VPC/ICC_A+SD, crude", cf$VPC_ASD, "VPC_ASD")
 line("CCI, crude", cf$GCCI, "GCCI")
 line("VPC_A std, uncorrected", cf$VPC_A_s, "VPC_A_s")
 line("VPC_A std, corrected", cf$VPC_A_c, "VPC_A_c")
 line("VPC_SD std, uncorrected", cf$VPC_SD_s, "VPC_SD_s")
 line("VPC_SD std, corrected", cf$VPC_SD_c, "VPC_SD_c")
 line("VPC/ICC_A+SD std, uncorr", cf$VPC_ASD_s, "VPC_ASD_s")
 line("VPC/ICC_A+SD std, corrected", cf$VPC_ASD_c, "VPC_ASD_c")
 line("CCI, std, uncorrected", cf$GCCI_s, "GCCI_s")
 line("CCI, std, corrected", cf$GCCI_c, "GCCI_c")
 line("coAUC (A-SD strata)", A, "AUC")
 line("coAUC (area level)", Aa, "AUC_area")
 line("coAUC* (A-SD strata)", As, "AUC_s")
 line("coAUC* (area, size)", Aas, "AUC_area_s")
 line("coAUC*EAR (area)", Aae, "AUC_area_ear")
 cl <- classify_sm(n, p, dd$grand)
 cat(sprintf(" threshold (pop. prevalence) = %.2f%%\n", cl$threshold * 100))
 cat(sprintf(" strata flagged high-risk	= %d\n", cl$n_flag))
 cat(sprintf(" sensitivity / specificity = %.1f%% / %.1f%%\n", cl$sens * 100, cl$spec * 100))
 cat(sprintf(" positive predictive value	= %.1f%%\n", cl$ppv * 100))
 cat(sprintf(" TP / FP / FN / TN (indiv.) = %.0f / %.0f / %.0f / %.0f\n",
 cl$TP, cl$FP, cl$FN, cl$TN))
 tw <- three_way(area, sd, n, p)
 cat(" contextual decomposition (standardised; CMI noise-corrected):\n")
 cat(sprintf("	CCI clustering	= %5.1f%% (%.1f to %.1f)\n", tw["CCI"], ci[1, "CCI"], ci[2, "CCI"]))
 cat(sprintf("	G	additive	= %5.1f%% (%.1f to %.1f)\n", tw["G"], ci[1, "G"], ci[2, "G"]))
 cat(sprintf(" CMI modification = %5.1f%% (%.1f to %.1f)\n", tw["CMI"], ci[1, "CMI"], ci[2, "CMI"]))
 flag <- function(val, key) if (!is.na(val) && (val < ci[1, key] || val > ci[2, key])) " (!)" else ""
 f <- paste0(flag(tw["CCI"], "CCI"), flag(tw["G"], "G"), flag(tw["CMI"], "CMI")) # shares and their CIs are both in percent
 if (nchar(f) > 0)
 cat("	(!) a point estimate lies outside its own percentile interval\n")
 }
}

main()

**Output on the Malmö dataset (B = 2000, seed 2026)**

smaihda R v1.9.0 | minn = 10 | B = 2000 | seed = 2026
STEP 1 Data verification: N=43291 areas=25 strata=300

===== psycmed (prevalence 26.0%) =====
 cell-size diagnostic: 300 occupied of 300 area x stratum cells (0 empty); median n = 144, min n = 35
 n < 5	: 0 cells (0.0%), 0 individuals (0.0%)
 n < 10 : 0 cells (0.0%), 0 individuals (0.0%)
 n < 30 : 0 cells (0.0%), 0 individuals (0.0%)
 no events (p = 0) : 0 cells (0.0%), 0 individuals (0.0%)
 all events (p = 1) : 0 cells (0.0%), 0 individuals (0.0%)
 identity weighted : 0.19234832 vs 0.19234832
 identity standard.: 0.19230650 vs 0.19230650
 STEP 4 deviations (Wilson shift): 125 of 300 significant; range -0.188 to 0.250
 VPC_A, crude	0.22% (0.13 to 0.32)
 VPC_SD, crude	3.70% (3.33 to 4.06)
 VPC/ICC_A+SD, crude	3.92% (3.54 to 4.30)
 CCI, crude	5.64% (3.85 to 7.68)
 VPC_A std, uncorrected	0.20% (0.11 to 0.31)
 VPC_A std, corrected	0.14% (0.05 to 0.25)
 VPC_SD std, uncorrected	3.86% (3.47 to 4.26)
 VPC_SD std, corrected	3.16% (2.76 to 3.56)
 VPC/ICC_A+SD std, uncorr	4.06% (3.64 to 4.48)
 VPC/ICC_A+SD std, corrected	3.30% (2.87 to 3.72)
 CCI, std, uncorrected	4.87% (3.07 to 7.02)
 CCI, std, corrected	4.14% (2.00 to 6.70)
 coAUC (A-SD strata)	62.71% (62.14 to 63.27)
 coAUC (area level)	52.97% (52.40 to 53.55)
 coAUC* (A-SD strata)	62.97% (62.35 to 63.56)
 coAUC* (area, size)	53.22% (52.61 to 53.84)
 coAUC*EAR (area)	52.86% (52.25 to 53.52)
 threshold (pop. prevalence) = 25.99%
 strata flagged high-risk	= 142
 sensitivity / specificity	= 59.7% / 58.3%
 positive predictive value	= 33.5%
 TP / FP / FN / TN (indiv.) = 6717 / 13352 / 4534 / 18688
 contextual decomposition (standardised; CMI noise-corrected):
 CCI clustering	=	4.1% (2.0 to 6.7)
 G	additive	= 95.9% (90.6 to 100.0)
 CMI modification =	0.0% (0.0 to 5.0)

===== private (prevalence 22.1%) =====
 cell-size diagnostic: 300 occupied of 300 area x stratum cells (0 empty); median n = 144, min n = 35
 n < 5	: 0 cells (0.0%), 0 individuals (0.0%)
 n < 10 : 0 cells (0.0%), 0 individuals (0.0%)
 n < 30 : 0 cells (0.0%), 0 individuals (0.0%)
 no events (p = 0) : 0 cells (0.0%), 0 individuals (0.0%)
 all events (p = 1) : 0 cells (0.0%), 0 individuals (0.0%)
 identity weighted : 0.17216788 vs 0.17216788
 identity standard.: 0.17493753 vs 0.17493753
 STEP 4 deviations (Wilson shift): 122 of 300 significant; range -0.313 to 0.236
 VPC_A, crude	7.37% (6.87 to 7.84)
 VPC_SD, crude	3.80% (3.43 to 4.16)
 VPC/ICC_A+SD, crude	11.17% (10.56 to 11.80)
 CCI, crude	65.95% (63.43 to 68.48)
 VPC_A std, uncorrected	6.91% (6.38 to 7.44)
 VPC_A std, corrected	6.86% (6.32 to 7.38)
 VPC_SD std, uncorrected	4.53% (4.05 to 5.00)
 VPC_SD std, corrected	3.87% (3.39 to 4.35)
 VPC/ICC_A+SD std, uncorr	11.44% (10.76 to 12.13)
 VPC/ICC_A+SD std, corrected 10.72% (10.04 to 11.42)
 CCI, std, uncorrected	60.43% (57.39 to 63.68)
 CCI, std, corrected	63.94% (60.69 to 67.42)
 coAUC (A-SD strata)	71.77% (71.20 to 72.29)
 coAUC (area level)	67.90% (67.31 to 68.45)
 coAUC* (A-SD strata)	71.90% (71.30 to 72.46)
 coAUC* (area, size)	68.38% (67.79 to 68.92)
 coAUC*EAR (area)	67.46% (66.82 to 68.05)
 threshold (pop. prevalence) = 22.10%
 strata flagged high-risk	= 130
 sensitivity / specificity	= 66.5% / 65.7%
 positive predictive value	= 35.5%
 TP / FP / FN / TN (indiv.) = 6366 / 11583 / 3202 / 22140
 contextual decomposition (standardised; CMI noise-corrected):
 CCI clustering	= 63.9% (60.7 to 67.4)
 G	additive	= 19.0% (16.7 to 21.3)
 CMI modification = 17.1% (14.3 to 20.0)
