## Supplementary material for "Describing health inequalities without distortion: Simple-Means MAIHDA vs Random-Effects MAIHDA": S6 File. Programs that draw the result figures: fig3_decomposition.py

```
# -*- coding: utf-8 -*-
"""Figure 3 of the paper. How an observed prevalence splits into four absolute differences, and
what the general contextual measures summarise.

The same intersectional position, high-income women aged 45 to 54, in three areas ordered by
their level, for private GP choice. Each observed prevalence is the exact sum of the reference
p* (the plain mean of the 300 stratum proportions), the area ARD (the composition-standardised
prevalence of the area minus p*), the common ARD of the stratum (its deviation from the level of
its area averaged over the 25 areas) and the modification of the ARD by the area. No model is
fitted. The first two terms are area terms: together they give the level of the area, its EAR.
The last two are stratum terms: together they give the deviation of the stratum in that area from
the level of its area. The common ARD is the part the stratum carries into every area; the
modification belongs to the stratum in that area. The dashed line across the white bar marks the
expected prevalence, the level of the area plus the common ARD. The variances of the area ARD*,
the common ARDs and the modifications over all strata give the three shares CCI, G and CMI,
labelled in the third panel.
Adapted from the program of Figure 2 of the companion study of the European Union.
Reads the stratum table (S5 File) and writes the figure next to this script.
    python fig3_decomposition.py
Needs pandas and matplotlib.
"""
import os
import pandas as pd, matplotlib
matplotlib.use("Agg")
import matplotlib.pyplot as plt
from matplotlib.patches import Rectangle

HERE = os.path.dirname(os.path.abspath(__file__))
t = pd.read_csv(os.path.join(HERE, "..", "S5_File_stratum_table", "dataset-smaihda_stratum_table.csv"))
g = t[t.outcome == "private"].copy(); g["p"] = g.cases / g.n
pstar = g.p.mean(); ear = g.groupby("area").p.mean()
g["dev"] = g.p - g.area.map(ear)
delta = g.groupby("strata_sociodem_id").dev.mean(); g["gam"] = g.dev - g.strata_sociodem_id.map(delta)
key = g.groupby("strata_sociodem_id")[["sex", "age3", "income"]].first()
SD = int(key[(key.sex == 0) & (key.age3 == 2) & (key.income == 1)].index[0])   # women, 45-54, high income
AREAS = (21, 18, 24)
COL = {"ref": "0.75", "area": "#4c72b0", "add": "#55a868", "mod": "#dd8452"}
YMAX = 92
fig, axes = plt.subplots(1, 3, figsize=(16.0, 5.8), sharey=True)
for ax, a in zip(axes, AREAS):
    r = g[(g.area == a) & (g.strata_sociodem_id == SD)].iloc[0]
    comp = [("Reference\np*", pstar * 100, "ref"), ("Area\nARD*", (ear[a] - pstar) * 100, "area"),
            ("Common\nARD", delta[SD] * 100, "add"), ("Modification\nof the ARD", r.gam * 100, "mod")]
    level = 0; tops = {}
    for k, (lab, val, key_) in enumerate(comp):
        if key_ == "ref":
            ax.add_patch(Rectangle((k - 0.35, 0), 0.7, val, fc=COL[key_], ec="black", lw=0.8)); level = val
            ax.text(k, val + 1.5, f"{val:.1f}", ha="center", fontsize=10.5)
        else:
            y0 = level if val >= 0 else level + val
            ax.add_patch(Rectangle((k - 0.35, y0), 0.7, abs(val), fc=COL[key_], ec="black", lw=0.8))
            ax.plot([k - 1 + 0.35, k - 0.35], [level, level], color="black", lw=0.7, ls=":")
            ax.text(k, max(level, level + val) + 1.5, "0.0" if abs(val) < 0.05 else f"{val:+.1f}", ha="center", fontsize=10.5)
            tops[key_] = (k, y0 + abs(val) / 2)
            level += val
    assert abs(level - r.p * 100) < 1e-9          # the four terms add up to the observed prevalence
    ax.add_patch(Rectangle((4 - 0.35, 0), 0.7, r.p * 100, fc="white", ec="black", lw=1.4))
    expected = (ear[a] + delta[SD]) * 100
    ax.plot([4 - 0.45, 4 + 0.45], [expected, expected], color="black", lw=1.3, ls=(0, (4, 2)), zorder=5)
    ax.text(4 + 0.47, expected, f"expected\n{expected:.1f}", ha="left", va="center", fontsize=8.5)
    # the two blocks: area terms and stratum terms
    for x0, x1, txt in ((-0.35, 1.35, "Area terms\n= level of the area"),
                        (1.65, 3.35, "Stratum terms\n= the stratum in this area")):
        yb = YMAX - 9
        ax.plot([x0, x0, x1, x1], [yb - 1.5, yb, yb, yb - 1.5], color="0.2", lw=1.0, clip_on=False)
        ax.text((x0 + x1) / 2, yb + 1.0, txt, ha="center", va="bottom", fontsize=8.6, color="0.15")
    ax.plot([3.35, 4 - 0.35], [level, level], color="black", lw=0.7, ls=":")
    ax.text(4, r.p * 100 + 1.5, f"{r.p*100:.1f}", ha="center", fontsize=11, fontweight="bold")
    if a == AREAS[-1]:
        arr = dict(arrowstyle="-", color="0.25", lw=0.9, shrinkA=0, shrinkB=0)
        for key_, name, dx, dy in (("area", "CCI", -0.55, 14), ("add", "G", -0.6, 12), ("mod", "CMI", -0.65, 10)):
            k, ym = tops[key_]
            ax.annotate(name, xy=(k - 0.3, ym), xytext=(k + dx, min(ym + dy, YMAX - 4)), fontsize=10.5, fontweight="bold",
                        color=COL[key_], ha="center", arrowprops=arr)
    ax.set_xticks(range(5)); ax.set_xticklabels([c[0] for c in comp] + [f"Observed\nprevalence\n(n = {int(r.n)})"], fontsize=8.8)
    kind = "rich" if g[g.area == a].area_income.iloc[0] == 1 else "poor"
    ax.set_title(f"Area {a} ({kind}), level {ear[a]*100:.1f}%", fontsize=12, fontweight="bold")
    ax.tick_params(axis="y", labelsize=10); ax.set_xlim(-0.65, 5.0); ax.set_ylim(0, YMAX)
    for s in ("top", "right"): ax.spines[s].set_visible(False)
    print(f"area {a}: {pstar*100:.1f} {100*(ear[a]-pstar):+.1f} {100*delta[SD]:+.1f} {100*r.gam:+.1f} = {100*r.p:.1f} ({r.cases}/{r.n})")
axes[0].set_ylabel("Private general practitioner choice (%)", fontsize=10.5)
fig.suptitle("The same intersectional position in three areas: high-income women aged 45 to 54", fontsize=10.5, y=0.995)
fig.tight_layout(rect=(0, 0, 1, 0.96))
fig.savefig(os.path.join(HERE, "Fig3_decomposition.png"), dpi=400); fig.savefig(os.path.join(HERE, "Fig3_decomposition.pdf"))
```
