## Supplementary material for "Describing health inequalities without distortion: Simple-Means MAIHDA vs Random-Effects MAIHDA": S6 File. Programs that draw the result figures: fig8_modification_radar.py

```
# -*- coding: utf-8 -*-
"""Figures 8a and 8b of the paper. Modification of the sociodemographic
differences by the area.

One polar panel per sociodemographic stratum. The 25 geographical areas are the
spokes, ordered clockwise by their standardised prevalence; the order is the same
in every panel and in both outcomes.

Blue   the standardised prevalence of the area (EAR: each of the 12 strata counts once)
Green  the expectation under additivity: area level + the difference of that stratum
       common to all areas
Black  the observed prevalence
Orange the modification, that is the gap between green and black

A filled point marks a modification larger than 1.96 standard errors of the
observed proportion.

Reads the stratum table (S5 File) and writes the figures next to this script.
    python fig8_modification_radar.py
Needs numpy, pandas and matplotlib.
"""
import os
import numpy as np, pandas as pd, matplotlib
matplotlib.use('Agg')
import matplotlib.pyplot as plt

HERE = os.path.dirname(os.path.abspath(__file__))
TABLE = os.path.join(HERE, "..", "S5_File_stratum_table", "dataset-smaihda_stratum_table.csv")
t = pd.read_csv(TABLE)

SEX = {0: 'Women', 1: 'Men'}
AGE = {1: '35-44', 2: '45-54', 3: '55-64'}
INC = {0: 'Low income', 1: 'High income'}
OUT = {'psycmed': ('Psychotropic medication use', 'Fig8a_modification_psycmed'),
       'private': ('Private GP choice', 'Fig8b_modification_private')}

key = t.groupby('strata_sociodem_id')[['sex', 'age3', 'income']].first()
area_income = t.groupby('area').area_income.first()        # 0 poor, 1 rich
BLUE, GREEN, ORANGE = '#4c72b0', '#55a868', '#dd8452'
CPOOR, CRICH = '#a03030', '#33556f'

for y, (title, stem) in OUT.items():
    g = (t[t.outcome == y][['area', 'strata_sociodem_id', 'n', 'cases']]
           .sort_values(['area', 'strata_sociodem_id']).reset_index(drop=True))
    g['p'] = g.cases / g.n * 100
    ear = g.groupby('area').p.mean()
    g['D'] = g.p - g.area.map(ear)
    delta = g.groupby('strata_sociodem_id').D.mean()
    order = ear.sort_values().index

    lo, hi = min(0, g.p.min()), np.ceil(g.p.max() / 10) * 10
    fig, axes = plt.subplots(3, 4, figsize=(17, 13.5), subplot_kw=dict(polar=True))
    strata = [int(key[(key.sex == s) & (key.age3 == a) & (key.income == i)].index[0])
              for a in (1, 2, 3) for s in (0, 1) for i in (0, 1)]
    nsig = 0
    for ax, sid in zip(axes.ravel(), strata):
        sub = g[g.strata_sociodem_id == sid].set_index('area').loc[order]
        lev = ear.loc[sub.index].values
        exp = np.maximum(lev + delta[sid], 0)
        obs = sub.p.values
        th = np.linspace(0, 2 * np.pi, len(sub), endpoint=False)
        thc = np.r_[th, th[:1]]
        cl = lambda v: np.r_[v, v[:1]]
        ax.fill_between(thc, cl(exp), cl(obs), color=ORANGE, alpha=0.5, lw=0)
        ax.plot(thc, cl(lev), color=BLUE, lw=1.4)
        ax.plot(thc, cl(exp), color=GREEN, lw=1.4)
        ax.plot(thc, cl(obs), color='black', lw=1.7)
        pa = (sub.p.values / 100 * sub.n.values + 1) / (sub.n.values + 2)
        se = np.sqrt(pa * (1 - pa) / sub.n.values) * 100
        sig = np.abs(obs - exp) > 1.96 * se
        nsig += int(sig.sum())
        ax.scatter(th[sig], obs[sig], color='black', s=13, zorder=5)
        ax.scatter(th[~sig], obs[~sig], facecolor='white', edgecolor='black',
                   s=13, lw=0.7, zorder=5)
        ax.set_xticks(th); ax.set_xticklabels([str(a) for a in sub.index], fontsize=7)
        for tl, a, s in zip(ax.get_xticklabels(), sub.index, sig):
            tl.set_color(CPOOR if area_income[a] == 0 else CRICH)
            tl.set_fontweight('bold' if s else 'normal')
        ax.set_theta_zero_location('N'); ax.set_theta_direction(-1)
        ax.set_ylim(lo, hi); ax.set_yticks(np.arange(10, hi + 1, 10))
        ax.set_rlabel_position(8); ax.tick_params(axis='y', labelsize=6.5)
        r = key.loc[sid]
        ax.set_title(f"{SEX[r.sex]}, {AGE[r.age3]}, {INC[r.income]}", fontsize=10, pad=14)

    h = [plt.Line2D([], [], color=BLUE, lw=1.6, label='Area level (EAR-standardised)'),
         plt.Line2D([], [], color=GREEN, lw=1.6,
                    label='Expected = area level + common difference'),
         plt.Line2D([], [], color='black', lw=1.8, label='Observed'),
         plt.Rectangle((0, 0), 1, 1, color=ORANGE, alpha=0.5,
                       label='Modification by the area'),
         plt.Line2D([], [], marker='o', color='black', lw=0, ms=5,
                    label='Modification beyond 1.96 SE'),
         plt.Line2D([], [], marker='o', mfc='white', mec='black', color='w', lw=0, ms=5,
                    label='Modification within 1.96 SE')]
    fig.legend(handles=h, loc='lower center', ncol=3, frameon=False, fontsize=10,
               bbox_to_anchor=(0.5, -0.005))
    fig.suptitle(f"{title}. Spokes are the 25 geographical areas, "
                 f"ordered by their standardised prevalence.\n"
                 f"Area numbers in red are poor areas, in blue rich areas.",
                 fontsize=12, y=0.995)
    fig.tight_layout(rect=[0, 0.035, 1, 0.965])
    for ext in ('png', 'pdf'):
        fig.savefig(os.path.join(HERE, f"{stem}.{ext}"),
                    dpi=250 if ext == 'png' else None, bbox_inches='tight')
    plt.close(fig)
    print(f"{title}: {nsig} of {len(g)} strata ({nsig/len(g)*100:.1f}%) with a modification "
          f"beyond 1.96 SE  ->  {stem}.png and .pdf")
```
