## Supplementary material for "Describing health inequalities without distortion: Simple-Means MAIHDA vs Random-Effects MAIHDA": S6 File. Programs that draw the result figures: fig9_roc_discriminatory_accuracy.py

```
# -*- coding: utf-8 -*-
"""Figure 9 of the paper. Discriminatory accuracy of the context.

Receiver operating characteristic curves for the two outcomes, with individuals
ranked by the observed prevalence of their area-sociodemographic stratum (black)
and by that of their area alone (blue).

The curves join the vertices, one per distinct prevalence, so the area under the
plotted curve is the trapezoidal AUC, which equals the coAUC with ties counted as one
half. The script checks that identity, and that the classification counts at the
population prevalence are those of Table 7. The coAUC and its 95% interval are taken
from the Python implementation of the paper (S2 File): within-context bootstrap,
2,000 replicates, seed 2026.

Reads the stratum table (S5 File) and S2_File_smaihda_python.py, and writes the
figure next to this script.
    python fig9_roc_discriminatory_accuracy.py
Needs numpy, pandas and matplotlib.
"""
import importlib.util
import os
import sys
import numpy as np, pandas as pd, matplotlib
matplotlib.use('Agg')
import matplotlib.pyplot as plt

HERE = os.path.dirname(os.path.abspath(__file__))
TABLE = os.path.join(HERE, "..", "S5_File_stratum_table", "dataset-smaihda_stratum_table.csv")
ENGINE = os.path.join(HERE, "..", "S2_File_smaihda_python.py")

spec = importlib.util.spec_from_file_location("smaihda_engine", ENGINE)
eng = importlib.util.module_from_spec(spec)
argv, sys.argv = sys.argv, sys.argv[:1]      # the engine reads its own command line
spec.loader.exec_module(eng)
sys.argv = argv
rows = eng.load(TABLE)
trapezoid = getattr(np, "trapezoid", None) or np.trapz


def roc_points(score, pos, cnt):
    g = (pd.DataFrame({'s': score, 'pos': pos, 'n': cnt}).groupby('s')
           .sum().sort_index(ascending=False))
    tp = np.r_[0, np.cumsum(g.pos.values)]
    fp = np.r_[0, np.cumsum((g.n - g.pos).values)]
    return fp / fp[-1], tp / tp[-1]


OUT = {'psycmed': 'Psychotropic medication use', 'private': 'Private GP choice'}
TAB7 = {'psycmed': dict(TP=6717, FP=13352, FN=4534, TN=18688),
        'private': dict(TP=6366, FP=11583, FN=3202, TN=22140)}

res = {}
for y in OUT:
    area, sd, n, ev, p = eng.per_cell(rows, y)
    ci = eng.bootstrap(area, sd, n, p)
    areas = np.unique(area)
    n_a = np.array([n[area == a].sum() for a in areas])
    ev_a = np.array([ev[area == a].sum() for a in areas])
    out = {}
    for k, sc, pos, cnt, est, key in (('asd', p, ev, n, eng.auc(n, p), 'AUC'),
                                      ('area', ev_a / n_a, ev_a, n_a,
                                       eng.auc_area(area, n, p), 'AUC_area')):
        f, tpr = roc_points(sc, pos, cnt)
        assert abs(trapezoid(tpr, f) - est) < 1e-9, "trapezoidal area and AUC differ"
        out[k] = (f, tpr, est * 100, ci[key][0] * 100, ci[key][1] * 100)
    prev = ev.sum() / n.sum() * 100
    flag = p * 100 > prev
    TP, FP = int(ev[flag].sum()), int((n - ev)[flag].sum())
    FN, TN = int(ev[~flag].sum()), int((n - ev)[~flag].sum())
    assert (TP, FP, FN, TN) == (TAB7[y]['TP'], TAB7[y]['FP'],
                                TAB7[y]['FN'], TAB7[y]['TN']), "counts differ from Table 7"
    out['op'] = (1 - TN / (TN + FP), TP / (TP + FN), prev,
                 TP / (TP + FN) * 100, TN / (TN + FP) * 100)
    res[y] = out
    print(f"{OUT[y]}: AUC A-SD {out['asd'][2]:.2f}% ({out['asd'][3]:.2f} to {out['asd'][4]:.2f}), "
          f"area {out['area'][2]:.2f}% ({out['area'][3]:.2f} to {out['area'][4]:.2f}); "
          f"TP/FP/FN/TN as in Table 7")

BLACK, BLUE, GREY = '#000000', '#4c72b0', '#aaaaaa'
fig, axes = plt.subplots(1, 2, figsize=(10.5, 5.2))
for ax, y in zip(axes, OUT):
    r = res[y]
    ax.plot([0, 1], [0, 1], color=GREY, lw=0.9, ls=(0, (4, 4)), zorder=1)
    f, tpr, a, lo, hi = r['area']
    ax.plot(f, tpr, color=BLUE, lw=1.7, zorder=2,
            label=f"Area only, coAUC {a:.1f}% ({lo:.1f} to {hi:.1f})")
    f, tpr, a, lo, hi = r['asd']
    ax.plot(f, tpr, color=BLACK, lw=1.9, zorder=3,
            label=f"Area-sociodemographic strata, coAUC {a:.1f}% ({lo:.1f} to {hi:.1f})")
    x0, y0, prev, sens, spec_ = r['op']
    ax.scatter([x0], [y0], s=52, color=BLACK, zorder=5)
    ax.annotate(f"threshold {prev:.1f}%\nsens {sens:.1f}%, spec {spec_:.1f}%",
                (x0, y0), textcoords="offset points", xytext=(14, 4),
                fontsize=8.5, ha='left', va='bottom')
    ax.set_xlim(0, 1); ax.set_ylim(0, 1.005)
    ax.set_xlabel("1 − specificity"); ax.set_ylabel("Sensitivity")
    ax.set_title(OUT[y], fontsize=11)
    ax.legend(loc='lower right', fontsize=8, frameon=False)
    ax.set_aspect('equal')
    for s in ('top', 'right'):
        ax.spines[s].set_visible(False)
fig.tight_layout()
for ext in ('png', 'pdf'):
    fig.savefig(os.path.join(HERE, f"Fig9_ROC_discriminatory_accuracy.{ext}"),
                dpi=400 if ext == 'png' else None, bbox_inches='tight')
print("-> Fig9_ROC_discriminatory_accuracy.png and .pdf")
```
