## Supplementary material for "Describing health inequalities without distortion: Simple-Means MAIHDA vs Random-Effects MAIHDA": S6 File. Programs that draw the result figures: fig10_compare_diagnostic.R

```
# Figure 10 of the paper. The compare diagnostic, stratum by stratum.
#
# Fits the same random-intercept logistic model that the compare option of the
# smaihda command fits (S1 File), here with lme4::glmer instead of Stata's
# melogit. The model has no covariates, so it is fitted to the stratum table in
# binomial form, cbind(cases, n - cases), which has the same likelihood as the
# individual records.
#
# Top row     the observed proportion of each of the 300 strata against the
#             random-effects prediction, with the identity line. The compression
#             of the range is the compression of the cloud.
# Bottom row  the displacement of each stratum against its distance from the
#             population prevalence, with the area of the point proportional to
#             the number of individuals. The largest displacement is not the
#             smallest stratum.
#
# The script stops unless it agrees closely with Table 8 of the paper, which
# comes from Stata's melogit: the range compression factor within one point,
# the maximum absolute attenuation within 0.3 points, and the same size of the
# stratum where that maximum falls.
#
# It also prints the latent intraclass correlation and the median odds ratio of
# the same model, and of the model with a random intercept for the area, which
# Section 3.4 reports for comparison with the literature.
#
# Reads the stratum table (S5 File) and writes the figure next to this script.
#   Rscript fig10_compare_diagnostic.R
# Needs the lme4 package.

suppressMessages(library(lme4))
arg <- grep("^--file=", commandArgs(FALSE), value = TRUE)
HERE <- if (length(arg)) dirname(normalizePath(sub("^--file=", "", arg[1]))) else getwd()
tab <- read.csv(file.path(HERE, "..", "S5_File_stratum_table", "dataset-smaihda_stratum_table.csv"))

TAB8 <- list(psycmed = list(comp = 83.1, att = 6.44, n = 75,
                            lab = "Psychotropic medication use"),
             private = list(comp = 94.3, att = 4.14, n = 44,
                            lab = "Private GP choice"))
GREY <- "#999999"; BLUE <- "#4c72b0"; ORANGE <- "#dd8452"
R <- list()

for (y in names(TAB8)) {
  s <- tab[tab$outcome == y, ]
  s$asd <- factor(paste(s$area, s$strata_sociodem_id, sep = "_"))
  m <- glmer(cbind(cases, n - cases) ~ 1 + (1 | asd), data = s,
             family = binomial, nAGQ = 7)
  pre <- plogis(as.numeric(fixef(m)) + ranef(m)$asd[, 1]) * 100
  names(pre) <- rownames(ranef(m)$asd)
  nn <- tapply(s$n, s$asd, sum)
  obs <- tapply(s$cases, s$asd, sum) / nn * 100
  pre <- pre[names(obs)]
  gm <- sum(s$cases) / sum(s$n) * 100
  att <- pre - obs
  comp <- (max(pre) - min(pre)) / (max(obs) - min(obs)) * 100
  i <- which.max(abs(att))
  tg <- TAB8[[y]]

  cat(sprintf("%-28s range compression %.2f%% (Table 8: %.1f), maximum attenuation %.2f pp at n = %d (Table 8: %.2f at n = %d)\n",
              tg$lab, comp, tg$comp, max(abs(att)), as.integer(nn[i]), tg$att, tg$n))
  stopifnot(abs(comp - tg$comp) < 1.0,
            abs(max(abs(att)) - tg$att) < 0.3,
            as.integer(nn[i]) == tg$n)
  # Latent-scale quantities of Section 3.4: between-stratum variance, latent
  # intraclass correlation and median odds ratio (MOR), for the A-SD strata and
  # for the areas alone (V2.6)
  ma <- glmer(cbind(cases, n - cases) ~ 1 + (1 | area), data = s,
              family = binomial, nAGQ = 7)
  v <- c(asd = as.numeric(VarCorr(m)$asd), area = as.numeric(VarCorr(ma)$area))
  mor <- exp(sqrt(2 * v) * qnorm(0.75)); icc <- 100 * v / (v + pi^2 / 3)
  cat(sprintf("%-28s A-SD strata: variance %.2f, latent ICC %.1f%%, MOR %.2f | areas: latent ICC %.1f%%, MOR %.2f\n",
              tg$lab, v[["asd"]], icc[["asd"]], mor[["asd"]], icc[["area"]], mor[["area"]]))
  TEXT <- list(psycmed = c(1.50, 1.10), private = c(2.15, 1.89))
  stopifnot(all(abs(round(mor, 2) - TEXT[[y]]) < 0.006))
  R[[y]] <- list(obs = obs, pre = pre, n = as.numeric(nn), att = att, gm = gm,
                 comp = comp, i = i, lab = tg$lab)
  cat(sprintf("   correlation of |attenuation| with n: %+.2f, with the distance from the prevalence: %+.2f\n",
              cor(abs(att), nn), cor(abs(att), abs(obs - gm))))
}

for (ext in c("png", "pdf")) {
  f <- file.path(HERE, paste0("Fig10_compare_diagnostic.", ext))
  if (ext == "png") png(f, width = 4200, height = 4000, res = 380)
  else pdf(f, width = 11, height = 10.5)
  par(mfrow = c(2, 2), mar = c(4.4, 4.4, 3.0, 1.2))

  for (y in names(R)) {                       # top row: observed against model
    r <- R[[y]]
    lim <- range(c(r$obs, r$pre))
    plot(r$obs, r$pre, pch = 21, bg = adjustcolor(BLUE, 0.45), col = BLUE,
         cex = 0.35 + 1.5 * sqrt(r$n / max(r$n)), xlim = lim, ylim = lim,
         xlab = "Observed proportion (%)",
         ylab = "Random-effects prediction (%)", bty = "n",
         main = r$lab, cex.main = 1.05)
    abline(0, 1, col = "black", lwd = 1.4)
    abline(h = r$gm, v = r$gm, col = GREY, lty = 3, lwd = 1.0)
    points(r$obs[r$i], r$pre[r$i], pch = 21, bg = ORANGE, col = "black",
           cex = 0.35 + 1.5 * sqrt(r$n[r$i] / max(r$n)), lwd = 1.3)
    legend("topleft", bty = "n", cex = 0.82,
           legend = c(sprintf("range compression %.1f%%", r$comp),
                      "area of the point is the stratum size",
                      "dotted lines: the population prevalence"))
  }
  for (y in names(R)) {                       # bottom row: displacement and distance
    r <- R[[y]]
    dist <- r$obs - r$gm
    plot(dist, r$att, pch = 21, bg = adjustcolor(BLUE, 0.45), col = BLUE,
         cex = 0.35 + 1.5 * sqrt(r$n / max(r$n)),
         xlab = "Distance of the observed proportion from the population prevalence (pp)",
         ylab = "Displacement by the model (pp)", bty = "n",
         main = r$lab, cex.main = 1.05)
    abline(h = 0, col = GREY, lty = 3); abline(v = 0, col = GREY, lty = 3)
    points(dist[r$i], r$att[r$i], pch = 21, bg = ORANGE, col = "black",
           cex = 0.35 + 1.5 * sqrt(r$n[r$i] / max(r$n)), lwd = 1.3)
    text(dist[r$i], r$att[r$i], sprintf("largest displacement\n%.1f pp, n = %d   ",
                                        r$att[r$i], as.integer(r$n[r$i])),
         adj = c(1, 0.1), cex = 0.72)
    legend("topright", bty = "n", cex = 0.82,
           legend = c("every stratum is pulled towards the mean",
                      "the pull grows with the distance,",
                      "and shrinks as the stratum grows",
                      sprintf("correlation with the distance %+.2f, with the size %+.2f",
                              cor(abs(r$att), abs(dist)), cor(abs(r$att), r$n))))
  }
  invisible(dev.off())
}
cat("-> Fig10_compare_diagnostic.png and .pdf\n")
```
