## Supplementary material for "Describing health inequalities without distortion: Simple-Means MAIHDA vs Random-Effects MAIHDA": S6 File. Programs that draw the result figures: fig45_prevalence_and_deviations.py

```
# -*- coding: utf-8 -*-
"""Figures 4a, 4b, 5a and 5b of the paper.

Figure 4  the observed prevalence of each of the 300 area-sociodemographic
          strata, with its Wilson 95% interval; a black tick marks the prevalence
          of each sociodemographic stratum in all areas pooled (V2.6, as Figure 3a
          of the companion study of the European Union)
Figure 5  the deviation of each stratum from the level of its own area, the
          composition-standardised prevalence (EAR, the plain mean of the 12
          stratum proportions of the area), with the Wilson-shift interval
          (V2.6, 7 October 2026; the earlier versions used the crude area
          prevalence, which the graph option of smaihda still draws)

The construction follows the graph option of the smaihda command (S1 File):
  y position   yy = s + 0.72*((rank-1)/(n-1) - 0.5), rank by prevalence within
               the sociodemographic stratum s, axis reversed
  interval     Wilson 95% interval of the stratum proportion
  deviation    dev = p_stratum - p*_area; its interval is the Wilson interval of
               the stratum shifted by p*_area, which is treated as known. With
               this level the deviations of every area average zero, and their
               mean over the 25 areas is the common ARD of the stratum
  significance dev_lo > 0 or dev_hi < 0
  colour       poor areas blue, rich areas orange

Reads the stratum table (S5 File) and writes the figures next to this script.
    python fig45_prevalence_and_deviations.py
Needs numpy, pandas and matplotlib.
"""
import os
import numpy as np, pandas as pd, matplotlib
matplotlib.use('Agg')
import matplotlib.pyplot as plt

HERE = os.path.dirname(os.path.abspath(__file__))
TABLE = os.path.join(HERE, "..", "S5_File_stratum_table", "dataset-smaihda_stratum_table.csv")
t = pd.read_csv(TABLE)
Z = 1.96

DOT_P, DOT_R = '#1f77b4', '#ff7f0e'          # 31 119 180 | 255 127 14
BAR_P, BAR_R = '#aec7e8', '#ffc896'          # 174 199 232 | 255 200 150
SEX = {0: 'W', 1: 'M'}
AGE = {1: '35-44', 2: '45-54', 3: '55-64'}
INC = {0: 'Low', 1: 'High'}
OUT = {'psycmed': ('Psychotropic medication use', 'psycmed'),
       'private': ('Private GP choice', 'private')}


def wilson(k, n, z=Z):
    p = k / n
    den = 1 + z * z / n
    c = (p + z * z / (2 * n)) / den
    h = z * np.sqrt(p * (1 - p) / n + z * z / (4 * n * n)) / den
    return c - h, c + h


key = t.groupby('strata_sociodem_id')[['sex', 'age3', 'income']].first()
LAB = {s: f"{SEX[r.sex]}, {AGE[r.age3]}, {INC[r.income]}" for s, r in key.iterrows()}
area_income = t.groupby('area').area_income.first()          # 0 poor, 1 rich

for y, (title, stem) in OUT.items():
    g = (t[t.outcome == y][['area', 'strata_sociodem_id', 'cases', 'n']]
           .rename(columns={'cases': 'k'})
           .sort_values(['area', 'strata_sociodem_id']).reset_index(drop=True))
    g['p'] = g.k / g.n
    g['lo'], g['hi'] = wilson(g.k.values, g.n.values)
    parea = g.groupby('area').p.mean()          # EAR: composition-standardised level
    g['parea'] = g.area.map(parea)
    g['dev'] = g.p - g.parea
    g['dev_lo'] = g.lo - g.parea
    g['dev_hi'] = g.hi - g.parea
    g['sig'] = (g.dev_lo > 0) | (g.dev_hi < 0)
    g['rich'] = g.area.map(area_income).values == 1
    for c in ('p', 'lo', 'hi', 'dev', 'dev_lo', 'dev_hi'):
        g[c] *= 100

    g['rk'] = g.groupby('strata_sociodem_id').p.rank(method='first')
    kk = g.groupby('strata_sociodem_id').p.transform('size')
    g['yy'] = g.strata_sociodem_id + 0.72 * ((g.rk - 1) / (kk - 1) - 0.5)
    thr = g.k.sum() / g.n.sum() * 100

    # ---------------- Figure 4: prevalence
    fig, ax = plt.subplots(figsize=(9.6, 8.2))
    for rich, cbar, cdot in ((False, BAR_P, DOT_P), (True, BAR_R, DOT_R)):
        s = g[g.rich == rich]
        ax.hlines(s.yy, s.lo, s.hi, color=cbar, lw=0.9, zorder=2)
        ax.scatter(s.p, s.yy, s=15, color=cdot, zorder=3,
                   label='Rich area' if rich else 'Poor area')
    ax.axvline(thr, color='black', lw=1.0, ls=(0, (5, 4)), zorder=4)
    ax.text(thr, 0.35, f"{thr:.1f}%", ha='center', va='bottom', fontsize=9)
    sk = g.groupby('strata_sociodem_id')[['k', 'n']].sum(); pooled = sk.k / sk.n * 100
    for sd, v in pooled.items():                     # the stratum in all areas pooled
        ax.plot([v, v], [sd - 0.42, sd + 0.42], color='black', lw=1.8, zorder=5)
    ax.plot([], [], color='black', lw=1.8, label='Stratum, all areas pooled')
    for s in range(1, 12):
        ax.axhline(s + 0.5, color='#dddddd', lw=0.8, zorder=1)
    ax.set_yticks(range(1, 13)); ax.set_yticklabels([LAB[s] for s in range(1, 13)],
                                                    fontsize=9)
    ax.invert_yaxis(); ax.set_ylim(12.6, 0.3)
    ax.set_xlabel("Prevalence (%)", fontsize=11)
    ax.legend(loc='upper center', bbox_to_anchor=(0.5, -0.075), ncol=3, fontsize=9,
              frameon=False, markerscale=1.2)   # below the axes: no overlap with the dots
    for sp in ('top', 'right'):
        ax.spines[sp].set_visible(False)
    fig.tight_layout()
    lab = 'a' if y == 'psycmed' else 'b'
    for ext in ('png', 'pdf'):
        fig.savefig(os.path.join(HERE, f"Fig4{lab}_prevalence_{stem}.{ext}"),
                    dpi=400 if ext == 'png' else None, bbox_inches='tight')
    plt.close(fig)

    # ---------------- Figure 5: deviations
    fig, ax = plt.subplots(figsize=(9.6, 8.2))
    for rich, cbar, cdot, nm in ((False, BAR_P, DOT_P, 'Poor area'),
                                 (True, BAR_R, DOT_R, 'Rich area')):
        s = g[g.rich == rich]
        ax.hlines(s.yy, s.dev_lo, s.dev_hi, color=cbar, lw=0.9, zorder=2)
        a = s[s.sig]
        ax.scatter(a.dev, a.yy, s=17, color=cdot, zorder=3,
                   label=f"{nm}, 95% CI excludes 0")
        b = s[~s.sig]
        ax.scatter(b.dev, b.yy, s=17, facecolor='white', edgecolor=cdot, lw=1.0,
                   zorder=3)
    ax.scatter([], [], s=17, facecolor='white', edgecolor='black', lw=1.0,
               label='95% CI includes 0')
    ax.axvline(0, color='black', lw=1.0, ls=(0, (5, 4)), zorder=4)
    for s in range(1, 12):
        ax.axhline(s + 0.5, color='#dddddd', lw=0.8, zorder=1)
    ax.set_yticks(range(1, 13)); ax.set_yticklabels([LAB[s] for s in range(1, 13)],
                                                    fontsize=9)
    ax.invert_yaxis(); ax.set_ylim(12.6, 0.3)
    ax.set_xlabel(r"$\Delta_{stratum}=p_{stratum}-p^{*}_{area}$ (percentage points)", fontsize=11)
    ax.legend(loc='upper center', bbox_to_anchor=(0.5, -0.075), ncol=3,
              fontsize=8.5, frameon=False)   # below the axes: no overlap with the dots
    for sp in ('top', 'right'):
        ax.spines[sp].set_visible(False)
    fig.tight_layout()
    for ext in ('png', 'pdf'):
        fig.savefig(os.path.join(HERE, f"Fig5{lab}_deviations_{stem}.{ext}"),
                    dpi=400 if ext == 'png' else None, bbox_inches='tight')
    plt.close(fig)

    print(f"{title}")
    print(f"   prevalence from {g.p.min():.1f} to {g.p.max():.1f}%, threshold {thr:.1f}%")
    print(f"   deviation from {g.dev.min():+.1f} to {g.dev.max():+.1f} percentage points")
    print(f"   strata whose interval excludes zero: {int(g.sig.sum())} of {len(g)} "
          f"({g.sig.mean()*100:.1f}%)")
    print(f"   -> Fig4{lab}_prevalence_{stem} and Fig5{lab}_deviations_{stem}")
```
