## Supplementary material for "Describing health inequalities without distortion: Simple-Means MAIHDA vs Random-Effects MAIHDA": S6 File. Programs that draw the result figures: fig67_heatmaps.py

```
# -*- coding: utf-8 -*-
"""Figures 6a, 6b, 7a and 7b of the paper: the grid of 25 areas by 12 sociodemographic strata.

Figure 6  the deviation of each area-sociodemographic stratum from the level of its area,
          in percentage points. The level of the area is its composition-standardised
          prevalence (EAR), the plain mean of its 12 stratum proportions, so the deviations
          of every area average zero. Bottom row: the common ARD of each sociodemographic
          stratum, the mean of its column over the 25 areas. Bold: the Wilson 95% interval
          of the stratum, shifted by the level of its area, excludes zero (as in Fig 5).
Figure 7  the modification of the ARD by the area, gamma = observed prevalence minus the
          expected one, where the expected one is the level of the area plus the common ARD
          of the stratum, that is, the deviation of Figure 6 minus the common ARD of its
          column. Bold: |gamma| exceeds 1.96 binomial standard errors of the cell, computed
          with (cases + 1)/(n + 2), the criterion with which the text counts the cells.
Rows: the 25 areas ordered by their composition-standardised prevalence, lowest at the top;
in brackets, that prevalence. Square before the area number: blue, poor area; orange, rich.
Columns: women on the left and men on the right; by age and, within age, low and high income.
Adapted from the programs of Figures 3b and 3c of the companion study of the European Union.
Reads the stratum table (S5 File) and writes the figures next to this script.
    python fig67_heatmaps.py
Needs numpy, pandas and matplotlib.
"""
import os
import numpy as np, pandas as pd, matplotlib
matplotlib.use("Agg")
import matplotlib.pyplot as plt

HERE = os.path.dirname(os.path.abspath(__file__))
t0 = pd.read_csv(os.path.join(HERE, "..", "S5_File_stratum_table", "dataset-smaihda_stratum_table.csv"))
Z = 1.959963984540054
C_POOR, C_RICH = "#1f77b4", "#ff7f0e"           # the colours of Figs 4 and 5
AGE = {1: "35-44", 2: "45-54", 3: "55-64"}; INC = {0: "Low", 1: "High"}
key = t0.groupby("strata_sociodem_id")[["sex", "age3", "income"]].first()
COLS = sorted(key.index, key=lambda s: (key.loc[s, "sex"], key.loc[s, "age3"], key.loc[s, "income"]))
CLAB = ["%s\n%s" % (AGE[key.loc[s, "age3"]], INC[key.loc[s, "income"]]) for s in COLS]
NWOMEN = int((key.sex == 0).sum())
OUT = {"psycmed": ("a", "psycmed"), "private": ("b", "private")}


def fmt(v):
    return "0.0" if abs(v) < 0.05 else "%+.1f" % v


def draw(M, bold, rows, rlab, rich, lim, ticks, label, out, extra=None):
    R, C = M.shape
    fig = plt.figure(figsize=(7.3, 8.4))
    ax = fig.add_axes([0.16, 0.17, 0.82, 0.78])
    cmap = plt.get_cmap("RdBu_r").copy(); cmap.set_bad("white")
    if extra is None:
        Mv, nr, ylab = M, R, list(rlab)
    else:
        Mv = np.vstack([M, np.full((1, C), np.nan), np.asarray(extra[1], float)[None, :]])
        nr, ylab = R + 2, list(rlab) + ["", extra[0]]
    im = ax.imshow(Mv, cmap=cmap, vmin=-lim, vmax=lim, aspect="auto")
    for i in range(R):
        for j in range(C):
            v = M[i, j]
            ax.text(j, i, fmt(v), ha="center", va="center", fontsize=6.3,
                    fontweight="bold" if bold[i, j] else "normal",
                    color="white" if abs(v) > 0.55 * lim else "0.1")
    if extra is not None:
        for j in range(C):
            v = Mv[R + 1, j]
            ax.text(j, R + 1, fmt(v), ha="center", va="center", fontsize=6.3,
                    color="white" if abs(v) > 0.55 * lim else "0.1")
    ax.set_xticks(np.arange(-0.5, C, 1), minor=True); ax.set_yticks(np.arange(-0.5, nr, 1), minor=True)
    ax.grid(which="minor", color="white", lw=1.0); ax.tick_params(which="minor", length=0)
    ax.vlines(NWOMEN - 0.5, -0.5, R - 0.5, color="black", lw=1.3)
    if extra is not None:
        ax.vlines(NWOMEN - 0.5, R + 0.5, R + 1.5, color="black", lw=1.3)
    ax.set_xticks(range(C), CLAB, fontsize=6.8)
    ax.set_yticks(range(nr), ylab, fontsize=7.3)
    if extra is not None:
        ax.get_yticklabels()[-1].set_fontweight("bold")
    ax.tick_params(axis="x", length=0, pad=2); ax.tick_params(axis="y", length=0, pad=16)
    for s in ("top", "right", "left", "bottom"):
        ax.spines[s].set_visible(False)
    for x, txt in ((NWOMEN / 2 - 0.5, "Women"), (NWOMEN + (C - NWOMEN) / 2 - 0.5, "Men")):
        ax.text(x, -0.8, txt, ha="center", va="bottom", fontsize=8.5, fontweight="bold")
    for i, a in enumerate(rows):
        ax.plot(-0.75, i, marker="s", ms=4.0, color=C_RICH if rich[a] else C_POOR, clip_on=False, zorder=5)
    ax.set_xlim(-0.5, C - 0.5); ax.set_ylim(nr - 0.5, -0.5)
    cax = fig.add_axes([0.16, 0.085, 0.82, 0.016])
    cb = fig.colorbar(im, cax=cax, orientation="horizontal", ticks=ticks)
    cb.ax.tick_params(labelsize=7, length=2); cb.set_label(label, fontsize=7.5); cb.outline.set_visible(False)
    h = [plt.Line2D([], [], marker="s", ls="", color=C_POOR, ms=5, label="Poor area"),
         plt.Line2D([], [], marker="s", ls="", color=C_RICH, ms=5, label="Rich area")]
    fig.legend(handles=h, loc="lower center", ncol=2, frameon=False, fontsize=7, bbox_to_anchor=(0.57, 0.005))
    fig.savefig(out + ".png", dpi=400); fig.savefig(out + ".pdf"); plt.close(fig)


for y, (lab, stem) in OUT.items():
    t = t0[t0.outcome == y].copy(); t["p"] = t.cases / t.n
    ear = t.groupby("area").p.mean()
    rich = t.groupby("area").area_income.first() == 1
    t["lev"] = t.area.map(ear)
    t["dev"] = 100 * (t.p - t.lev)
    n = t.n; c = (t.p + Z**2 / (2 * n)) / (1 + Z**2 / n)
    h = Z * np.sqrt(t.p * (1 - t.p) / n + Z**2 / (4 * n * n)) / (1 + Z**2 / n)
    t["sig_dev"] = (t.lev < c - h) | (t.lev > c + h)
    t["gam"] = t.dev - t.groupby("strata_sociodem_id").dev.transform("mean")
    pa = (t.cases + 1) / (t.n + 2)
    t["sig_gam"] = t.gam.abs() > 1.96 * 100 * np.sqrt(pa * (1 - pa) / t.n)
    rows = ear.sort_values().index.tolist()
    k = t.set_index(["area", "strata_sociodem_id"])
    grid = lambda v: np.array([[k.loc[(a, s), v] for s in COLS] for a in rows])
    D, SD, G, SG = grid("dev"), grid("sig_dev").astype(bool), grid("gam"), grid("sig_gam").astype(bool)
    rlab = ["Area %d (%.1f)" % (a, 100 * ear[a]) for a in rows]
    common = D.mean(axis=0)
    limD = int(np.ceil(np.abs(D).max() / 10) * 10); limG = int(np.ceil(np.abs(G).max() / 5) * 5)
    draw(D, SD, rows, rlab, rich, limD, list(range(-limD, limD + 1, 10)),
         "Deviation from the level of the area (percentage points)",
         os.path.join(HERE, "Fig6%s_deviation_heatmap_%s" % (lab, stem)), extra=("Common ARD", common))
    tG = list(range(-limG, limG + 1, 5)) if limG <= 15 else list(range(-(limG // 10) * 10, (limG // 10) * 10 + 1, 10))
    draw(G, SG, rows, rlab, rich, limG, tG,
         "Modification of the ARD, observed minus expected (percentage points)",
         os.path.join(HERE, "Fig7%s_modification_heatmap_%s" % (lab, stem)))
    print("%s | Fig 6%s: deviations %.1f to %.1f, bold %d of %d, common ARD %.1f to %.1f, largest row mean %.1e | "
          "Fig 7%s: gamma %.1f to %.1f, bold %d of %d" % (y, lab, D.min(), D.max(), SD.sum(), D.size, common.min(), common.max(),
                                                     np.abs(D.mean(axis=1)).max(), lab, G.min(), G.max(), SG.sum(), G.size))
```
