## Supplementary material for "Describing health inequalities without distortion: Simple-Means MAIHDA vs Random-Effects MAIHDA": S6 File. Programs that draw the result figures: README.md

# S6 File. Programs for the result figures

Each program draws one or more result figures of the paper from the stratum table
of S5 File. Every figure is computed from the number of individuals and the number
of cases in each stratum, like every measure of the paper. The figures are written
next to the programs.

| Figure | Program | Language |
|---|---|---|
| Fig 3. How an observed prevalence splits into four absolute differences (three areas) | `fig3_decomposition.py` | Python |
| Fig 4a, 4b. Prevalence of the 300 area-sociodemographic strata, with the stratum in all areas pooled | `fig45_prevalence_and_deviations.py` | Python |
| Fig 5a, 5b. Deviation from the composition-standardised (EAR) area prevalence | `fig45_prevalence_and_deviations.py` | Python |
| Fig 6a, 6b. Deviation from the level of the area, map of 25 areas by 12 strata | `fig67_heatmaps.py` | Python |
| Fig 7a, 7b. Modification of the ARD by the area, map of 25 areas by 12 strata | `fig67_heatmaps.py` | Python |
| Fig 8a, 8b. Modification of the sociodemographic differences by the area, observed and expected | `fig8_modification_radar.py` | Python |
| Fig 9. Discriminatory accuracy of the context | `fig9_roc_discriminatory_accuracy.py` | Python |
| Fig 10. The compare diagnostic, stratum by stratum | `fig10_compare_diagnostic.R` | R |
| Numbers of Sections 2.2, 3.1.3 and 3.2.2 that the command does not print, checked against the text | `text_numbers_decomposition.py` | Python |

Figs 1 and 2 are didactic illustrations and plot no data.

## Running the programs

The programs expect the folder layout of the supporting information: this folder,
`S5_File_stratum_table` and `S2_File_smaihda_python.py` side by side. Paths are
resolved from the location of each program, so they can be run from any folder.

```
python fig3_decomposition.py
python fig45_prevalence_and_deviations.py
python fig67_heatmaps.py
python fig8_modification_radar.py
python fig9_roc_discriminatory_accuracy.py
Rscript fig10_compare_diagnostic.R
python text_numbers_decomposition.py
```

The Python programs need numpy, pandas and matplotlib. `fig9_roc_discriminatory_accuracy.py`
takes the coAUC and its 95% interval from the Python implementation of the paper
(S2 File), and checks the classification counts of Table 7. `fig10_compare_diagnostic.R`
needs the lme4 package; it fits the random-intercept logistic model of the compare
diagnostic and stops unless it agrees closely with Table 8, which comes from Stata's
melogit: the range compression factor within one point, the maximum absolute
attenuation within 0.3 points, and the same size of the stratum where that maximum
falls.

The Stata command also draws Fig 4 with its graph option (S1 Text, Step 9), and the deviations from the
crude area prevalence that S2 Text tabulates (Tables A to D). Fig 5 takes the deviations from the
composition-standardised level of the area instead, which average zero within every area (V2.6).

`fig10_compare_diagnostic.R` also prints the latent intraclass correlation and the median odds
ratio of the random-intercept model, for the A-SD strata and for the areas alone (Section 3.4).
