## Supplementary material for "Describing health inequalities without distortion: Simple-Means MAIHDA vs Random-Effects MAIHDA": S6 File. Programs that draw the result figures: text_numbers_decomposition.py

```
# -*- coding: utf-8 -*-
"""Numbers quoted in Sections 2.2, 3.1.3 and 3.2.2 of the paper that are not printed by the
smaihda command. Every number is computed from the stratum table (S5 File), and the program
stops if one of them differs from the text.

  Section 2.2    the variances of the three terms with the crude level of the area add up to
                 102.0% (psychotropic medication use) and 112.1% (private GP choice) of the
                 between-stratum variance, before the noise correction; exactly 100% with the
                 composition-standardised level (EAR)
  Section 3.1.3  the composition gap of the areas (EAR minus crude), on average 0.6 and 1.2
                 percentage points and at most 4.2; the mean deviation in rich minus poor areas
                 of the low-income and high-income strata, private GP choice; the four terms
                 for low-income men aged 35 to 44 in areas 18, 24 and 21, private GP choice
  Section 3.2.2  the cells whose expected prevalence under additivity is below 0% or above
                 100%, and their share of the modification variance
    python text_numbers_decomposition.py
Needs numpy and pandas.
"""
import os
import numpy as np, pandas as pd
HERE = os.path.dirname(os.path.abspath(__file__))
t0 = pd.read_csv(os.path.join(HERE, "..", "S5_File_stratum_table", "dataset-smaihda_stratum_table.csv"))
r1 = lambda v: round(float(v), 1)
for y, total_crude, gap_mean in (("psycmed", 102.0, 0.6), ("private", 112.1, 1.2)):
    g = t0[t0.outcome == y].copy(); g["p"] = g.cases / g.n
    a = g.groupby("area")[["cases", "n"]].sum()
    g["crude"] = g.area.map(a.cases / a.n); g["ear"] = g.area.map(g.groupby("area").p.mean())
    pstar = g.p.mean(); VB = ((g.p - pstar) ** 2).mean()
    share = {}
    for lev in ("crude", "ear"):
        d = g.p - g[lev]; delta = d.groupby(g.strata_sociodem_id).transform("mean"); gam = d - delta
        VC = ((g.groupby("area")[lev].first() - pstar) ** 2).mean()
        VG = (delta.groupby(g.strata_sociodem_id).first() ** 2).mean(); VM = (gam ** 2).mean()
        share[lev] = 100 * (VC + VG + VM) / VB
    gap = 100 * (g.groupby("area").ear.first() - g.groupby("area").crude.first())
    print(f"{y}: sum of the three variances {share['crude']:.1f}% (crude level), {share['ear']:.1f}% (EAR); "
          f"composition gap mean {gap.abs().mean():.2f}, maximum {gap.abs().max():.2f} pp")
    assert r1(share["crude"]) == total_crude and abs(share["ear"] - 100) < 1e-9 and r1(gap.abs().mean()) == gap_mean
    d = g.p - g.ear; delta = d.groupby(g.strata_sociodem_id).transform("mean"); gam = d - delta; exp_ = g.ear + delta
    out = (exp_ < 0) | (exp_ > 1)
    print(f"   expected prevalence outside 0-100%: {out.sum()} cells, {100 * (gam[out] ** 2).sum() / (gam ** 2).sum():.1f}% of the modification variance")
    if y == "private":
        assert r1(gap.abs().max()) == 4.2 and out.sum() == 2 and r1(100 * (gam[out] ** 2).sum() / (gam ** 2).sum()) == 0.5
        rich = g.area_income == 1
        for lev in ("crude", "ear"):
            dd = 100 * (g.p - g[lev])
            lo = dd[rich & (g.income == 0)].mean() - dd[~rich & (g.income == 0)].mean()
            hi = dd[rich & (g.income == 1)].mean() - dd[~rich & (g.income == 1)].mean()
            print(f"   rich minus poor areas, {lev} level: low-income {lo:+.1f}, high-income {hi:+.1f} pp")
            if lev == "ear": assert (r1(lo), r1(hi)) == (-1.3, 1.3)
            else: assert r1(lo) == -2.8
        s = 7          # low-income men aged 35 to 44
        for area, obs in ((18, 10.0), (24, 15.9), (21, 3.5)):
            r = g[(g.area == area) & (g.strata_sociodem_id == s)].iloc[0]; i = r.name
            print(f"   area {area}: reference {100*pstar:.1f} + area ARD {100*(r.ear-pstar):+.1f} + common ARD {100*delta[i]:+.1f}"
                  f" + modification {100*gam[i]:+.1f} = {100*r.p:.1f}% ({r.cases} of {r.n}); expected {100*exp_[i]:.1f}%")
            assert r1(100 * r.p) == obs
    else:
        assert out.sum() == 0
print("all numbers agree with the text")
```
