## Supplementary material for "Describing health inequalities without distortion: Simple-Means MAIHDA vs Random-Effects MAIHDA": S7 Text. Simulation supporting the standardised contextual AUC

### **S7 Text. Simulation supporting the standardised contextual AUC (coAUC*)**

*Companion to Section 2.2.2 of the main text. It supports the claims made there about coAUC*, and it also states what coAUC* costs.*

#### **1. Purpose**

Four questions are answered.

Question 1. Does coAUC* order contexts in the same way as VPC*, and does that agreement survive when the strata differ in size?

Question 2. Is coAUC* invariant to the distribution of stratum sizes when the true contextual structure is held fixed?

Question 3. How do the two measures behave when the prevalence of the outcome changes and the contextual structure is held fixed on the latent scale?

Question 4. What does size standardisation cost in precision?

#### **2. Design**

The simulation has 100 strata and 200,000 individuals, with 400 replicates per scenario and seed 2026. A latent normal effect per stratum gives the stratum risk on the probability scale. Stratum sizes are drawn from a lognormal distribution whose spread is the experimental factor, with the total sample size held constant, so that the only thing changing across scenarios is how unequally the same people are distributed over the same strata. The overall prevalence is 15% except in Question 3, where it is the factor of interest.

Three measures are computed from the stratum risks and sizes alone. VPC*, the standardised VPC, gives all strata the same size and does not involve the sizes. The contextual AUC, coAUC, counts pairs of individuals and therefore depends on the sizes. coAUC* gives every stratum the same total weight, which is the value that would be obtained if all strata were of equal size. Ties are counted at one half throughout.

#### **3. Question 1. Agreement with VPC***

The true contextual effect varies across replicates, so the rank correlation measures whether the two summaries order the same set of populations in the same way.

| **Spread of log size** | **Largest stratum** | **Largest / smallest** | **Spearman (VPC*, coAUC)** | **Spearman (VPC*, coAUC*)** |
| --- | --- | --- | --- | --- |
| 0.0 | 1.0% | 1 | 0.994 | 0.994 |
| 0.5 | 3.0% | 12 | 0.989 | 0.996 |
| 1.0 | 7.0% | 141 | 0.951 | 0.995 |
| 1.5 | 14.3% | 1,994 | 0.879 | 0.994 |
| 2.0 | 21.1% | 8,175 | 0.782 | 0.992 |
| 2.5 | 33.6% | 13,432 | 0.632 | 0.993 |

*Table S7.1. Rank correlation with VPC* across six degrees of size inequality.*

coAUC* keeps a rank correlation of 0.99 with VPC* in every scenario. The coAUC keeps it only while the strata are of similar size, and loses it as the population concentrates, falling to 0.63 when the largest stratum holds a third of the individuals. coAUC* is therefore a reading of the same quantity that VPC* measures, expressed on the discrimination scale.

#### **4. Question 2. Invariance to the distribution of sizes**

Here the contextual structure is identical in every row, so any movement of a measure is an artefact of how the population is distributed.

| **Spread of log size** | **Largest stratum** | **VPC* (%)** | **coAUC (%)** | **coAUC* (%)** |
| --- | --- | --- | --- | --- |
| 0.0 | 1.0% | 12.88 | 74.29 | 74.29 |
| 0.5 | 3.0% | 12.90 | 74.27 | 74.30 |
| 1.0 | 7.0% | 13.04 | 74.00 | 74.39 |
| 1.5 | 14.7% | 12.85 | 73.17 | 74.24 |
| 2.0 | 21.4% | 12.90 | 72.04 | 74.29 |
| 2.5 | 31.8% | 12.77 | 69.59 | 74.19 |

*Table S7.2. The same underlying stratum risks with the size distribution changing.*

The coAUC falls by 4.7 points, from 74.3% to 69.6%, although nothing about the contextual boundaries has changed. coAUC* stays between 74.2% and 74.4%, as does VPC*. Two studies of the same phenomenon with differently distributed populations would report different coAUC and the same coAUC*.

#### **5. Question 3. Behaviour under the prevalence of the outcome**

The contextual structure is now held fixed on the latent scale, which is the scale on which it is free of the prevalence, and the prevalence is moved from 5% to 45%. The strata are of equal size, so the coAUC and coAUC* coincide and only coAUC* is shown.

| **Latent ICC** | **5%** | **10%** | **20%** | **30%** | **45%** | **Ratio** |
| --- | --- | --- | --- | --- | --- | --- |
| 1% | 0.16 | 0.30 | 0.52 | 0.69 | 0.80 | 5.0 |
| 5% | 0.91 | 1.64 | 2.80 | 3.42 | 3.92 | 4.3 |
| 10% | 2.12 | 3.69 | 5.71 | 6.91 | 7.59 | 3.6 |
| 20% | 5.83 | 8.80 | 12.12 | 13.86 | 14.89 | 2.6 |
| 35% | 13.94 | 18.30 | 22.17 | 24.12 | 25.13 | 1.8 |

*Table S7.3. Size-standardised VPC (%) at five prevalences, with the ratio of the largest to the smallest value in the row.*

| **Latent ICC** | **5%** | **10%** | **20%** | **30%** | **45%** | **Swing** |
| --- | --- | --- | --- | --- | --- | --- |
| 1% | 55.08 | 55.07 | 55.04 | 55.06 | 55.06 | 0.04 |
| 5% | 61.39 | 61.34 | 61.41 | 61.24 | 61.23 | 0.18 |
| 10% | 66.14 | 66.12 | 65.98 | 65.85 | 65.69 | 0.44 |
| 20% | 73.03 | 72.73 | 72.40 | 72.25 | 72.13 | 0.91 |
| 35% | 80.29 | 79.70 | 79.22 | 79.01 | 78.86 | 1.44 |

*Table S7.4. coAUC* (%) at the same five prevalences, with the range of the row in percentage points.*

With the same contextual structure, VPC* reads five times higher at a prevalence of 45% than at 5% when the latent ICC is 1%, and 1.8 times higher when it is 35%. coAUC* moves by 0.04 points in the first case and by 1.44 points in the second. The reason is that a rank-based measure does not see the base rate, whereas a measure in percentage points is bounded by the room that the base rate leaves.

The agreement is closest where the categorisation of Table 1 does most of its work, that is, at contextual effects that are small to moderate. At very large contextual effects coAUC* begins to drift, so a comparison across outcomes in that range should be read with the prevalences in view.

#### **6. Question 4. The price of standardising**

Individuals are sampled in this section, so each stratum proportion carries binomial noise, and the table reports the standard deviation of each measure over 400 samples from the same population.

| **Spread of log size** | **Smallest stratum (n)** | **SD of coAUC (points)** | **SD of coAUC* (points)** | **Ratio** |
| --- | --- | --- | --- | --- |
| 0.0 | 2,000 | 0.136 | 0.136 | 1.00 |
| 0.5 | 334 | 0.126 | 0.159 | 1.26 |
| 1.0 | 146 | 0.131 | 0.227 | 1.73 |
| 1.5 | 38 | 0.104 | 0.315 | 3.02 |
| 2.0 | 5 | 0.120 | 0.550 | 4.60 |
| 2.5 | 5 | 0.148 | 1.097 | 7.40 |

*Table S7.5. Sampling variability of the two measures under binomial sampling.*

Size standardisation gives full weight to the smallest strata, which are the noisiest, so coAUC* is the less precise of the two. With strata of equal size the two are equally precise. With a spread of 2.5, where the smallest stratum holds five individuals, coAUC* is seven times more variable than the coAUC. This is the same trade that an age-standardised rate makes against a crude rate, and it is the reason the interval is reported alongside the estimate. The command excludes strata below a minimum size, ten individuals by default, for this reason. The simulation keeps them, since its purpose here is to price the cost of counting them.

#### **7. What the simulation does not show**

The stratum risks are generated from a normal distribution on the logit scale. This is the assumption that Simple-Means MAIHDA declines in estimation, and it is used here only to produce populations with a known contextual structure against which the measures can be compared. Nothing in the definition of coAUC* depends on it.

The simulation compares measures computed on known stratum risks. It does not address the estimation of those risks, except in Question 4, where binomial sampling is introduced for the single purpose of measuring precision.

The results concern one contextual level with strata that are exchangeable in size only. They do not speak to the area and stratum decomposition of the main text, which is a separate question.

#### **8. Reproducing the results**

Every number in this text is produced by S7 File, a self-contained Python program that requires only numpy and runs in under a minute. Seed 2026.

*python S7_File_simulation_AUCstar.py*
