## Supplementary material for "Describing health inequalities without distortion: Simple-Means MAIHDA vs Random-Effects MAIHDA": S7 File. The simulation as a runnable Python script: S7_File_simulation_AUCstar.py

```
"""
S7 Text. Simulation supporting the size-standardised AUC (AUC*).

Four questions are answered.

  1. Does AUC* order contexts in the same way as the size-standardised VPC,
     and does that agreement survive when the strata differ in size?
  2. Is AUC* invariant to the distribution of stratum sizes when the true
     contextual structure is held fixed?
  3. How do the measures behave when the prevalence of the outcome changes
     and the contextual structure is held fixed on the latent scale?
  4. What does size standardisation cost in precision?

Design. K strata. A latent normal effect per stratum, u_k ~ N(0, s2u), gives
the stratum risk p_k = logistic(b0 + u_k). Stratum sizes are drawn from a
lognormal whose spread SIGMA_N is the experimental factor, with the total
sample size held constant, so the only thing that changes across scenarios is
how unequally the same people are distributed over the same strata.

Measures, all computed from the stratum risks and sizes alone.

  VPC*   var(p_k) / (p* (1 - p*)) with every stratum counted once, p* the
         plain mean of the stratum risks. Does not involve the sizes.
  AUC    probability that a randomly drawn case belongs to a stratum of higher
         risk than a randomly drawn non-case, counting individuals, ties at
         one half. Depends on the sizes.
  AUC*   the same quantity with every stratum contributing the same total
         weight, which is the value that would be obtained if all strata were
         of equal size.

Run: python S7_File_simulation_AUCstar.py
"""
import numpy as np

RNG = np.random.default_rng(2026)
K, N_TOTAL, R, PREV = 100, 200_000, 400, 0.15
SIGMA_N = [0.0, 0.5, 1.0, 1.5, 2.0, 2.5]
PI3 = np.pi ** 2 / 3

logistic = lambda x: 1.0 / (1.0 + np.exp(-x))
logit = lambda p: np.log(p / (1 - p))


def auc(n, p):
    """AUC with each stratum weighted by n; ties at one half."""
    o = np.argsort(p)
    ca, no = (n * p)[o], (n * (1 - p))[o]
    below = np.concatenate(([0.0], np.cumsum(no)[:-1]))
    return ((ca * below).sum() + 0.5 * (ca * no).sum()) / (ca.sum() * no.sum())


def auc_std(p):
    """AUC* : every stratum contributes the same total weight."""
    return auc(np.ones(len(p)), p)


def vpc_std(p):
    ps = p.mean()
    return p.var() / (ps * (1 - ps))


def sizes(sigma_n):
    w = np.exp(sigma_n * RNG.normal(0.0, 1.0, K))
    return np.maximum(np.round(N_TOTAL * w / w.sum()), 5.0)


def spearman(a, b):
    ra = np.argsort(np.argsort(a)).astype(float)
    rb = np.argsort(np.argsort(b)).astype(float)
    return np.corrcoef(ra, rb)[0, 1]


b0 = logit(PREV)
line = lambda n=92: print("-" * n)

print(f"S7 Text simulation | K = {K} strata | N = {N_TOTAL:,} | {R} replicates "
      f"per scenario | prevalence {PREV:.0%} | seed 2026\n")

# ---- Question 1 ---------------------------------------------------------
print("QUESTION 1. Rank correlation with the size-standardised VPC.")
print("The true contextual effect varies across replicates; only the size spread changes.\n")
print(f"{'spread of':>10}{'largest':>10}{'max/min':>10} | {'rho(VPC*, AUC)':>16}{'rho(VPC*, AUC*)':>17}")
print(f"{'log size':>10}{'stratum':>10}{'size':>10} |")
line(68)
for sg in SIGMA_N:
    V, A, As, top, rat = [], [], [], [], []
    for _ in range(R):
        s2u = RNG.uniform(0.05, 2.5)
        p = logistic(b0 + RNG.normal(0, np.sqrt(s2u), K))
        n = sizes(sg)
        V.append(vpc_std(p)); A.append(auc(n, p)); As.append(auc_std(p))
        top.append(n.max() / n.sum()); rat.append(n.max() / n.min())
    print(f"{sg:10.1f}{np.median(top):9.1%}{np.median(rat):10.0f} | "
          f"{spearman(V, A):16.3f}{spearman(V, As):17.3f}")

# ---- Question 2 ---------------------------------------------------------
print("\n\nQUESTION 2. The same underlying risks; only the size distribution changes.")
print("s2u fixed at 1.0, so the contextual structure is identical in every row.\n")
print(f"{'spread of':>10}{'largest':>10} | {'VPC* %':>9}{'AUC %':>9}{'AUC* %':>9}")
print(f"{'log size':>10}{'stratum':>10} |")
line(50)
for sg in SIGMA_N:
    V, A, As, top = [], [], [], []
    for _ in range(R):
        p = logistic(b0 + RNG.normal(0, 1.0, K))
        n = sizes(sg)
        V.append(vpc_std(p)); A.append(auc(n, p)); As.append(auc_std(p))
        top.append(n.max() / n.sum())
    print(f"{sg:10.1f}{np.median(top):9.1%} | {np.mean(V)*100:9.2f}"
          f"{np.mean(A)*100:9.2f}{np.mean(As)*100:9.2f}")

# ---- Question 3 ---------------------------------------------------------
print("\n\nQUESTION 3. The same contextual structure on the latent scale; the prevalence moves.")
print("Strata of equal size, so the crude AUC and AUC* coincide and only AUC* is shown.\n")
PREVS = (0.05, 0.10, 0.20, 0.30, 0.45)
print(f"{'latent ICC':>11} | " + "".join(f"{f'{q:.0%}':>20}" for q in PREVS) + f"{'  swing':>10}")
print(f"{'':11} | " + "".join(f"{'VPC*      AUC*':>20}" for _ in PREVS))
line(11 + 3 + 20 * len(PREVS) + 10)
for ic in (0.01, 0.05, 0.10, 0.20, 0.35):
    s = np.sqrt(ic * PI3 / (1 - ic))
    cells, Vv, Av = [], [], []
    for q in PREVS:
        V, As = [], []
        for _ in range(R):
            p = logistic(logit(q) + RNG.normal(0, s, K))
            V.append(vpc_std(p)); As.append(auc_std(p))
        v, a = np.mean(V) * 100, np.mean(As) * 100
        Vv.append(v); Av.append(a)
        cells.append(f"{v:9.2f}{a:11.2f}")
    print(f"{ic:10.0%} | " + "".join(cells) +
          f"   VPC* x{max(Vv)/min(Vv):.1f}, AUC* {max(Av)-min(Av):.2f} pts")

# ---- Question 4 ---------------------------------------------------------
print("\n\nQUESTION 4. The price of standardising: precision.")
print("Individuals are now sampled, so each stratum proportion carries binomial noise.")
print("Standard deviation over 400 replicates of the same population.\n")
print(f"{'spread of':>10}{'smallest':>10} | {'SD of AUC':>12}{'SD of AUC*':>13}{'ratio':>8}")
print(f"{'log size':>10}{'stratum':>10} |")
line(55)
for sg in SIGMA_N:
    n = sizes(sg)
    p_true = logistic(b0 + RNG.normal(0, 1.0, K))
    A, As = [], []
    for _ in range(R):
        p_hat = RNG.binomial(n.astype(int), p_true) / n
        A.append(auc(n, p_hat)); As.append(auc_std(p_hat))
    sa, sas = np.std(A) * 100, np.std(As) * 100
    print(f"{sg:10.1f}{n.min():10.0f} | {sa:12.3f}{sas:13.3f}{sas/sa:8.2f}")

print("\nSize standardisation gives full weight to the smallest and noisiest strata,")
print("so AUC* is the less precise of the two. This is the same trade that an")
print("age-standardised rate makes against a crude rate.")
```
