## Supplementary material for "Describing health inequalities without distortion: Simple-Means MAIHDA vs Random-Effects MAIHDA": S1 Checklist. STROBE checklist

**S1 Checklist. STROBE checklist for cross-sectional studies**

Manuscript: Describing health inequalities without distortion: Simple-Means MAIHDA vs Random-Effects MAIHDA. Locations are given as manuscript section numbers rather than page numbers, so that they remain valid across typeset versions.

| **Item** | **Recommendation** | **Section** | **Note** |
| --- | --- | --- | --- |
| **Title and abstract** | | | |
| **1a** | Indicate the study's design with a commonly used term in the title or the abstract | Abstract | See note 1 |
| **1b** | Provide in the abstract an informative and balanced summary of what was done and what was found | Abstract |  |
| **Introduction** | | | |
| **2** | Explain the scientific background and rationale for the investigation being reported | 1 |  |
| **3** | State specific objectives, including any prespecified hypotheses | 1 |  |
| **Methods** | | | |
| **4** | Present key elements of study design early in the paper | 2.1 | See note 2 |
| **5** | Describe the setting, locations, and relevant dates, including periods of recruitment, exposure, follow-up, and data collection | 2.1 | Malmo, Sweden, calendar year 2006 |
| **6a** | Give the eligibility criteria, and the sources and methods of selection of participants | 2.1 | Residents aged 35 to 64 with at least one primary healthcare contact in 2006 |
| **7** | Clearly define all outcomes, exposures, predictors, potential confounders, and effect modifiers; give diagnostic criteria if applicable | 2.1 | Two binary outcomes, ATC codes given; strata defined by sex, age group and income |
| **8** | For each variable of interest, give sources of data and details of methods of assessment; describe comparability of assessment methods if there is more than one group | 2.1; Ethics statement | Record linkage of Swedish national registers |
| **9** | Describe any efforts to address potential sources of bias | 4.3; 4.4; 4.7 | Shrinkage bias is the object of the paper; the compare diagnostic makes it visible; binomial noise is discounted explicitly |
| **10** | Explain how the study size was arrived at | 2.1 | No sample size calculation. The study includes the whole eligible population, N = 43,291 |
| **11** | Explain how quantitative variables were handled in the analyses; if applicable, describe which groupings were chosen and why | 2.1; 4.7 | Income dichotomised at the Malmo median equivalised household disposable income; age in three ten-year groups |
| **12a** | Describe all statistical methods, including those used to control for confounding | 2.2; 2.2.1 to 2.2.4 | Descriptive study; no confounding control is attempted, and the reason is given in the Discussion |
| **12b** | Describe any methods used to examine subgroups and interactions | 2.2.2; 3.2.2 | Decomposition into clustering, additive differences and modification |
| **12c** | Explain how missing data were addressed | 2.1 | See note 3 |
| **12d** | If applicable, describe analytical methods taking account of sampling strategy | 2.3 | See note 4 |
| **12e** | Describe any sensitivity analyses | 2.2.4; 3.4; S1 Text; S3 Text | Comparison against random-effects MAIHDA through the compare diagnostic, and a worked example in the supporting information |
| **Results** | | | |
| **13a** | Report numbers of individuals at each stage of the study | 2.1; Fig 1 | See note 5 |
| **13b** | Give reasons for non-participation at each stage | 2.1 | Not applicable, see note 5 |
| **13c** | Consider use of a flow diagram | Fig 1 | Fig 1 shows the nested structure and the numbers at each level |
| **14a** | Give characteristics of study participants and information on exposures and potential confounders | 3.1.1; Table 2 |  |
| **14b** | Indicate number of participants with missing data for each variable of interest | 2.1 | See note 3 |
| **15** | Report numbers of outcome events or summary measures | 3.1; Tables 2 to 4 | Observed prevalences with 95% confidence intervals for every stratum |
| **16a** | Give unadjusted estimates and, if applicable, confounder-adjusted estimates and their precision; make clear which confounders were adjusted for and why they were included | 3.1 to 3.3 | All estimates are unadjusted observed proportions, by design |
| **16b** | Report category boundaries when continuous variables were categorized | 2.1 | Median of the Malmo distribution for income; ten-year bands for age |
| **16c** | If relevant, consider translating estimates of relative risk into absolute risk for a meaningful time period | 2.2.1; 3.1 | All measures are reported on the probability scale, as prevalences and absolute risk differences in percentage points |
| **17** | Report other analyses done, such as analyses of subgroups and interactions, and sensitivity analyses | 3.2.2; 3.4 |  |
| **Discussion** | | | |
| **18** | Summarise key results with reference to study objectives | 4.1 |  |
| **19** | Discuss limitations of the study, taking into account sources of potential bias or imprecision; discuss both direction and magnitude of any potential bias | 4.7 | Direction and magnitude of the shrinkage bias are quantified in 3.4 and 4.4 |
| **20** | Give a cautious overall interpretation of results considering objectives, limitations, multiplicity of analyses, results from similar studies, and other relevant evidence | 4.2 to 4.6; 4.8; 4.9 |  |
| **21** | Discuss the generalisability (external validity) of the study results | 4.7 | One city, one calendar year, Swedish primary healthcare |
| **Other information** | | | |
| **22** | Give the source of funding and the role of the funders for the present study and, if applicable, for the original study on which the present article is based | Funding statement |  |

**Notes**

Note 1. The design term is stated in the Abstract and again at the start of section 2.1.

Note 2. The study is a cross-sectional analysis of a register-based dataset for one calendar year.

Note 3. The analytic dataset is a complete register extract for the eligible population and contains no missing values on the variables used, so no imputation or exclusion was required.

Note 4. The dataset is the eligible population rather than a probability sample, so no survey weights apply. Uncertainty for the summary measures is estimated by a within-context bootstrap. The areas and the area-sociodemographic strata are the observed set and are held fixed, and individuals are resampled within each stratum, so the interval reflects the uncertainty of measuring each proportion and not the variability of which contexts were observed.

Note 5. The study population is defined by the eligibility criteria in section 2.1 and no participants were excluded thereafter, so there are no successive stages of participation and no non-participation to report. Fig 1 gives the numbers at each level of the structure.

STROBE: Strengthening the Reporting of Observational Studies in Epidemiology. Checklist version for cross-sectional studies, available at https://www.strobe-statement.org
